# The genetic risk for COVID-19 severity is associated with defective innate immune responses

**DOI:** 10.1101/2020.11.10.20229203

**Authors:** Yunus Kuijpers, Xiaojing Chu, Martin Jaeger, Simone J.C.F.M. Moorlag, Valerie A.C.M. Koeken, Bowen Zhang, Aline de Nooijer, Inge Grondman, Nico Janssen, Vera P. Mourits, L. Charlotte J. de Bree, Quirijn de Mast, Frank L. van de Veerdonk, Leo A.B. Joosten, Yang Li, Mihai G. Netea, Cheng-Jian Xu

## Abstract

Recent genome-wide association studies (GWASs) of COVID-19 patients of European ancestry have identified genetic loci significantly associated with disease severity (*1*). Here, we employed the detailed clinical, immunological and multi-omics dataset of the Human Functional Genomics Projects (HFGP) to explore the physiological significance of the host genetic variants that influence susceptibility to severe COVID-19. A genomics investigation intersected with functional characterization of individuals with high genetic risk for severe COVID-19 susceptibility identified several major patterns: i. a large impact of genetically determined innate immune responses in COVID-19, with increased susceptibility for severe disease in individuals with defective monocyte-derived cytokine production; ii. genetic susceptibility related to ABO blood groups is probably mediated through the von Willebrand factor (VWF) and endothelial dysfunction. We further validated these identified associations at transcript and protein levels by using independent disease cohorts. These insights allow a physiological understanding of genetic susceptibility to severe COVID-19, and indicate pathways that could be targeted for prevention and therapy.

**One Sentence summary:** In this study, we explore the physiological significance of the genetic variants associated with COVID-19 severity using detailed clinical, immunological and multi-omics data from large cohorts. Our findings allow a physiological understanding of genetic susceptibility to severe COVID-19, and indicate pathways that could be targeted for prevention and therapy.

## Introduction

The novel coronavirus disease 2019 (COVID-19), caused by the severe acute respiratory syndrome coronavirus 2 (SARS-CoV-2) (*2*)(*3*), firstly emerged in late December 2019 and has been spreading worldwide very quickly. The COVID-19 pandemic creates a severe disruption to the healthcare system and endangers the economy. While much has been learned about the pathophysiology of the disease, and immune-based treatment proven to be effective has included dexamethasone (*4*) and anti-IL-6/IL-6R monoclonal antibodies(*5*)(*6*), the prognosis is still poor in many patients. Therefore, there is an urgent need to better understand the exact host-pathogen interactions leading to increased severity and mortality, in order to design additional prophylactic and therapeutic strategies in future (*7*)(*8*).

The severity of SARS-CoV-2 infection is highly variable, and ranges from asymptomatic to mild disease, and even to severe Acute Respiratory Distress Syndrome with a fatal outcome (*9*). However, the causes for this broad variability in disease outcome between individuals are largely unknown. A recent study indicates that human host factors rather than viral genetic variation affect COVID-19 severity outcome (*10*). Additionally, clinical and epidemiological data have shown that old age, male sex, and chronic comorbidity are associated with higher mortality (*11*)(*12*). The first genome-wide association study in individuals with genetic European ancestry has identified several chemokine receptor genes, including *CCR9*, *CXCR6* and *XCR1* and the locus controlling the ABO blood type to be associated with severe symptoms of COVID-19 (*1*). Nevertheless, little is known about the mechanisms through which these genetic variants influence COVID-19 severity. For example, several competing hypotheses may be envisaged for the involvement of immune genes in susceptibility to severe COVID-19: on the one hand, it may be hypothesized that genetic risk for severe COVID-19 is associated with defective innate immune responses that would allow viral multiplication with high viral loads; on the other hand, the opposite hypothesis may also be true, with an exaggerated genetically-mediated cytokine production being responsible for the late phase hyperinflammation and poor outcome. A purely genetic study cannot respond to this crucial question, that would have important consequences for the approach to prophylaxis and therapy.

By making use of resources from the Human Functional Genomics Project (HFGP) (*13, 14*), we assessed the impact of COVID-19 associated genetic polymorphisms on variability of immune responses at the population level (Fig. 1), and we validated our findings using single cell transcriptomics and proteomics data from two independent COVID-19 cohorts. This study will help us to understand how genetic variability is related to disease susceptibility through the regulation of immune responses and endothelial function.

**Fig. 1.**
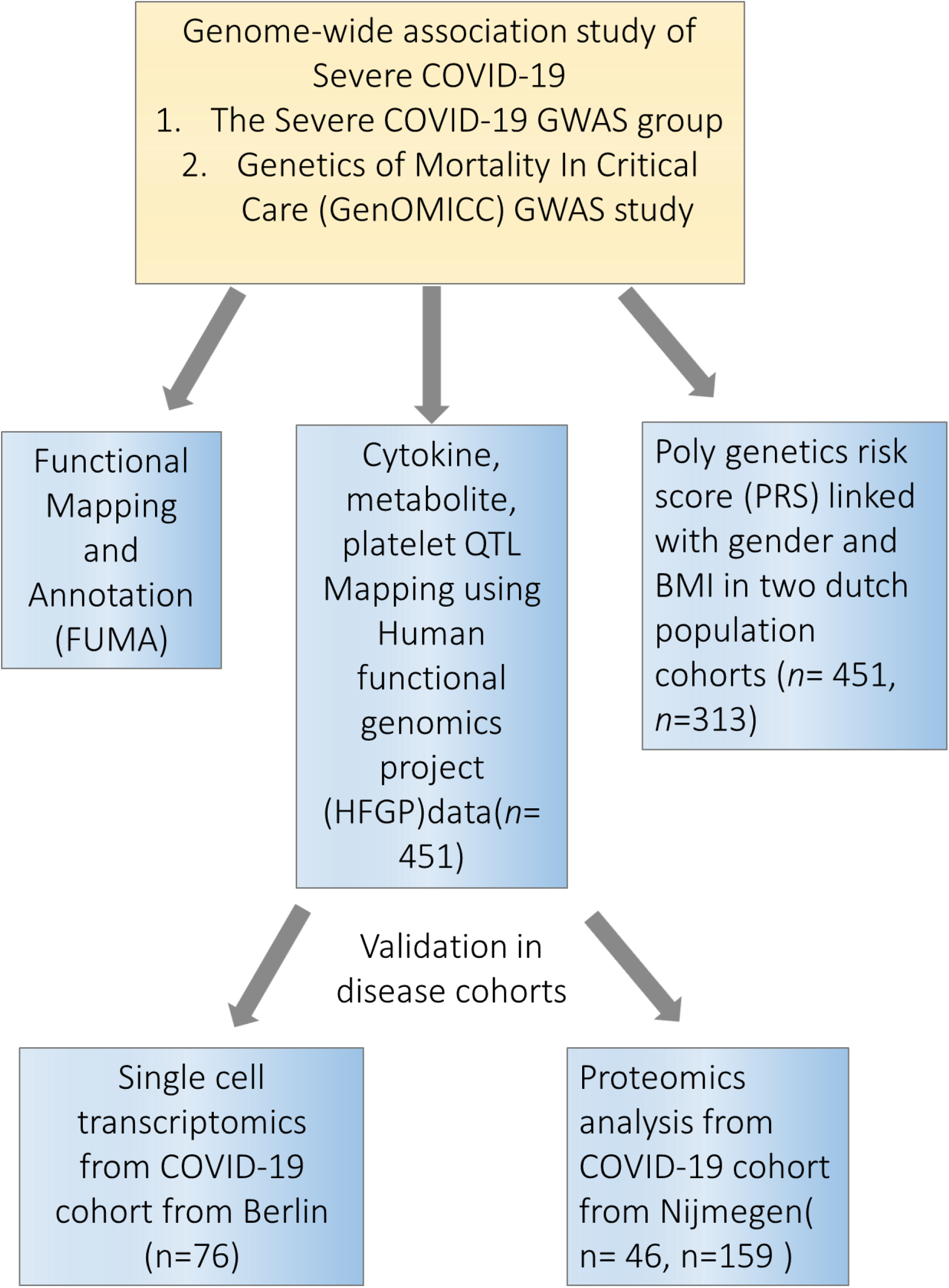
Study overview. Firstly, we performed a functional mapping and annotation (FUMA) to link COVID-19 SNPs to gene expression and identified important pathways and tissues contributing to the pathophysiology of COVID-19. Secondly, we utilized the cytokine quantitative trait loci (QTL), metabolite QTL and plate QTL from Human Functional Genomics Projects (HFGP) 500FG data (n=451) to test if specific loci are associated with immune functions. Thirdly, we linked PRS score with gender and BMI in 500FG (n=451) and 300BCG (n=313) cohorts. Lastly, we validated our findings in disease cohorts in single- cell transcriptomics data from Berlin (n=76), and proteomics data from Nijmegen (n=46, n=159). SNPs, single-nucleotide polymorphisms.

## Results

### COVID-19 loci are enriched for expression in immune organs, chemokine signaling pathways and enhancer region

To explore the functional impact of the identified COVID-19 loci, we firstly investigated if the identified independent genetic loci (p value <1×10^-5^) from the first COVID-19 GWAS study by Severe COVID-19 GWAS Group (*1*) are associated with any phenotypes available at the GWAS catalog (https://www.ebi.ac.uk/gwas/). We found that many of these loci are associated with traits such as blood proteins/biomarkers, LDL and VLDL concentrations (Table S1). We next performed functional annotation of significant loci and gene-mapping using Functional Mapping and Annotation (FUMA) (*15*). The SNP2GENE function identified 32 independent SNPs located in 26 different loci which reached suggestive significance in this study (p-value <1×10^-5^, Table S2). Using multiple independent expression quantitative trait loci (eQTL) datasets, FUMA mapped 115 genes to these 26 genomic risk loci. Using RNA-seq data of 30 tissues from GTEx database (v8), we found significant enrichment of candidate genes in expression in immune organs such as spleen and blood (Fig. 2A), suggesting that they are important tissues contributing to the pathophysiology of COVID-19 (*16*)(*17*). Moreover, we observed the enrichment of candidate genes to be mainly expressed in small intestine and lung (Fig. 2A), suggesting that COVID-19 represents a multisystem illness with involvement of different organs, consistent with the respiratory and intestinal symptoms of the disease (*18*).

**Fig. 2.**
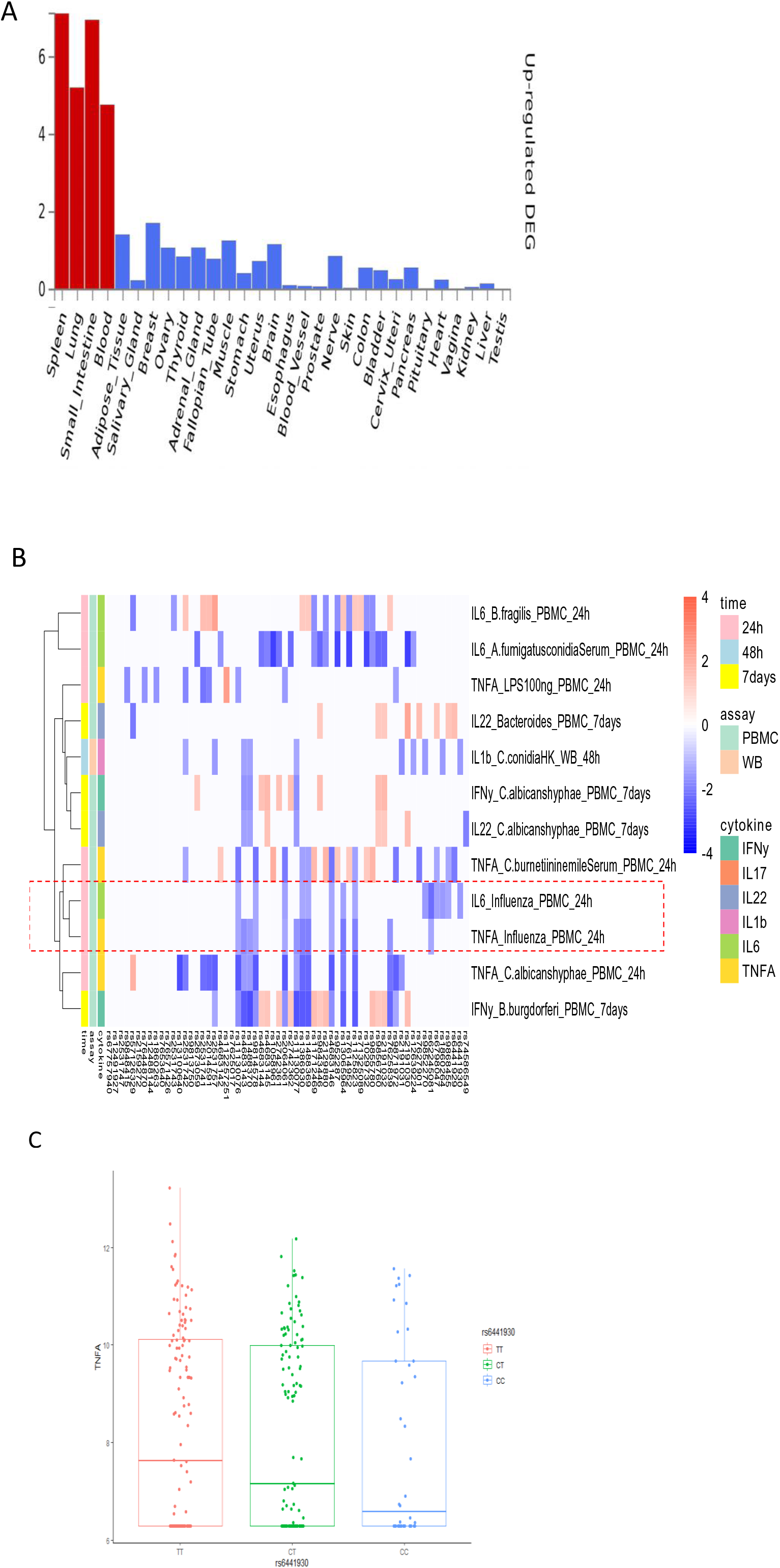
Functional annotation of COVID-19 loci using the FUMA pipeline and association 3p21.31 loci with immune traits. A) MAGMA Tissue expression results on 30 general tissues type (GTEx v8). FUMA analysis was done based on genes identified after using their genomic location, eQTL associations, and histone activity. B) The heatmap showing the assoication between 3p21.31 loci with cytokine production upon *in vitro* stimulations. Red color in heatmap indicates higher cytokine production leaded by risk allele in COVID-19 GWAS profiles, Blue color indicates lower cytokine production leaded by risk allele in COVID-19 GWAS profiles. C) a boxplot showing COVID-19 risk allele(rs6441930-C) associated with reduced IL6 production with influenza stimulation of PBMC for 24 hours (p- value = 0.026).

Pathway analysis using these 115 genes showed a strong enrichment in chemokine binding and chemokine receptors binding (Fig. S1), which is in line with the fact that chemokines can recruit immune cells to the site of infection and are critical for the function of the immune response (*19*). In addition, chemokines have been reported as the most significantly elevated biomarkers in patients with severe COVID-19 on the intensive care unit (*17*).

Considering that all SNPs in Linkage Disequilibrium (LD) with the 32 independent loci (p- value <1×10^-5^) identified by the COVID-19 GWAS, were significantly enriched in the non-coding intronic region (p value = 0.036, Fisher’s exact test) (Table S3), we next examined whether the COVID-19 associated variants are enriched in regulatory DNA elements. We interrogated all significant SNPs (p-value<1×10^-5^ and stricter thresholds) with histone marks and chromatin states of 24 blood cell-types in the Roadmap Epigenome Project (*20*). We found that the COVID-19 genetic loci were strongly enriched for enhancer markers and weakly enriched in promoter marker (Table S4). The strong enrichment of COVID-19 loci in enhancer marks indicates that the associated genetic variants are likely to be involved in the regulation of immunologically-related functions. This finding also suggests that epigenetic mechanisms/regulation may play an important role in the pathogenesis of COVID-19 infection.

### 3p21.31 loci are associated with lower production of monocyte-derived cytokines

Severe COVID-19 is characterized by complex immune dysregulation, combining immune defective features with hyperinflammatory innate immune traits (*21*)(*22*). However, these analyses in patients could be performed only late during the disease, and whether genetic risk for severe COVID-19 is characterized by low or high innate immune responses in a non-infected person is not known. We therefore used the cytokine QTL data from the 500FG cohort (*14*) of the HFGP to test whether SNPs in 3p21.31 influence cytokine production upon stimulation. We checked all SNPs located within a 50 kilobase window of top variant rs11385942, and showed all nominal significant associations (p-value < 0.05, Fig. 2A). Interestingly, we observed that the risk alleles for a severe course of COVID-19 are consistently associated with lower production of monocyte-dependent cytokines (IL-6, IL-1β and TNF-α) upon various *in-vitro* stimulations (Fig. 2B). Of note, COVID-19 risk alleles also correspond to lower monocyte-derived cytokine production after influenza stimulation, a viral stimulus (Fig. 2B and C, Table S5). It is thus tempting to speculate that the people who carry risk alleles may not respond properly to an initial virus infection, leading to high viral loads, subsequent systemic inflammation and poor outcome. Next, we tested whether the COVID-19 risk SNPs are associated with the concentrations of circulating cytokines and levels of metabolites in blood (Table S6-7). Using the same cohort, we found that IL-18 and IL-18BP concentrations show a suggestive positive association with genetic risk of COVID-19 (Fig. S2, Table S6). Additionally, one of the 3p21.31 loci, rs2191031 is suggestively negative associated with high density lipoprotein cholesterol (HDL) (P value =0.004, Table S7-S8), which is consistent with previous findings that HDL levels were significantly lower in the severe COVID-19 disease group (*23*).

### Von Willebrand Factor (VWF) and lymphocytes are strongly colocalized with ABO loci

It is known that ABO blood group influences the plasma levels of von Willebrand factor (VWF)(*24*) and elevated VWF levels are associated with severe COVID-19 (*25*). We therefore tested the association of ABO locus with VWF circulating concentrations from the individuals in the 500FG cohort. Of note, we found the risk allele rs687621-G is significantly associated with elevated levels of VWF (p-value = 9.58×10^-20^) (Fig. 3A and 3B). Recent studies have reported that the VWF level is highly related to COVID-19 severity (*26, 27*). As VWF level in plasma is an indicator of inflammation, endothelial activation and damage (*28*), our results suggest that the association of VWF and COVID-19 severity is very likely mediated through genetic regulation.

**Fig 3.**
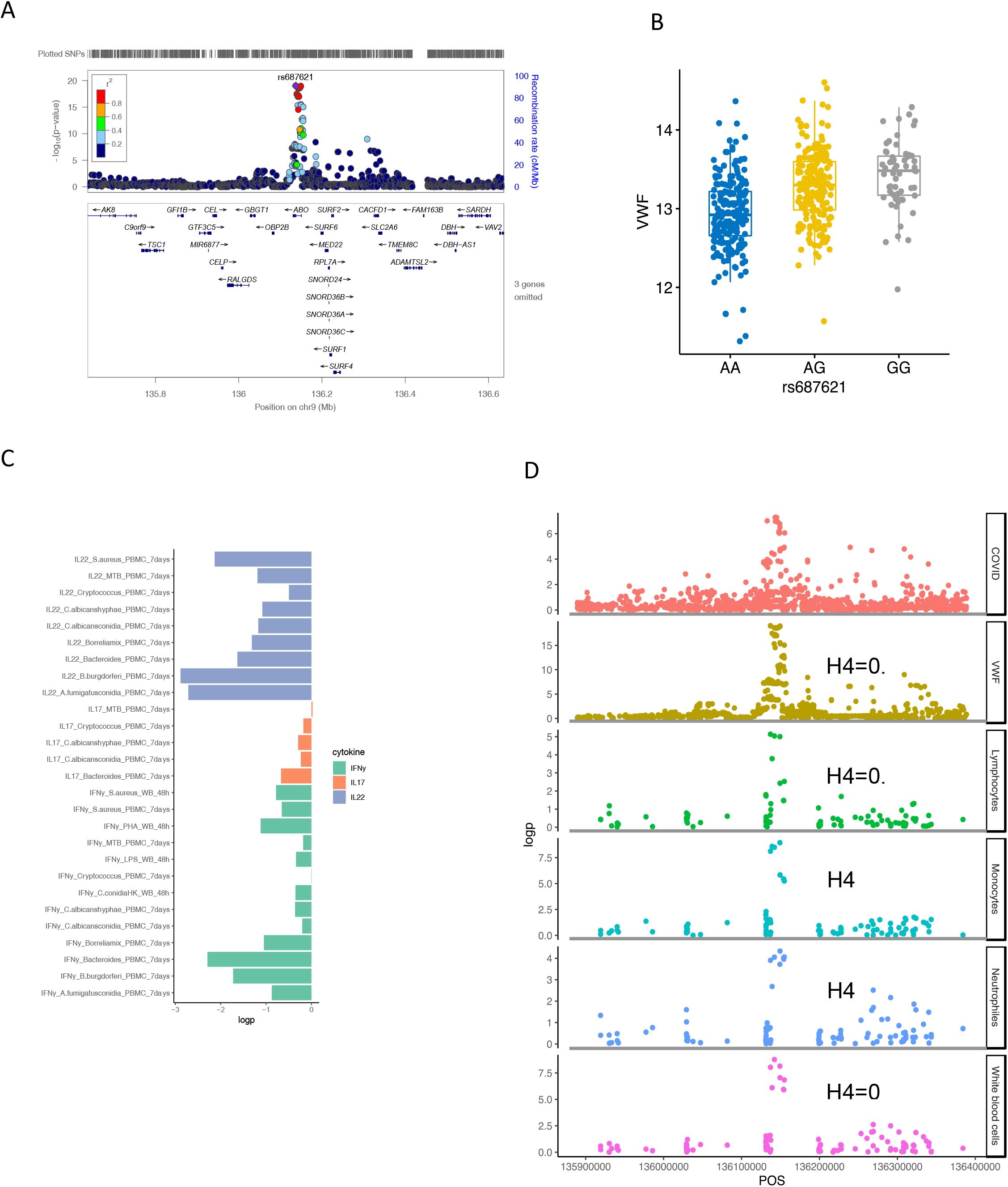
Functional annotation of ABO loci. A) locus zoom plot showing the significant association between ABO loci and VWF level. B) a boxplot showing COVID-19 risk allele(rs687621-G) associated with increasing VWF level (p-value = 9.58×10^-20^). C) a barplot showing consistent negative correlations between VWF levels and T cell-derived cytokines D) scatter plots showing colocalization between ABO loci with VWF, lymphocytes, monocytes, neutrophils and white blood cell counts.

We next tested if this specific locus is associated with immune functions. Interestingly, we observed consistent negative correlation of VWF and T-cell derived cytokine production in response to various *ex-vivo* stimulations (Fig. 3C and Fig. S3). In addition, the ABO locus led by the variant rs687621 also showed statistically significant co-localization with several immune-mediated traits, including cell counts of lymphocytes (Coloc analysis H4: 0.98), monocytes (Coloc analysis H4: 1), neutrophils (Coloc analysis H4: 0.8), and whole blood cells (Coloc analysis H4: 1) (Fig. 3D).

### Association between genetic risk score of COVID-19 severity with gender and BMI

Polygenic risk scores (PRS) combine multiple risk alleles and capture an individual’s load of common genetic variants associated with a disease phenotype (*29*). Using the summary statistics provided in the GWAS study (*1*), we calculated the PRS for the samples from 500FG and 300BCG cohorts. Since higher mortality of COVID-19 has been reported to be associated with male sex and BMI (*11, 12*), we investigated whether these host factors are associated with the PRS, a predictive measure of risk for development of severe COVID-19.

We firstly assessed if males have a higher genetic risk compared to females in 500FG. Hereby, we defined people with top 10% PRS as a high-risk group and those with bottom 10% PRS as a low-risk group. As shown in Fig. 4A, male tend to have higher severe COVID-19 risk than female (odd ratio: 1.47, 95% CI: 0.98-2.22, p-value = 0.045 (Fisher’s exact test)) (Table S9). We next used different percentile cut-offs (15%, 20%, 25% and 30%) to re-define low and high-risk groups. Interestingly, we observed a consistent pattern that males have higher genetic risk (PRS) than females at different percentile cut-offs. These results can be replicated in a similar, but independent, cohort (300BCG cohort, Fig. 4B). Meanwhile, the genetic risk difference between male and female can be attenuated when a loose cut-off has been defined. The meta-analysis of two cohorts showed a significant p-value at various percentile cut-offs (10%, 15%, 20%, 25%) and marginal significant p-value of 0.051 at the percentile cut-off of 30%. Furthermore, this result persisted when PRS was computed using summary statistics from the GWAS model after age and sex correction, reported in the original GWAS study (Table S10). However, when excluding variants from the X and Y chromosomes in the PRS calculation, the enrichment of men in the higher PRS group was not significant anymore (Fig. 4C and 4D). This suggested that higher genetic severity risk at least partially originates from the SNPs in the sex chromosomes.

**Fig. 4.**
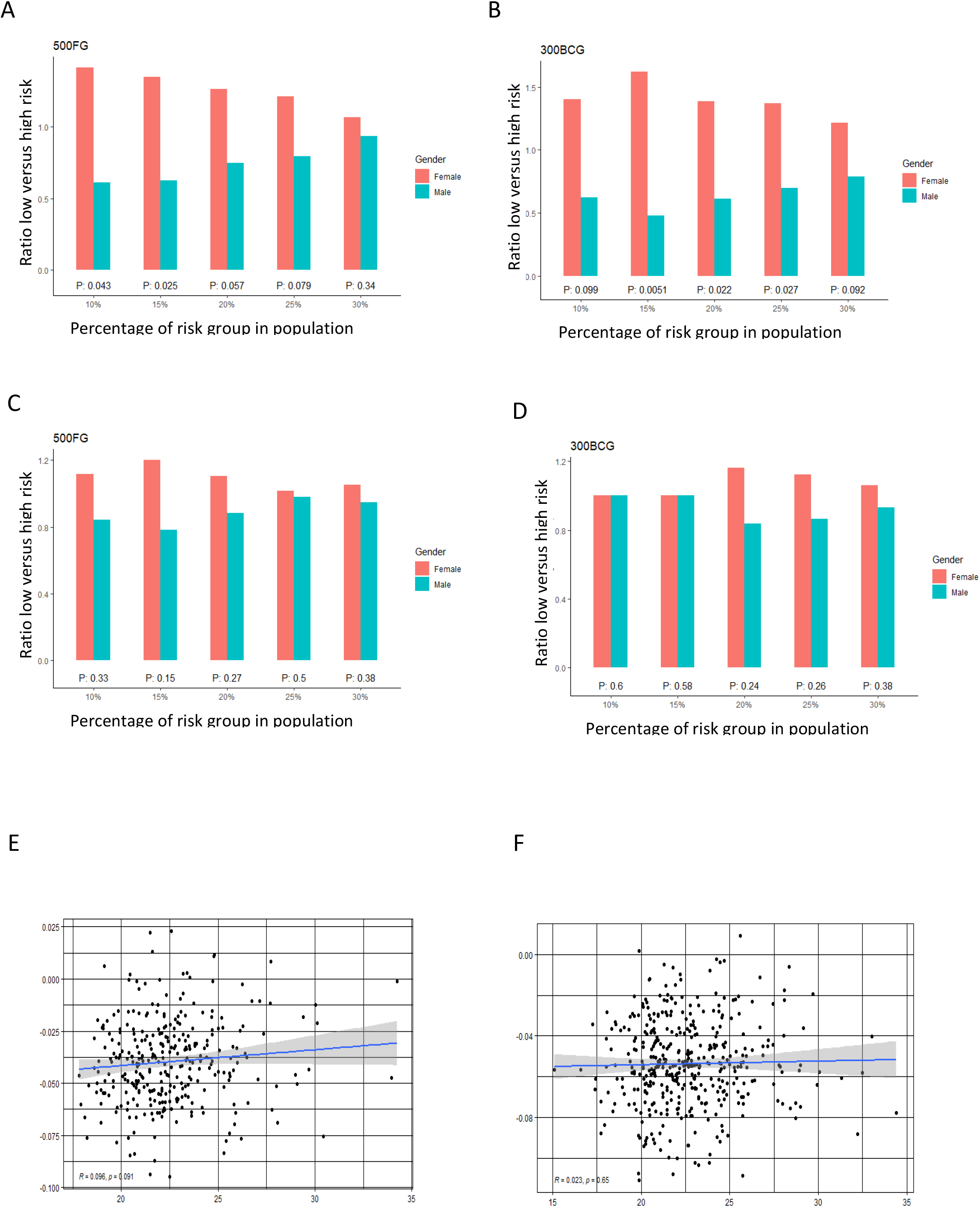
Correlation of COVID-19 PRS with gender and BMI. A) bar plot representing the ratio of low risk versus high risk in 500FG. The X-axis shows the range of different quantiles (e.g.,10% corresponds to those individuals with PRS between 0^th^ and 10^th^ percentile of the population), and the Y-axis shows the odds ratio when comparing low PRS risk and high PRS risk in the male and female group from different quantiles. B) bar plot representing the ratio of low risk versus high risk in 300BCG. C) Bar plot representing the ratio of low versus high PRS based risk between men and women in 500FG calculated without including the sex chromosomes. D). Bar plot representing the ratio of low versus high PRS based risk between men and women in 300BCG calculated without including the sex chromosomes. E). Scatter plot showing the correlation between PRS with BMI in 500FG F) in Scatter plot showing the correlation between PRS with BMI in 300BCG.

As obesity or overweight has been reported as a risk factor for serious illness or death from COVID-19, we tested if the PRS is associated with BMI (Fig. 4E and 4F). We did not observe any significant correlation between PRS and BMI.

### Association between severity risk factors and cytokine response was replicated in a second GWAS study

To investigate whether our findings are only specific to one cohort, we implemented the same functional analyses in the recent published GenOMICC GWAS study(*30*). We firstly run FUMA analyses with the same parameter settings, and the 135 mapped genes show significant enrichment in expression in spleen, blood, lung and small intestine (Fig. S4), which is consistent with our interpretation of the function of the genetic loci on immune organs. Next, we sought to test whether COVID-19 GWAS SNPs identified in GenOMICC studies influence cytokine production upon stimulation. We again observed risk alleles from 3p21.31 loci are consistently associated with lower production of monocyte-dependent cytokines (IL-6, IL-1β and TNF-α) upon various *in-vitro* stimulations (Fig. 5A and Table S11). Table S11 listed 816 normally significantly stimulation pairs SNP- cytokine (p value <0.05). Notably, 777 out of 816 SNP- cytokine stimulation pairs are from 3p21.31 loci. As 3p21.31 loci is the major genetic risk factor explaining severity outcome(https://www.covid19hg.org), our result may indicate a potential mechanism of major genetic factor impacting the severity of COVID-19 through an innate immune response.

**Fig. 5.**
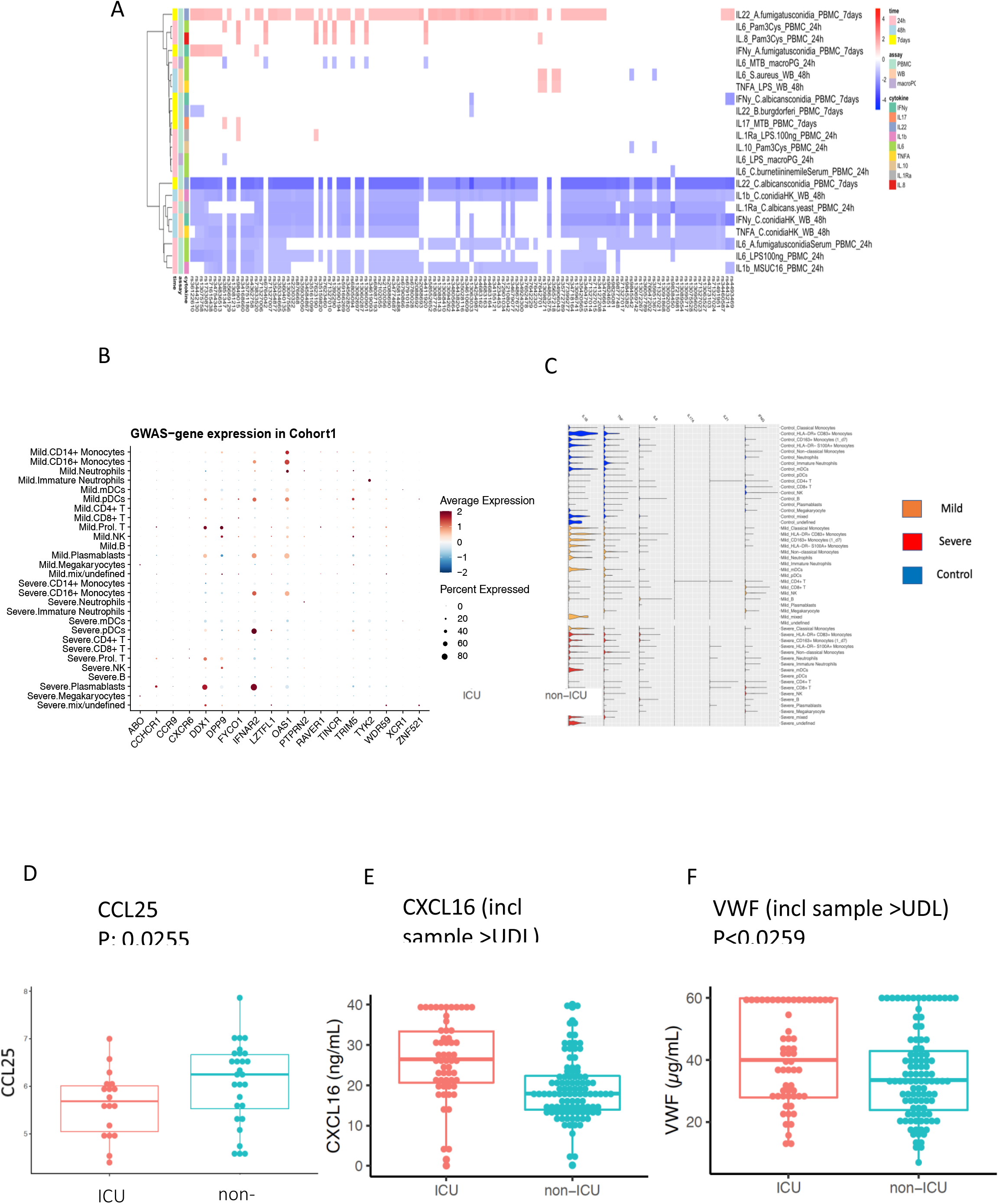
Replication and validation. A) Heatmap showing the association between 3p21.31 loci and immune traits can be replicated in an independent cohort. The red color corresponds to higher cytokine production leaded by risk allele in COVID-19 GWAS profiles, whereas blue color indicates lower cytokine production leaded by risk allele in COVID-19 GWAS profiles. B) Dot plots of expression of GWAS genes in single-cell transcriptomics of COVID-19 patients. The GWAS genes were selected from the Severe COVID-19 GWAS(*1*) and GenOMICC study(*30*). C) Violin plots of the expression of monocyte-derived cytokine genes and T cell derived cytokine genes in COVID-19 Berlin cohort based on single cell RNA-seq data. D) A boxplot of the differential protein levels of CCL25 between 18 ICU and 28 non- ICU COVID-19 patients from another independent cohort. E) A boxplot of the differential protein levels of CCL25 between 57 ICU and 102 non-ICU COVID-19 patients. F) A boxplot of the differential expression of VWF between 57 ICU and 102 non-ICU COVID-19 patients.

### Validation in single-cell transcriptomes and proteomics data of COVID-19 cohorts

To investigate in which cells those COVID-19 GWAS genes expressed, we utilized single-cell transcriptomics data from a COVID-19 German Berlin cohort(*31*) to illustrate the cell type-specific expression of COVID-19 risk genes (Fig. 5B). Among 37 COVID-19 risk genes, 10 were significantly differential expressed between severe and mild COVID-19. In general, the people with mild symptoms show high expression of those risk genes that those with severe symptoms (p-value χ2test =6.9×10^-13^) (Fig.S5). Interestingly, *OAS1* (2’-5’-Oligoadenylate Synthetase 1) shows higher expression in mild symptoms patients, which agrees with the recent findings that higher plasma OAS1 levels were associated with reduced COVID-19 severity(*32*) . We next tested the expression of monocyte-derived and T cell-derived cytokine genes in single-cell transcriptomics data. We found that monocyte-derived cytokine genes are expressed in patients, while T cell-derived cytokine genes are not, which indicates that innate immune response is of importance in disease development (Fig.5C). Therefore, we further tested the longitudinal expression change of *IL-1β* and *TNF*-*α* using the data from the same cohort (Fig. S6). Notedly, we found that patients with mild symptoms usually show a decreasing pattern of expression in cytokines (*IL-1β*, *TNF*-*α*) during the disease course (from high to low), while the severe symptom patients show an increasing pattern of expression in cytokine genes (*IL-1β*, *TNF*-*α*) with time (from low to high). Such a pattern may suggest that mild patients have a sufficiently good response in the first phase of the infection, which inhibits efficiently the viral replication, subsequently resulting in decreased inflammation. On the contrary, severe patients show defective activation of antiviral innate immunity in the beginning, leading to prolongation of the disease process with inefficient and deleterious inflammatory response in the end. This observation is in line with the recent findings of immunosuppression in severely affected patients(*33*).

We next validate whether two inflammatory proteins CCL25 and CXCL16, which are ligands for Chromosome 3 GWAS gene *CCR9* and *CXCR6*, respectively, are associated with disease severity. We measured CCL25 and CXCL16 protein levels in two different patient cohorts by using Olink platform technology and ELISA, respectively (Methods). We observed a significantly lower expression of CCL25 in ICU patients (P-value = 0.0255) (Fig. 5D), while a significantly higher expression of CXCL16 in ICU patient (P-value <0.0001) (Fig. 5E and Fig. S7). These results are in line with the reported protective effect of *CXCR6* and the risk effect of *CCR9* in transcriptomic regulation (*34*). Interestingly, the longitudinal data from COVID-19 patients showed that a clinical improvement is often associated with increased CCL25 concentration (Fig. S8).

Additionally, we measured VWF levels in 159 COVID-19 patients (Methods) and found that VWF levels were significantly higher in the ICU patients (Fig.5F and Fig.S9). This further supported our findings that the ABO genetic risk is associated with VWF levels.

## Discussion

Understanding the pathophysiology of COVID-19 is urgently needed for designing novel preventive and therapeutic approaches against the disease. One important tool for identifying the most important mechanisms mediating severity of a disease process is genomics: genetic variants that influence susceptibility or severity to a disease are usually located in genetic loci that impact important mechanisms for that particular disease. Using the information of a recently published GWAS assessing the severity of COVID-19 (*1*), and the rich datasets available in the HFGP, we interrogated the mechanisms through which genetic variants associated with severe COVID-19 exert their effects.

Among the genetic loci associated with severe COVID-19, the 3p21.31 gene cluster has been well replicated by independent studies from the COVID-19 Host Genetics Initiative (https://www.covid19hg.org), and it was reported to be inherited by Neanderthals (*35*). This locus is currently regarded as a marker of COVID-19 severity, but crucial information is missing: are the risk alleles in this locus (that encode several cytokines and chemokines) associated with a lower or higher cytokine production? The answer to this question is crucial for understanding COVID-19: a genetic risk associated with low cytokine production would imply that severe COVID-19 is the consequence of a relative immunodeficiency, while a high cytokine production associated with genetic risk would mean that severe COVID-19 is a genetic hyperinflammatory disease. In our study, the 3p21.31 genetic polymorphisms associated with a high risk of severe COVID-19 were associated with lower production of monocyte-derived cytokines, especially to viral (influenza) stimuli. This important discovery has significant prophylactic and therapeutic consequences. On the one hand, it implies that improvement of innate immune responses in healthy individuals would decrease the probability that they undergo a severe form of COVID-19: this supports the rationale of clinical trials that improve innate immune responses through induction of trained innate immunity (*7*). On the other hand, this also implies that the dysregulated immune responses that have been described at late time points in patients with severe COVID-19 (*31*)(*36*) are likely the consequence of accelerated viral multiplication due to defective innate immune responses, and subsequent systemic inflammation due to high viral loads.

Several studies have shown that ABO blood types are associated with COVID-19 severity (*37*)(*38*) and susceptibility (*39*)(*40*)(*41*). It is still not well-known how the ABO locus regulates COVID-19 susceptibility. As ABO blood group are also expressed on endothelial cells and platelets, it has been speculated that this effect may manifest itself via elevated plasma VWF (*42*). Our results provide evidence supporting this hypothesis, by showing that the risk alleles in the ABO locus are associated with high concentrations of VWF. Moreover, interesting associations have been found between polymorphisms in this locus and the number of various immune cell populations, especially lymphocytes, since lymphopenia is also consistently associated with severe COVID-19 (*43*). This suggests that genetic factors are relevant to the host thrombo-inflammatory response. However, a note of caution should be mentioned, as the association between the genetics of ABO group with severity in COVID-19 Host Genetics Initiative data did not reach a genome-wide level of significance (p value <5×10^-8^) (Table S12) (as of 21^st^ of October 2020), and thus the association might be population specific.

Another observation is that the impact of genetic polymorphisms on the severity of COVID- 19 is likely mediated through sex chromosomes, i.e. chromosome X, which is known to encode many genes related to the immune system. Indeed, men in both 500FG and 300BCG cohorts had a higher genetic risk than women, and this difference was largely lost when sex chromosomes were excluded from the PRS analysis. This finding was not able to be replicated in GenOMICC study(*30*). (Table S13), due to fact that no significant association were reported on sex chromosomes in the other GenOMICC study. Therefore, these data suggest that at least part of the well-known increase of COVID-19 severity in men is genetically determined. The recent description by our group of rare mutations in the RNA receptor TLR7 located on chromosome X as a cause of very severe COVID-19 in young men also supports this hypothesis (*44*).

While our study sheds further light on how COVID-19 genetic risk affects the human immune system, there are several limitations of this study: firstly, due to different sets of stimuli used in measuring cytokine production to stimulations in the two healthy cohorts, we are not able to replicate all our findings of genetic associations with cytokine responses from the 500FG cohort in the 300BCG cohort. Secondly, young adults (< 30 years) and normal weight BMI are overrepresented in both healthy cohorts (500FG and 300BCG), which may lead to a biased conclusion which cannot be generalized to the whole population, especially since the severe COVID-19 cases often occur in the elderly population.

Collectively, our data demonstrate that genetic variability explains an important component of the increased susceptibility to severe COVID-19. The genetic risk for severe COVID-19 is associated with defective innate immune responses (low cytokine production), dysregulated endothelial function. These findings may contribute to the development of novel treatment and prevention strategies for severe COVID-19.

## Materials and Methods

### Study cohort

The cohorts involved in this study are from the Human Functional Genomics Project (HFGP)(*45*). 500FG consists of 451 healthy individuals of European ancestry with genotype measurement. Within this cohort, immune cell counts, cytokine production upon stimulations, platelets, globulins, and gut microbiome were measured (for detailed information see (*13, 14, 46, 47*)). 300BCG consists of 313 healthy Europeans that participated in a BCG vaccination study (*48*)(*49*). The basic characteristics of study populations are shown in Table S14. Within this cohort, blood was collected before vaccination and cytokine production was measured upon *ex-vivo* stimulation of PBMCs with microbial stimuli.

### Genotype quality control and imputation

Genotyping on samples from 500FG and 300BCG was performed using Illumina humanOmniExpress Exome-8 v1.0 SNP chip Calling by Opticall 0.7.0(*50*) with default settings. All individuals of non-European ancestry, ambiguous sex, call rate ≤ 0.99, excess of autosomal heterozygosity (F<mean-3SD), cryptic relatedness (π>0.185) were removed. SNPs with low genotyping rate (<95%), with low minor allele frequency (<0.001), deviation from Hardy-Weinberg equilibrium (p<10^-4^) were excluded. The detailed QC steps have been published in reference(*14*). Genotype data of 500FG and 300BCG were imputed respectively. The imputation was performed on the Michigan imputation server(*51*). The cohorts were phased using Eagle v2.4 with the European population of HRC 1.1 2016 hg 2019 reference panel. After imputation, variants with a MAF < 0.01, an imputation quality score R2 < 0.5, or a Hardy-Weinberg-Equilibrium P < 10^-12^ were excluded. All quality control steps were performed using Plink v1.9. After imputation and quality control, 451 individuals from 500FG and 313 individuals from 300BCG were available for downstream analyses.

### Immune parameter quantitative trait locus (QTL) profiles

We acquired summary statistics of cytokine QTLs (*14*), cell proportion QTLs(*46*) and circulating mediators and metabolite QTLs(*52*) from our previous studies performed with 500FG. Metabolites were measured on the Brainshake Metabolomics/Nightingale Health metabolic platform. These samples were processed following the automated standard protocol provided by Nightingale’s technology (Finland), and blood metabolites were quantified in absolute concentrations (e.g. mmol/L) and percentages using nuclear magnetic resonance (NMR) spectroscopy.We performed QTL mapping for circulating mediators and platelet traits in 500FG using an R package MatrixeQTL(*53*). The measurement of circulating mediators including IL-18BP, resistin, leptin, adiponectin, alpha-1 antitrypsin (AAT), and IL-18 have been described previously(*13*). Platelet traits(*54*) include Thrombin-Antithrombin Complex (TAT), Beta-thromboglobulin total, beta-thromboglobulin, fibrinogen binding, collagen-related peptide (CRP) P-selectin, CRP fibrinogen, ADP P-selectin, ADP fibrinogen, P-selectin, platelet−monocyte complex, total platelet count, and von Willebrand factor (VWF). The circulating mediator levels and platelet traits were log2 transformed. A linear model was applied to the platelet data and genetic data by taking age and sex as covariates. We considered p-value < 5×10^-8^ to be genome-wide significant.

### Colocalization analysis

We performed co-localization analysis (*55*) to look at the overlapping profile between molecular QTLs, COVID-19 GWAS, and other GWAS profiles using the R package ‘coloc’.

### PRS calculation

Polygenic risk scores (PRS) were calculated by first intersecting the variants from the COVID- 19 summary statistics(*1*) with the variants present in our samples. Clumping was done starting at the most significant variant. All variants within a 250kb window around that variant were excluded if they were in greater LD than 0.1 before continuing to the most significant variant outside of the previous window. For each sample specifically, we then multiplied the dosage of the effect allele with its effect size while substituting missing genotype data with the average dosage of that variant in the entire sample set. These values were then summed to form the PRS for each specific sample. As the GWAS summary statistics for creating PRS from Eillinghaus *et.al* (*1*) did not correct age and sex, we also performed a sensitivity analysis with the PRS created from the GWAS model corrected for age and sex.

### PRS based correlations

Linear models were constructed using the computed PRS and various phenotype data available for each cohort. Samples within the top/bottom 10% PRS were classified as high/low-risk, respectively. Using the PRS of the samples in these risk groups, we performed a Student T-test to test for significant correlation between gender and PRS. Furthermore, we tested for enrichment of any specific gender in these risk groups using a Fisher’s exact test.

### Functional analysis of genomic loci

We used the FUMA pipeline in order to identify genes linked to COVID-19 with severe respiratory failure. FUMA identified significant independent SNPs as variants with P < 1×10^-5^ that were independent from each other using an LD threshold of r2 < 0.6. Within these independent significant SNPs variants lead SNPs are identified as the most significant variants that are independent using an LD threshold of r2 < 0.1. We mapped Genes to these SNPs based on their genomic position allowing for a maximum distance of 10kb. In addition to this, genes were also mapped based on eQTL effects. Genes were selected based on significant SNP-gene pairs at FDR < 0.05 using cis- and trans-eQTLs from eQTLGen (https://www.eqtlgen.org).

As part of the FUMA pipeline we used these mapped genes in order to generate gene expression heatmaps using GTEx v8 (54 tissue types and 30 general tissue types). Gene expression values with a pseudocount of 1 were normalized across tissue types using winsorization at 50 and log2 transformed. Using the hypergeometric test, we tested for significant enrichment of our input genes in DEG sets for the different tissue types using a Bonferroni corrected P value ≤ 0.05. Finally, we tested for overrepresentation of our input genes in predetermined gene-sets using hypergeometric tests. Gene-sets were obtained from MsigDB, WikiPathways, and GWAS-catalog reported gene-sets. We used Benjamini-Hochberg FDR correction for each of the categories within these gene-set sources separately using a threshold of 0.05 for our adjusted P value.

### Roadmap epigenetic state enrichment

Based on the Roadmap 15-core epigenetic state database(*20*), we used data obtained from 23 blood samples spanning 127 epigenomes to map the QTLs in the summary statistics to their respective epigenetic states. Epigenetic state information was available for bins of 200bp. we aggregated this information into 4 categories; active enhancer states (Enh, EnhG), active promotor states (TssA, TssAFlnk), all enhancer states (Enh, EnhG, EnhBiv), and all promotor states (TssA, TssAFlnk, TssBiv). We tested for enrichment using a Fisher’s exact test based on the number of unique 200bp bins variants mapped to. This was done after filtering the QTL’s down based on their p value using different thresholds (1×10^-5^, 1×10^-6^ and 1×10^-7^). Enrichment P values were obtained after FDR correction.

### Biomarker measurements

Hospitalized patients with presumed COVID-19 disease were included in a prospective cohort between March and April 2020 at the Radboudumc (Nijmegen, the Netherlands). Disease severity was defined based on the patient’s need for intensive care at the time of sampling. The inclusion and clinical characteristics of this patient cohort has been previously described in detail(*56*). The basic characteristics of the studies samples are shown in Table S15. Plasma samples were collected from EDTA blood and stored at -80°C. The plasma concentrations of CCL25 were determined using the commercially available Inflammation panel from Olink Proteomics AB (Uppsala, Sweden). The procedure of this immunoassay was performed as previously described(*57*). CCL25 levels are expressed on a log2-scale as normalized protein expression (NPX) values and were normalized using control samples to correct for batch effects. Values under the detection threshold were replaced with the lower limit of detection. CCL25 levels were compared between severe COVID-19 patients (ICU ward = 18) and non-severe COVID-19 patients (non-ICU ward, N = 28) using a linear regression analysis with age and sex as covariates. CCL25 levels was measured every 2 days until an endpoint was reached (either patients left the hospital, or died of the disease).

Plasma concentrations of CXCL16 were measured using a commercially available enzyme-linked immunosorbent assays (ELISA, Invitrogen, Thermo Fisher Scientific) according to the manufacturer’s protocol, with a lower and upper detection limits of 0.055 and 40 ng/mL, respectively. Plasma concentrations of VWF were measured using a commercially available ELISA (Abcam, ab223864) according to the manufacturer’s protocol, with a lower and upper detection limits of 0.94 and 60 µg/mL, respectively. Values under or above the range of detection were replaced with the lower or upper detection limits, respectively. The basic characteristics of the studies samples are shown in Table S16. The student’s *t* test was used to compare protein levels between 57 ICU patient and 102 non-ICU patients.

### scRNA-seq *analysis*

Samples from patients with COVID-19 were collected during the first wave of the pandemic from Berlin between March and July 2020 in Germany. Berlin cohort consists of 25 mild and 29 severe COVID-19 patients, and 22 controls samples from publicly available scNRA-seq data. The detailed clinical characteristics of those samples have been previously described (*31*). Gene expression levels were compared between severe patients and mild or healthy controls using FindMarkers functions in *Seurat* v3.2.2 (Stuart, Cell, 2019) with Wilcoxon Rank Sum Test. Genes at least 10% expressed in tested groups and Bonferroni-corrected p-values < 0.05 were regarded as significant differentially expressed genes.

### Visualization

R package ggplot2 was used to perform bar charts, box plots and scatter plots. We applied an online tool Locus zoom to present genes within candidate loci. We used R package pheatmap to generate heat maps.

## Data Availability

500FG data used in this project have been archived in the BBMRI-NL data infrastructure (https://hfgp.bbmri.nl/). This allows flexible data querying and download, including sufficiently rich metadata and interfaces for machine processing and using FAIR principles to optimize Findability, Accessibility, Interoperability and Reusability.

## Acknowledgments

We thank all volunteers in the 500FG and 300BCG cohorts of the Human Functional Genomics Project (HFGP) for their participation.

## Funding

MGN was supported by an ERC Advanced Grant (833247) and a Spinoza Grant of the Netherlands Organization for Scientific Research. YL was supported by an ERC starting Grant (948207) and a Radboud University Medical Centre Hypatia Grant (2018). CJX was supported by Helmholtz Initiative and Networking Fund (1800167).

## Author contributions

M.G.N., C.J.X. and Y.L. designed the study, Y.K. and X.C. performed statistical analyses supervised by Y.L. and C.J.X., M.J., S.M., V.K., V.M., C.d.B., Q.d.M. and L.A.B.J. established the cohorts, helped with the data analysis and interpretation of results. B.Z., V.K., A.d. N, I. G. and N.J. performed the validation. C.J.X., M.G.N., Y.L., Y.K. and X.C. wrote the manuscript with input from all authors.

## Competing interests

The authors declare no competing interests.

## Data and materials availability

500FG data used in this project have been archived in the BBMRI-NL data infrastructure (https://hfgp.bbmri.nl/). This allows flexible data querying and download, including sufficiently rich metadata and interfaces for machine processing and using FAIR principles to optimize Findability, Accessibility, Interoperability and Reusability. scRNA-seq data generated during this study are deposited at the European Genome-phenome Archive (EGA) under access number EGAS00001004571. The processed count tables of the datasets used in the analysis were available at The FASTGenomics platform(fastgrnomics.org).

## Ethics statement

The 300BCG (NL58553.091.16) and 500FG (NL42561.091.12) studies were approved by the Arnhem-Nijmegen Medical Ethical Committee. Experiments were conducted according to the principles expressed in the Declaration of Helsinki. Samples of venous blood were drawn after informed consent was obtained.

---

**Fig. S1.**
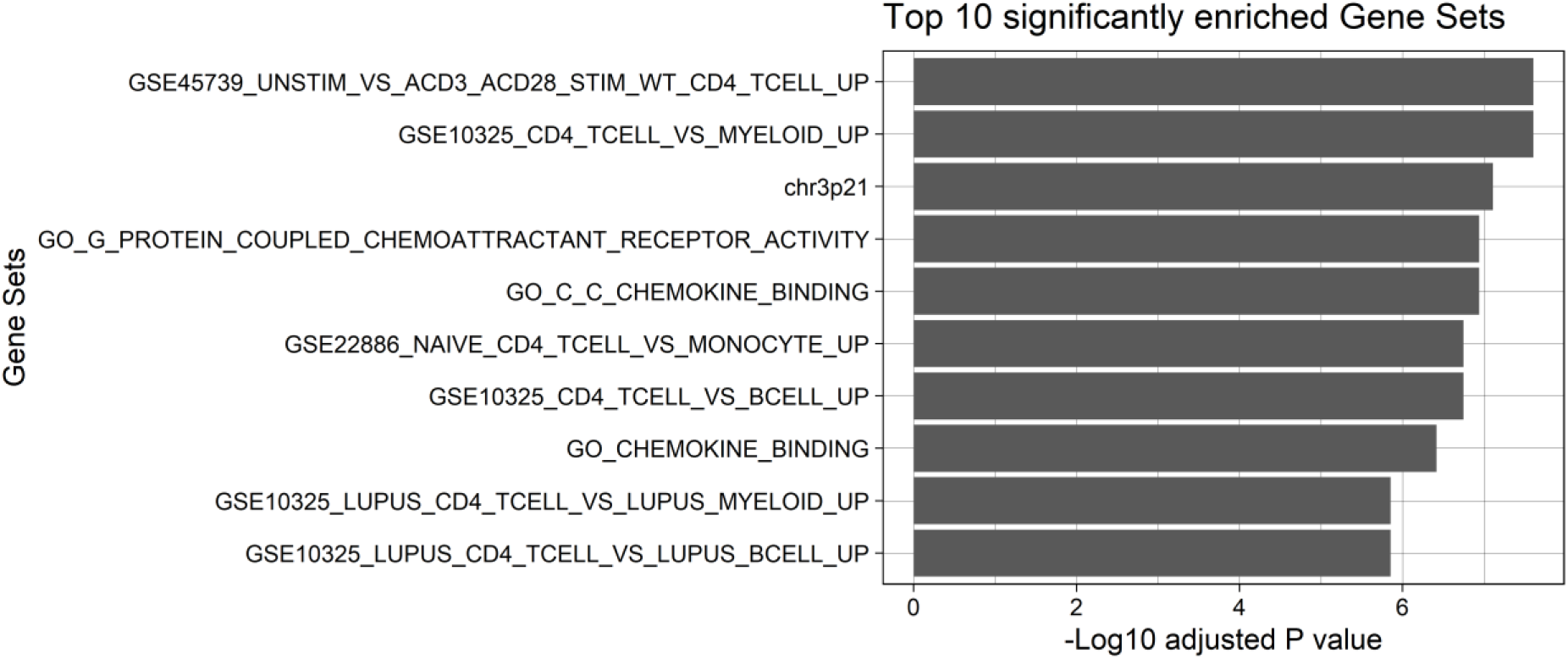
A bar plot showing the top 10 significant enriched gene sets using functional annotation of COVID-19 loci by FUMA pipeline.

**Fig. S2.**
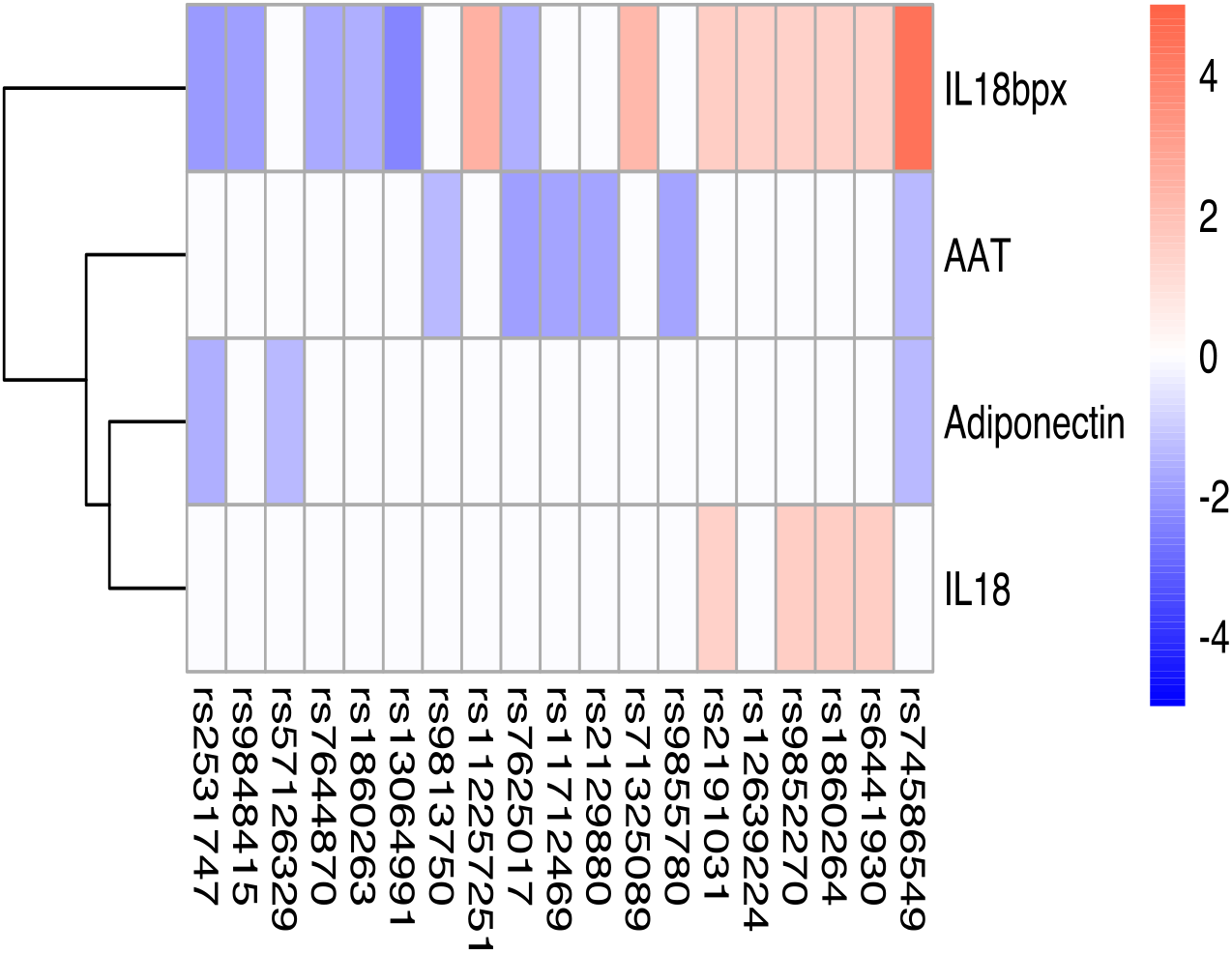
Heatmap of the genetic associations between 3p21.31 loci and circulating mediator.

**Fig. S3.**
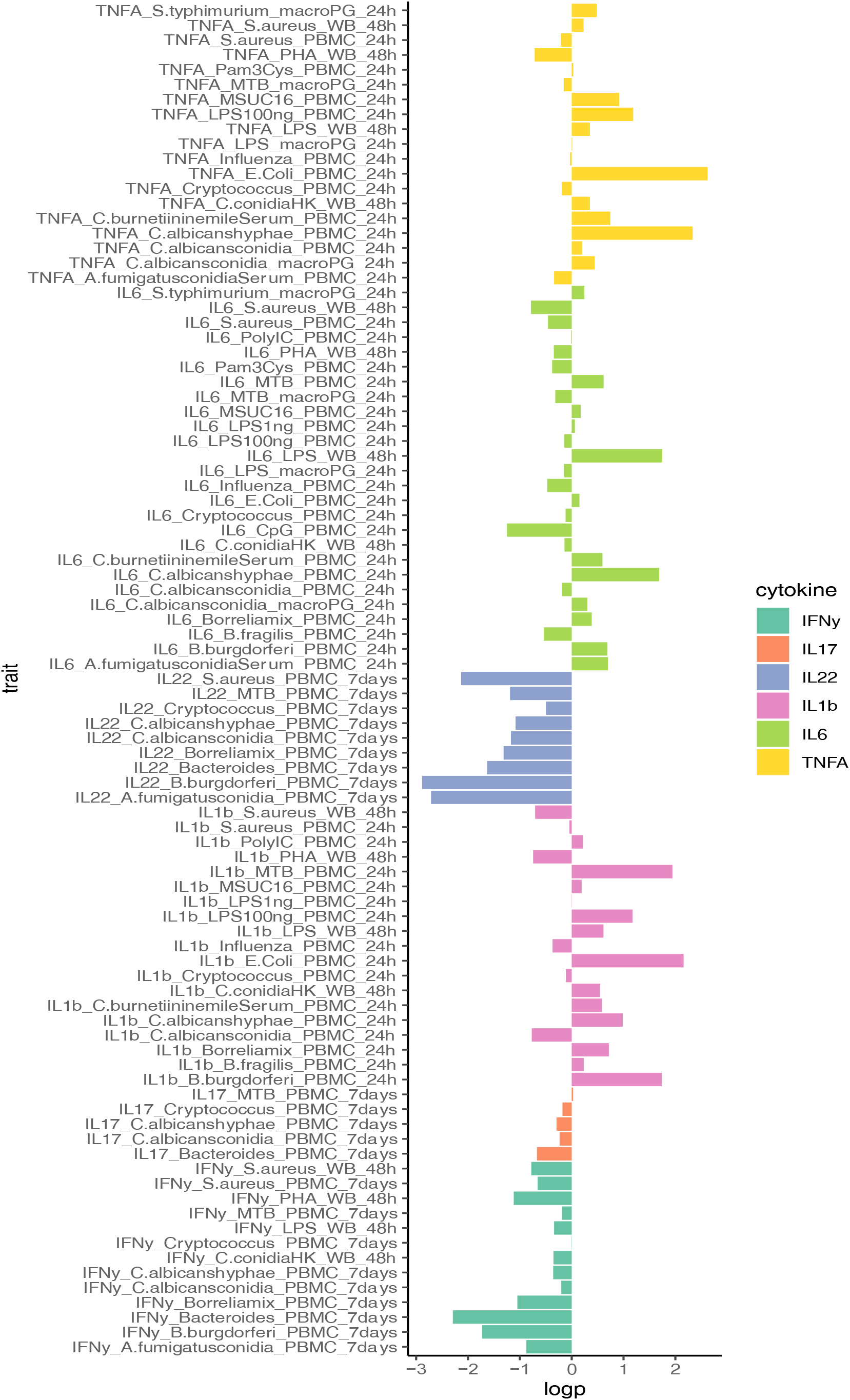
A barplot showing associations between VWF levels and cytokine

**Fig. S4.**
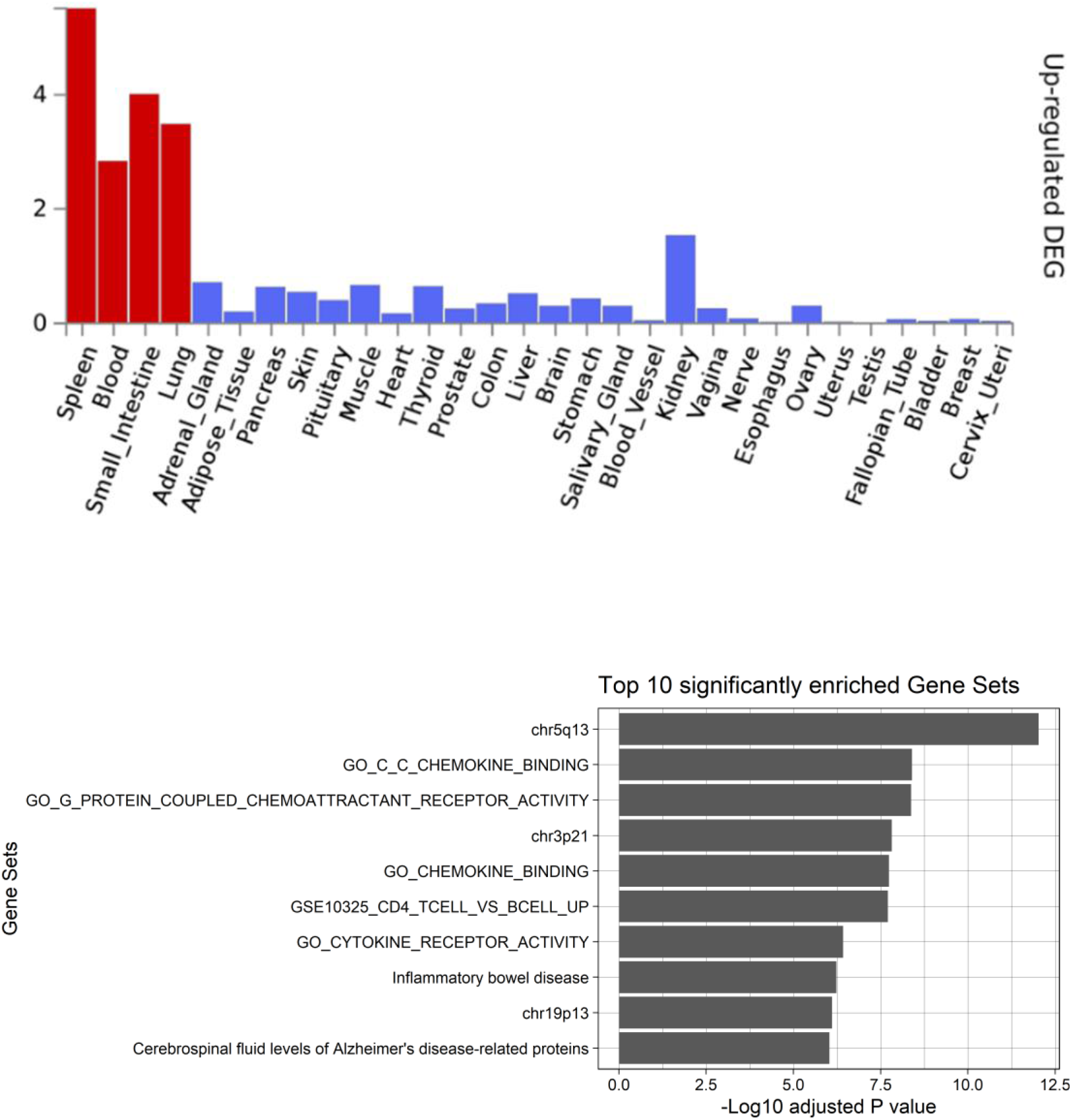
Functional annotation of COVID-19 loci from GenOMICC study of European ancestry group using the FUMA pipeline. This was done based on genes identified after using FUMA to map QTLs based on their genomic location, eQTL associations, and histone activity. A) MAGMA Tissue expression results on 30 general tissues type (GTEx v8), B) The top 10 significant enriched gene sets.

**Fig. S5.**
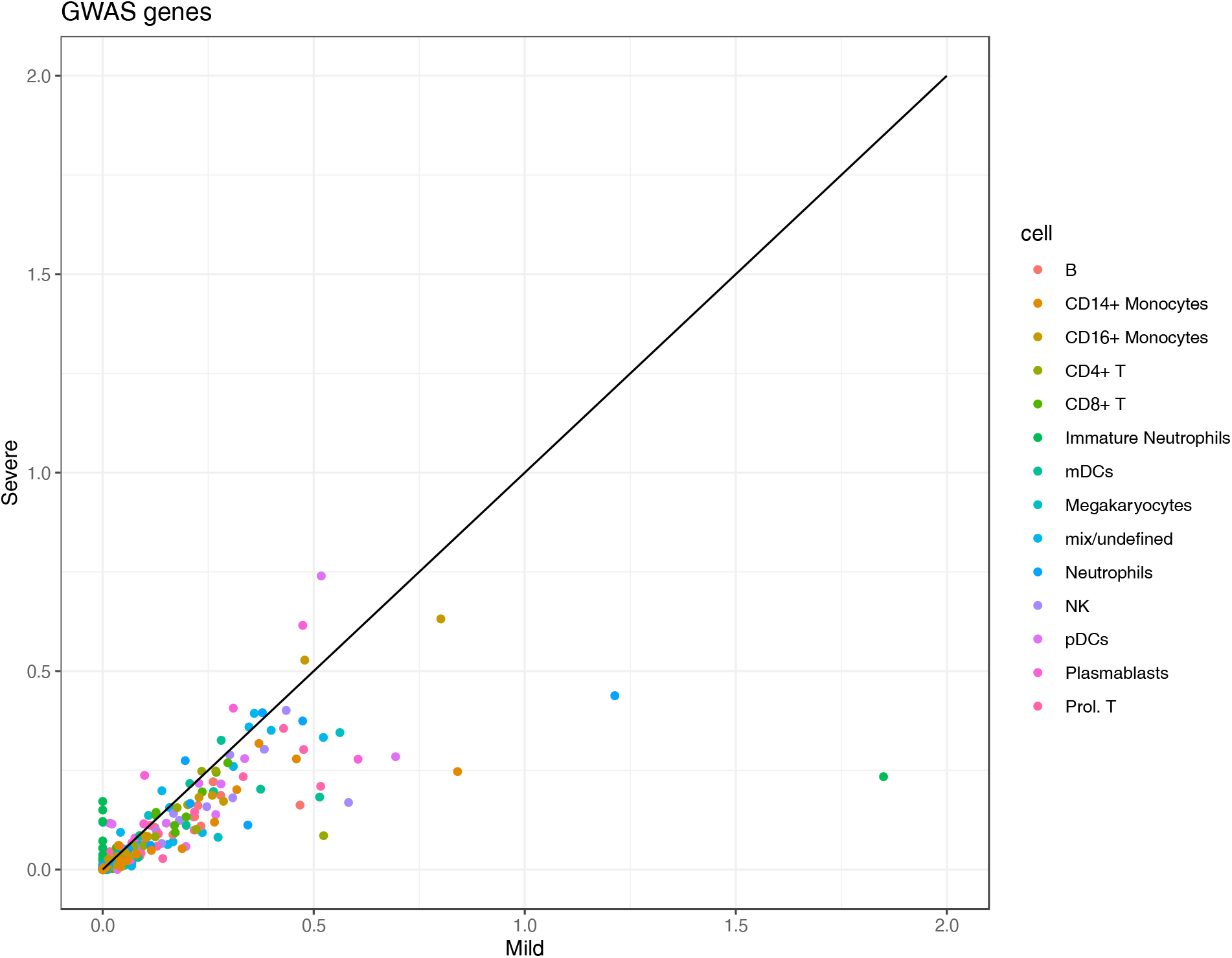
A scatter plot showing the average cell type-specific expression of GWAS genes between mild and severe COVID-19 patients.

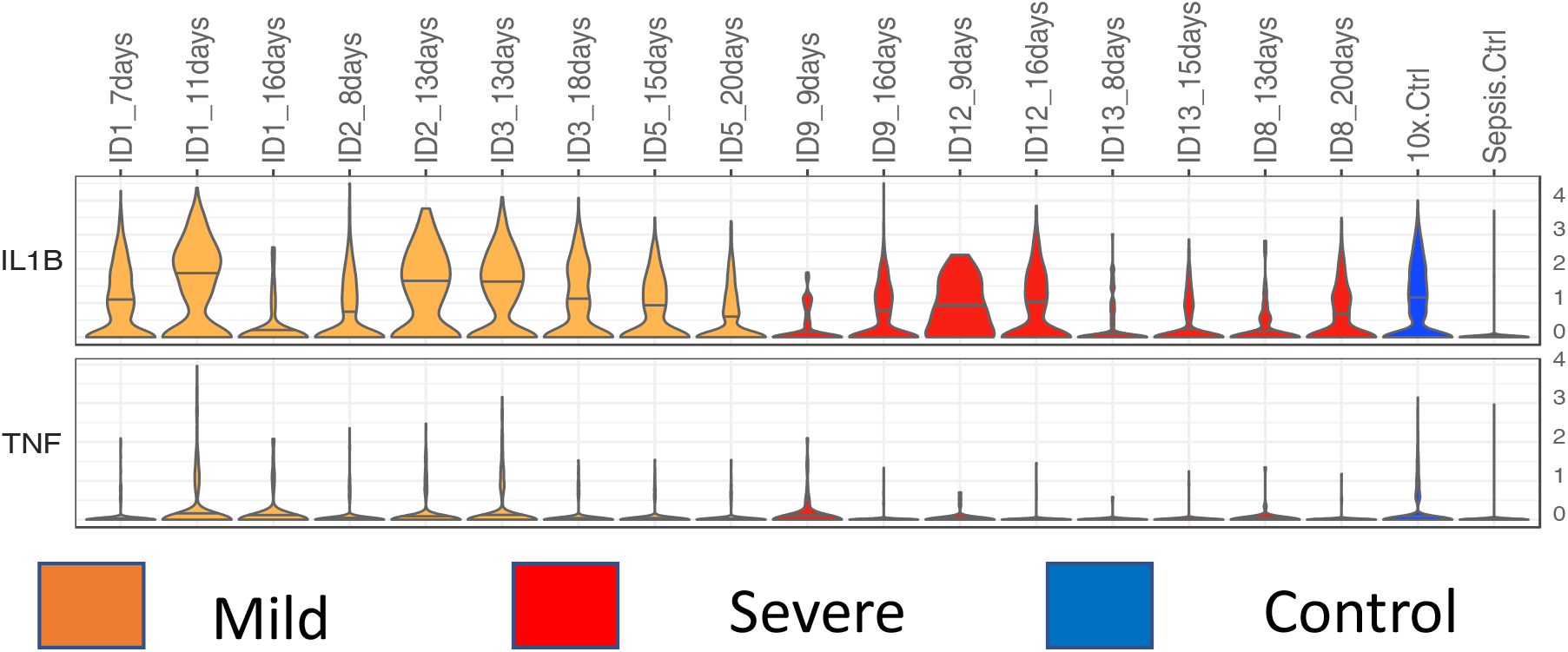
The longitudinal change of *IL-1 β* and *TNF-α* expression (violin-plots) in monocytes of COVID-19 Berlin cohort.

**Fig. S7.**
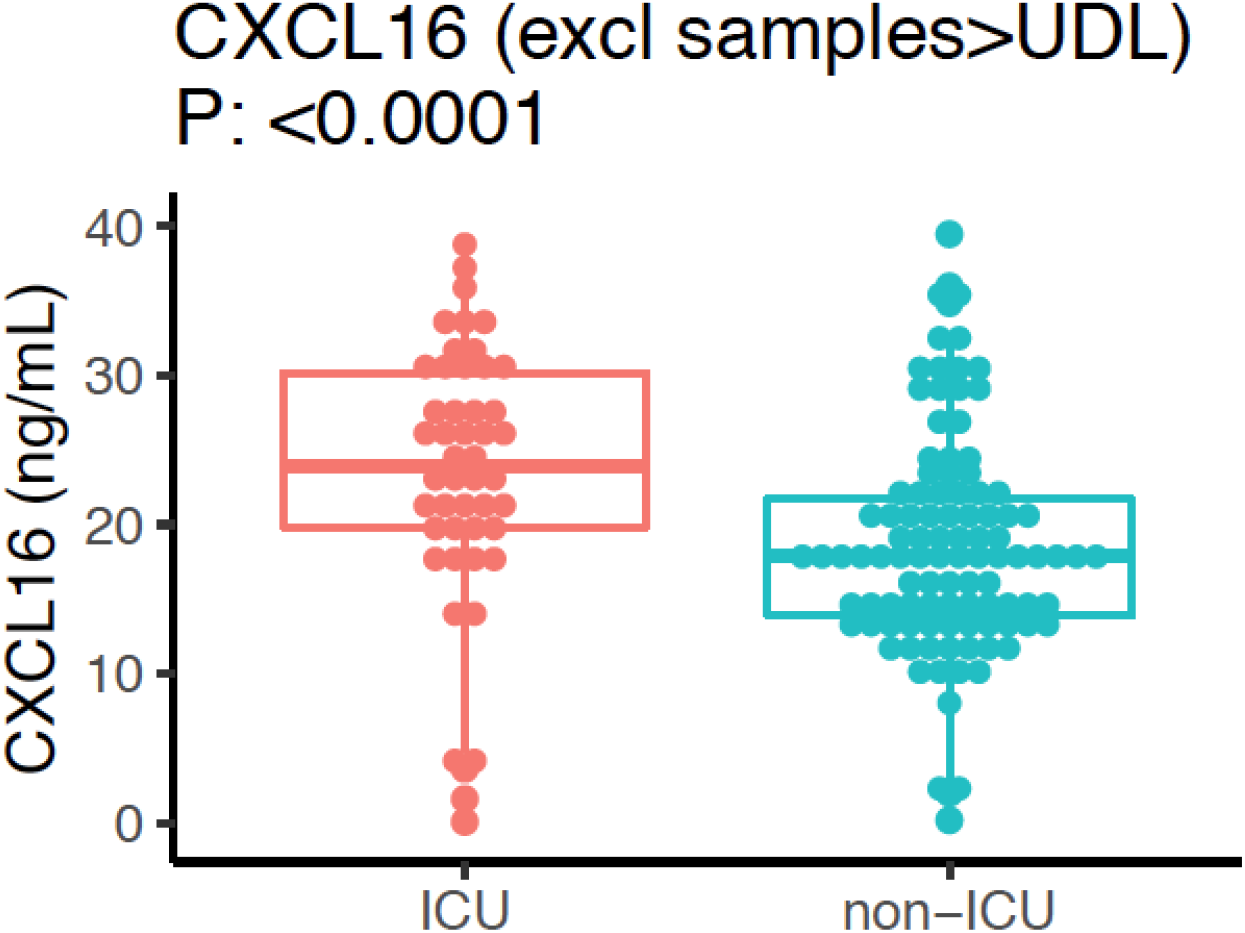
A boxplot of the differential expression of CXCL16 between ICU and non-ICU COVID-19 patients after excluding samples which are above upper detection limit (UDL).

**Fig. S8.**
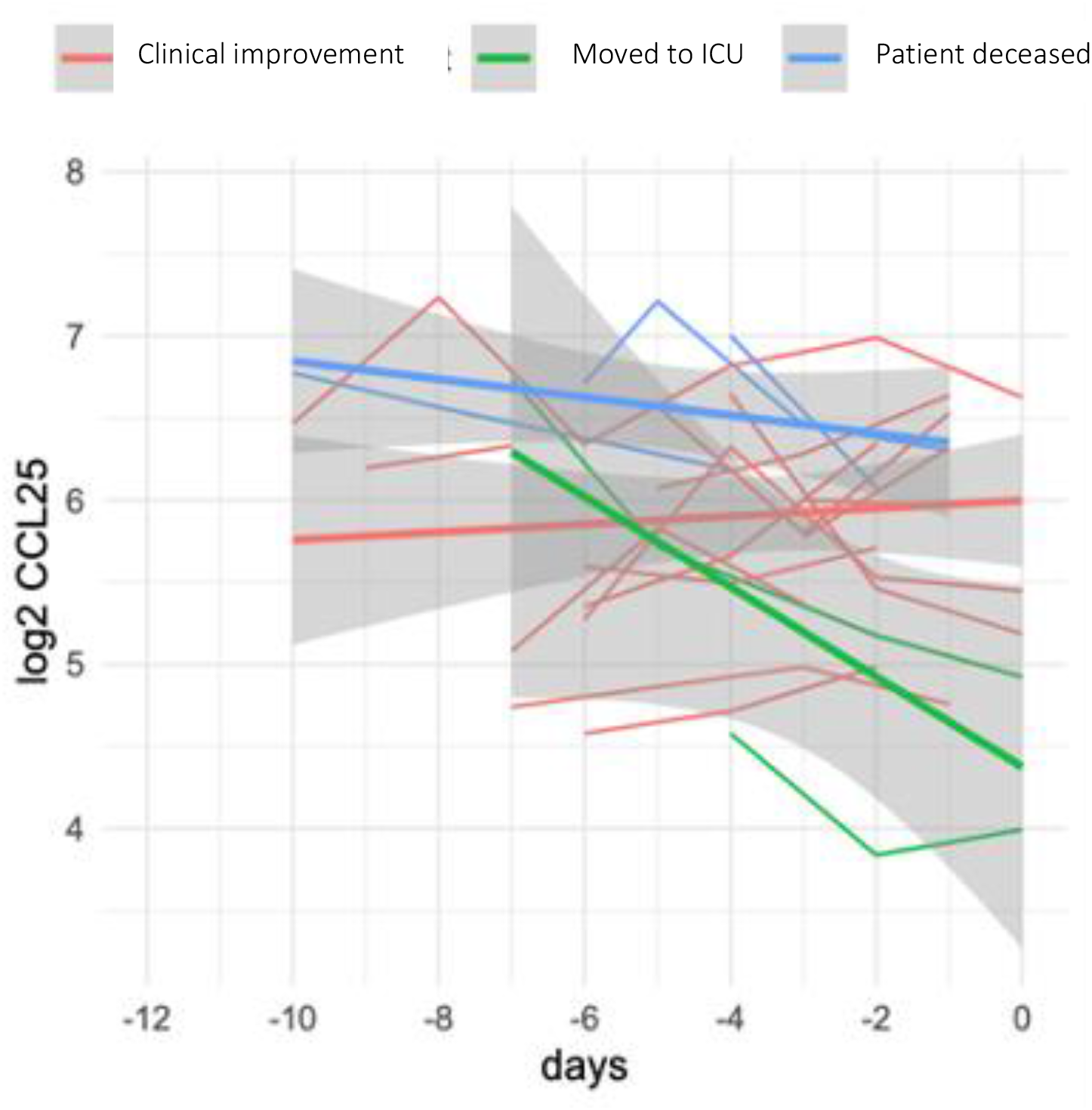
The expression of CCL25 at different time points in three clinical groups (clinical improvement, moved to ICU and patient deceased).

**Fig. S9.**
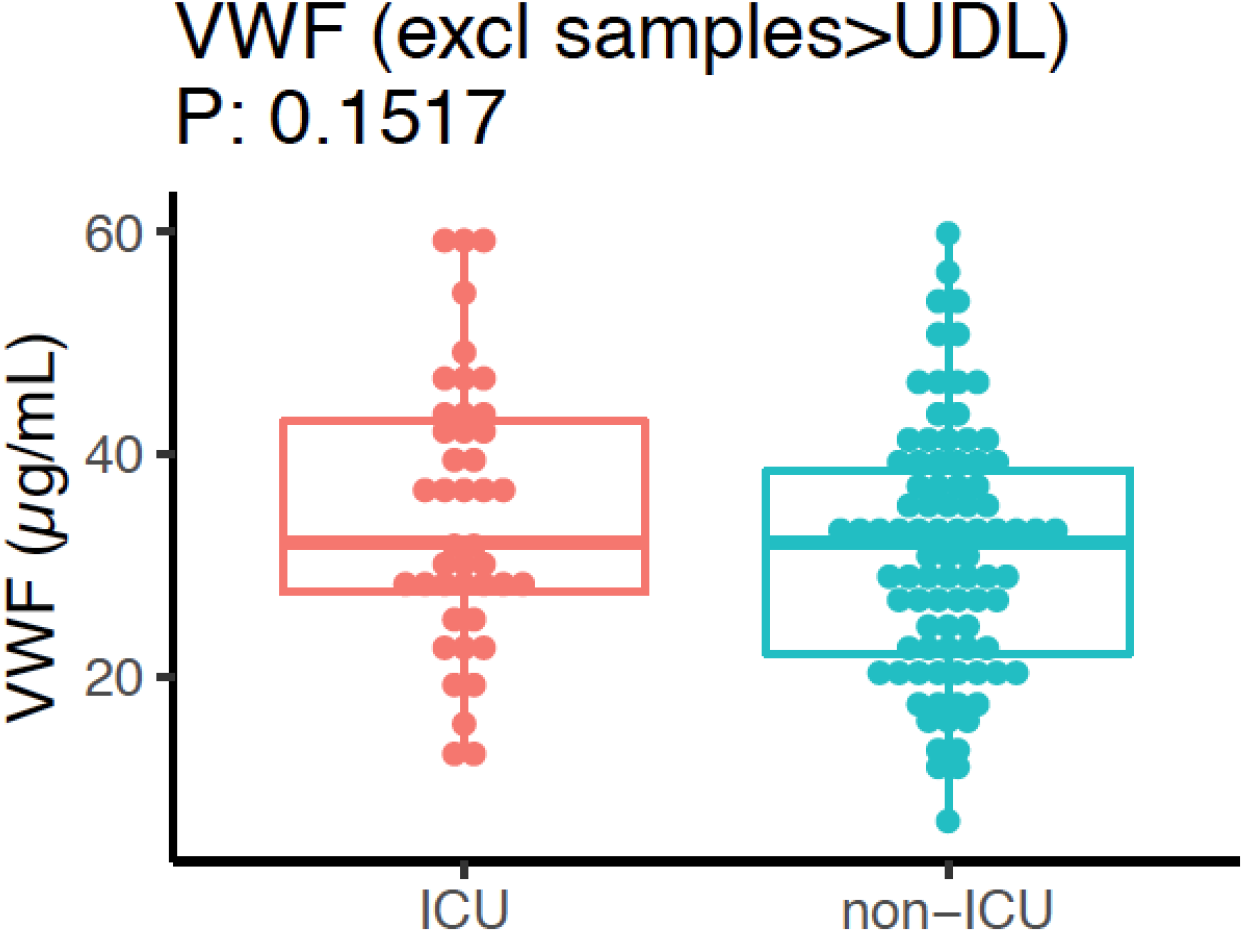
A boxplot of the differential expression of VWF between ICU and non-ICU COVID- 19 patients after excluding samples which are above upper detection limit (UDL).

## Notes

### Competing Interest Statement

The authors have declared no competing interest.

### Author Declarations

The 300BCG (NL58553.091.16) and 500FG (NL42561.091.12) studies were approved by the Arnhem-Nijmegen Medical Ethical Committee. Experiments were conducted according to the principles expressed in the Declaration of Helsinki. Samples of venous blood were drawn after informed consent was obtained. The COVID-19 Single cell RNA sequencing study is approved by the Institutional Review board of Charite (EA2/066/20). Written informed consent was provided by all patients or legal representatives for participation in the study. This COVID-19 protemics study was performed according the latest version of the declaration of Helskini and guidelines for good clinical practice. The Arnhem-Nijmegen Medical Ethical Committee approved the study protocol (CMO 2020-6344 and CMO 2016-2923).

### Summary of Updates

During the revision, we have added the following validation data to further strengthen our conclusions: 1) By using single-cell transcriptome data from healthy, mild and severe COVID-19 patients (n=76), we demonstrated that monocyte-derived cytokine genes are highly expressed in patients, while T cell-derived cytokine genes are not. This is in line with our original findings that innate immune response is of importance in COVID-19 development. 2) To better understand the function of GWAS CXCR6/CCR9 locus on Chr 3, we have further measured their ligands, i.e. CXCL16 in 159 ICU and non-ICU COVID-19 patients using ELISA and CCL25 in 46 patients using OLink platform technology. Our new data revealed that CCL25 and CXCL16 are differentially in the ICU and non-ICU patients, suggesting a protective effect of CXCR6, and the detrimental effect of CCR9. 3) To validate our finding on the colocalization of COVID-19 GWAS and vWF levels on chr9:ABO locus, we have measured the plasma concentrations of vWF in 159 COVID-19 ICU and non-ICU patients. Interestingly, the ICU patients had significantly higher levels of vWF levels than non-ICU ones, which confirmed our genetic colocalization results. 4) In the new version of the manuscript, we have made use of data from the recent GWAS COVID-19 study (Pairo-Castineira et.al Nature, 2021) and we were able to replicate our findings and conclusions which were based on Ellinghaus et.al NEJM 2020.

