## supplemental table for "The genetic risk for COVID-19 severity is associated with defective innate immune responses"

**Tabel S1 the association of COVID-19 risk loci with phenotypes in GWAS catalog (last accessed on 15-July-2020)**

| SNP | P | P-value annota | RAF | OR | Beta | CI | Mapped gene | Reported trait | Trait(s) | Study accession | Location | P-value_in_COVID_GWAS |
|---|---|---|---|---|---|---|---|---|---|---|---|---|
| rs505922-C | 4 x 10-15 | | 0.35 | 1.81 | '- | [1.56-2.11] | ABO | Venous thromboembolism | venous thromboembolism | GCST000354 | 9:133273813 | 3.21E-07 |
| rs505922-C | 5 x 10-8 | | 0.35 | 1.2 | '- | [1.12-1.28] | ABO | Pancreatic cancer | pancreatic carcinoma | GCST000456 | 9:133273813 | 3.21E-07 |
| rs8176719-G | 4 x 10-21 | | 0.37 | 1.48 | '- | [NR] | ABO | Malaria | malaria | GCST001637 | 9:133257522 | 9.93E-08 |
| rs505922-T | 5 x 10-57 | (vWF) | 0.68 | '- | 0.561 unit | [0.49-0.63] | ABO | End-stage coagulation | von Willebrand factor measurement | GCST001798 | 9:133273813 | 3.21E-07 |
| rs505922-? | 7 x 10-40 | (TNFA) | 0.34 | '- | '- | '- | ABO | Protein quantitative trait loci | tumor necrosis factor-alpha measurement | GCST000189 | 9:133273813 | 3.21E-07 |
| rs507666-G | 5 x 10-29 | | 0.20 | '- | 17.73 umc | [NR] | ABO | Soluble ICAM-1 | ICAM-1 measurement | GCST000210 | 9:133273983 | 2.24E-05 |
| rs505922-? | 2 x 10-10 | | 0.53 | 1.14 | '- | [1.1-1.19] | ABO | Graves' disease | Graves disease | GCST001982 | 9:133273813 | 3.21E-07 |
| rs8176719-G | 6 x 10-12 | | 0.419 | 1.47 | '- | [1.32-1.64] | ABO | Venous thromboembolism | venous thromboembolism | GCST001557 | 9:133257522 | 9.93E-08 |
| rs2519093-A | 8 x 10-16 | | 0.243 | 1.69 | '- | [1.48-1.91] | ABO | Venous thromboembolism | venous thromboembolism | GCST001557 | 9:133266456 | 4.18E-05 |
| rs507666-A | 3 x 10-91 | | 0.20 | '- | 17.3 ng/m | [NR] | ABO | Soluble ICAM-1 | ICAM-1 measurement | GCST001047 | 9:133273983 | 2.24E-05 |
| rs505922-T | 1 x 10-10 | (Recessive) | 0.54 | 1.32 | '- | [NR] | ABO | Duodenal ulcer | duodenal ulcer | GCST001433 | 9:133273813 | 3.21E-07 |
| rs505922-T | 8 x 10-6 | | 0.53 | 1.13 | '- | [1.07-1.20] | ABO | Graves' disease | Graves disease | GCST001200 | 9:133273813 | 3.21E-07 |
| rs612169-G | 9 x 10-40 | (SM-8 + 3 otl | 0.335 | '- | 0.202 unit | [NR] | ABO | Metabolic traits | metabolite measurement | GCST001217 | 9:133268030 | 1.08E-07 |
| rs505922-C | 1 x 10-34 | | 0.43 | 1.92 | '- | [NR] | ABO | Venous thromboembolism | venous thromboembolism | GCST001253 | 9:133273813 | 3.21E-07 |
| rs8176685-G | 7 x 10-10 | | 0.1811 | '- | 0.0288057 | [0.02-0.038] | ABO | Platelet distribution width | platelet component distribution width | GCST004616 | 9:133263363 | 3.34E-05 |
| rs505922-C | 5 x 10-8 | | NR | 1.06 | '- | [1.04-1.09] | ABO | Type 2 diabetes | type II diabetes mellitus | GCST005414 | 9:133273813 | 3.21E-07 |
| rs507666-? | 8 x 10-118 | (EA, E-Selec | NR | '- | '- | '- | ABO | Blood protein levels | e-selectin measurement | GCST004365 | 9:133273983 | 2.24E-05 |
| rs2519093-C | 2 x 10-48 | (EA) | 0.806 | '- | 0.087 unit | '- | ABO | Low density lipoprotein chol | low density lipoprotein cholesterol measurement | GCST007141 | 9:133266456 | 4.18E-05 |
| rs2519093-C | 1 x 10-14 | (Hispanic) | 0.831 | '- | 0.155 unit | '- | ABO | Low density lipoprotein chol | low density lipoprotein cholesterol measurement | GCST007141 | 9:133266456 | 4.18E-05 |
| rs2519093-C | 1 x 10-6 | (East Asian) | 0.797 | '- | 0.098 unit | '- | ABO | Low density lipoprotein chol | low density lipoprotein cholesterol measurement | GCST007141 | 9:133266456 | 4.18E-05 |
| rs2519093-C | 7 x 10-65 | | NR | '- | 0.092 unit | '- | ABO | Low density lipoprotein chol | low density lipoprotein cholesterol measurement | GCST007141 | 9:133266456 | 4.18E-05 |
| rs507666-G | 5 x 10-48 | (EA) | 0.806 | '- | 0.087 unit | '- | ABO | Low density lipoprotein chol | low density lipoprotein cholesterol measurement | GCST007141 | 9:133273983 | 2.24E-05 |
| rs507666-G | 2 x 10-14 | (Hispanic) | 0.831 | '- | 0.155 unit | '- | ABO | Low density lipoprotein chol | low density lipoprotein cholesterol measurement | GCST007141 | 9:133273983 | 2.24E-05 |
| rs507666-G | 1 x 10-63 | | NR | '- | 0.092 unit | '- | ABO | Low density lipoprotein chol | low density lipoprotein cholesterol measurement | GCST007141 | 9:133273983 | 2.24E-05 |
| rs507666-A | 2 x 10-11 | (LDL) | 0.23 | '- | 0.073 mm | [0.044-0.102] | ABO | Lipid traits | low density lipoprotein cholesterol measurement | GCST002321 | 9:133273983 | 2.24E-05 |
| rs507666-A | 4 x 10-11 | (TC) | 0.23 | '- | 0.015 mm | [0.0091-0.0209 | ABO | Lipid traits | total cholesterol measurement | GCST002321 | 9:133273983 | 2.24E-05 |
| rs505922-? | 2 x 10-13 | | NR | 1.27 | '- | [1.19-1.35] | ABO | Pancreatic cancer | pancreatic carcinoma | GCST002991 | 9:133273813 | 3.21E-07 |
| rs505922-? | 8 x 10-65 | (alkaline phos | '- | '- | '- | '- | ABO | Clinical laboratory measurer | alkaline phosphatase measurement, clinical laboratory m | GCST003540 | 9:133273813 | 3.21E-07 |
| rs2519093-T | 1 x 10-11 | | 0.190872 | 1.08 | '- | [1.06- 1.11] | ABO | Coronary artery disease | coronary artery disease | GCST003116 | 9:133266456 | 4.18E-05 |
| rs8176685-G | 3 x 10-9 | | 0.1809 | '- | 0.0274764 | [0.018-0.037] | ABO | Sum eosinophil basophil cou | basophil count, eosinophil count | GCST004624 | 9:133263363 | 3.34E-05 |
| rs2519093-C | 3 x 10-41 | (EA) | 0.806 | '- | 0.078 unit | '- | ABO | Total cholesterol levels | total cholesterol measurement | GCST007143 | 9:133266456 | 4.18E-05 |
| rs2519093-C | 1 x 10-10 | (Hispanic) | 0.831 | '- | 0.129 unit | '- | ABO | Total cholesterol levels | total cholesterol measurement | GCST007143 | 9:133266456 | 4.18E-05 |
| rs2519093-C | 2 x 10-53 | | NR | '- | 0.082 unit | '- | ABO | Total cholesterol levels | total cholesterol measurement | GCST007143 | 9:133266456 | 4.18E-05 |
| rs507666-G | 9 x 10-41 | (EA) | 0.806 | '- | 0.078 unit | '- | ABO | Total cholesterol levels | total cholesterol measurement | GCST007143 | 9:133273983 | 2.24E-05 |
| rs507666-G | 1 x 10-10 | (Hispanic) | 0.831 | '- | 0.129 unit | '- | ABO | Total cholesterol levels | total cholesterol measurement | GCST007143 | 9:133273983 | 2.24E-05 |
| rs507666-G | 4 x 10-52 | | NR | '- | 0.081 unit | '- | ABO | Total cholesterol levels | total cholesterol measurement | GCST007143 | 9:133273983 | 2.24E-05 |
| rs115478735- | 8 x 10-24 | (Protein-tyros | 0.178 | '- | 0.32 unit i | [0.26-0.38] | ABO | Blood protein levels | blood protein measurement | GCST005806 | 9:133274295 | 1.68E-05 |
| rs115478735- | 1 x 10-46 | (Immunoglob | 0.178 | '- | 0.45 unit c | [0.39-0.51] | ABO | Blood protein levels | blood protein measurement | GCST005806 | 9:133274295 | 1.68E-05 |
| rs2519093-? | 2 x 10-887 | | NR | '- | 0.3104 uni | [0.3-0.32] | ABO | Serum alkaline phosphatase | alkaline phosphatase measurement | GCST006016 | 9:133266456 | 4.18E-05 |
| rs8176719-? | 4 x 10-403 | | 0.63 | '- | 0.1488 uni | [0.14-0.16] | ABO | Factor VIII levels | factor VIII measurement | GCST007445 | 9:133257522 | 9.93E-08 |
| rs8176719-? | 2 x 10-307 | (EA) | 0.62 | '- | 0.1442 uni | [0.14-0.15] | ABO | Factor VIII levels | factor VIII measurement | GCST007445 | 9:133257522 | 9.93E-08 |
| rs2519093-T | 4 x 10-305 | | 0.188 | '- | 0.903 unit | '- | ABO | Soluble E-selectin levels | e-selectin measurement | GCST008202 | 9:133266456 | 4.18E-05 |
| rs2519093-T | 7 x 10-48 | | 0.188 | '- | 0.352 unit | '- | ABO | Soluble ICAM-1 | ICAM-1 measurement | GCST008210 | 9:133266456 | 4.18E-05 |
| rs2519093-? | 4 x 10-34 | | NR | '- | 2.633 unit | [2.2-3.06] | ABO | LDL cholesterol levels x sh | sleep duration, low density lipoprotein cholesterol measu | GCST009365 | 9:133266456 | 4.18E-05 |
| rs2519093-T | 4 x 10-169 | | NR | 1.4 | '- | [1.37-1.43] | ABO | Venous thromboembolism | venous thromboembolism | GCST009030 | 9:133266456 | 4.18E-05 |
| rs2519093-T | 8 x 10-102 | | 0.1808 | '- | 0.0737 mg | [0.067-0.08] | ABO | LDL cholesterol | low density lipoprotein cholesterol measurement | GCST006612 | 9:133266456 | 4.18E-05 |
| rs507666-A | 3 x 10-9 | | 0.1859 | 1,046 | '- | [1.031-1.061] | ABO | Hay fever and/or eczema | Eczema, allergic rhinitis | GCST009717 | 9:133273983 | 2.24E-05 |
| rs507666-? | 5 x 10-15 | (CDH1) | '- | '- | 0.3397 uni | [NR] | ABO | Blood protein levels | blood protein measurement | GCST010104 | 9:133273983 | 2.24E-05 |
| rs507666-? | 1 x 10-58 | (PECAM1) | '- | '- | 0.641 unit | [NR] | ABO | Blood protein levels | blood protein measurement | GCST010104 | 9:133273983 | 2.24E-05 |
| rs507666-? | 2 x 10-104 | (SELE) | '- | '- | 0.9406 uni | [NR] | ABO | Blood protein levels | blood protein measurement | GCST010104 | 9:133273983 | 2.24E-05 |
| rs507666-A | 5 x 10-12 | (gal-8) | 0.1316 | '- | 0.52713 u | [0.38-0.68] | ABO | Neurological blood protein b | blood protein measurement | GCST008478 | 9:133273983 | 2.24E-05 |
| rs2519093-? | 4 x 10-24 | | NR | '- | '- | '- | ABO | Eosinophil counts | eosinophil count | GCST007065 | 9:133266456 | 4.18E-05 |
| rs2519093-? | 6 x 10-52 | | NR | '- | '- | '- | ABO | White blood cell count | leukocyte count | GCST007070 | 9:133266456 | 4.18E-05 |
| rs495828-A | 3 x 10-8 | | 0.17 | '- | 4.9 | [NR] | AL772161.2, A | Angiotensin-converting enzy | angiotensin converting enzyme activity measurement | GCST000565 | 9:133279294 | 1.18E-05 |
| rs495828-T | 3 x 10-12 | | 0.28 | '- | 0.091 unit | [0.066-0.116] | AL772161.2, A | Red blood cell count | erythrocyte count | GCST000588 | 9:133279294 | 1.18E-05 |
| rs495828-T | 6 x 10-10 | (Ht) | 0.28 | '- | 0.081 unit | [0.056-0.106] | AL772161.2, A | Hematological and biochem | hematocrit | GCST000583 | 9:133279294 | 1.18E-05 |
| rs495828-T | 4 x 10-59 | (ALP) | 0.28 | '- | 0.308 unit | [0.27-0.35] | AL772161.2, A | Hematological and biochem | alkaline phosphatase measurement | GCST000583 | 9:133279294 | 1.18E-05 |
| rs495828-T | 1 x 10-11 | (Hb) | 0.28 | '- | 0.089 unit | [0.064-0.114] | AL772161.2, A | Hematological and biochem | hemoglobin measurement | GCST000583 | 9:133279294 | 1.18E-05 |
| rs579459-C | 1 x 10-29 | | 0.20 | '- | '- | '- | ABO, AL7721( | Soluble E-selectin levels | e-selectin measurement | GCST000476 | 9:133278724 | 1.36E-05 |
| rs579459-C | 4 x 10-14 | | 0.21 | 1.1 | '- | [1.07-1.13] | ABO, AL7721( | Coronary heart disease | coronary heart disease | GCST000998 | 9:133278724 | 1.36E-05 |
| rs579459-T | 2 x 10-41 | (P-Selectin) | NR | '- | 14 % incr | [12.04-15.96] | ABO, AL7721( | Soluble levels of adhesion m | adhesion molecule measurement, soluble P-selectin mea | GCST000599 | 9:133278724 | 1.36E-05 |
| rs579459-T | 3 x 10-123 | | 0.80 | '- | 8.8 % incr | [7.40-10.2] | ABO, AL7721( | Liver enzyme levels (alkalin | alkaline phosphatase measurement | GCST001276 | 9:133278724 | 1.36E-05 |
| rs495828-T | 3 x 10-16 | | 0.272 | 1.65 | '- | [1.46-1.86] | AL772161.2, A | Venous thromboembolism | venous thromboembolism | GCST001557 | 9:133279294 | 1.18E-05 |
| rs579459-C | 2 x 10-9 | | NR | '- | '- | '- | ABO, AL7721( | Coronary artery disease or i | stroke, coronary heart disease | GCST002287 | 9:133278724 | 1.36E-05 |
| rs579459-C | 2 x 10-7 | | NR | 1.1 | '- | [1.06-1.14] | ABO, AL7721( | Coronary artery disease | coronary heart disease | GCST002289 | 9:133278724 | 1.36E-05 |
| rs579459-? | 3 x 10-8 | | '- | '- | '- | '- | ABO, AL7721( | Coronary artery disease or l | large artery stroke, coronary heart disease | GCST002290 | 9:133278724 | 1.36E-05 |
| rs579459-T | 1 x 10-28 | (2.0525, Unk | 0.27 | '- | 0.49 unit i | [NR] | ABO, AL7721( | Urinary metabolites (H-NM | urinary metabolite measurement | GCST002364 | 9:133278724 | 1.36E-05 |
| rs579459-T | 2 x 10-32 | (5.1825, Unk | 0.27 | '- | 0.53 unit i | [NR] | ABO, AL7721( | Urinary metabolites (H-NM | urinary metabolite measurement | GCST002364 | 9:133278724 | 1.36E-05 |
| rs579459-T | 7 x 10-13 | (BMI unadjus | 0.78 | '- | 0.082 unit | [0.06-0.104] | ABO, AL7721( | Total cholesterol levels | total cholesterol measurement | GCST004231 | 9:133278724 | 1.36E-05 |
| rs579459-T | 2 x 10-51 | (Trans-ethnic | 0.785 | '- | 0.0665 uni | [0.058-0.075] | ABO, AL7721( | LDL cholesterol levels | low density lipoprotein cholesterol measurement | GCST004233 | 9:133278724 | 1.36E-05 |
| rs579459-T | 2 x 10-9 | (Asian initial, | 0.65 | '- | 0.057 unit | [0.039-0.075] | ABO, AL7721( | LDL cholesterol levels | low density lipoprotein cholesterol measurement | GCST004233 | 9:133278724 | 1.36E-05 |
| rs579459-T | 1 x 10-43 | (EA) | 0.786 | '- | 0.079 unit | '- | ABO, AL7721( | Low density lipoprotein chol | low density lipoprotein cholesterol measurement | GCST007141 | 9:133278724 | 1.36E-05 |
| rs579459-T | 3 x 10-14 | (Hispanic) | 0.812 | '- | 0.147 unit | '- | ABO, AL7721( | Low density lipoprotein chol | low density lipoprotein cholesterol measurement | GCST007141 | 9:133278724 | 1.36E-05 |
| rs579459-T | 4 x 10-7 | (East Asian) | 0.79 | '- | 0.099 unit | '- | ABO, AL7721( | Low density lipoprotein chol | low density lipoprotein cholesterol measurement | GCST007141 | 9:133278724 | 1.36E-05 |
| rs579459-T | 6 x 10-60 | | NR | '- | 0.085 unit | '- | ABO, AL7721( | Low density lipoprotein chol | low density lipoprotein cholesterol measurement | GCST007141 | 9:133278724 | 1.36E-05 |
| rs495828-T | 6 x 10-34 | (DSGEGDFX | 0.22 | '- | 0.088 unit | [0.074-0.102] | AL772161.2, A | Blood metabolite ratios | blood metabolite measurement | GCST002442 | 9:133279294 | 1.18E-05 |
| rs579459-T | 1 x 10-28 | (ADpSGEGD | 0.79 | '- | 0.124 unit | [0.1-0.15] | ABO, AL7721( | Blood metabolite ratios | blood metabolite measurement | GCST002442 | 9:133278724 | 1.36E-05 |
| rs579459-T | 9 x 10-18 | (EA, RBCC) | 0.8 | '- | 0.021 unit | [0.015-0.027] | ABO, AL7721( | Red blood cell traits | erythrocyte count | GCST001765 | 9:133278724 | 1.36E-05 |
| rs633862-A | 9 x 10-7 | | 0.54 | 1.19 | '- | [1.02-1.39] | AL772161.2, A | Epithelial ovarian cancer | Malignant epithelial tumor of ovary | GCST002576 | 9:133279871 | 8.79E-07 |
| rs579459-C | 3 x 10-10 | | 0.2 | '- | 3.038 unit | [2.1-3.98] | ABO, AL7721( | Cholesterol, total | total cholesterol measurement | GCST003214 | 9:133278724 | 1.36E-05 |
| rs579459-C | 6 x 10-13 | | 0.2 | '- | 2.853 unit | [2.08-3.62] | ABO, AL7721( | LDL cholesterol | low density lipoprotein cholesterol measurement | GCST003216 | 9:133278724 | 1.36E-05 |
| rs579459-C | 9 x 10-9 | | 0.2389 | '- | 0.0107 uni | [0.007-0.0144] | ABO, AL7721( | Glycated hemoglobin levels | HbA1c measurement | GCST007954 | 9:133278724 | 1.36E-05 |
| rs579459-T | 9 x 10-37 | (EA) | 0.786 | '- | 0.071 unit | '- | ABO, AL7721( | Total cholesterol levels | total cholesterol measurement | GCST007143 | 9:133278724 | 1.36E-05 |
| rs579459-T | 2 x 10-10 | (Hispanic) | 0.812 | '- | 0.122 unit | '- | ABO, AL7721( | Total cholesterol levels | total cholesterol measurement | GCST007143 | 9:133278724 | 1.36E-05 |
| rs579459-T | 1 x 10-48 | | NR | '- | 0.075 unit | '- | ABO, AL7721( | Total cholesterol levels | total cholesterol measurement | GCST007143 | 9:133278724 | 1.36E-05 |
| rs495828-T | 1 x 10-6 | | NR | '- | '- | '- | AL772161.2, A | Triglyceride levels x alcohol | triglyceride measurement, alcohol drinking | GCST008083 | 9:133279294 | 1.18E-05 |
| rs495828-T | 5 x 10-69 | (EA) | NR | '- | '- | '- | AL772161.2, A | LDL cholesterol levels x alc | low density lipoprotein cholesterol measurement, alcoho | GCST008079 | 9:133279294 | 1.18E-05 |
| rs495828-T | 1 x 10-7 | (Asian) | NR | '- | '- | '- | AL772161.2, A | Triglyceride levels x alcohol | triglyceride measurement, alcohol drinking | GCST008083 | 9:133279294 | 1.18E-05 |
| rs495828-T | 4 x 10-86 | | NR | '- | '- | '- | AL772161.2, A | LDL cholesterol levels x alc | low density lipoprotein cholesterol measurement, alcoho | GCST008079 | 9:133279294 | 1.18E-05 |
| rs495828-T | 4 x 10-103 | (EA) | NR | '- | '- | '- | AL772161.2, A | LDL cholesterol levels x alc | low density lipoprotein cholesterol measurement, alcoho | GCST008078 | 9:133279294 | 1.18E-05 |
| rs495828-T | 2 x 10-7 | (Asian) | NR | '- | '- | '- | AL772161.2, A | Triglyceride levels x alcohol | triglyceride measurement, alcohol consumption measure | GCST008074 | 9:133279294 | 1.18E-05 |
| rs600038-C | 4 x 10-9 | | 0.21 | 1.06 | '- | [1.04-1.08] | ABO, AL7721( | Heart failure | heart failure | GCST009541 | 9:133276354 | 1.49E-05 |
| rs579459-C | 4 x 10-145 | | NR | 1.36 | '- | [1.33-1.39] | ABO, AL7721( | Venous thromboembolism | venous thromboembolism | GCST009030 | 9:133278724 | 1.36E-05 |
| rs579459-C | 9 x 10-193 | (SELE) | 0.202 | '- | 0.963 unit | [NR] | ABO, AL7721( | Blood protein levels | blood protein measurement | GCST006585 | 9:133278724 | 1.36E-05 |
| rs579459-C | 8 x 10-125 | (ADGRF5) | 0.202 | '- | 0.815 unit | [NR] | ABO, AL7721( | Blood protein levels | blood protein measurement | GCST006585 | 9:133278724 | 1.36E-05 |
| rs579459-C | 1 x 10-114 | (ROBO4) | 0.202 | '- | 0.73 unit c | [NR] | ABO, AL7721( | Blood protein levels | blood protein measurement | GCST006585 | 9:133278724 | 1.36E-05 |
| rs579459-C | 5 x 10-81 | (IL3RA) | 0.202 | '- | 0.613 unit | [NR] | ABO, AL7721( | Blood protein levels | blood protein measurement | GCST006585 | 9:133278724 | 1.36E-05 |
| rs579459-C | 4 x 10-73 | (QSOX2) | 0.202 | '- | 0.625 unit | [NR] | ABO, AL7721( | Blood protein levels | blood protein measurement | GCST006585 | 9:133278724 | 1.36E-05 |
| rs579459-C | 3 x 10-70 | (INSR) | 0.202 | '- | 0.595 unit | [NR] | ABO, AL7721( | Blood protein levels | blood protein measurement | GCST006585 | 9:133278724 | 1.36E-05 |
| rs579459-C | 3 x 10-67 | (ICAM2) | 0.202 | '- | 0.593 unit | [NR] | ABO, AL7721( | Blood protein levels | blood protein measurement | GCST006585 | 9:133278724 | 1.36E-05 |
| rs579459-C | 8 x 10-66 | (FAM3D) | 0.202 | '- | 0.581 unit | [NR] | ABO, AL7721( | Blood protein levels | blood protein measurement | GCST006585 | 9:133278724 | 1.36E-05 |
| rs579459-C | 5 x 10-50 | (KDR) | 0.202 | '- | 0.508 unit | [NR] | ABO, AL7721( | Blood protein levels | blood protein measurement | GCST006585 | 9:133278724 | 1.36E-05 |
| rs579459-C | 2 x 10-49 | (EPHA4) | 0.202 | '- | 0.492 unit | [NR] | ABO, AL7721( | Blood protein levels | blood protein measurement | GCST006585 | 9:133278724 | 1.36E-05 |
| rs579459-C | 5 x 10-40 | (ICAM5) | 0.202 | '- | 0.457 unit | [NR] | ABO, AL7721( | Blood protein levels | blood protein measurement | GCST006585 | 9:133278724 | 1.36E-05 |
| rs579459-C | 1 x 10-36 | (FLT4) | 0.202 | '- | 0.442 unit | [NR] | ABO, AL7721( | Blood protein levels | blood protein measurement | GCST006585 | 9:133278724 | 1.36E-05 |
| rs579459-C | 9 x 10-32 | (F8) | 0.202 | '- | 0.393 unit | [NR] | ABO, AL7721( | Blood protein levels | blood protein measurement | GCST006585 | 9:133278724 | 1.36E-05 |
| rs579459-C | 1 x 10-30 | (ENG) | 0.202 | '- | 0.394 unit | [NR] | ABO, AL7721( | Blood protein levels | blood protein measurement | GCST006585 | 9:133278724 | 1.36E-05 |
| rs579459-C | 1 x 10-28 | (ISLR2) | 0.202 | '- | 0.387 unit | [NR] | ABO, AL7721( | Blood protein levels | blood protein measurement | GCST006585 | 9:133278724 | 1.36E-05 |
| rs579459-C | 2 x 10-28 | (KIN) | 0.202 | '- | 0.376 unit | [NR] | ABO, AL7721( | Blood protein levels | blood protein measurement | GCST006585 | 9:133278724 | 1.36E-05 |
| rs579459-C | 2 x 10-28 | (GOLM1) | 0.202 | '- | 0.379 unit | [NR] | ABO, AL7721( | Blood protein levels | blood protein measurement | GCST006585 | 9:133278724 | 1.36E-05 |
| rs579459-C | 2 x 10-25 | (CD200) | 0.202 | '- | 0.357 unit | [NR] | ABO, AL7721( | Blood protein levels | blood protein measurement | GCST006585 | 9:133278724 | 1.36E-05 |

| | | | | | | | | | | | | |
|---|---|---|---|---|---|---|---|---|---|---|---|---|
| rs579459-C | 3 x 10-25 | (MET) | 0.202 | '- | 0.367 unit | [NR] | ABO, AL7721( | Blood protein levels | blood protein measurement | GCST006585 | 9:133278724 | 1.36E-05 |
| rs579459-C | 1 x 10-24 | (GLCE) | 0.202 | '- | 0.36 unit i | [NR] | ABO, AL7721( | Blood protein levels | blood protein measurement | GCST006585 | 9:133278724 | 1.36E-05 |
| rs579459-C | 1 x 10-22 | (LIFR) | 0.202 | '- | 0.337 unit | [NR] | ABO, AL7721( | Blood protein levels | blood protein measurement | GCST006585 | 9:133278724 | 1.36E-05 |
| rs579459-C | 2 x 10-20 | (C1GALT1C | 0.202 | '- | 0.326 unit | [NR] | ABO, AL7721( | Blood protein levels | blood protein measurement | GCST006585 | 9:133278724 | 1.36E-05 |
| rs579459-C | 4 x 10-20 | (SHANK3) | 0.202 | '- | 0.314 unit | [NR] | ABO, AL7721( | Blood protein levels | blood protein measurement | GCST006585 | 9:133278724 | 1.36E-05 |
| rs579459-C | 5 x 10-20 | (ICAM4) | 0.202 | '- | 0.323 unit | [NR] | ABO, AL7721( | Blood protein levels | blood protein measurement | GCST006585 | 9:133278724 | 1.36E-05 |
| rs579459-C | 2 x 10-19 | (ACE) | 0.202 | '- | 0.316 unit | [NR] | ABO, AL7721( | Blood protein levels | blood protein measurement | GCST006585 | 9:133278724 | 1.36E-05 |
| rs579459-C | 1 x 10-17 | (CHST15) | 0.202 | '- | 0.292 unit | [NR] | ABO, AL7721( | Blood protein levels | blood protein measurement | GCST006585 | 9:133278724 | 1.36E-05 |
| rs579459-C | 7 x 10-13 | (SELP) | 0.202 | '- | 0.248 unit | [NR] | ABO, AL7721( | Blood protein levels | blood protein measurement | GCST006585 | 9:133278724 | 1.36E-05 |
| rs579459-C | 1 x 10-12 | (IGF1R) | 0.202 | '- | 0.237 unit | [NR] | ABO, AL7721( | Blood protein levels | blood protein measurement | GCST006585 | 9:133278724 | 1.36E-05 |
| rs579459-C | 1 x 10-12 | (CDH5) | 0.202 | '- | 0.242 unit | [NR] | ABO, AL7721( | Blood protein levels | blood protein measurement | GCST006585 | 9:133278724 | 1.36E-05 |
| rs579459-C | 2 x 10-12 | (VWF) | 0.202 | '- | 0.244 unit | [NR] | ABO, AL7721( | Blood protein levels | blood protein measurement | GCST006585 | 9:133278724 | 1.36E-05 |
| rs579459-C | 6 x 10-12 | (SEMA6A) | 0.202 | '- | 0.236 unit | [NR] | ABO, AL7721( | Blood protein levels | blood protein measurement | GCST006585 | 9:133278724 | 1.36E-05 |
| rs579459-C | 2 x 10-11 | (L1CAM) | 0.202 | '- | 0.232 unit | [NR] | ABO, AL7721( | Blood protein levels | blood protein measurement | GCST006585 | 9:133278724 | 1.36E-05 |
| rs579459-C | 7 x 10-10 | (CD109) | 0.202 | '- | 0.211 unit | [NR] | ABO, AL7721( | Blood protein levels | blood protein measurement | GCST006585 | 9:133278724 | 1.36E-05 |
| rs579459-C | 8 x 10-10 | (CCL28) | 0.202 | '- | 0.217 unit | [NR] | ABO, AL7721( | Blood protein levels | blood protein measurement | GCST006585 | 9:133278724 | 1.36E-05 |
| rs579459-C | 3 x 10-9 | (IL6ST) | 0.202 | '- | 0.202 unit | [NR] | ABO, AL7721( | Blood protein levels | blood protein measurement | GCST006585 | 9:133278724 | 1.36E-05 |
| rs495828-T | 3 x 10-10 | (gal-8) | 0.1582 | '- | 0.44528 u | [0.31-0.58] | AL772161.2, A | Neurological blood protein b | blood protein measurement | GCST008478 | 9:133279294 | 1.18E-05 |
| rs495828-? | 6 x 10-9 | | NR | '- | '- | '- | AL772161.2, A | Eczema | Eczema | GCST007075 | 9:133279294 | 1.18E-05 |
| rs507666-A | 2 x 10-18 | | 0.18 | '- | 0.067 s.d. | [0.051-0.083] | ABO | Cholesterol, total | total cholesterol measurement | GCST002896 | 9:133273983 | 2.24E-05 |
| rs2519093-? | 4 x 10-8 | | NR | 1,055 | '- | [1.035-1.075] | ABO | Allergy | allergy | GCST003990 | 9:133266456 | 4.18E-05 |
| rs507666-G | 8 x 10-6 | | '- | '- | 0.145 unit | '- | ABO | Pulse pressure | pulse pressure measurement | GCST007096 | 9:133273983 | 2.24E-05 |
| rs507666-G | 2 x 10-8 | | '- | '- | 0.169 unit | '- | ABO | Diastolic blood pressure | diastolic blood pressure | GCST007094 | 9:133273983 | 2.24E-05 |
| rs8176643-A | 3 x 10-71 | | 0.1761 | '- | 0.0835811 | [0.074-0.093] | ABO | Hemoglobin concentration | hemoglobin measurement | GCST004615 | 9:133274294 | 1.73E-05 |
| rs8176643-? | 4 x 10-11 | | 0.36 | 1.06 | '- | [1.05-1.07] | ABO | Childhood ear infection | susceptibility to childhood ear infection measurement | GCST005013 | 9:133274294 | 1.73E-05 |
| rs505922-C | 8 x 10-86 | (CD209 antig | 0.3544177 | '- | 0.8177 uni | [0.74-0.89] | ABO | Blood protein levels | CD209 antigen measurement | GCST004365 | 9:133273813 | 3.21E-07 |
| rs505922-C | 2 x 10-30 | | NR | '- | 0.3315 uni | [0.28-0.39] | ABO | vWF levels in ischaemic stro | Ischemic stroke, von Willebrand factor measurement, h | GCST004598 | 9:133273813 | 3.21E-07 |
| rs2519093-T | 6 x 10-26 | | 0.19 | 1.41 | '- | [1.32,Äì1.50] | ABO | Venous thromboembolism | venous thromboembolism | GCST004256 | 9:133266456 | 4.18E-05 |
| rs8176645-A | 4 x 10-21 | | 0.33 | 1.28 | '- | [1.22,Äì1.35] | ABO | Venous thromboembolism | venous thromboembolism | GCST004256 | 9:133273682 | 1.74E-07 |
| rs8176643-A | 6 x 10-64 | | 0.176 | '- | 0.0787670 | [0.07-0.088] | ABO | Hematocrit | hematocrit | GCST004604 | 9:133274294 | 1.73E-05 |
| rs612169-G | 1 x 10-20 | (von Willebra | 0.3539157 | '- | 0.417 unit | [0.33-0.5] | ABO | Blood protein levels | von Willebrand factor measurement | GCST004365 | 9:133268030 | 1.08E-07 |
| rs507666-A | 1 x 10-12 | | 0.192 | 1.08 | '- | [1.06-1.10] | ABO | Coronary artery disease (my | coronary artery disease | GCST004787 | 9:133273983 | 2.24E-05 |
| rs2519093-T | 1 x 10-78 | (D-glucurony | 0.178 | '- | 0.58 unit i | [0.52-0.64] | ABO | Blood protein levels | blood protein measurement | GCST005806 | 9:133266456 | 4.18E-05 |
| rs2519093-T | 4 x 10-48 | (P-selectin, S | 0.178 | '- | 0.46 unit c | [0.4-0.52] | ABO | Blood protein levels | blood protein measurement | GCST005806 | 9:133266456 | 4.18E-05 |
| rs2519093-T | 2 x 10-474 | (E-selectin, S | 0.178 | '- | 1.17 unit c | [1.11-1.23] | ABO | Blood protein levels | blood protein measurement | GCST005806 | 9:133266456 | 4.18E-05 |
| rs2519093-T | 8 x 10-198 | (Interleukin-3 | 0.178 | '- | 0.86 unit c | [0.8-0.92] | ABO | Blood protein levels | blood protein measurement | GCST005806 | 9:133266456 | 4.18E-05 |
| rs2519093-T | 6 x 10-80 | (Vascular enc | 0.178 | '- | 0.58 unit c | [0.52-0.64] | ABO | Blood protein levels | blood protein measurement | GCST005806 | 9:133266456 | 4.18E-05 |
| rs2519093-T | 3 x 10-20 | (Protein FAM | 0.178 | '- | 0.29 unit i | [0.23-0.35] | ABO | Blood protein levels | blood protein measurement | GCST005806 | 9:133266456 | 4.18E-05 |
| rs2519093-T | 1 x 10-155 | (Adhesion G | 0.178 | '- | 0.78 unit c | [0.72-0.84] | ABO | Blood protein levels | blood protein measurement | GCST005806 | 9:133266456 | 4.18E-05 |
| rs2519093-T | 2 x 10-89 | (C1GALT1-s | 0.178 | '- | 0.61 unit i | [0.55-0.67] | ABO | Blood protein levels | blood protein measurement | GCST005806 | 9:133266456 | 4.18E-05 |
| rs2519093-T | 3 x 10-19 | (Thrombospo | 0.178 | '- | 0.29 unit c | [0.23-0.35] | ABO | Blood protein levels | blood protein measurement | GCST005806 | 9:133266456 | 4.18E-05 |
| rs2519093-T | 4 x 10-23 | (N-acetyllact | 0.178 | '- | 0.31 unit i | [0.25-0.37] | ABO | Blood protein levels | blood protein measurement | GCST005806 | 9:133266456 | 4.18E-05 |
| rs505922-C | 7 x 10-27 | (EA) | 0.35 | 1.27 | '- | [1.22-1.31] | ABO | Pancreatic cancer | pancreatic carcinoma | GCST005434 | 9:133273813 | 3.21E-07 |
| rs2519093-T | 2 x 10-14 | | 0.1841 | '- | 0.0554 uni | [0.041-0.07] | ABO | Coronary artery disease | coronary artery disease | GCST005195 | 9:133266456 | 4.18E-05 |
| rs2519093-? | 1 x 10-33 | | NR | '- | 0.05699 u | [0.048-0.066] | ABO | Hematocrit | hematocrit | GCST005994 | 9:133266456 | 4.18E-05 |
| rs2519093-? | 1 x 10-35 | | NR | '- | 0.0588 uni | [0.05-0.068] | ABO | Hemoglobin | hemoglobin measurement | GCST005995 | 9:133266456 | 4.18E-05 |
| rs2519093-? | 7 x 10-45 | | NR | '- | 0.06646 u | [0.057-0.076] | ABO | Red blood cell count | erythrocyte count | GCST005996 | 9:133266456 | 4.18E-05 |
| rs507666-A | 5 x 10-20 | | 0.1918 | '- | 0.0734 uni | [0.058-0.089] | ABO | Coronary artery disease | coronary artery disease | GCST005196 | 9:133273983 | 2.24E-05 |
| rs505922-C | 7 x 10-11 | | 0.3371 | 1.06 | '- | [1.04-1.07] | ABO | Peripheral artery disease | peripheral arterial disease | GCST008474 | 9:133273813 | 3.21E-07 |
| rs8176685-? | 1 x 10-320 | (EA) | NR | '- | '- | '- | ABO | vWF levels | von Willebrand factor measurement | GCST007446 | 9:133263363 | 3.34E-05 |
| rs8176719-? | 1 x 10-320 | (EA) | 0.65 | '- | 0.172 unit | [0.17-0.18] | ABO | vWF levels | von Willebrand factor measurement | GCST007446 | 9:133257522 | 9.93E-08 |
| rs8176643-A | 4 x 10-24 | (Toll-like rece | 0.171 | '- | 0.33 unit c | [0.27-0.39] | ABO | Blood protein levels | blood protein measurement | GCST005806 | 9:133274294 | 1.73E-05 |
| rs8176643-A | 6 x 10-12 | (Mannosyl-oli | 0.171 | '- | 0.23 unit i | [0.17-0.29] | ABO | Blood protein levels | blood protein measurement | GCST005806 | 9:133274294 | 1.73E-05 |
| rs8176643-A | 2 x 10-64 | (Coagulation | 0.171 | '- | 0.54 unit i | [0.48-0.6] | ABO | Blood protein levels | blood protein measurement | GCST005806 | 9:133274294 | 1.73E-05 |
| rs8176643-A | 1 x 10-162 | (CD209 antig | 0.171 | '- | 0.81 unit i | [0.75-0.87] | ABO | Blood protein levels | blood protein measurement | GCST005806 | 9:133274294 | 1.73E-05 |
| rs8176685-? | 6 x 10-507 | | NR | '- | '- | '- | ABO | vWF levels | von Willebrand factor measurement | GCST007446 | 9:133263363 | 3.34E-05 |
| rs505922-C | 4 x 10-12 | | 0.3317 | 1.05 | '- | [1.03-1.06] | ABO | Type 2 diabetes | type II diabetes mellitus | GCST009379 | 9:133273813 | 3.21E-07 |
| rs2519093-? | 1 x 10-33 | | NR | '- | 2.472 unit | [2.04-2.91] | ABO | LDL cholesterol levels x lon | sleep duration, low density lipoprotein cholesterol measu | GCST009366 | 9:133266456 | 4.18E-05 |
| rs507666-A | 5 x 10-16 | | 0.14995 | '- | 0.55 unit c | [0.42-0.68] | ABO | E-selectin levels | e-selectin measurement | GCST009572 | 9:133273983 | 2.24E-05 |
| rs507666-A | 1 x 10-57 | | 0.1853 | '- | '- | '- | ABO | LDL cholesterol x physical | low density lipoprotein cholesterol measurement, physic | GCST007284 | 9:133273983 | 2.24E-05 |
| rs2519093-T | 3 x 10-16 | | 0.1957 | 1.05 | '- | [1.04-1.07] | ABO | Allergic rhinitis | allergic rhinitis | GCST006409 | 9:133266456 | 4.18E-05 |
| rs505922-C | 1 x 10-1828 | (Histo-blood | 0.313 | '- | 1.3 unit in | [1.28-1.32] | ABO | Blood protein levels | blood protein measurement | GCST005806 | 9:133273813 | 3.21E-07 |
| rs505922-C | 1 x 10-301 | (CD209 antig | 0.313 | '- | 0.83 unit i | [0.79-0.87] | ABO | Blood protein levels | blood protein measurement | GCST005806 | 9:133273813 | 3.21E-07 |
| rs507666-A | 2 x 10-17 | (Intercellular | 0.179 | '- | 0.27 unit c | [0.21-0.33] | ABO | Blood protein levels | blood protein measurement | GCST005806 | 9:133273983 | 2.24E-05 |
| rs507666-A | 5 x 10-80 | (Insulin recep | 0.179 | '- | 0.58 unit c | [0.52-0.64] | ABO | Blood protein levels | blood protein measurement | GCST005806 | 9:133273983 | 2.24E-05 |
| rs2519093-T | 7 x 10-31 | | 0.185023 | '- | 0.0835069 | [0.069-0.098] | ABO | Medication use (HMG CoA | HMG CoA reductase inhibitor use measurement | GCST007931 | 9:133266456 | 4.18E-05 |
| rs507666-? | 1 x 10-14 | | NR | '- | 3.665804 | [2.73-4.6] | ABO | Total cholesterol levels | total cholesterol measurement | GCST008045 | 9:133273983 | 2.24E-05 |
| rs2769071-A | 6 x 10-17 | (EA) | 0.6491 | '- | 0.037 unit | [0.027-0.047] | ABO | Circulating fibroblast growth | fibroblast growth factor 23 measurement | GCST006491 | 9:133270565 | 1.17E-07 |
| rs507666-? | 4 x 10-15 | | NR | '- | 3.361036 | [2.52-4.2] | ABO | Low density lipoprotein chol | low density lipoprotein cholesterol measurement | GCST008037 | 9:133273983 | 2.24E-05 |
| rs505922-C | 4 x 10-9 | | 0.471 | '- | 0.038 unit | [0.026-0.05] | ABO | Insulin disposition index | disposition index measurement | GCST008111 | 9:133273813 | 3.21E-07 |
| rs495828-? | 1 x 10-8 | | NR | '- | '- | '- | AL772161.2, A | Red blood cell count | erythrocyte count | GCST004008 | 9:133279294 | 1.18E-05 |
| rs495828-T | 3 x 10-9 | (Asian) | NR | '- | 0.2708 pe | [0.18-0.36] | AL772161.2, A | Hematocrit | hematocrit | GCST004003 | 9:133279294 | 1.18E-05 |
| rs579459-T | 4 x 10-11 | (Asian) | NR | '- | 0.0086 uni | [0.0061-0.0111 | ABO, AL7721( | Red blood cell count | erythrocyte count | GCST004008 | 9:133278724 | 1.36E-05 |
| rs495828-T | 2 x 10-10 | (Asian) | NR | '- | 0.1033 gra | [0.072-0.135] | AL772161.2, A | Hemoglobin levels | hemoglobin measurement | GCST004005 | 9:133279294 | 1.18E-05 |
| rs495828-? | 1 x 10-19 | | NR | '- | '- | '- | AL772161.2, A | Hemoglobin levels | hemoglobin measurement | GCST004005 | 9:133279294 | 1.18E-05 |
| rs579459-T | 7 x 10-13 | (BMI unadjus | 0.78 | '- | 0.082 unit | [0.06-0.104] | ABO, AL7721( | LDL cholesterol levels | low density lipoprotein cholesterol measurement | GCST004236 | 9:133278724 | 1.36E-05 |
| rs579459-T | 4 x 10-49 | (Trans-ethnic | 0.785 | '- | 0.062 unit | [0.053-0.071] | ABO, AL7721( | Total cholesterol levels | total cholesterol measurement | GCST004235 | 9:133278724 | 1.36E-05 |
| rs579459-T | 5 x 10-10 | (Asian initial, | 0.65 | '- | 0.059 unit | [0.041-0.077] | ABO, AL7721( | Total cholesterol levels | total cholesterol measurement | GCST004235 | 9:133278724 | 1.36E-05 |
| rs495828-T | 9 x 10-12 | | 0.202 | '- | 0.06 unit i | [0.042-0.078] | AL772161.2, A | Coronary artery disease | coronary artery disease | GCST005194 | 9:133279294 | 1.18E-05 |
| rs495828-T | 2 x 10-36 | (EA) | NR | '- | 2.645 unit | NR | AL772161.2, A | LDL cholesterol levels in cu | low density lipoprotein cholesterol measurement, alcoho | GCST008086 | 9:133279294 | 1.18E-05 |
| rs495828-T | 6 x 10-7 | (Asian) | NR | '- | 1.684 unit | NR | AL772161.2, A | LDL cholesterol levels in cu | low density lipoprotein cholesterol measurement, alcoho | GCST008086 | 9:133279294 | 1.18E-05 |
| rs579459-? | 1 x 10-9 | | NR | '- | '- | '- | ABO, AL7721( | Platelet reactivity measurem | platelet reactivity measurement | GCST008457 | 9:133278724 | 1.36E-05 |
| rs495828-T | 1 x 10-7 | (Hispanic) | NR | '- | 3.35 unit i | NR | AL772161.2, A | LDL cholesterol levels in cu | low density lipoprotein cholesterol measurement, alcoho | GCST008086 | 9:133279294 | 1.18E-05 |
| rs495828-T | 8 x 10-47 | | NR | '- | 2.453 unit | NR | AL772161.2, A | LDL cholesterol levels in cu | low density lipoprotein cholesterol measurement, alcoho | GCST008086 | 9:133279294 | 1.18E-05 |
| rs495828-T | 3 x 10-7 | (Asian) | NR | '- | '- | '- | AL772161.2, A | LDL cholesterol levels x alc | low density lipoprotein cholesterol measurement, alcoho | GCST008078 | 9:133279294 | 1.18E-05 |
| rs495828-T | 2 x 10-8 | (Hispanic) | NR | '- | '- | '- | AL772161.2, A | LDL cholesterol levels x alc | low density lipoprotein cholesterol measurement, alcoho | GCST008078 | 9:133279294 | 1.18E-05 |
| rs495828-T | 1 x 10-114 | | NR | '- | '- | '- | AL772161.2, A | LDL cholesterol levels x alc | low density lipoprotein cholesterol measurement, alcoho | GCST008078 | 9:133279294 | 1.18E-05 |
| rs495828-T | 4 x 10-18 | (EA) | NR | '- | 2.471 unit | NR | AL772161.2, A | LDL cholesterol levels | low density lipoprotein cholesterol measurement | GCST008077 | 9:133279294 | 1.18E-05 |
| rs495828-T | 1 x 10-7 | (Asian) | NR | '- | 1.668 unit | NR | AL772161.2, A | LDL cholesterol levels | low density lipoprotein cholesterol measurement | GCST008077 | 9:133279294 | 1.18E-05 |
| rs495828-T | 2 x 10-8 | (Asian) | NR | '- | '- | '- | AL772161.2, A | LDL cholesterol levels x alc | low density lipoprotein cholesterol measurement, alcoho | GCST008079 | 9:133279294 | 1.18E-05 |
| rs495828-T | 8 x 10-9 | (H) | NR | '- | '- | '- | AL772161.2, A | LDL cholesterol levels x alc | low density lipoprotein cholesterol measurement, alcoho | GCST008079 | 9:133279294 | 1.18E-05 |
| rs495828-T | 7 x 10-29 | | NR | '- | 2.222 unit | NR | AL772161.2, A | LDL cholesterol levels | low density lipoprotein cholesterol measurement | GCST008077 | 9:133279294 | 1.18E-05 |
| rs579459-T | 1 x 10-26 | | 0.7972 | '- | 2.0948 uni | [1.71-2.48] | ABO, AL7721( | LDL cholesterol levels | low density lipoprotein cholesterol measurement | GCST008676 | 9:133278724 | 1.36E-05 |
| rs495828-T | 6 x 10-13 | | 0.25 | '- | 0.17 unit c | '- | AL772161.2, A | Blood protein levels in cardi | tissue factor measurement | GCST009731 | 9:133279294 | 1.18E-05 |
| rs495828-T | 1 x 10-124 | | '- | '- | 0.065 unit | [NR] | AL772161.2, A | Hemoglobin levels | hemoglobin measurement | GCST010083 | 9:133279294 | 1.18E-05 |
| rs579459-C | 2 x 10-8 | | 0.246 | '- | 3.045 unit | [1.99-4.1] | ABO, AL7721( | Serum alkaline phosphatase | alkaline phosphatase measurement | GCST010047 | 9:133278724 | 1.36E-05 |
| rs579459-T | 8 x 10-244 | (ABO) | 0.798 | '- | 1.087 unit | [1.07-1.11] | ABO, AL7721( | Blood protein levels | blood protein measurement | GCST006585 | 9:133278724 | 1.36E-05 |
| rs579459-? | 3 x 10-10 | | NR | '- | 2.857088 | [1.97-3.74] | ABO, AL7721( | Total cholesterol levels | total cholesterol measurement | GCST008045 | 9:133278724 | 1.36E-05 |
| rs579459-? | 7 x 10-11 | | NR | '- | 2.653243 | [1.86-3.45] | ABO, AL7721( | Low density lipoprotein chol | low density lipoprotein cholesterol measurement | GCST008037 | 9:133278724 | 1.36E-05 |
| rs17764831-? | 2 x 10-13 | | NR | - | - | - | LZTFL1 | Eosinophil counts | eosinophil count | GCST007065 | 3:4587672 | 4.11E-07 |

**Table S2: Independent lead GWAS loci reported in NEJM (hg38) and in hg19***

| GenomicLocus | uniqID | rsID | chr | pos-hg19 | p | nIndSigSNPs | IndSigSNPs |
|---|---|---|---|---|---|---|---|
| 1 | 1:88993151:C:T | rs75558547 | 1 | 88993151 | 4.49E-06 | 1 | rs75558547 |
| 2 | 2:15724310:A:G | rs2080811 | 2 | 15724310 | 1.12E-06 | 1 | rs2080811 |
| 3 | 3:45876459:G:GA | rs11385942 | 3 | 45876459 | 1.15E-10 | 3 | rs11385942;rs34901975;rs3774641 |
| 4 | 3:149805983:C:G | rs144582715 | 3 | 149805983 | 7.17E-06 | 1 | rs144582715 |
| 5 | 4:180344873:A:G | rs1455662 | 4 | 180344873 | 9.60E-06 | 1 | rs1455662 |
| 6 | 6:20314719:C:T | rs911360 | 6 | 20314719 | 4.09E-06 | 1 | rs911360 |
| 7 | 6:23379360:A:AT | rs5874914 | 6 | 23379360 | 4.64E-06 | 1 | rs5874914 |
| 8 | 6:91010824:A:ATTAC | rs3073485 | 6 | 91010824 | 5.72E-06 | 1 | rs3073485 |
| 9 | 7:123006727:A:G | rs12706520 | 7 | 123006727 | 9.63E-06 | 1 | rs12706520 |
| 10 | 7:158020147:A:G | rs6970487 | 7 | 158020147 | 4.89E-06 | 1 | rs6970487 |
| 11 | 8:122848829:A:G | rs10091098 | 8 | 122848829 | 4.55E-06 | 1 | rs10091098 |
| 12 | 8:126952467:A:T | rs28730361 | 8 | 126952467 | 8.11E-06 | 1 | rs28730361 |
| 13 | 9:136139265:A:C | rs657152 | 9 | 136139265 | 4.95E-08 | 3 | rs657152;rs550057;rs647800 |
| 14 | 11:5691474:C:G | rs12796811 | 11 | 5691474 | 3.62E-06 | 1 | rs12796811 |
| 15 | 12:62463979:G:T | rs10877786 | 12 | 62463979 | 6.89E-07 | 1 | rs10877786 |
| 16 | 14:32422746:C:G | rs7152677 | 14 | 32422746 | 1.52E-06 | 1 | rs7152677 |
| 17 | 16:74936031:A:G | rs2059266 | 16 | 74936031 | 4.01E-06 | 1 | rs2059266 |
| 18 | 16:82338406:C:T | rs114093749 | 16 | 82338406 | 3.86E-06 | 2 | rs114093749;rs4569267 |
| 19 | 17:10376558:G:T | rs117438562 | 17 | 10376558 | 7.49E-06 | 1 | rs117438562 |
| 20 | 18:3724602:A:G | rs13381043 | 18 | 3724602 | 1.13E-07 | 2 | rs13381043;rs4797120 |
| 21 | 18:22624707:A:G | rs56132597 | 18 | 22624707 | 8.94E-06 | 1 | rs56132597 |
| 22 | 19:4717672:A:G | rs12610495 | 19 | 4717672 | 5.20E-06 | 1 | rs12610495 |
| 23 | 19:5518492:C:T | rs183544391 | 19 | 5518492 | 7.29E-06 | 1 | rs183544391 |
| 24 | 19:17923554:T:TC | rs3833287 | 19 | 17923554 | 4.24E-06 | 1 | rs3833287 |
| 25 | 23:15138357:C:T | rs55634010 | 23 | 15138357 | 9.75E-06 | 1 | rs55634010 |
| 26 | 23:146325425:A:T | rs10126492 | 23 | 146325425 | 8.81E-06 | 1 | rs10126492 |

**Table S3 Functional annotation of SNPs on genes by ANNOVAR***

| annotation | ref.count | ref.prop | count | prop | enrichment | fisher.P |
|---|---|---|---|---|---|---|
| UTR3 | 233824 | 0.0093225 | 13 | 0.015795869 | 1.694381603 | 0.065857881 |
| UTR5 | 71546 | 0.00285252 | 3 | 0.0036452 | 1.277888192 | 0.511953737 |
| downstream | 284177 | 0.01133006 | 8 | 0.009720535 | 0.857942185 | 0.868279967 |
| exonic | 254736 | 0.01015625 | 8 | 0.009720535 | 0.957098471 | 1 |
| intergenic | 11684523 | 0.46585868 | 378 | 0.459295261 | 0.985911137 | 0.726822711 |
| intronic | 9137749 | 0.36431951 | 329 | 0.399756987 | 1.097270325 | 0.035680573 |
| ncRNA_exonic | 259951 | 0.01036417 | 2 | 0.002430134 | 0.234474417 | 0.015369465 |
| ncRNA_intronic | 2884355 | 0.11499843 | 79 | 0.095990279 | 0.83470947 | 0.090259384 |
| ncRNA_splicing | 1313 | 5.23E-05 | 0 | 0 | 0 | 1 |
| splicing | 2830 | 0.00011283 | 0 | 0 | 0 | 1 |
| upstream | 266686 | 0.0106327 | 3 | 0.0036452 | 0.342829352 | 0.057905561 |

* All SNPs in LD with the 32 independent significant SNPs with a functional annotation assigned by ANNOVAR were used

**Table S4 Roadmap epigenetic state enrichment analysis**

| Regulatory elements | total Bins | 1.00E-05 | 1.00E-06 | 1.00E-07 | P. 1E-5 | OR. 1E-5 | P. 1E-6 | OR. 1E-6 | P. 1E-7 | OR. 1E-7 |
|---|---|---|---|---|---|---|---|---|---|---|
| **Active enhancer states** | 2068209 | 129 | 34 | 10 | 2.96E-25 | 3.44887963 | 6.76E-12 | 5.87144643 | 0.04206038 | 2.6623966 |
| **Active promotor states** | 513015 | 31 | 15 | 0 | 4.86E-06 | 2.64845359 | 1.59E-08 | 7.71346187 | 0.63151025 | 0 |
| **All enhancer states** | 2133264 | 133 | 37 | 10 | 7.26E-26 | 3.48879309 | 1.35E-13 | 6.70804761 | 0.04206038 | 2.56830565 |
| **All promotor states** | 525189 | 31 | 15 | 0 | 8.36E-06 | 2.58494123 | 1.62E-08 | 7.5253286 | 0.63151025 | 0 |
| **Whole Genome** | 15282831 | 368 | 71 | 34 | | | | | | |

Table S5: Cytokine QTLs of COVID-19 GWAS loci from the 500 FG cohort study

| SNP* | chr | pos | variant_id | effect_allele | alternative_allele | beta-GWAS | standard_error-GWAS | p_value-GWAS | cytokine | beta-cQTL | pvalue-CQTL** |
|---|---|---|---|---|---|---|---|---|---|---|---|
| rs13063033 | 3 | 46040989 | chr3:46040989:G:A | A | G | 0.3555 | 0.0772 | 4.12E-06 | IL22_C.albicansconidia_PBMC_7days | -0.588665334 | 0.000602469 |
| rs9739956 | 12 | 62065731 | chr12:62065731:T:C | T | C | 0.2934 | 0.0598 | 9.45E-07 | IL22_B.burgdorferi_PBMC_7days | 0.404739861 | 0.000889539 |
| rs10877786 | 12 | 62070198 | chr12:62070198:T:G | T | G | 0.2964 | 0.0597 | 6.89E-07 | IL22_B.burgdorferi_PBMC_7days | 0.404611231 | 0.000893359 |
| rs74586549 | 3 | 45884551 | chr3:45884551:C:T | T | C | 0.3443 | 0.0768 | 7.34E-06 | IL22_C.albicansconidia_PBMC_7days | -0.571153018 | 0.000942702 |
| rs633862 | 9 | 133279871 | chr9:133279871:T:C | T | C | 0.2534 | 0.0515 | 8.79E-07 | TNFA_C.burnetiininemileSerum_PBMC_24h | 0.19300074 | 0.000967431 |
| rs660340 | 9 | 133272138 | chr9:133272138:G:A | A | G | -0.255 | 0.0517 | 7.97E-07 | TNFA_C.burnetiininemileSerum_PBMC_24h | -0.195176897 | 0.001009662 |
| rs581107 | 9 | 133272287 | chr9:133272287:T:C | T | C | 0.2553 | 0.0517 | 7.87E-07 | TNFA_C.burnetiininemileSerum_PBMC_24h | 0.195128539 | 0.001012258 |
| rs659104 | 9 | 133272408 | chr9:133272408:G:T | T | G | -0.255 | 0.0517 | 7.97E-07 | TNFA_C.burnetiininemileSerum_PBMC_24h | -0.195079446 | 0.001014882 |
| rs473533 | 9 | 133272620 | chr9:133272620:C:T | T | C | -0.2551 | 0.0517 | 7.89E-07 | TNFA_C.burnetiininemileSerum_PBMC_24h | -0.194991675 | 0.001020311 |
| rs475419 | 9 | 133272816 | chr9:133272816:T:C | T | C | 0.2576 | 0.0517 | 6.32E-07 | TNFA_C.burnetiininemileSerum_PBMC_24h | 0.194940739 | 0.001023012 |
| rs476410 | 9 | 133272953 | chr9:133272953:C:G | C | G | 0.2538 | 0.0517 | 8.96E-07 | TNFA_C.burnetiininemileSerum_PBMC_24h | 0.194889071 | 0.001025742 |
| rs645982 | 9 | 133272994 | chr9:133272994:G:A | A | G | -0.2536 | 0.0516 | 9.02E-07 | TNFA_C.burnetiininemileSerum_PBMC_24h | -0.194862962 | 0.001027118 |
| rs500498 | 9 | 133273232 | chr9:133273232:C:T | T | C | -0.2535 | 0.0516 | 9.06E-07 | TNFA_C.burnetiininemileSerum_PBMC_24h | -0.194793183 | 0.001030173 |
| rs500499 | 9 | 133273233 | chr9:133273233:C:G | C | G | 0.2535 | 0.0516 | 9.06E-07 | TNFA_C.burnetiininemileSerum_PBMC_24h | 0.194793183 | 0.001030173 |
| rs660340 | 9 | 133272138 | chr9:133272138:G:A | A | G | -0.255 | 0.0517 | 7.97E-07 | IL1b_C.burnetiininemileSerum_PBMC_24h | -0.270826347 | 0.001316142 |
| rs581107 | 9 | 133272287 | chr9:133272287:T:C | T | C | 0.2553 | 0.0517 | 7.87E-07 | IL1b_C.burnetiininemileSerum_PBMC_24h | 0.270705531 | 0.001322347 |
| rs659104 | 9 | 133272408 | chr9:133272408:G:T | T | G | -0.255 | 0.0517 | 7.97E-07 | IL1b_C.burnetiininemileSerum_PBMC_24h | -0.270583701 | 0.001328607 |
| rs473533 | 9 | 133272620 | chr9:133272620:C:T | T | C | -0.2551 | 0.0517 | 7.89E-07 | IL1b_C.burnetiininemileSerum_PBMC_24h | -0.270368448 | 0.001340629 |
| rs475419 | 9 | 133272816 | chr9:133272816:T:C | T | C | 0.2576 | 0.0517 | 6.32E-07 | IL1b_C.burnetiininemileSerum_PBMC_24h | 0.270244077 | 0.001347039 |
| rs476410 | 9 | 133272953 | chr9:133272953:C:G | C | G | 0.2538 | 0.0517 | 8.96E-07 | IL1b_C.burnetiininemileSerum_PBMC_24h | 0.270118698 | 0.001353507 |
| rs645982 | 9 | 133272994 | chr9:133272994:G:A | A | G | -0.2536 | 0.0516 | 9.02E-07 | IL1b_C.burnetiininemileSerum_PBMC_24h | -0.270055631 | 0.001356762 |
| rs500498 | 9 | 133273232 | chr9:133273232:C:T | T | C | -0.2535 | 0.0516 | 9.06E-07 | IL1b_C.burnetiininemileSerum_PBMC_24h | -0.269853067 | 0.001366636 |
| rs500499 | 9 | 133273233 | chr9:133273233:C:G | C | G | 0.2535 | 0.0516 | 9.06E-07 | IL1b_C.burnetiininemileSerum_PBMC_24h | 0.269853067 | 0.001366636 |
| rs633862 | 9 | 133279871 | chr9:133279871:T:C | T | C | 0.2534 | 0.0515 | 8.79E-07 | IL1b_C.burnetiininemileSerum_PBMC_24h | 0.263547123 | 0.001512703 |
| rs2230322 | 3 | 46021837 | chr3:46021837:T:C | T | C | -0.368 | 0.0768 | 1.68E-06 | IL22_C.albicansconidia_PBMC_7days | 0.559080107 | 0.001814071 |
| rs4362758 | 3 | 46015419 | chr3:46015419:T:A | A | T | 0.3681 | 0.0768 | 1.67E-06 | IL22_C.albicansconidia_PBMC_7days | -0.575914641 | 0.001828601 |
| rs35159820 | 3 | 46026981 | chr3:46026981:A:G | A | G | -0.3673 | 0.0769 | 1.76E-06 | IL22_C.albicansconidia_PBMC_7days | 0.561081238 | 0.001892859 |
| rs35161099 | 3 | 46026015 | chr3:46026015:T:G | T | G | -0.3625 | 0.0774 | 2.83E-06 | IL22_C.albicansconidia_PBMC_7days | 0.561036509 | 0.001892859 |
| rs71327010 | 3 | 46027343 | chr3:46027343:G:T | T | G | 0.3625 | 0.0774 | 2.83E-06 | IL22_C.albicansconidia_PBMC_7days | -0.561124527 | 0.001892885 |
| rs13090194 | 3 | 46028081 | chr3:46028081:A:C | A | C | -0.3672 | 0.0769 | 1.77E-06 | IL22_C.albicansconidia_PBMC_7days | 0.561206781 | 0.001893014 |
| rs59166269 | 3 | 46028845 | chr3:46028845:C:G | C | G | -0.3672 | 0.0769 | 1.77E-06 | IL22_C.albicansconidia_PBMC_7days | 0.561245745 | 0.001893118 |
| rs57437758 | 3 | 46028861 | chr3:46028861:A:C | A | C | -0.3672 | 0.0769 | 1.77E-06 | IL22_C.albicansconidia_PBMC_7days | 0.561245745 | 0.001893118 |
| rs35930050 | 3 | 46024271 | chr3:46024271:G:A | A | G | 0.3681 | 0.0768 | 1.67E-06 | IL22_C.albicansconidia_PBMC_7days | -0.560843215 | 0.001893119 |
| rs34584867 | 3 | 46024471 | chr3:46024471:G:A | A | G | 0.3681 | 0.0768 | 1.67E-06 | IL22_C.albicansconidia_PBMC_7days | -0.560843215 | 0.001893119 |
| rs34562820 | 3 | 46029589 | chr3:46029589:G:A | A | G | 0.3625 | 0.0775 | 2.87E-06 | IL22_C.albicansconidia_PBMC_7days | -0.561283264 | 0.001893248 |
| rs13082697 | 3 | 46030271 | chr3:46030271:T:C | T | C | -0.3671 | 0.0769 | 1.78E-06 | IL22_C.albicansconidia_PBMC_7days | 0.561319338 | 0.001893404 |
| rs13060287 | 3 | 46030376 | chr3:46030376:A:G | A | G | -0.3671 | 0.0769 | 1.79E-06 | IL22_C.albicansconidia_PBMC_7days | 0.561353967 | 0.001893586 |
| rs876668 | 3 | 46022261 | chr3:46022261:T:C | T | C | -0.368 | 0.0768 | 1.68E-06 | IL22_C.albicansconidia_PBMC_7days | 0.560626969 | 0.001893793 |
| rs34168660 | 3 | 46014670 | chr3:46014670:G:A | A | G | 0.375 | 0.0768 | 1.06E-06 | IL22_C.albicansconidia_PBMC_7days | -0.57391989 | 0.001901827 |
| rs34619093 | 3 | 46031027 | chr3:46031027:G:A | A | G | 0.3625 | 0.0775 | 2.89E-06 | IL22_C.albicansconidia_PBMC_7days | -0.561150268 | 0.001901837 |
| rs68087193 | 3 | 46031138 | chr3:46031138:C:T | T | C | 0.3673 | 0.0769 | 1.76E-06 | IL22_C.albicansconidia_PBMC_7days | -0.561150268 | 0.001901837 |
| rs2102055 | 3 | 46031212 | chr3:46031212:G:A | A | G | 0.3591 | 0.0767 | 2.83E-06 | IL22_C.albicansconidia_PBMC_7days | -0.561150268 | 0.001901837 |
| rs2102056 | 3 | 46031276 | chr3:46031276:G:A | A | G | 0.3671 | 0.0769 | 1.78E-06 | IL22_C.albicansconidia_PBMC_7days | -0.561150268 | 0.001901837 |
| rs2088690 | 3 | 46031347 | chr3:46031347:C:T | T | C | 0.3671 | 0.0769 | 1.78E-06 | IL22_C.albicansconidia_PBMC_7days | -0.561150268 | 0.001901837 |
| rs34774687 | 3 | 46031822 | chr3:46031822:T:C | T | C | -0.367 | 0.0769 | 1.80E-06 | IL22_C.albicansconidia_PBMC_7days | 0.561181975 | 0.001902072 |
| rs35814488 | 3 | 46032431 | chr3:46032431:A:G | A | G | -0.3672 | 0.0769 | 1.77E-06 | IL22_C.albicansconidia_PBMC_7days | 0.561212234 | 0.001902334 |
| rs6790866 | 3 | 46033219 | chr3:46033219:G:A | A | G | 0.367 | 0.0769 | 1.80E-06 | IL22_C.albicansconidia_PBMC_7days | -0.561241044 | 0.001902622 |
| rs6791016 | 3 | 46033366 | chr3:46033366:G:A | A | G | 0.367 | 0.0769 | 1.80E-06 | IL22_C.albicansconidia_PBMC_7days | -0.561268405 | 0.001902936 |
| rs6780028 | 3 | 46033598 | chr3:46033598:A:C | A | C | -0.367 | 0.0769 | 1.80E-06 | IL22_C.albicansconidia_PBMC_7days | 0.561268405 | 0.001902936 |
| rs35751180 | 3 | 46014681 | chr3:46014681:C:T | T | C | 0.3731 | 0.0774 | 1.42E-06 | IL22_C.albicansconidia_PBMC_7days | -0.573520315 | 0.001906391 |
| rs2088692 | 3 | 46035840 | chr3:46035840:A:G | A | G | -0.3654 | 0.0768 | 1.96E-06 | IL22_C.albicansconidia_PBMC_7days | 0.561116263 | 0.001912687 |
| rs34438204 | 3 | 46039814 | chr3:46039814:T:C | T | C | -0.3609 | 0.0774 | 3.13E-06 | IL22_C.albicansconidia_PBMC_7days | 0.561236412 | 0.001916194 |
| rs34679077 | 3 | 46046500 | chr3:46046500:G:A | A | G | 0.3609 | 0.0774 | 3.13E-06 | IL22_C.albicansconidia_PBMC_7days | -0.561042217 | 0.001932537 |
| rs13081213 | 3 | 46014417 | chr3:46014417:C:T | T | C | 0.3746 | 0.0768 | 1.08E-06 | IL22_C.albicansconidia_PBMC_7days | -0.573045954 | 0.001935398 |
| rs13081151 | 3 | 46014224 | chr3:46014224:G:A | A | G | 0.3751 | 0.0777 | 1.38E-06 | IL22_C.albicansconidia_PBMC_7days | -0.572708115 | 0.001947995 |
| rs13097556 | 3 | 46020505 | chr3:46020505:T:C | T | C | -0.3679 | 0.0768 | 1.69E-06 | IL22_C.albicansconidia_PBMC_7days | 0.559578702 | 0.001948125 |
| rs34863575 | 3 | 46052571 | chr3:46052571:A:G | A | G | -0.3543 | 0.0773 | 4.53E-06 | IL22_C.albicansconidia_PBMC_7days | 0.560733863 | 0.001953008 |
| rs35772789 | 3 | 46053612 | chr3:46053612:A:G | A | G | -0.3544 | 0.0773 | 4.52E-06 | IL22_C.albicansconidia_PBMC_7days | 0.560601039 | 0.001958772 |
| rs13074382 | 3 | 46020348 | chr3:46020348:A:G | A | G | -0.3679 | 0.0768 | 1.69E-06 | IL22_C.albicansconidia_PBMC_7days | 0.559344377 | 0.001958873 |
| rs13093179 | 3 | 46104822 | chr3:46104822:G:T | T | G | 0.3557 | 0.0779 | 4.91E-06 | IL22_C.albicansconidia_PBMC_7days | -0.565478533 | 0.001961131 |
| rs647800 | 9 | 133272585 | chr9:133272585:G:A | A | G | -0.2683 | 0.052 | 2.41E-07 | TNFA_C.burnetiininemileSerum_PBMC_24h | -0.183643848 | 0.001969629 |
| rs34340587 | 3 | 46054551 | chr3:46054551:G:A | A | G | 0.3542 | 0.0773 | 4.63E-06 | IL22_C.albicansconidia_PBMC_7days | -0.560258148 | 0.001972332 |
| rs77399277 | 3 | 46055306 | chr3:46055306:T:C | T | C | -0.3556 | 0.0773 | 4.25E-06 | IL22_C.albicansconidia_PBMC_7days | 0.559933905 | 0.001983987 |
| rs36040135 | 3 | 46019812 | chr3:46019812:A:G | A | G | -0.3679 | 0.0768 | 1.69E-06 | IL22_C.albicansconidia_PBMC_7days | 0.558757675 | 0.001987493 |
| rs34718164 | 3 | 46057034 | chr3:46057034:C:G | C | G | -0.3556 | 0.0773 | 4.25E-06 | IL22_C.albicansconidia_PBMC_7days | 0.559594189 | 0.001996244 |
| rs13096741 | 3 | 46057662 | chr3:46057662:C:T | T | C | 0.3556 | 0.0773 | 4.27E-06 | IL22_C.albicansconidia_PBMC_7days | -0.559484023 | 0.002000005 |
| rs35334665 | 3 | 46019726 | chr3:46019726:T:C | T | C | -0.3632 | 0.0774 | 2.70E-06 | IL22_C.albicansconidia_PBMC_7days | 0.558294585 | 0.002006736 |
| rs35420565 | 3 | 46058646 | chr3:46058646:C:G | C | G | -0.3556 | 0.0773 | 4.26E-06 | IL22_C.albicansconidia_PBMC_7days | 0.559246246 | 0.002007877 |
| rs34047915 | 3 | 46058999 | chr3:46058999:C:T | T | C | 0.3555 | 0.0773 | 4.29E-06 | IL22_C.albicansconidia_PBMC_7days | -0.559183171 | 0.002009918 |
| rs71327014 | 3 | 46059974 | chr3:46059974:C:T | T | C | 0.3556 | 0.0773 | 4.25E-06 | IL22_C.albicansconidia_PBMC_7days | -0.559118644 | 0.002011988 |
| rs71327015 | 3 | 46060681 | chr3:46060681:G:C | C | G | 0.3585 | 0.0773 | 3.57E-06 | IL22_C.albicansconidia_PBMC_7days | -0.559118644 | 0.002011988 |
| rs35669129 | 3 | 46013758 | chr3:46013758:G:A | A | G | 0.3746 | 0.0768 | 1.08E-06 | IL22_C.albicansconidia_PBMC_7days | -0.570873778 | 0.002018753 |
| rs35516580 | 3 | 46066006 | chr3:46066006:A:G | A | G | -0.3562 | 0.0773 | 4.13E-06 | IL22_C.albicansconidia_PBMC_7days | 0.558877142 | 0.002020576 |
| rs34766614 | 3 | 46066109 | chr3:46066109:A:G | A | G | -0.3484 | 0.0775 | 6.99E-06 | IL22_C.albicansconidia_PBMC_7days | 0.558877142 | 0.002020576 |
| rs34127208 | 3 | 46062188 | chr3:46062188:G:T | T | G | 0.3554 | 0.0773 | 4.31E-06 | IL22_C.albicansconidia_PBMC_7days | -0.558647621 | 0.002029788 |
| rs35454877 | 3 | 46017992 | chr3:46017992:T:C | T | C | -0.3679 | 0.0768 | 1.69E-06 | IL22_C.albicansconidia_PBMC_7days | 0.556279223 | 0.002102441 |
| rs71327007 | 3 | 46017757 | chr3:46017757:C:T | T | C | 0.3643 | 0.0774 | 2.51E-06 | IL22_C.albicansconidia_PBMC_7days | -0.555996498 | 0.002114927 |
| rs71327006 | 3 | 46017416 | chr3:46017416:G:A | A | G | 0.3628 | 0.0774 | 2.77E-06 | IL22_C.albicansconidia_PBMC_7days | -0.555227598 | 0.002148424 |
| rs1491951 | 3 | 46100352 | chr3:46100352:G:A | A | G | 0.3485 | 0.0777 | 7.30E-06 | IL22_C.albicansconidia_PBMC_7days | -0.545243213 | 0.00224495 |
| rs35539222 | 3 | 46112780 | chr3:46112780:T:C | T | C | -0.3556 | 0.0779 | 4.96E-06 | IL22_C.albicansconidia_PBMC_7days | 0.551595284 | 0.00229677 |
| rs35110864 | 3 | 46112965 | chr3:46112965:G:A | A | G | 0.3555 | 0.0779 | 4.99E-06 | IL22_C.albicansconidia_PBMC_7days | -0.551595284 | 0.00229677 |
| rs71327025 | 3 | 46106012 | chr3:46106012:A:G | A | G | -0.3556 | 0.0779 | 4.96E-06 | IL22_C.albicansconidia_PBMC_7days | 0.550934758 | 0.002314809 |
| rs34093271 | 3 | 46103489 | chr3:46103489:G:T | T | G | 0.3558 | 0.0779 | 4.89E-06 | IL22_C.albicansconidia_PBMC_7days | -0.546275924 | 0.002460276 |
| rs34452002 | 3 | 46101695 | chr3:46101695:C:T | T | C | 0.3556 | 0.0779 | 4.95E-06 | IL22_C.albicansconidia_PBMC_7days | -0.545733099 | 0.002474056 |
| rs13075758 | 3 | 45983556 | chr3:45983556:G:A | A | G | 0.3672 | 0.0775 | 2.19E-06 | IL22_C.albicansconidia_PBMC_7days | -0.560676768 | 0.002474993 |
| rs34442130 | 3 | 45983037 | chr3:45983037:T:A | A | T | 0.3674 | 0.0775 | 2.16E-06 | IL22_C.albicansconidia_PBMC_7days | -0.560695623 | 0.002475623 |
| rs34460587 | 3 | 46100972 | chr3:46100972:C:T | T | C | 0.3557 | 0.0779 | 4.93E-06 | IL22_C.albicansconidia_PBMC_7days | -0.545386348 | 0.002478423 |
| rs13091868 | 3 | 46081551 | chr3:46081551:A:G | A | G | -0.3538 | 0.0773 | 4.72E-06 | IL22_C.albicansconidia_PBMC_7days | 0.544813335 | 0.002488023 |
| rs13092030 | 3 | 46081563 | chr3:46081563:C:T | T | C | 0.354 | 0.0773 | 4.66E-06 | IL22_C.albicansconidia_PBMC_7days | -0.544813335 | 0.002488023 |
| rs17215981 | 3 | 46082926 | chr3:46082926:A:G | A | G | -0.3547 | 0.0773 | 4.47E-06 | IL22_C.albicansconidia_PBMC_7days | 0.544813335 | 0.002488023 |
| rs13089554 | 3 | 46086065 | chr3:46086065:G:A | A | G | 0.3547 | 0.0774 | 4.61E-06 | IL22_C.albicansconidia_PBMC_7days | -0.544813335 | 0.002488023 |
| rs13095717 | 3 | 46087050 | chr3:46087050:G:A | A | G | 0.3548 | 0.0774 | 4.58E-06 | IL22_C.albicansconidia_PBMC_7days | -0.544813335 | 0.002488023 |
| rs13075528 | 3 | 46087217 | chr3:46087217:T:C | T | C | -0.3549 | 0.0775 | 4.63E-06 | IL22_C.albicansconidia_PBMC_7days | 0.544813335 | 0.002488023 |
| rs13095602 | 3 | 46087221 | chr3:46087221:A:G | A | G | -0.3547 | 0.0774 | 4.60E-06 | IL22_C.albicansconidia_PBMC_7days | 0.544813335 | 0.002488023 |
| rs71327023 | 3 | 46089733 | chr3:46089733:C:G | C | G | -0.3539 | 0.0777 | 5.19E-06 | IL22_C.albicansconidia_PBMC_7days | 0.544813335 | 0.002488023 |
| rs13063527 | 3 | 46094198 | chr3:46094198:G:A | A | G | 0.3561 | 0.0777 | 4.51E-06 | IL22_C.albicansconidia_PBMC_7days | -0.544813335 | 0.002488023 |
| rs13068570 | 3 | 46094946 | chr3:46094946:A:G | A | G | -0.3561 | 0.0777 | 4.61E-06 | IL22_C.albicansconidia_PBMC_7days | 0.544813335 | 0.002488023 |
| rs4373103 | 3 | 46096963 | chr3:46096963:G:A | A | G | 0.3547 | 0.078 | 5.42E-06 | IL22_C.albicansconidia_PBMC_7days | -0.544813335 | 0.002488023 |
| rs71327024 | 3 | 46098581 | chr3:46098581:G:T | T | G | 0.3547 | 0.078 | 5.37E-06 | IL22_C.albicansconidia_PBMC_7days | -0.544813335 | 0.002488023 |
| rs36122610 | 3 | 45981341 | chr3:45981341:G:A | A | G | 0.3673 | 0.0775 | 2.18E-06 | IL22_C.albicansconidia_PBMC_7days | -0.560186614 | 0.002498444 |
| rs71327017 | 3 | 46069721 | chr3:46069721:T:C | T | C | -0.3576 | 0.0774 | 3.80E-06 | IL22_C.albicansconidia_PBMC_7days | 0.544589858 | 0.002499018 |
| rs13072267 | 3 | 46074518 | chr3:46074518:T:A | A | T | 0.3543 | 0.0773 | 4.57E-06 | IL22_C.albicansconidia_PBMC_7days | -0.544589858 | 0.002499018 |
| rs36057789 | 3 | 46075604 | chr3:46075604:A:G | A | G | -0.3551 | 0.0773 | 4.36E-06 | IL22_C.albicansconidia_PBMC_7days | 0.544589858 | 0.002499018 |
| rs76647202 | 3 | 46076468 | chr3:46076468:A:G | A | G | -0.3542 | 0.0773 | 4.60E-06 | IL22_C.albicansconidia_PBMC_7days | 0.544589858 | 0.002499018 |
| rs71327021 | 3 | 46079025 | chr3:46079025:A:G | A | G | -0.3542 | 0.0773 | 4.61E-06 | IL22_C.albicansconidia_PBMC_7days | 0.544589858 | 0.002499018 |
| rs1491950 | 3 | 46096615 | chr3:46096615:G:A | A | G | 0.3544 | 0.078 | 5.55E-06 | IL22_C.albicansconidia_PBMC_7days | -0.536881521 | 0.002822749 |
| rs17330872 | 3 | 45993605 | chr3:45993605:A:G | A | G | -0.3702 | 0.0774 | 1.74E-06 | IL22_C.albicansconidia_PBMC_7days | 0.553990152 | 0.002848216 |
| rs36023124 | 3 | 45997568 | chr3:45997568:G:C | C | G | 0.3665 | 0.0771 | 2.01E-06 | IL22_C.albicansconidia_PBMC_7days | -0.55044912 | 0.003014781 |
| rs73833520 | 3 | 46016375 | chr3:46016375:A:G | A | G | -0.3679 | 0.0768 | 1.69E-06 | IL22_C.albicansconidia_PBMC_7days | 0.540628427 | 0.003038221 |
| rs71615438 | 3 | 45998376 | chr3:45998376:A:G | A | G | -0.3666 | 0.0772 | 2.04E-06 | IL22_C.albicansconidia_PBMC_7days | 0.549526464 | 0.003058006 |
| rs34492478 | 3 | 46097350 | chr3:46097350:A:T | A | T | -0.3562 | 0.078 | 4.95E-06 | IL22_C.albicansconidia_PBMC_7days | 0.533523963 | 0.003132196 |
| rs34754340 | 3 | 46000345 | chr3:46000345:C:T | T | C | 0.3673 | 0.0772 | 1.94E-06 | IL22_C.albicansconidia_PBMC_7days | -0.547472361 | 0.00315361 |
| rs34836513 | 3 | 46004967 | chr3:46004967:G:A | A | G | 0.3674 | 0.0772 | 1.96E-06 | IL22_C.albicansconidia_PBMC_7days | -0.542341868 | 0.003395566 |
| rs10877787 | 12 | 62072981 | chr12:62072981:A:G | A | G | 0.29 | 0.0604 | 1.58E-06 | IL22_B.burgdorferi_PBMC_7days | 0.357687822 | 0.003474381 |
| rs9706516 | 12 | 62074403 | chr12:62074403:C:A | A | C | -0.2923 | 0.0604 | 1.30E-06 | IL22_B.burgdorferi_PBMC_7days | -0.357653548 | 0.003485897 |
| rs10506432 | 12 | 62077116 | chr12:62077116:A:C | A | C | 0.29 | 0.0604 | 1.58E-06 | IL22_B.burgdorferi_PBMC_7days | 0.35778809 | 0.003490773 |
| rs10747951 | 12 | 62075677 | chr12:62075677:C:T | T | C | -0.29 | 0.0604 | 1.58E-06 | IL22_B.burgdorferi_PBMC_7days | -0.357692945 | 0.003491101 |
| rs11174296 | 12 | 62077677 | chr12:62077677:C:G | C | G | 0.288 | 0.0602 | 1.75E-06 | IL22_B.burgdorferi_PBMC_7days | 0.357639883 | 0.003506854 |
| rs9738190 | 12 | 62078790 | chr12:62078790:G:A | A | G | -0.2898 | 0.0602 | 1.51E-06 | IL22_B.burgdorferi_PBMC_7days | -0.357610001 | 0.003525605 |
| rs11174294 | 12 | 62072129 | chr12:62072129:C:T | T | C | -0.29 | 0.0604 | 1.59E-06 | IL22_B.burgdorferi_PBMC_7days | -0.356678201 | 0.003566548 |
| rs10784279 | 12 | 62044016 | chr12:62044016:C:T | T | C | -0.2778 | 0.0599 | 3.50E-06 | IL22_B.burgdorferi_PBMC_7days | -0.352066079 | 0.004104088 |
| rs10784278 | 12 | 62042165 | chr12:62042165:T:A | A | T | -0.2836 | 0.06 | 2.32E-06 | IL22_B.burgdorferi_PBMC_7days | -0.351968607 | 0.004117457 |
| rs9739464 | 12 | 62037992 | chr12:62037992:A:T | A | T | 0.2846 | 0.06 | 2.10E-06 | IL22_B.burgdorferi_PBMC_7days | 0.351753989 | 0.004186996 |
| rs10506436 | 12 | 62028786 | chr12:62028786:T:A | A | T | -0.2811 | 0.06 | 2.84E-06 | IL22_B.burgdorferi_PBMC_7days | -0.348771886 | 0.004492644 |
| rs11627388 | 14 | 31953140 | chr14:31953140:A:C | A | C | -0.2482 | 0.0533 | 3.19E-06 | IL6_Borreliamix_PBMC_24h | -0.260397096 | 0.004802014 |
| rs1455664 | 4 | 179425171 | chr4:179425171:T:C | T | C | -0.3158 | 0.0714 | 9.77E-06 | IL22_B.burgdorferi_PBMC_7days | 0.416777203 | 0.005036079 |
| rs1455662 | 4 | 179423719 | chr4:179423719:A:G | A | G | -0.3156 | 0.0713 | 9.60E-06 | IL22_B.burgdorferi_PBMC_7days | 0.414968838 | 0.005175043 |
| rs1455663 | 4 | 179425076 | chr4:179425076:A:G | A | G | -0.3156 | 0.0714 | 9.87E-06 | IL22_B.burgdorferi_PBMC_7days | 0.41640833 | 0.005204446 |
| rs647800 | 9 | 133272585 | chr9:133272585:G:A | A | G | -0.2683 | 0.052 | 2.41E-07 | IL1b_C.burnetiininemileSerum_PBMC_24h | -0.232372393 | 0.005871546 |
| rs2059266 | 16 | 74902133 | chr16:74902133:A:G | A | G | -0.2457 | 0.0533 | 4.01E-06 | IL6_A.fumigatusconidiaSerum_PBMC_24h | 0.286273283 | 0.005993184 |
| rs2191031 | 3 | 45869378 | chr3:45869378:G:A | A | G | 0.2758 | 0.0613 | 6.80E-06 | IL6_C.burnetiininemileSerum_PBMC_24h | -0.196641887 | 0.007396957 |
| rs41264195 | 2 | 15606096 | chr2:15606096:C:T | T | C | -0.3359 | 0.0754 | 8.49E-06 | IL1b_MTB_PBMC_24h | 0.347253962 | 0.007413472 |
| rs79808866 | 2 | 15604860 | chr2:15604860:T:G | T | G | 0.3357 | 0.0754 | 8.59E-06 | IL1b_MTB_PBMC_24h | -0.347011707 | 0.007454685 |
| rs3770462 | 2 | 15609200 | chr2:15609200:T:G | T | G | 0.3409 | 0.0757 | 6.74E-06 | IL1b_MTB_PBMC_24h | -0.346783267 | 0.00748967 |
| rs4668941 | 2 | 15610007 | chr2:15610007:T:G | T | G | 0.3359 | 0.0754 | 8.51E-06 | IL1b_MTB_PBMC_24h | -0.346783267 | 0.00748967 |
| rs3774641 | 3 | 45896341 | chr3:45896341:G:T | T | G | 0.2741 | 0.0605 | 5.95E-06 | IL6_C.burnetiininemileSerum_PBMC_24h | -0.18727043 | 0.009301355 |
| rs76860840 | 2 | 15618674 | chr2:15618674:C:G | C | G | 0.3536 | 0.0764 | 3.66E-06 | IL1b_MTB_PBMC_24h | -0.340876768 | 0.009443143 |
| rs4797120 | 18 | 3725189 | chr18:3725189:G:C | C | G | -0.2956 | 0.0621 | 1.91E-06 | IFNy_A.fumigatusconidia_PBMC_7days | -0.336828996 | 0.010289777 |
| rs10088118 | 8 | 121806381 | chr8:121806381:G:C | C | G | -0.2329 | 0.0517 | 6.79E-06 | TNFA_Cryptococcus_PBMC_24h | 0.4104619 | 0.010435232 |
| rs2302929 | 2 | 15595524 | chr2:15595524:T:C | T | C | 0.2976 | 0.0672 | 9.54E-06 | IFNy_C.albicanshyphae_PBMC_7days | 0.412272501 | 0.010450043 |
| rs4389947 | 8 | 121816361 | chr8:121816361:G:A | A | G | -0.2364 | 0.0517 | 4.91E-06 | TNFA_Cryptococcus_PBMC_24h | 0.409287571 | 0.010630089 |
| rs10091098 | 8 | 121836590 | chr8:121836590:G:A | A | G | -0.233 | 0.0508 | 4.55E-06 | TNFA_Cryptococcus_PBMC_24h | 0.408212042 | 0.010817879 |
| rs10087789 | 8 | 121836324 | chr8:121836324:C:G | C | G | 0.2327 | 0.0518 | 7.00E-06 | TNFA_Cryptococcus_PBMC_24h | -0.40813143 | 0.010834382 |
| rs2059266 | 16 | 74902133 | chr16:74902133:A:G | A | G | -0.2457 | 0.0533 | 4.01E-06 | IFNy_MTB_PBMC_7days | -0.292326806 | 0.010919981 |
| rs550057 | 9 | 133271182 | chr9:133271182:T:C | T | C | 0.2681 | 0.0562 | 1.86E-06 | IL6_MSUC16_PBMC_24h | 0.243010022 | 0.011239002 |
| rs2302934 | 2 | 15613155 | chr2:15613155:C:G | C | G | 0.3393 | 0.076 | 8.11E-06 | IL1b_MTB_PBMC_24h | -0.332726853 | 0.011660372 |
| rs12616567 | 2 | 15615498 | chr2:15615498:A:C | A | C | 0.3427 | 0.0762 | 6.85E-06 | IL1b_MTB_PBMC_24h | -0.332726853 | 0.011660372 |
| rs74586549 | 3 | 45884551 | chr3:45884551:C:T | T | C | 0.3443 | 0.0768 | 7.34E-06 | IL22_C.albicanshyphae_PBMC_7days | -0.453315197 | 0.012126756 |
| rs807635 | 2 | 15629483 | chr2:15629483:C:T | T | C | -0.3579 | 0.0765 | 2.85E-06 | IL1b_MTB_PBMC_24h | 0.329706349 | 0.012147109 |
| rs7152677 | 14 | 31953540 | chr14:31953540:G:C | C | G | 0.2637 | 0.0548 | 1.52E-06 | IL6_Borreliamix_PBMC_24h | 0.243828758 | 0.012162277 |
| rs11627388 | 14 | 31953140 | chr14:31953140:A:C | A | C | -0.2482 | 0.0533 | 3.19E-06 | TNFA_MSUC16_PBMC_24h | -0.196502582 | 0.012174531 |
| rs807636 | 2 | 15627828 | chr2:15627828:C:T | T | C | -0.365 | 0.0772 | 2.25E-06 | IL1b_MTB_PBMC_24h | 0.327159629 | 0.012947455 |
| rs10088118 | 8 | 121806381 | chr8:121806381:G:C | C | G | -0.2329 | 0.0517 | 6.79E-06 | IL1b_Cryptococcus_PBMC_24h | 0.430210431 | 0.014697652 |
| rs4389947 | 8 | 121816361 | chr8:121816361:G:A | A | G | -0.2364 | 0.0517 | 4.91E-06 | IL1b_Cryptococcus_PBMC_24h | 0.429216578 | 0.01489285 |
| rs10091098 | 8 | 121836590 | chr8:121836590:G:A | A | G | -0.233 | 0.0508 | 4.55E-06 | IL1b_Cryptococcus_PBMC_24h | 0.428048399 | 0.015143372 |
| rs10087789 | 8 | 121836324 | chr8:121836324:C:G | C | G | 0.2327 | 0.0518 | 7.00E-06 | IL1b_Cryptococcus_PBMC_24h | -0.427952648 | 0.015167218 |
| rs12610495 | 19 | 4717660 | chr19:4717660:A:G | A | G | -0.2534 | 0.0556 | 5.20E-06 | IFNy_S.aureus_PBMC_7days | -0.285517699 | 0.016075367 |
| rs13063033 | 3 | 46040989 | chr3:46040989:G:A | A | G | 0.3555 | 0.0772 | 4.12E-06 | IL22_A.fumigatusconidia_PBMC_7days | 0.449241111 | 0.016222315 |
| rs12619874 | 2 | 15586997 | chr2:15586997:G:C | C | G | -0.3204 | 0.0665 | 1.46E-06 | IFNy_C.albicanshyphae_PBMC_7days | -0.378972389 | 0.016236963 |
| rs36122610 | 3 | 45981341 | chr3:45981341:G:A | A | G | 0.3673 | 0.0775 | 2.18E-06 | IL6_LPS100ng_PBMC_24h | -0.246349673 | 0.016412853 |
| rs34442130 | 3 | 45983037 | chr3:45983037:T:A | A | T | 0.3674 | 0.0775 | 2.16E-06 | IL6_LPS100ng_PBMC_24h | -0.246295606 | 0.016436759 |
| rs13075758 | 3 | 45983556 | chr3:45983556:G:A | A | G | 0.3672 | 0.0775 | 2.19E-06 | IL6_LPS100ng_PBMC_24h | -0.246270068 | 0.016441633 |
| rs13381043 | 18 | 3724602 | chr18:3724602:G:A | A | G | -0.325 | 0.0613 | 1.13E-07 | IL1b_MSUC16_PBMC_24h | -0.265837818 | 0.017162197 |
| rs13381043 | 18 | 3724602 | chr18:3724602:G:A | A | G | -0.325 | 0.0613 | 1.13E-07 | IL6_MSUC16_PBMC_24h | -0.245593079 | 0.017293966 |
| rs550057 | 9 | 133271182 | chr9:133271182:T:C | T | C | 0.2681 | 0.0562 | 1.86E-06 | TNFA_S.aureus_PBMC_24h | 0.253264567 | 0.017475876 |
| rs807577 | 2 | 15664270 | chr2:15664270:C:G | C | G | 0.3181 | 0.0704 | 6.28E-06 | IFNy_Cryptococcus_PBMC_7days | -0.304988357 | 0.017930364 |
| rs12619874 | 2 | 15586997 | chr2:15586997:G:C | C | G | -0.3204 | 0.0665 | 1.46E-06 | TNFA_S.aureus_PBMC_24h | -0.305557944 | 0.017931009 |
| rs807615 | 2 | 15648378 | chr2:15648378:A:G | A | G | 0.309 | 0.068 | 5.51E-06 | IFNy_Cryptococcus_PBMC_7days | -0.303170064 | 0.018002902 |
| rs807613 | 2 | 15649443 | chr2:15649443:C:T | T | C | -0.3086 | 0.0679 | 5.60E-06 | IFNy_Cryptococcus_PBMC_7days | 0.30250602 | 0.018283847 |
| rs807590 | 2 | 15657801 | chr2:15657801:G:T | T | G | -0.3146 | 0.0696 | 6.11E-06 | IFNy_Cryptococcus_PBMC_7days | 0.304758059 | 0.018314779 |
| rs807611 | 2 | 15649857 | chr2:15649857:G:T | T | G | -0.3084 | 0.0672 | 4.51E-06 | IFNy_Cryptococcus_PBMC_7days | 0.302292012 | 0.018377032 |
| rs807587 | 2 | 15659856 | chr2:15659856:C:G | C | G | 0.3246 | 0.0702 | 3.76E-06 | IFNy_Cryptococcus_PBMC_7days | -0.303970795 | 0.018399149 |
| rs807586 | 2 | 15660004 | chr2:15660004:T:C | T | C | 0.3125 | 0.0703 | 8.76E-06 | IFNy_Cryptococcus_PBMC_7days | -0.303970795 | 0.018399149 |
| rs807583 | 2 | 15660597 | chr2:15660597:G:A | A | G | -0.3121 | 0.0703 | 9.01E-06 | IFNy_Cryptococcus_PBMC_7days | 0.303970795 | 0.018399149 |
| rs807579 | 2 | 15663460 | chr2:15663460:T:C | T | C | 0.3124 | 0.0703 | 8.88E-06 | IFNy_Cryptococcus_PBMC_7days | -0.303970795 | 0.018399149 |
| rs807578 | 2 | 15663516 | chr2:15663516:G:C | C | G | -0.3124 | 0.0703 | 8.81E-06 | IFNy_Cryptococcus_PBMC_7days | 0.303970795 | 0.018399149 |
| rs807589 | 2 | 15658661 | chr2:15658661:C:A | A | C | -0.326 | 0.0702 | 3.48E-06 | IFNy_Cryptococcus_PBMC_7days | 0.303930465 | 0.01846398 |
| rs12185539 | 2 | 15650052 | chr2:15650052:A:G | A | G | 0.3079 | 0.067 | 4.31E-06 | IFNy_Cryptococcus_PBMC_7days | -0.301754822 | 0.018603847 |
| rs12185540 | 2 | 15650056 | chr2:15650056:A:G | A | G | 0.3058 | 0.0677 | 6.19E-06 | IFNy_Cryptococcus_PBMC_7days | -0.301754822 | 0.018603847 |
| rs3774641 | 3 | 45896341 | chr3:45896341:G:T | T | G | 0.2741 | 0.0605 | 5.95E-06 | TNFA_C.albicanshyphae_PBMC_24h | -0.291622518 | 0.019052037 |
| rs807610 | 2 | 15650440 | chr2:15650440:C:G | C | G | 0.3082 | 0.0673 | 4.64E-06 | IFNy_Cryptococcus_PBMC_7days | -0.300674126 | 0.019056122 |
| rs76880770 | 2 | 15618488 | chr2:15618488:G:A | A | G | -0.3455 | 0.0767 | 6.62E-06 | IL1b_MTB_PBMC_24h | 0.324307215 | 0.019373464 |
| rs12610495 | 19 | 4717660 | chr19:4717660:A:G | A | G | -0.2534 | 0.0556 | 5.20E-06 | IL1b_Influenza_PBMC_24h | 0.399352327 | 0.020013466 |
| rs807607 | 2 | 15651223 | chr2:15651223:G:C | C | G | -0.3087 | 0.0679 | 5.52E-06 | IFNy_Cryptococcus_PBMC_7days | 0.297963669 | 0.02018846 |
| rs17330872 | 3 | 45993605 | chr3:45993605:A:G | A | G | -0.3702 | 0.0774 | 1.74E-06 | IL6_LPS100ng_PBMC_24h | 0.238873635 | 0.020254553 |
| rs2191031 | 3 | 45869378 | chr3:45869378:G:A | A | G | 0.2758 | 0.0613 | 6.80E-06 | TNFA_C.albicanshyphae_PBMC_24h | -0.293944857 | 0.020504578 |
| rs74586549 | 3 | 45884551 | chr3:45884551:C:T | T | C | 0.3443 | 0.0768 | 7.34E-06 | IFNy_A.fumigatusconidia_PBMC_7days | 0.388900192 | 0.020520136 |
| rs807576 | 2 | 15664332 | chr2:15664332:T:C | T | C | 0.3185 | 0.0704 | 6.15E-06 | IFNy_Cryptococcus_PBMC_7days | -0.296307965 | 0.020673276 |
| rs807606 | 2 | 15651602 | chr2:15651602:C:A | A | C | -0.3042 | 0.0679 | 7.55E-06 | IFNy_Cryptococcus_PBMC_7days | 0.296465222 | 0.020787171 |
| rs550057 | 9 | 133271182 | chr9:133271182:T:C | T | C | 0.2681 | 0.0562 | 1.86E-06 | IL6_LPS1ng_PBMC_24h | 0.329312252 | 0.020930981 |
| rs9739464 | 12 | 62037992 | chr12:62037992:A:T | A | T | 0.2846 | 0.06 | 2.10E-06 | IL22_C.albicansconidia_PBMC_7days | 0.322027108 | 0.020941805 |
| rs10784279 | 12 | 62044016 | chr12:62044016:C:T | T | C | -0.2778 | 0.0599 | 3.50E-06 | IL22_C.albicansconidia_PBMC_7days | -0.321292761 | 0.021069012 |
| rs10784278 | 12 | 62042165 | chr12:62042165:T:A | A | T | -0.2836 | 0.06 | 2.32E-06 | IL22_C.albicansconidia_PBMC_7days | -0.321249122 | 0.021096708 |
| rs807623 | 2 | 15642467 | chr2:15642467:C:T | T | C | -0.312 | 0.0679 | 4.30E-06 | IFNy_Cryptococcus_PBMC_7days | 0.291446011 | 0.02119158 |
| rs807622 | 2 | 15642631 | chr2:15642631:A:G | A | G | 0.3099 | 0.0675 | 4.43E-06 | IFNy_Cryptococcus_PBMC_7days | -0.291446011 | 0.02119158 |
| rs807620 | 2 | 15643012 | chr2:15643012:C:T | T | C | -0.312 | 0.0679 | 4.30E-06 | IFNy_Cryptococcus_PBMC_7days | 0.291446011 | 0.02119158 |
| rs807605 | 2 | 15652128 | chr2:15652128:G:A | A | G | -0.3007 | 0.0679 | 9.38E-06 | IFNy_Cryptococcus_PBMC_7days | 0.295397603 | 0.02122353 |

| | | | | | | | | | | | |
|---|---|---|---|---|---|---|---|---|---|---|---|
| rs36023124 | 3 | 45997568 | chr3:45997568:G:C | C | G | 0.3665 | 0.0771 | 2.01E-06 | IL6_LPS100ng_PBMC_24h | -0.236859203 | 0.021260946 |
| rs807602 | 2 | 15652485 | chr2:15652485:C:A | A | C | -0.3021 | 0.0679 | 8.60E-06 | IFNy_Cryptococcus_PBMC_7days | 0.295297149 | 0.02126677 |
| rs807604 | 2 | 15652384 | chr2:15652384:T:A | A | T | -0.3037 | 0.0679 | 7.64E-06 | IFNy_Cryptococcus_PBMC_7days | 0.295269716 | 0.021276414 |
| rs62120724 | 2 | 15593845 | chr2:15593845:G:A | A | G | -0.2986 | 0.0675 | 9.73E-06 | IFNy_C.albicanshyphae_PBMC_7days | -0.376600851 | 0.021336543 |
| rs2032778 | 2 | 15596573 | chr2:15596573:G:A | A | G | -0.299 | 0.0675 | 9.48E-06 | IFNy_C.albicanshyphae_PBMC_7days | -0.376600851 | 0.021336543 |
| rs71615438 | 3 | 45998376 | chr3:45998376:A:G | A | G | -0.3666 | 0.0772 | 2.04E-06 | IL6_LPS100ng_PBMC_24h | 0.236346008 | 0.021516651 |
| rs13263570 | 8 | 121845942 | chr8:121845942:G:A | A | G | -0.2298 | 0.0516 | 8.50E-06 | TNFA_Cryptococcus_PBMC_24h | 0.365237884 | 0.021619567 |
| rs807618 | 2 | 15646936 | chr2:15646936:G:C | C | G | -0.3046 | 0.0673 | 6.02E-06 | IFNy_Cryptococcus_PBMC_7days | 0.290174709 | 0.021753714 |
| rs807595 | 2 | 15654550 | chr2:15654550:C:T | T | C | -0.3019 | 0.0682 | 9.52E-06 | IFNy_Cryptococcus_PBMC_7days | 0.294625421 | 0.02183808 |
| rs2059266 | 16 | 74902133 | chr16:74902133:A:G | A | G | -0.2457 | 0.0533 | 4.01E-06 | IFNy_B.burgdorferi_PBMC_7days | -0.193117577 | 0.021868037 |
| rs807596 | 2 | 15654457 | chr2:15654457:G:A | A | G | -0.3016 | 0.0682 | 9.70E-06 | IFNy_Cryptococcus_PBMC_7days | 0.294104324 | 0.022040411 |
| rs34754340 | 3 | 46000345 | chr3:46000345:C:T | T | C | 0.3673 | 0.0772 | 1.94E-06 | IL6_LPS100ng_PBMC_24h | -0.235205446 | 0.022082841 |
| rs10506436 | 12 | 62028786 | chr12:62028786:T:A | A | T | -0.2811 | 0.06 | 2.84E-06 | IL22_C.albicansconidia_PBMC_7days | -0.318813328 | 0.022163963 |
| rs807599 | 2 | 15652990 | chr2:15652990:T:C | T | C | 0.3084 | 0.0674 | 4.79E-06 | IFNy_Cryptococcus_PBMC_7days | -0.293457013 | 0.022169454 |
| rs807615 | 2 | 15648378 | chr2:15648378:A:G | A | G | 0.309 | 0.068 | 5.51E-06 | IFNy_C.albicanshyphae_PBMC_7days | 0.375967771 | 0.022444199 |
| rs807597 | 2 | 15653780 | chr2:15653780:T:C | T | C | 0.3087 | 0.0675 | 4.80E-06 | IFNy_Cryptococcus_PBMC_7days | -0.292633263 | 0.022507684 |
| rs807598 | 2 | 15653525 | chr2:15653525:G:A | A | G | -0.3069 | 0.0682 | 6.74E-06 | IFNy_Cryptococcus_PBMC_7days | 0.292613375 | 0.022542267 |
| rs9738190 | 12 | 62078790 | chr12:62078790:G:A | A | G | -0.2898 | 0.0602 | 1.51E-06 | IFNy_C.albicanshyphae_PBMC_7days | -0.350275517 | 0.022567642 |
| rs807613 | 2 | 15649443 | chr2:15649443:C:T | T | C | -0.3086 | 0.0679 | 5.60E-06 | IFNy_C.albicanshyphae_PBMC_7days | -0.374911588 | 0.022857509 |
| rs807611 | 2 | 15649857 | chr2:15649857:G:T | T | G | -0.3084 | 0.0672 | 4.51E-06 | IFNy_C.albicanshyphae_PBMC_7days | -0.374370976 | 0.023067963 |
| rs12185539 | 2 | 15650052 | chr2:15650052:A:G | A | G | 0.3079 | 0.067 | 4.31E-06 | IFNy_C.albicanshyphae_PBMC_7days | 0.373565458 | 0.023386494 |
| rs12185540 | 2 | 15650056 | chr2:15650056:A:G | A | G | 0.3058 | 0.0677 | 6.19E-06 | IFNy_C.albicanshyphae_PBMC_7days | 0.373565458 | 0.023386494 |
| rs11174296 | 12 | 62077677 | chr12:62077677:C:G | C | G | 0.288 | 0.0602 | 1.75E-06 | IFNy_C.albicanshyphae_PBMC_7days | 0.347948073 | 0.023419578 |
| rs34836513 | 3 | 46004967 | chr3:46004967:G:A | A | G | 0.3674 | 0.0772 | 1.96E-06 | IL6_LPS100ng_PBMC_24h | -0.232409611 | 0.023484604 |
| rs10506432 | 12 | 62077116 | chr12:62077116:A:C | A | C | 0.29 | 0.0604 | 1.58E-06 | IFNy_C.albicanshyphae_PBMC_7days | 0.347620147 | 0.023541418 |
| rs41264195 | 2 | 15606096 | chr2:15606096:C:T | T | C | -0.3359 | 0.0754 | 8.49E-06 | IL17_C.albicanshyphae_PBMC_7days | -0.348584218 | 0.023773422 |
| rs35669129 | 3 | 46013758 | chr3:46013758:G:A | A | G | 0.3746 | 0.0768 | 1.08E-06 | IL6_LPS100ng_PBMC_24h | -0.231716379 | 0.023809114 |
| rs13081213 | 3 | 46014417 | chr3:46014417:C:T | T | C | 0.3746 | 0.0768 | 1.08E-06 | IL6_LPS100ng_PBMC_24h | -0.231663216 | 0.023813451 |
| rs13081151 | 3 | 46014224 | chr3:46014224:G:A | A | G | 0.3751 | 0.0777 | 1.38E-06 | IL6_LPS100ng_PBMC_24h | -0.231648963 | 0.023825405 |
| rs34168660 | 3 | 46014670 | chr3:46014670:G:A | A | G | 0.375 | 0.0768 | 1.06E-06 | IL6_LPS100ng_PBMC_24h | -0.231544198 | 0.02386828 |
| rs3770462 | 2 | 15609200 | chr2:15609200:T:G | T | G | 0.3409 | 0.0757 | 6.74E-06 | IL17_C.albicanshyphae_PBMC_7days | 0.34830602 | 0.023873854 |
| rs4668941 | 2 | 15610007 | chr2:15610007:T:G | T | G | 0.3359 | 0.0754 | 8.51E-06 | IL17_C.albicanshyphae_PBMC_7days | 0.34830602 | 0.023873854 |
| rs79808866 | 2 | 15604860 | chr2:15604860:T:G | T | G | 0.3357 | 0.0754 | 8.59E-06 | IL17_C.albicanshyphae_PBMC_7days | 0.348266507 | 0.023900774 |
| rs4362758 | 3 | 46015419 | chr3:46015419:T:A | A | T | 0.3681 | 0.0768 | 1.67E-06 | IL6_LPS100ng_PBMC_24h | -0.231438833 | 0.023902514 |
| rs10747951 | 12 | 62075677 | chr12:62075677:C:T | T | C | -0.29 | 0.0604 | 1.58E-06 | IFNy_C.albicanshyphae_PBMC_7days | -0.346628004 | 0.02390767 |
| rs10877786 | 12 | 62070198 | chr12:62070198:T:G | T | G | 0.2964 | 0.0597 | 6.89E-07 | IFNy_C.albicanshyphae_PBMC_7days | 0.345111273 | 0.023969569 |
| rs35751180 | 3 | 46014681 | chr3:46014681:C:T | T | C | 0.3731 | 0.0774 | 1.42E-06 | IL6_LPS100ng_PBMC_24h | -0.231082275 | 0.024084677 |
| rs807610 | 2 | 15650440 | chr2:15650440:C:G | C | G | 0.3082 | 0.0673 | 4.64E-06 | IFNy_C.albicanshyphae_PBMC_7days | 0.371579086 | 0.024165368 |
| rs9706516 | 12 | 62074403 | chr12:62074403:C:A | A | C | -0.2923 | 0.0604 | 1.30E-06 | IFNy_C.albicanshyphae_PBMC_7days | -0.345660986 | 0.024266993 |
| rs62122279 | 2 | 15621842 | chr2:15621842:T:C | T | C | 0.3047 | 0.0678 | 6.91E-06 | IFNy_C.albicanshyphae_PBMC_7days | 0.368857676 | 0.024415712 |
| rs4668946 | 2 | 15624494 | chr2:15624494:A:G | A | G | 0.3109 | 0.0679 | 4.72E-06 | IFNy_C.albicanshyphae_PBMC_7days | 0.368651836 | 0.024477419 |
| rs12614308 | 2 | 15624914 | chr2:15624914:G:A | A | G | -0.3109 | 0.0679 | 4.72E-06 | IFNy_C.albicanshyphae_PBMC_7days | -0.368651836 | 0.024477419 |
| rs10877787 | 12 | 62072981 | chr12:62072981:A:G | A | G | 0.29 | 0.0604 | 1.58E-06 | IFNy_C.albicanshyphae_PBMC_7days | 0.344863846 | 0.024562136 |
| rs807594 | 2 | 15654931 | chr2:15654931:C:T | T | C | -0.302 | 0.0682 | 9.44E-06 | IFNy_C.albicanshyphae_PBMC_7days | -0.371625707 | 0.024575247 |
| rs9739956 | 12 | 62065731 | chr12:62065731:T:C | T | C | 0.2934 | 0.0598 | 9.45E-07 | IFNy_C.albicanshyphae_PBMC_7days | 0.343617544 | 0.024584637 |
| rs12610495 | 19 | 4717660 | chr19:4717660:A:G | A | G | -0.2534 | 0.0556 | 5.20E-06 | IL1b_Cryptococcus_PBMC_24h | 0.384151651 | 0.025222458 |
| rs11174294 | 12 | 62072129 | chr12:62072129:C:T | T | C | -0.29 | 0.0604 | 1.59E-06 | IFNy_C.albicanshyphae_PBMC_7days | -0.343240833 | 0.025240247 |
| rs34836513 | 3 | 46004967 | chr3:46004967:G:A | A | G | 0.3674 | 0.0772 | 1.96E-06 | IL1b_MSUC16_PBMC_24h | -0.364013419 | 0.025583958 |
| rs13263570 | 8 | 121845942 | chr8:121845942:G:A | A | G | -0.2298 | 0.0516 | 8.50E-06 | IL1b_Cryptococcus_PBMC_24h | 0.390417408 | 0.0256092 |
| rs11627388 | 14 | 31953140 | chr14:31953140:A:C | A | C | -0.2482 | 0.0533 | 3.19E-06 | IL6_B.burgdorferi_PBMC_24h | -0.22584737 | 0.025699809 |
| rs807627 | 2 | 15639565 | chr2:15639565:T:C | T | C | 0.3105 | 0.0679 | 4.79E-06 | IFNy_C.albicanshyphae_PBMC_7days | 0.363107973 | 0.026357215 |
| rs807628 | 2 | 15638176 | chr2:15638176:A:G | A | G | 0.3095 | 0.0678 | 5.04E-06 | IFNy_C.albicanshyphae_PBMC_7days | 0.363147078 | 0.026358477 |
| rs807607 | 2 | 15651223 | chr2:15651223:G:C | C | G | -0.3087 | 0.0679 | 5.52E-06 | IFNy_C.albicanshyphae_PBMC_7days | -0.3660826 | 0.026376583 |
| rs2059266 | 16 | 74902133 | chr16:74902133:A:G | A | G | -0.2457 | 0.0533 | 4.01E-06 | IL22_MTB_PBMC_7days | -0.227339809 | 0.026453247 |
| rs13063033 | 3 | 46040989 | chr3:46040989:G:A | A | G | 0.3555 | 0.0772 | 4.12E-06 | IFNy_C.albicansconidia_PBMC_7days | -0.332180822 | 0.026815875 |
| rs807602 | 2 | 15652485 | chr2:15652485:C:A | A | C | -0.3021 | 0.0679 | 8.60E-06 | IFNy_C.albicanshyphae_PBMC_7days | -0.363333607 | 0.027446957 |
| rs807606 | 2 | 15651602 | chr2:15651602:C:A | A | C | -0.3042 | 0.0679 | 7.55E-06 | IFNy_C.albicanshyphae_PBMC_7days | -0.36323067 | 0.027528422 |
| rs13263570 | 8 | 121845942 | chr8:121845942:G:A | A | G | -0.2298 | 0.0516 | 8.50E-06 | IL1b_E.Coli_PBMC_24h | 0.200629057 | 0.027796815 |
| rs807594 | 2 | 15654931 | chr2:15654931:C:T | T | C | -0.302 | 0.0682 | 9.44E-06 | IFNy_Cryptococcus_PBMC_7days | 0.282603386 | 0.028088102 |
| rs3965316 | 7 | 158231123 | chr7:158231123:C:T | T | C | 0.2305 | 0.0509 | 6.03E-06 | TNFA_A.fumigatusconidiaSerum_PBMC_24h | -0.118104142 | 0.028123992 |
| rs13063033 | 3 | 46040989 | chr3:46040989:G:A | A | G | 0.3555 | 0.0772 | 4.12E-06 | IL6_LPS100ng_PBMC_24h | -0.209198867 | 0.028138756 |
| rs6963124 | 7 | 158230897 | chr7:158230897:T:A | A | T | 0.2323 | 0.0509 | 5.05E-06 | TNFA_A.fumigatusconidiaSerum_PBMC_24h | -0.118050037 | 0.028192764 |
| rs2335848 | 7 | 158229411 | chr7:158229411:A:G | A | G | -0.2321 | 0.051 | 5.36E-06 | TNFA_A.fumigatusconidiaSerum_PBMC_24h | 0.118023898 | 0.028197873 |
| rs10273930 | 7 | 158232429 | chr7:158232429:A:G | A | G | -0.2288 | 0.0509 | 7.00E-06 | TNFA_A.fumigatusconidiaSerum_PBMC_24h | 0.118043575 | 0.028199238 |
| rs807605 | 2 | 15652128 | chr2:15652128:G:A | A | G | -0.3007 | 0.0679 | 9.38E-06 | IFNy_C.albicanshyphae_PBMC_7days | -0.361538482 | 0.02822652 |
| rs807604 | 2 | 15652384 | chr2:15652384:T:A | A | T | -0.3037 | 0.0679 | 7.64E-06 | IFNy_C.albicanshyphae_PBMC_7days | -0.361516922 | 0.028231669 |
| rs10276960 | 7 | 158232334 | chr7:158232334:T:C | T | C | -0.2305 | 0.0509 | 5.93E-06 | TNFA_A.fumigatusconidiaSerum_PBMC_24h | 0.118018433 | 0.028233777 |
| rs6974918 | 7 | 158227592 | chr7:158227592:T:G | T | G | -0.2313 | 0.0509 | 5.55E-06 | TNFA_A.fumigatusconidiaSerum_PBMC_24h | 0.117923464 | 0.028301855 |
| rs6951312 | 7 | 158227595 | chr7:158227595:C:A | A | C | 0.2315 | 0.0509 | 5.48E-06 | TNFA_A.fumigatusconidiaSerum_PBMC_24h | -0.117923464 | 0.028301855 |
| rs6974924 | 7 | 158227601 | chr7:158227601:T:C | T | C | -0.2315 | 0.0509 | 5.48E-06 | TNFA_A.fumigatusconidiaSerum_PBMC_24h | 0.117923464 | 0.028301855 |
| rs2302929 | 2 | 15595524 | chr2:15595524:T:C | T | C | 0.2976 | 0.0672 | 9.54E-06 | IL17_C.albicanshyphae_PBMC_7days | 0.325416361 | 0.028402278 |
| rs12706520 | 7 | 123366673 | chr7:123366673:G:A | A | G | -0.2835 | 0.0641 | 9.63E-06 | IL1b_MSUC16_PBMC_24h | -0.265699801 | 0.028411319 |
| rs807592 | 2 | 15657078 | chr2:15657078:T:C | T | C | 0.3573 | 0.0788 | 5.82E-06 | IL1b_MTB_PBMC_24h | -0.294327974 | 0.028510767 |
| rs807627 | 2 | 15639565 | chr2:15639565:T:C | T | C | 0.3105 | 0.0679 | 4.79E-06 | IFNy_Cryptococcus_PBMC_7days | -0.278632728 | 0.028568338 |
| rs807628 | 2 | 15638176 | chr2:15638176:A:G | A | G | 0.3095 | 0.0678 | 5.04E-06 | IFNy_Cryptococcus_PBMC_7days | -0.278541271 | 0.028639461 |
| rs6949642 | 7 | 158232636 | chr7:158232636:C:A | A | C | 0.2273 | 0.0509 | 8.02E-06 | TNFA_A.fumigatusconidiaSerum_PBMC_24h | -0.119409549 | 0.029179325 |
| rs35669129 | 3 | 46013758 | chr3:46013758:G:A | A | G | 0.3746 | 0.0768 | 1.08E-06 | IL1b_MSUC16_PBMC_24h | -0.355231004 | 0.029274541 |
| rs36122610 | 3 | 45981341 | chr3:45981341:G:A | A | G | 0.3673 | 0.0775 | 2.18E-06 | IL22_A.fumigatusconidia_PBMC_7days | 0.439264687 | 0.029324556 |
| rs13081151 | 3 | 46014224 | chr3:46014224:G:A | A | G | 0.3751 | 0.0777 | 1.38E-06 | IL1b_MSUC16_PBMC_24h | -0.354636453 | 0.02951907 |
| rs6970487 | 7 | 158227455 | chr7:158227455:A:G | A | G | -0.2327 | 0.0509 | 4.89E-06 | TNFA_A.fumigatusconidiaSerum_PBMC_24h | 0.117110796 | 0.029553992 |
| rs13081213 | 3 | 46014417 | chr3:46014417:C:T | T | C | 0.3746 | 0.0768 | 1.08E-06 | IL1b_MSUC16_PBMC_24h | -0.354361958 | 0.029642076 |
| rs807623 | 2 | 15642467 | chr2:15642467:C:T | T | C | -0.312 | 0.0679 | 4.30E-06 | IFNy_C.albicanshyphae_PBMC_7days | -0.353365053 | 0.029704141 |
| rs807622 | 2 | 15642631 | chr2:15642631:A:G | A | G | 0.3099 | 0.0675 | 4.43E-06 | IFNy_C.albicanshyphae_PBMC_7days | 0.353365053 | 0.029704141 |
| rs807620 | 2 | 15643012 | chr2:15643012:C:T | T | C | -0.312 | 0.0679 | 4.30E-06 | IFNy_C.albicanshyphae_PBMC_7days | -0.353365053 | 0.029704141 |
| rs34168660 | 3 | 46014670 | chr3:46014670:G:A | A | G | 0.375 | 0.0768 | 1.06E-06 | IL1b_MSUC16_PBMC_24h | -0.354004579 | 0.029787175 |
| rs35751180 | 3 | 46014681 | chr3:46014681:C:T | T | C | 0.3731 | 0.0774 | 1.42E-06 | IL1b_MSUC16_PBMC_24h | -0.353818025 | 0.029796447 |
| rs36122610 | 3 | 45981341 | chr3:45981341:G:A | A | G | 0.3673 | 0.0775 | 2.18E-06 | IFNy_A.fumigatusconidia_PBMC_7days | 0.390998369 | 0.029802731 |
| rs2302929 | 2 | 15595524 | chr2:15595524:T:C | T | C | 0.2976 | 0.0672 | 9.54E-06 | TNFA_C.albicansconidia_PBMC_24h | 0.176564951 | 0.029908784 |
| rs10156036 | 7 | 158224085 | chr7:158224085:A:C | A | C | -0.2322 | 0.0509 | 4.96E-06 | TNFA_A.fumigatusconidiaSerum_PBMC_24h | 0.116806822 | 0.02995493 |
| rs807618 | 2 | 15646936 | chr2:15646936:G:C | C | G | -0.3046 | 0.0673 | 6.02E-06 | IFNy_C.albicanshyphae_PBMC_7days | -0.352738521 | 0.029981835 |
| rs10784279 | 12 | 62044016 | chr12:62044016:C:T | T | C | -0.2778 | 0.0599 | 3.50E-06 | IFNy_C.albicanshyphae_PBMC_7days | -0.333411069 | 0.030105227 |
| rs4362758 | 3 | 46015419 | chr3:46015419:T:A | A | T | 0.3681 | 0.0768 | 1.67E-06 | IL1b_MSUC16_PBMC_24h | -0.353029273 | 0.030208025 |
| rs34442130 | 3 | 45983037 | chr3:45983037:T:A | A | T | 0.3674 | 0.0775 | 2.16E-06 | IL22_A.fumigatusconidia_PBMC_7days | 0.436830925 | 0.030238686 |
| rs13075758 | 3 | 45983556 | chr3:45983556:G:A | A | G | 0.3672 | 0.0775 | 2.19E-06 | IL22_A.fumigatusconidia_PBMC_7days | 0.436543352 | 0.030338485 |
| rs13075758 | 3 | 45983556 | chr3:45983556:G:A | A | G | 0.3672 | 0.0775 | 2.19E-06 | IFNy_A.fumigatusconidia_PBMC_7days | 0.38959212 | 0.030390113 |
| rs34442130 | 3 | 45983037 | chr3:45983037:T:A | A | T | 0.3674 | 0.0775 | 2.16E-06 | IFNy_A.fumigatusconidia_PBMC_7days | 0.389610777 | 0.030391925 |
| rs2080811 | 2 | 15584186 | chr2:15584186:G:A | A | G | -0.329 | 0.0676 | 1.12E-06 | IFNy_C.albicanshyphae_PBMC_7days | -0.346908494 | 0.030568944 |
| rs10784278 | 12 | 62042165 | chr12:62042165:T:A | A | T | -0.2836 | 0.06 | 2.32E-06 | IFNy_C.albicanshyphae_PBMC_7days | -0.332119797 | 0.030766006 |
| rs6948273 | 7 | 158222765 | chr7:158222765:T:C | T | C | -0.2286 | 0.0509 | 6.99E-06 | TNFA_A.fumigatusconidiaSerum_PBMC_24h | 0.116046495 | 0.030968978 |
| rs4506155 | 7 | 158211289 | chr7:158211289:G:A | A | G | 0.2258 | 0.0508 | 8.62E-06 | TNFA_A.fumigatusconidiaSerum_PBMC_24h | -0.116094507 | 0.031009703 |
| rs2302929 | 2 | 15595524 | chr2:15595524:T:C | T | C | 0.2976 | 0.0672 | 9.54E-06 | TNFA_S.aureus_PBMC_24h | 0.284447289 | 0.031119016 |
| rs12619108 | 2 | 15586369 | chr2:15586369:G:A | A | G | -0.315 | 0.0658 | 1.68E-06 | IFNy_C.albicanshyphae_PBMC_7days | -0.345289563 | 0.031123081 |
| rs12610495 | 19 | 4717660 | chr19:4717660:A:G | A | G | -0.2534 | 0.0556 | 5.20E-06 | IL6_A.fumigatusconidiaSerum_PBMC_24h | 0.234889741 | 0.031217237 |
| rs4595081 | 7 | 158215231 | chr7:158215231:G:T | T | G | 0.2285 | 0.0507 | 6.62E-06 | TNFA_A.fumigatusconidiaSerum_PBMC_24h | -0.115909054 | 0.031229106 |
| rs2191031 | 3 | 45869378 | chr3:45869378:G:A | A | G | 0.2758 | 0.0613 | 6.80E-06 | IFNy_A.fumigatusconidia_PBMC_7days | 0.297689315 | 0.03129479 |
| rs34836513 | 3 | 46004967 | chr3:46004967:G:A | A | G | 0.3674 | 0.0772 | 1.96E-06 | IL6_A.fumigatusconidiaSerum_PBMC_24h | -0.366044886 | 0.031311893 |
| rs807577 | 2 | 15664270 | chr2:15664270:C:G | C | G | 0.3181 | 0.0704 | 6.28E-06 | IFNy_C.albicanshyphae_PBMC_7days | 0.356214725 | 0.031514973 |
| rs9739464 | 12 | 62037992 | chr12:62037992:A:T | A | T | 0.2846 | 0.06 | 2.10E-06 | IFNy_C.albicanshyphae_PBMC_7days | 0.330964427 | 0.031564145 |
| rs807587 | 2 | 15659856 | chr2:15659856:C:G | C | G | 0.3246 | 0.0702 | 3.76E-06 | IFNy_C.albicanshyphae_PBMC_7days | 0.356003235 | 0.031733199 |
| rs807586 | 2 | 15660004 | chr2:15660004:T:C | T | C | 0.3125 | 0.0703 | 8.76E-06 | IFNy_C.albicanshyphae_PBMC_7days | 0.356003235 | 0.031733199 |
| rs807583 | 2 | 15660597 | chr2:15660597:G:A | A | G | -0.3121 | 0.0703 | 9.01E-06 | IFNy_C.albicanshyphae_PBMC_7days | -0.356003235 | 0.031733199 |
| rs807579 | 2 | 15663460 | chr2:15663460:T:C | T | C | 0.3124 | 0.0703 | 8.88E-06 | IFNy_C.albicanshyphae_PBMC_7days | 0.356003235 | 0.031733199 |
| rs807578 | 2 | 15663516 | chr2:15663516:G:C | C | G | -0.3124 | 0.0703 | 8.81E-06 | IFNy_C.albicanshyphae_PBMC_7days | -0.356003235 | 0.031733199 |
| rs807589 | 2 | 15658661 | chr2:15658661:C:A | A | C | -0.326 | 0.0702 | 3.48E-06 | IFNy_C.albicanshyphae_PBMC_7days | -0.356054354 | 0.031780296 |
| rs34754340 | 3 | 46000345 | chr3:46000345:C:T | T | C | 0.3673 | 0.0772 | 1.94E-06 | IL6_A.fumigatusconidiaSerum_PBMC_24h | -0.365661987 | 0.031783607 |
| rs2191031 | 3 | 45869378 | chr3:45869378:G:A | A | G | 0.2758 | 0.0613 | 6.80E-06 | IL22_C.albicansconidia_PBMC_7days | -0.306430354 | 0.031789233 |
| rs34754340 | 3 | 46000345 | chr3:46000345:C:T | T | C | 0.3673 | 0.0772 | 1.94E-06 | IL1b_MSUC16_PBMC_24h | -0.35074504 | 0.031799565 |
| rs7152677 | 14 | 31953540 | chr14:31953540:G:C | C | G | 0.2637 | 0.0548 | 1.52E-06 | IL6_B.burgdorferi_PBMC_24h | 0.22846225 | 0.031999139 |
| rs71615438 | 3 | 45998376 | chr3:45998376:A:G | A | G | -0.3666 | 0.0772 | 2.04E-06 | IL6_A.fumigatusconidiaSerum_PBMC_24h | 0.365225646 | 0.032087674 |
| rs6949642 | 7 | 158232636 | chr7:158232636:C:A | A | C | 0.2273 | 0.0509 | 8.02E-06 | IFNy_C.albicanshyphae_PBMC_7days | 0.270015054 | 0.032136512 |
| rs36023124 | 3 | 45997568 | chr3:45997568:G:C | C | G | 0.3665 | 0.0771 | 2.01E-06 | IL6_A.fumigatusconidiaSerum_PBMC_24h | -0.365080383 | 0.03219389 |
| rs17330872 | 3 | 45993605 | chr3:45993605:A:G | A | G | -0.3702 | 0.0774 | 1.74E-06 | IL6_A.fumigatusconidiaSerum_PBMC_24h | 0.364100466 | 0.032769909 |
| rs633862 | 9 | 133279871 | chr9:133279871:T:C | T | C | 0.2534 | 0.0515 | 8.79E-07 | IL6_Influenza_PBMC_24h | 0.389243738 | 0.033626907 |
| rs1491950 | 3 | 46096615 | chr3:46096615:G:A | A | G | 0.3544 | 0.078 | 5.55E-06 | IL6_A.fumigatusconidiaSerum_PBMC_24h | -0.350732108 | 0.033677535 |
| rs2080811 | 2 | 15584186 | chr2:15584186:G:A | A | G | -0.329 | 0.0676 | 1.12E-06 | TNFA_S.aureus_PBMC_24h | -0.278824971 | 0.033745954 |
| rs6949642 | 7 | 158232636 | chr7:158232636:C:A | A | C | 0.2273 | 0.0509 | 8.02E-06 | IL22_C.albicanshyphae_PBMC_7days | 0.252986566 | 0.033769578 |
| rs12619108 | 2 | 15586369 | chr2:15586369:G:A | A | G | -0.315 | 0.0658 | 1.68E-06 | TNFA_S.aureus_PBMC_24h | -0.278209121 | 0.033893413 |
| rs12619874 | 2 | 15586997 | chr2:15586997:G:C | C | G | -0.3204 | 0.0665 | 1.46E-06 | IL1b_MTB_PBMC_24h | 0.259577179 | 0.033964947 |
| rs13263570 | 8 | 121845942 | chr8:121845942:G:A | A | G | -0.2298 | 0.0516 | 8.50E-06 | IL1b_MSUC16_PBMC_24h | 0.224984493 | 0.034797776 |
| rs4668946 | 2 | 15624494 | chr2:15624494:A:G | A | G | 0.3109 | 0.0679 | 4.72E-06 | IFNy_Cryptococcus_PBMC_7days | -0.269308496 | 0.034808535 |
| rs12614308 | 2 | 15624914 | chr2:15624914:G:A | A | G | -0.3109 | 0.0679 | 4.72E-06 | IFNy_Cryptococcus_PBMC_7days | 0.269308496 | 0.034808535 |
| rs71615438 | 3 | 45998376 | chr3:45998376:A:G | A | G | -0.3666 | 0.0772 | 2.04E-06 | IL1b_MSUC16_PBMC_24h | 0.344679638 | 0.034998099 |
| rs62122279 | 2 | 15621842 | chr2:15621842:T:C | T | C | 0.3047 | 0.0678 | 6.91E-06 | IFNy_Cryptococcus_PBMC_7days | -0.269047112 | 0.035009727 |
| rs807576 | 2 | 15664332 | chr2:15664332:T:C | T | C | 0.3185 | 0.0704 | 6.15E-06 | IFNy_C.albicanshyphae_PBMC_7days | 0.347053817 | 0.035009877 |
| rs807590 | 2 | 15657801 | chr2:15657801:G:T | T | G | -0.3146 | 0.0696 | 6.11E-06 | IFNy_C.albicanshyphae_PBMC_7days | -0.349595134 | 0.035298012 |
| rs17330872 | 3 | 45993605 | chr3:45993605:A:G | A | G | -0.3702 | 0.0774 | 1.74E-06 | IFNy_A.fumigatusconidia_PBMC_7days | -0.378470354 | 0.035863852 |
| rs10506436 | 12 | 62028786 | chr12:62028786:T:A | A | T | -0.2811 | 0.06 | 2.84E-06 | IFNy_C.albicanshyphae_PBMC_7days | -0.322808715 | 0.035889074 |
| rs7152677 | 14 | 31953540 | chr14:31953540:G:C | C | G | 0.2637 | 0.0548 | 1.52E-06 | IL1b_B.fragilis_PBMC_24h | -0.23324202 | 0.036111224 |
| rs17330872 | 3 | 45993605 | chr3:45993605:A:G | A | G | -0.3702 | 0.0774 | 1.74E-06 | IL22_A.fumigatusconidia_PBMC_7days | -0.423353027 | 0.036126825 |
| rs36023124 | 3 | 45997568 | chr3:45997568:G:C | C | G | 0.3665 | 0.0771 | 2.01E-06 | IL22_A.fumigatusconidia_PBMC_7days | 0.422522193 | 0.036379774 |
| rs12619874 | 2 | 15586997 | chr2:15586997:G:C | C | G | -0.3204 | 0.0665 | 1.46E-06 | TNFA_C.albicansconidia_PBMC_24h | -0.166557496 | 0.036426701 |
| rs71615438 | 3 | 45998376 | chr3:45998376:A:G | A | G | -0.3666 | 0.0772 | 2.04E-06 | IL22_A.fumigatusconidia_PBMC_7days | -0.422300755 | 0.036437812 |
| rs10877786 | 12 | 62070198 | chr12:62070198:T:G | T | G | 0.2964 | 0.0597 | 6.89E-07 | IL22_C.albicansconidia_PBMC_7days | 0.289959918 | 0.036443572 |
| rs4797120 | 18 | 3725189 | chr18:3725189:G:C | C | G | -0.2956 | 0.0621 | 1.91E-06 | TNFA_Pam3Cys_PBMC_24h | -0.221026368 | 0.036486102 |
| rs36122610 | 3 | 45981341 | chr3:45981341:G:A | A | G | 0.3673 | 0.0775 | 2.18E-06 | IL6_A.fumigatusconidiaSerum_PBMC_24h | -0.356013009 | 0.036490392 |
| rs34442130 | 3 | 45983037 | chr3:45983037:T:A | A | T | 0.3674 | 0.0775 | 2.16E-06 | IL6_A.fumigatusconidiaSerum_PBMC_24h | -0.356011396 | 0.03649144 |
| rs13075758 | 3 | 45983556 | chr3:45983556:G:A | A | G | 0.3672 | 0.0775 | 2.19E-06 | IL6_A.fumigatusconidiaSerum_PBMC_24h | -0.355935626 | 0.036520469 |
| rs36023124 | 3 | 45997568 | chr3:45997568:G:C | C | G | 0.3665 | 0.0771 | 2.01E-06 | IL1b_MSUC16_PBMC_24h | -0.341899266 | 0.036543543 |
| rs807636 | 2 | 15627828 | chr2:15627828:C:T | T | C | -0.365 | 0.0772 | 2.25E-06 | IL17_C.albicanshyphae_PBMC_7days | -0.327077453 | 0.036612279 |
| rs36023124 | 3 | 45997568 | chr3:45997568:G:C | C | G | 0.3665 | 0.0771 | 2.01E-06 | IFNy_A.fumigatusconidia_PBMC_7days | 0.376710962 | 0.036622041 |
| rs73833520 | 3 | 46016375 | chr3:46016375:A:G | A | G | -0.3679 | 0.0768 | 1.69E-06 | IL22_A.fumigatusconidia_PBMC_7days | -0.414639837 | 0.036685905 |
| rs35669129 | 3 | 46013758 | chr3:46013758:G:A | A | G | 0.3746 | 0.0768 | 1.08E-06 | IL6_A.fumigatusconidiaSerum_PBMC_24h | -0.355002925 | 0.036689575 |
| rs34754340 | 3 | 46000345 | chr3:46000345:C:T | T | C | 0.3673 | 0.0772 | 1.94E-06 | IL22_A.fumigatusconidia_PBMC_7days | 0.421338601 | 0.036760744 |
| rs13063033 | 3 | 46040989 | chr3:46040989:G:A | A | G | 0.3555 | 0.0772 | 4.12E-06 | IL6_A.fumigatusconidiaSerum_PBMC_24h | -0.329783606 | 0.03676413 |
| rs10087789 | 8 | 121836324 | chr8:121836324:C:G | C | G | 0.2327 | 0.0518 | 7.00E-06 | IL1b_MSUC16_PBMC_24h | -0.224414153 | 0.036771598 |
| rs71615438 | 3 | 45998376 | chr3:45998376:A:G | A | G | -0.3666 | 0.0772 | 2.04E-06 | IFNy_A.fumigatusconidia_PBMC_7days | -0.376289402 | 0.036792968 |
| rs9739956 | 12 | 62065731 | chr12:62065731:T:C | T | C | 0.2934 | 0.0598 | 9.45E-07 | IL22_C.albicansconidia_PBMC_7days | 0.289344129 | 0.036838041 |
| rs10091098 | 8 | 121836590 | chr8:121836590:G:A | A | G | -0.233 | 0.0508 | 4.55E-06 | IL1b_MSUC16_PBMC_24h | 0.224307264 | 0.036860043 |
| rs13081151 | 3 | 46014224 | chr3:46014224:G:A | A | G | 0.3751 | 0.0777 | 1.38E-06 | IL6_A.fumigatusconidiaSerum_PBMC_24h | -0.354498178 | 0.036925805 |
| rs807635 | 2 | 15629483 | chr2:15629483:C:T | T | C | -0.3579 | 0.0765 | 2.85E-06 | IL17_C.albicanshyphae_PBMC_7days | -0.326092314 | 0.036948372 |
| rs13081213 | 3 | 46014417 | chr3:46014417:C:T | T | C | 0.3746 | 0.0768 | 1.08E-06 | IL6_A.fumigatusconidiaSerum_PBMC_24h | -0.35418827 | 0.037087634 |
| rs34836513 | 3 | 46004967 | chr3:46004967:G:A | A | G | 0.3674 | 0.0772 | 1.96E-06 | IL22_A.fumigatusconidia_PBMC_7days | 0.41937273 | 0.037324622 |
| rs35751180 | 3 | 46014681 | chr3:46014681:C:T | T | C | 0.3731 | 0.0774 | 1.42E-06 | IL6_A.fumigatusconidiaSerum_PBMC_24h | -0.35351761 | 0.037335399 |
| rs34168660 | 3 | 46014670 | chr3:46014670:G:A | A | G | 0.375 | 0.0768 | 1.06E-06 | IL6_A.fumigatusconidiaSerum_PBMC_24h | -0.353605825 | 0.037377714 |
| rs34754340 | 3 | 46000345 | chr3:46000345:C:T | T | C | 0.3673 | 0.0772 | 1.94E-06 | IFNy_A.fumigatusconidia_PBMC_7days | 0.374868258 | 0.037404892 |
| rs4389947 | 8 | 121816361 | chr8:121816361:G:A | A | G | -0.2364 | 0.0517 | 4.91E-06 | IL1b_MSUC16_PBMC_24h | 0.223356136 | 0.037721181 |
| rs4362758 | 3 | 46015419 | chr3:46015419:T:A | A | T | 0.3681 | 0.0768 | 1.67E-06 | IL6_A.fumigatusconidiaSerum_PBMC_24h | -0.352644029 | 0.037860033 |
| rs7152677 | 14 | 31953540 | chr14:31953540:G:C | C | G | 0.2637 | 0.0548 | 1.52E-06 | IL1b_E.Coli_PBMC_24h | -0.209361304 | 0.038283582 |
| rs34836513 | 3 | 46004967 | chr3:46004967:G:A | A | G | 0.3674 | 0.0772 | 1.96E-06 | IFNy_A.fumigatusconidia_PBMC_7days | 0.37196481 | 0.038577233 |
| rs10088118 | 8 | 121806381 | chr8:121806381:G:C | C | G | -0.2329 | 0.0517 | 6.79E-06 | IL1b_MSUC16_PBMC_24h | 0.222025401 | 0.038955343 |
| rs807581 | 2 | 15662227 | chr2:15662227:T:G | T | G | 0.3698 | 0.0806 | 4.51E-06 | IL1b_MTB_PBMC_24h | -0.274824591 | 0.039288161 |
| rs71327014 | 3 | 46059974 | chr3:46059974:C:T | T | C | 0.3556 | 0.0773 | 4.25E-06 | IL22_A.fumigatusconidia_PBMC_7days | 0.40607718 | 0.039349956 |
| rs71327015 | 3 | 46060681 | chr3:46060681:G:C | C | G | 0.3585 | 0.0773 | 3.57E-06 | IL22_A.fumigatusconidia_PBMC_7days | 0.40607718 | 0.039349956 |
| rs34492478 | 3 | 46097350 | chr3:46097350:A:T | A | T | -0.3562 | 0.078 | 4.95E-06 | IL6_A.fumigatusconidiaSerum_PBMC_24h | 0.341687958 | 0.039418835 |
| rs34047915 | 3 | 46058999 | chr3:46058999:C:T | T | C | 0.3555 | 0.0773 | 4.29E-06 | IL22_A.fumigatusconidia_PBMC_7days | 0.405865717 | 0.039456551 |
| rs34127208 | 3 | 46062188 | chr3:46062188:G:T | T | G | 0.3554 | 0.0773 | 4.31E-06 | IL22_A.fumigatusconidia_PBMC_7days | 0.405838068 | 0.039465661 |
| rs35516580 | 3 | 46066006 | chr3:46066006:A:G | A | G | -0.3562 | 0.0773 | 4.13E-06 | IL22_A.fumigatusconidia_PBMC_7days | -0.405750975 | 0.039503801 |
| rs34766614 | 3 | 46066109 | chr3:46066109:A:G | A | G | -0.3484 | 0.0775 | 6.99E-06 | IL22_A.fumigatusconidia_PBMC_7days | -0.405750975 | 0.039503801 |
| rs35420565 | 3 | 46058646 | chr3:46058646:C:G | C | G | -0.3556 | 0.0773 | 4.26E-06 | IL22_A.fumigatusconidia_PBMC_7days | -0.405653183 | 0.039563653 |
| rs2191031 | 3 | 45869378 | chr3:45869378:G:A | A | G | 0.2758 | 0.0613 | 6.80E-06 | IL22_A.fumigatusconidia_PBMC_7days | 0.318673009 | 0.039657828 |
| rs2302934 | 2 | 15613155 | chr2:15613155:C:G | C | G | 0.3393 | 0.076 | 8.11E-06 | IL17_C.albicanshyphae_PBMC_7days | 0.32218619 | 0.039964791 |
| rs12616567 | 2 | 15615498 | chr2:15615498:A:C | A | C | 0.3427 | 0.0762 | 6.85E-06 | IL17_C.albicanshyphae_PBMC_7days | 0.32218619 | 0.039964791 |
| rs633862 | 9 | 133279871 | chr9:133279871:T:C | T | C | 0.2534 | 0.0515 | 8.79E-07 | IL1b_C.albicanshyphae_PBMC_24h | 0.157685277 | 0.039972279 |
| rs13096741 | 3 | 46057662 | chr3:46057662:C:T | T | C | 0.3556 | 0.0773 | 4.27E-06 | IL22_A.fumigatusconidia_PBMC_7days | 0.404792371 | 0.039997157 |
| rs34718164 | 3 | 46057034 | chr3:46057034:C:G | C | G | -0.3556 | 0.0773 | 4.25E-06 | IL22_A.fumigatusconidia_PBMC_7days | -0.404355582 | 0.040216995 |
| rs71327006 | 3 | 46017416 | chr3:46017416:G:A | A | G | 0.3628 | 0.0774 | 2.77E-06 | IL22_A.fumigatusconidia_PBMC_7days | 0.402794549 | 0.04083317 |
| rs77399277 | 3 | 46055306 | chr3:46055306:T:C | T | C | -0.3556 | 0.0773 | 4.25E-06 | IL22_A.fumigatusconidia_PBMC_7days | -0.402793532 | 0.041002875 |
| rs34340587 | 3 | 46054551 | chr3:46054551:G:A | A | G | 0.3542 | 0.0773 | 4.63E-06 | IL22_A.fumigatusconidia_PBMC_7days | 0.402662937 | 0.041072924 |
| rs12610495 | 19 | 4717660 | chr19:4717660:A:G | A | G | -0.2534 | 0.0556 | 5.20E-06 | TNFA_MSUC16_PBMC_24h | 0.149402284 | 0.041095819 |
| rs71327007 | 3 | 46017757 | chr3:46017757:C:T | T | C | 0.3643 | 0.0774 | 2.51E-06 | IL22_A.fumigatusconidia_PBMC_7days | 0.401965603 | 0.041226197 |
| rs613534 | 9 | 133267707 | chr9:133267707:G:A | A | G | -0.2801 | 0.0517 | 6.16E-08 | IL6_MSUC16_PBMC_24h | -0.202206274 | 0.041243421 |
| rs35772789 | 3 | 46053612 | chr3:46053612:A:G | A | G | -0.3544 | 0.0773 | 4.52E-06 | IL22_A.fumigatusconidia_PBMC_7days | -0.401883351 | 0.041459757 |
| rs35454877 | 3 | 46017992 | chr3:46017992:T:C | T | C | -0.3679 | 0.0768 | 1.69E-06 | IL22_A.fumigatusconidia_PBMC_7days | -0.40140222 | 0.041500556 |
| rs34452002 | 3 | 46101695 | chr3:46101695:C:T | T | C | 0.3556 | 0.0779 | 4.95E-06 | IL22_A.fumigatusconidia_PBMC_7days | 0.400026003 | 0.041527018 |
| rs34093271 | 3 | 46103489 | chr3:46103489:G:T | T | G | 0.3558 | 0.0779 | 4.89E-06 | IL22_A.fumigatusconidia_PBMC_7days | 0.400162886 | 0.041547642 |
| rs74586549 | 3 | 45884551 | chr3:45884551:C:T | T | C | 0.3443 | 0.0768 | 7.34E-06 | TNFA_S.aureus_PBMC_24h | 0.319181099 | 0.041589605 |
| rs73833520 | 3 | 46016375 | chr3:46016375:A:G | A | G | -0.3679 | 0.0768 | 1.69E-06 | IL6_A.fumigatusconidiaSerum_PBMC_24h | 0.341038333 | 0.04184029 |
| rs34863575 | 3 | 46052571 | chr3:46052571:A:G | A | G | -0.3543 | 0.0773 | 4.53E-06 | IL22_A.fumigatusconidia_PBMC_7days | -0.400946855 | 0.041930171 |
| rs12610495 | 19 | 4717660 | chr19:4717660:A:G | A | G | -0.2534 | 0.0556 | 5.20E-06 | IL6_Influenza_PBMC_24h | 0.40993399 | 0.04220645 |
| rs13091868 | 3 | 46081551 | chr3:46081551:A:G | A | G | -0.3538 | 0.0773 | 4.72E-06 | IL6_A.fumigatusconidiaSerum_PBMC_24h | 0.336020745 | 0.04233629 |

| | | | | | | | | | | | |
|---|---|---|---|---|---|---|---|---|---|---|---|
| rs13092030 | 3 | 46081563 | chr3:46081563:C:T | T | C | 0.354 | 0.0773 | 4.66E-06 | IL6_A.fumigatusconidiaSerum_PBMC_24h | -0.336020745 | 0.04233629 |
| rs17215981 | 3 | 46082926 | chr3:46082926:A:G | A | G | -0.3547 | 0.0773 | 4.47E-06 | IL6_A.fumigatusconidiaSerum_PBMC_24h | 0.336020745 | 0.04233629 |
| rs13089554 | 3 | 46086065 | chr3:46086065:G:A | A | G | 0.3547 | 0.0774 | 4.61E-06 | IL6_A.fumigatusconidiaSerum_PBMC_24h | -0.336020745 | 0.04233629 |
| rs13095717 | 3 | 46087050 | chr3:46087050:G:A | A | G | 0.3548 | 0.0774 | 4.58E-06 | IL6_A.fumigatusconidiaSerum_PBMC_24h | -0.336020745 | 0.04233629 |
| rs13075528 | 3 | 46087217 | chr3:46087217:T:C | T | C | -0.3549 | 0.0775 | 4.63E-06 | IL6_A.fumigatusconidiaSerum_PBMC_24h | 0.336020745 | 0.04233629 |
| rs13095602 | 3 | 46087221 | chr3:46087221:A:G | A | G | -0.3547 | 0.0774 | 4.60E-06 | IL6_A.fumigatusconidiaSerum_PBMC_24h | 0.336020745 | 0.04233629 |
| rs71327023 | 3 | 46089733 | chr3:46089733:C:G | C | G | -0.3539 | 0.0777 | 5.19E-06 | IL6_A.fumigatusconidiaSerum_PBMC_24h | 0.336020745 | 0.04233629 |
| rs13063527 | 3 | 46094198 | chr3:46094198:G:A | A | G | 0.3561 | 0.0777 | 4.51E-06 | IL6_A.fumigatusconidiaSerum_PBMC_24h | -0.336020745 | 0.04233629 |
| rs13068570 | 3 | 46094946 | chr3:46094946:A:G | A | G | -0.3561 | 0.0777 | 4.61E-06 | IL6_A.fumigatusconidiaSerum_PBMC_24h | 0.336020745 | 0.04233629 |
| rs4373103 | 3 | 46096963 | chr3:46096963:G:A | A | G | 0.3547 | 0.078 | 5.42E-06 | IL6_A.fumigatusconidiaSerum_PBMC_24h | -0.336020745 | 0.04233629 |
| rs71327024 | 3 | 46098581 | chr3:46098581:G:T | T | G | 0.3547 | 0.078 | 5.37E-06 | IL6_A.fumigatusconidiaSerum_PBMC_24h | -0.336020745 | 0.04233629 |
| rs71327017 | 3 | 46069721 | chr3:46069721:T:C | T | C | -0.3576 | 0.0774 | 3.80E-06 | IL6_A.fumigatusconidiaSerum_PBMC_24h | 0.335921054 | 0.042403215 |
| rs13072267 | 3 | 46074518 | chr3:46074518:T:A | A | T | 0.3543 | 0.0773 | 4.57E-06 | IL6_A.fumigatusconidiaSerum_PBMC_24h | -0.335921054 | 0.042403215 |
| rs36057789 | 3 | 46075604 | chr3:46075604:A:G | A | G | -0.3551 | 0.0773 | 4.36E-06 | IL6_A.fumigatusconidiaSerum_PBMC_24h | 0.335921054 | 0.042403215 |
| rs76647202 | 3 | 46076468 | chr3:46076468:A:G | A | G | -0.3542 | 0.0773 | 4.60E-06 | IL6_A.fumigatusconidiaSerum_PBMC_24h | 0.335921054 | 0.042403215 |
| rs71327021 | 3 | 46079025 | chr3:46079025:A:G | A | G | -0.3542 | 0.0773 | 4.61E-06 | IL6_A.fumigatusconidiaSerum_PBMC_24h | 0.335921054 | 0.042403215 |
| rs34460587 | 3 | 46100972 | chr3:46100972:C:T | T | C | 0.3557 | 0.0779 | 4.93E-06 | IL6_A.fumigatusconidiaSerum_PBMC_24h | -0.335925944 | 0.042533821 |
| rs807594 | 2 | 15654931 | chr2:15654931:C:T | T | C | -0.302 | 0.0682 | 9.44E-06 | IL17_C.albicanshyphae_PBMC_7days | -0.308920861 | 0.042619711 |
| rs34452002 | 3 | 46101695 | chr3:46101695:C:T | T | C | 0.3556 | 0.0779 | 4.95E-06 | IL6_A.fumigatusconidiaSerum_PBMC_24h | -0.335690766 | 0.042775835 |
| rs6970487 | 7 | 158227455 | chr7:158227455:A:G | A | G | -0.2327 | 0.0509 | 4.89E-06 | IFNy_C.albicanshyphae_PBMC_7days | -0.250983403 | 0.042815635 |
| rs62122279 | 2 | 15621842 | chr2:15621842:T:C | T | C | 0.3047 | 0.0678 | 6.91E-06 | IL17_C.albicanshyphae_PBMC_7days | 0.305877948 | 0.042893892 |
| rs4797120 | 18 | 3725189 | chr18:3725189:G:C | C | G | -0.2956 | 0.0621 | 1.91E-06 | IL6_MSUC16_PBMC_24h | -0.223300036 | 0.04295502 |
| rs34093271 | 3 | 46103489 | chr3:46103489:G:T | T | G | 0.3558 | 0.0779 | 4.89E-06 | IL6_A.fumigatusconidiaSerum_PBMC_24h | -0.335505901 | 0.042983541 |
| rs79808866 | 2 | 15604860 | chr2:15604860:T:G | T | G | 0.3357 | 0.0754 | 8.59E-06 | IL1b_Borreliamix_PBMC_24h | -0.219991416 | 0.042989856 |
| rs3770462 | 2 | 15609200 | chr2:15609200:T:G | T | G | 0.3409 | 0.0757 | 6.74E-06 | IL1b_Borreliamix_PBMC_24h | -0.219924142 | 0.043037808 |
| rs4668941 | 2 | 15610007 | chr2:15610007:T:G | T | G | 0.3359 | 0.0754 | 8.51E-06 | IL1b_Borreliamix_PBMC_24h | -0.219924142 | 0.043037808 |
| rs4668946 | 2 | 15624494 | chr2:15624494:A:G | A | G | 0.3109 | 0.0679 | 4.72E-06 | IL17_C.albicanshyphae_PBMC_7days | 0.305538424 | 0.043099467 |
| rs12614308 | 2 | 15624914 | chr2:15624914:G:A | A | G | -0.3109 | 0.0679 | 4.72E-06 | IL17_C.albicanshyphae_PBMC_7days | -0.305538424 | 0.043099467 |
| rs41264195 | 2 | 15606096 | chr2:15606096:C:T | T | C | -0.3359 | 0.0754 | 8.49E-06 | IL1b_Borreliamix_PBMC_24h | 0.219870672 | 0.043107002 |
| rs74586549 | 3 | 45884551 | chr3:45884551:C:T | T | C | 0.3443 | 0.0768 | 7.34E-06 | IL22_A.fumigatusconidia_PBMC_7days | 0.38061677 | 0.043121375 |
| rs34127208 | 3 | 46062188 | chr3:46062188:G:T | T | G | 0.3554 | 0.0773 | 4.31E-06 | IL1b_MSUC16_PBMC_24h | -0.322841177 | 0.043124608 |
| rs71327014 | 3 | 46059974 | chr3:46059974:C:T | T | C | 0.3556 | 0.0773 | 4.25E-06 | IL1b_MSUC16_PBMC_24h | -0.322764359 | 0.043175523 |
| rs71327015 | 3 | 46060681 | chr3:46060681:G:C | C | G | 0.3585 | 0.0773 | 3.57E-06 | IL1b_MSUC16_PBMC_24h | -0.322764359 | 0.043175523 |
| rs35516580 | 3 | 46066006 | chr3:46066006:A:G | A | G | -0.3562 | 0.0773 | 4.13E-06 | IL1b_MSUC16_PBMC_24h | 0.322678507 | 0.043225751 |
| rs34766614 | 3 | 46066109 | chr3:46066109:A:G | A | G | -0.3484 | 0.0775 | 6.99E-06 | IL1b_MSUC16_PBMC_24h | 0.322678507 | 0.043225751 |
| rs34047915 | 3 | 46058999 | chr3:46058999:C:T | T | C | 0.3555 | 0.0773 | 4.29E-06 | IL1b_MSUC16_PBMC_24h | -0.322653694 | 0.043251239 |
| rs35420565 | 3 | 46058646 | chr3:46058646:C:G | C | G | -0.3556 | 0.0773 | 4.26E-06 | IL1b_MSUC16_PBMC_24h | 0.322542182 | 0.043327344 |
| rs3965316 | 7 | 158231123 | chr7:158231123:C:T | T | C | 0.2305 | 0.0509 | 6.03E-06 | IFNy_C.albicanshyphae_PBMC_7days | 0.250207635 | 0.043372788 |
| rs6930922 | 6 | 20314659 | chr6:20314659:C:T | T | C | 0.265 | 0.058 | 4.86E-06 | IL17_MTB_PBMC_7days | 0.213327097 | 0.043388001 |
| rs6974918 | 7 | 158227592 | chr7:158227592:T:G | T | G | -0.2313 | 0.0509 | 5.55E-06 | IFNy_C.albicanshyphae_PBMC_7days | -0.250065851 | 0.04340594 |
| rs6951312 | 7 | 158227595 | chr7:158227595:C:A | A | C | 0.2315 | 0.0509 | 5.48E-06 | IFNy_C.albicanshyphae_PBMC_7days | 0.250065851 | 0.04340594 |
| rs6974924 | 7 | 158227601 | chr7:158227601:T:C | T | C | -0.2315 | 0.0509 | 5.48E-06 | IFNy_C.albicanshyphae_PBMC_7days | -0.250065851 | 0.04340594 |
| rs6963124 | 7 | 158230897 | chr7:158230897:T:A | A | T | 0.2323 | 0.0509 | 5.05E-06 | IFNy_C.albicanshyphae_PBMC_7days | 0.250048502 | 0.043501816 |
| rs633862 | 9 | 133279871 | chr9:133279871:T:C | T | C | 0.2534 | 0.0515 | 8.79E-07 | TNFA_Influenza_PBMC_24h | 0.2606462 | 0.043507152 |
| rs10273930 | 7 | 158232429 | chr7:158232429:A:G | A | G | -0.2288 | 0.0509 | 7.00E-06 | IFNy_C.albicanshyphae_PBMC_7days | -0.250024065 | 0.043519504 |
| rs10276960 | 7 | 158232334 | chr7:158232334:T:C | T | C | -0.2305 | 0.0509 | 5.93E-06 | IFNy_C.albicanshyphae_PBMC_7days | -0.249983507 | 0.04355475 |
| rs2335848 | 7 | 158229411 | chr7:158229411:A:G | A | G | -0.2321 | 0.051 | 5.36E-06 | IFNy_C.albicanshyphae_PBMC_7days | -0.249880386 | 0.043603854 |
| rs13096741 | 3 | 46057662 | chr3:46057662:C:T | T | C | 0.3556 | 0.0773 | 4.27E-06 | IL1b_MSUC16_PBMC_24h | -0.322087676 | 0.043635665 |
| rs911360 | 6 | 20314488 | chr6:20314488:C:T | T | C | 0.2673 | 0.058 | 4.09E-06 | IL17_MTB_PBMC_7days | 0.213219166 | 0.043759461 |
| rs34718164 | 3 | 46057034 | chr3:46057034:C:G | C | G | -0.3556 | 0.0773 | 4.25E-06 | IL1b_MSUC16_PBMC_24h | 0.32185536 | 0.043792183 |
| rs550057 | 9 | 133271182 | chr9:133271182:T:C | T | C | 0.2681 | 0.0562 | 1.86E-06 | IL1b_LPS1ng_PBMC_24h | 0.265999284 | 0.043906316 |
| rs17330872 | 3 | 45993605 | chr3:45993605:A:G | A | G | -0.3702 | 0.0774 | 1.74E-06 | IL1b_MSUC16_PBMC_24h | 0.329715651 | 0.043936927 |
| rs13063033 | 3 | 46040989 | chr3:46040989:G:A | A | G | 0.3555 | 0.0772 | 4.12E-06 | IL22_B.burgdorferi_PBMC_7days | -0.306326433 | 0.044037466 |
| rs807581 | 2 | 15662227 | chr2:15662227:T:G | T | G | 0.3698 | 0.0806 | 4.51E-06 | IL17_C.albicanshyphae_PBMC_7days | 0.318790515 | 0.044088958 |
| rs13093179 | 3 | 46104822 | chr3:46104822:G:T | T | G | 0.3557 | 0.0779 | 4.91E-06 | IL22_A.fumigatusconidia_PBMC_7days | 0.400344914 | 0.044091489 |
| rs34679077 | 3 | 46046500 | chr3:46046500:G:A | A | G | 0.3609 | 0.0774 | 3.13E-06 | IL22_A.fumigatusconidia_PBMC_7days | 0.396530246 | 0.044157411 |
| rs807592 | 2 | 15657078 | chr2:15657078:T:C | T | C | 0.3573 | 0.0788 | 5.82E-06 | IL17_C.albicanshyphae_PBMC_7days | 0.32124205 | 0.044199511 |
| rs73833520 | 3 | 46016375 | chr3:46016375:A:G | A | G | -0.3679 | 0.0768 | 1.69E-06 | IL1b_MSUC16_PBMC_24h | 0.323429415 | 0.044240474 |
| rs71327025 | 3 | 46106012 | chr3:46106012:A:G | A | G | -0.3556 | 0.0779 | 4.96E-06 | IL6_A.fumigatusconidiaSerum_PBMC_24h | 0.334239812 | 0.044314017 |
| rs73833520 | 3 | 46016375 | chr3:46016375:A:G | A | G | -0.3679 | 0.0768 | 1.69E-06 | IL6_LPS100ng_PBMC_24h | 0.203437811 | 0.044315986 |
| rs77399277 | 3 | 46055306 | chr3:46055306:T:C | T | C | -0.3556 | 0.0773 | 4.25E-06 | IL1b_MSUC16_PBMC_24h | 0.32101579 | 0.044352523 |
| rs3774641 | 3 | 45896341 | chr3:45896341:G:T | T | G | 0.2741 | 0.0605 | 5.95E-06 | IL6_Influenza_PBMC_24h | -0.477004432 | 0.044388521 |
| rs41264195 | 2 | 15606096 | chr2:15606096:C:T | T | C | -0.3359 | 0.0754 | 8.49E-06 | IL1b_B.burgdorferi_PBMC_24h | 0.224397577 | 0.0444111 |
| rs79808866 | 2 | 15604860 | chr2:15604860:T:G | T | G | 0.3357 | 0.0754 | 8.59E-06 | IL1b_B.burgdorferi_PBMC_24h | -0.224341564 | 0.044462335 |
| rs34340587 | 3 | 46054551 | chr3:46054551:G:A | A | G | 0.3542 | 0.0773 | 4.63E-06 | IL1b_MSUC16_PBMC_24h | -0.320853393 | 0.044465031 |
| rs3770462 | 2 | 15609200 | chr2:15609200:T:G | T | G | 0.3409 | 0.0757 | 6.74E-06 | IL1b_B.burgdorferi_PBMC_24h | -0.224266965 | 0.044517044 |
| rs4668941 | 2 | 15610007 | chr2:15610007:T:G | T | G | 0.3359 | 0.0754 | 8.51E-06 | IL1b_B.burgdorferi_PBMC_24h | -0.224266965 | 0.044517044 |
| rs13093179 | 3 | 46104822 | chr3:46104822:G:T | T | G | 0.3557 | 0.0779 | 4.91E-06 | IL6_A.fumigatusconidiaSerum_PBMC_24h | -0.337089313 | 0.044663971 |
| rs71327025 | 3 | 46106012 | chr3:46106012:A:G | A | G | -0.3556 | 0.0779 | 4.96E-06 | IL22_A.fumigatusconidia_PBMC_7days | -0.395227335 | 0.04467704 |
| rs35772789 | 3 | 46053612 | chr3:46053612:A:G | A | G | -0.3544 | 0.0773 | 4.52E-06 | IL1b_MSUC16_PBMC_24h | 0.320312496 | 0.044817964 |
| rs35539222 | 3 | 46112780 | chr3:46112780:T:C | T | C | -0.3556 | 0.0779 | 4.96E-06 | IL6_A.fumigatusconidiaSerum_PBMC_24h | 0.333507638 | 0.044877306 |
| rs35110864 | 3 | 46112965 | chr3:46112965:G:A | A | G | 0.3555 | 0.0779 | 4.99E-06 | IL6_A.fumigatusconidiaSerum_PBMC_24h | -0.333507638 | 0.044877306 |
| rs807595 | 2 | 15654550 | chr2:15654550:C:T | T | C | -0.3019 | 0.0682 | 9.52E-06 | IFNy_C.albicanshyphae_PBMC_7days | -0.331239691 | 0.044948406 |
| rs77399277 | 3 | 46055306 | chr3:46055306:T:C | T | C | -0.3556 | 0.0773 | 4.25E-06 | IL6_LPS100ng_PBMC_24h | 0.201339048 | 0.045046459 |
| rs34340587 | 3 | 46054551 | chr3:46054551:G:A | A | G | 0.3542 | 0.0773 | 4.63E-06 | IL6_LPS100ng_PBMC_24h | -0.20132653 | 0.045064268 |
| rs34127208 | 3 | 46062188 | chr3:46062188:G:T | T | G | 0.3554 | 0.0773 | 4.31E-06 | IL6_LPS100ng_PBMC_24h | -0.20129305 | 0.045067522 |
| rs34718164 | 3 | 46057034 | chr3:46057034:C:G | C | G | -0.3556 | 0.0773 | 4.25E-06 | IL6_LPS100ng_PBMC_24h | 0.201307246 | 0.045075691 |
| rs807599 | 2 | 15652990 | chr2:15652990:T:C | T | C | 0.3084 | 0.0674 | 4.79E-06 | IFNy_C.albicanshyphae_PBMC_7days | 0.330535103 | 0.045085794 |
| rs13096741 | 3 | 46057662 | chr3:46057662:C:T | T | C | 0.3556 | 0.0773 | 4.27E-06 | IL6_LPS100ng_PBMC_24h | -0.201293444 | 0.045086574 |
| rs34863575 | 3 | 46052571 | chr3:46052571:A:G | A | G | -0.3543 | 0.0773 | 4.53E-06 | IL6_LPS100ng_PBMC_24h | 0.201287557 | 0.045088082 |
| rs35772789 | 3 | 46053612 | chr3:46053612:A:G | A | G | -0.3544 | 0.0773 | 4.52E-06 | IL6_LPS100ng_PBMC_24h | 0.201291398 | 0.045092995 |
| rs35516580 | 3 | 46066006 | chr3:46066006:A:G | A | G | -0.3562 | 0.0773 | 4.13E-06 | IL6_LPS100ng_PBMC_24h | 0.201255877 | 0.045102575 |
| rs34766614 | 3 | 46066109 | chr3:46066109:A:G | A | G | -0.3484 | 0.0775 | 6.99E-06 | IL6_LPS100ng_PBMC_24h | 0.201255877 | 0.045102575 |
| rs35420565 | 3 | 46058646 | chr3:46058646:C:G | C | G | -0.3556 | 0.0773 | 4.26E-06 | IL6_LPS100ng_PBMC_24h | 0.201259555 | 0.045111715 |
| rs34047915 | 3 | 46058999 | chr3:46058999:C:T | T | C | 0.3555 | 0.0773 | 4.29E-06 | IL6_LPS100ng_PBMC_24h | -0.201249775 | 0.045118705 |
| rs34679077 | 3 | 46046500 | chr3:46046500:G:A | A | G | 0.3609 | 0.0774 | 3.13E-06 | IL6_LPS100ng_PBMC_24h | -0.201166629 | 0.045121564 |
| rs71327014 | 3 | 46059974 | chr3:46059974:C:T | T | C | 0.3556 | 0.0773 | 4.25E-06 | IL6_LPS100ng_PBMC_24h | -0.201239471 | 0.045125975 |
| rs71327015 | 3 | 46060681 | chr3:46060681:G:C | C | G | 0.3585 | 0.0773 | 3.57E-06 | IL6_LPS100ng_PBMC_24h | -0.201239471 | 0.045125975 |
| rs34863575 | 3 | 46052571 | chr3:46052571:A:G | A | G | -0.3543 | 0.0773 | 4.53E-06 | IL1b_MSUC16_PBMC_24h | 0.319797738 | 0.045155519 |
| rs34438204 | 3 | 46039814 | chr3:46039814:T:C | T | C | -0.3609 | 0.0774 | 3.13E-06 | IL6_LPS100ng_PBMC_24h | 0.200992173 | 0.045210696 |
| rs807596 | 2 | 15654457 | chr2:15654457:G:A | A | G | -0.3016 | 0.0682 | 9.70E-06 | IFNy_C.albicanshyphae_PBMC_7days | -0.33072743 | 0.045228017 |
| rs35539222 | 3 | 46112780 | chr3:46112780:T:C | T | C | -0.3556 | 0.0779 | 4.96E-06 | IL6_LPS100ng_PBMC_24h | 0.200967423 | 0.045243882 |
| rs35110864 | 3 | 46112965 | chr3:46112965:G:A | A | G | 0.3555 | 0.0779 | 4.99E-06 | IL6_LPS100ng_PBMC_24h | -0.200967423 | 0.045243882 |
| rs9739464 | 12 | 62037992 | chr12:62037992:A:T | A | T | 0.2846 | 0.06 | 2.10E-06 | IL6_MTB_PBMC_24h | 0.225777343 | 0.045251416 |
| rs2088692 | 3 | 46035840 | chr3:46035840:A:G | A | G | -0.3654 | 0.0768 | 1.96E-06 | IL6_LPS100ng_PBMC_24h | 0.200859488 | 0.045269877 |
| rs4389947 | 8 | 121816361 | chr8:121816361:G:A | A | G | -0.2364 | 0.0517 | 4.91E-06 | IFNy_Bacteroides_PBMC_7days | 0.140306239 | 0.045298337 |
| rs35334665 | 3 | 46019726 | chr3:46019726:T:C | T | C | -0.3632 | 0.0774 | 2.70E-06 | IL22_A.fumigatusconidia_PBMC_7days | -0.393898749 | 0.04529905 |
| rs6791016 | 3 | 46033366 | chr3:46033366:G:A | A | G | 0.367 | 0.0769 | 1.80E-06 | IL6_LPS100ng_PBMC_24h | -0.200756624 | 0.045335239 |
| rs6780028 | 3 | 46033598 | chr3:46033598:A:C | A | C | -0.367 | 0.0769 | 1.80E-06 | IL6_LPS100ng_PBMC_24h | 0.200756624 | 0.045335239 |
| rs6790866 | 3 | 46033219 | chr3:46033219:G:A | A | G | 0.367 | 0.0769 | 1.80E-06 | IL6_LPS100ng_PBMC_24h | -0.200734056 | 0.045345663 |
| rs807598 | 2 | 15653525 | chr2:15653525:G:A | A | G | -0.3069 | 0.0682 | 6.74E-06 | IFNy_C.albicanshyphae_PBMC_7days | -0.330093002 | 0.045349627 |
| rs10506436 | 12 | 62028786 | chr12:62028786:T:A | A | T | -0.2811 | 0.06 | 2.84E-06 | IL6_MTB_PBMC_24h | -0.225503185 | 0.045353783 |
| rs35814488 | 3 | 46032431 | chr3:46032431:A:G | A | G | -0.3672 | 0.0769 | 1.77E-06 | IL6_LPS100ng_PBMC_24h | 0.200710973 | 0.045356368 |
| rs34774687 | 3 | 46031822 | chr3:46031822:T:C | T | C | -0.367 | 0.0769 | 1.80E-06 | IL6_LPS100ng_PBMC_24h | 0.200687376 | 0.045367354 |
| rs34619093 | 3 | 46031027 | chr3:46031027:G:A | A | G | 0.3625 | 0.0775 | 2.89E-06 | IL6_LPS100ng_PBMC_24h | -0.200663264 | 0.04537862 |
| rs68087193 | 3 | 46031138 | chr3:46031138:C:T | T | C | 0.3673 | 0.0769 | 1.76E-06 | IL6_LPS100ng_PBMC_24h | -0.200663264 | 0.04537862 |
| rs2102055 | 3 | 46031212 | chr3:46031212:G:A | A | G | 0.3591 | 0.0767 | 2.83E-06 | IL6_LPS100ng_PBMC_24h | -0.200663264 | 0.04537862 |
| rs2102056 | 3 | 46031276 | chr3:46031276:G:A | A | G | 0.3671 | 0.0769 | 1.78E-06 | IL6_LPS100ng_PBMC_24h | -0.200663264 | 0.04537862 |
| rs2088690 | 3 | 46031347 | chr3:46031347:C:T | T | C | 0.3671 | 0.0769 | 1.78E-06 | IL6_LPS100ng_PBMC_24h | -0.200663264 | 0.04537862 |
| rs13060287 | 3 | 46030376 | chr3:46030376:A:G | A | G | -0.3671 | 0.0769 | 1.79E-06 | IL6_LPS100ng_PBMC_24h | 0.200620851 | 0.045414428 |
| rs550057 | 9 | 133271182 | chr9:133271182:T:C | T | C | 0.2681 | 0.0562 | 1.86E-06 | IL1b_S.aureus_PBMC_24h | 0.111119621 | 0.045419262 |
| rs13082697 | 3 | 46030271 | chr3:46030271:T:C | T | C | -0.3671 | 0.0769 | 1.78E-06 | IL6_LPS100ng_PBMC_24h | 0.200595715 | 0.04542626 |
| rs10088118 | 8 | 121806381 | chr8:121806381:G:C | C | G | -0.2329 | 0.0517 | 6.79E-06 | IFNy_Bacteroides_PBMC_7days | 0.140274555 | 0.045427402 |
| rs36040135 | 3 | 46019812 | chr3:46019812:A:G | A | G | -0.3679 | 0.0768 | 1.69E-06 | IL22_A.fumigatusconidia_PBMC_7days | -0.393619808 | 0.045433926 |
| rs34438204 | 3 | 46039814 | chr3:46039814:T:C | T | C | -0.3609 | 0.0774 | 3.13E-06 | IL22_A.fumigatusconidia_PBMC_7days | -0.393995054 | 0.045438016 |
| rs34562820 | 3 | 46029589 | chr3:46029589:G:A | A | G | 0.3625 | 0.0775 | 2.87E-06 | IL6_LPS100ng_PBMC_24h | -0.200570066 | 0.045438372 |
| rs59166269 | 3 | 46028845 | chr3:46028845:C:G | C | G | -0.3672 | 0.0769 | 1.77E-06 | IL6_LPS100ng_PBMC_24h | 0.200543905 | 0.045450766 |
| rs57437758 | 3 | 46028861 | chr3:46028861:A:C | A | C | -0.3672 | 0.0769 | 1.77E-06 | IL6_LPS100ng_PBMC_24h | 0.200543905 | 0.045450766 |
| rs10091098 | 8 | 121836590 | chr8:121836590:G:A | A | G | -0.233 | 0.0508 | 4.55E-06 | IFNy_Bacteroides_PBMC_7days | 0.140170409 | 0.045452084 |
| rs10087789 | 8 | 121836324 | chr8:121836324:C:G | C | G | 0.2327 | 0.0518 | 7.00E-06 | IFNy_Bacteroides_PBMC_7days | -0.140167507 | 0.045458636 |
| rs13090194 | 3 | 46028081 | chr3:46028081:A:C | A | C | -0.3672 | 0.0769 | 1.77E-06 | IL6_LPS100ng_PBMC_24h | 0.200517231 | 0.04546344 |
| rs71327010 | 3 | 46027343 | chr3:46027343:G:T | T | G | 0.3625 | 0.0774 | 2.83E-06 | IL6_LPS100ng_PBMC_24h | -0.20046235 | 0.045489633 |
| rs35159820 | 3 | 46026981 | chr3:46026981:A:G | A | G | -0.3673 | 0.0769 | 1.76E-06 | IL6_LPS100ng_PBMC_24h | 0.200434142 | 0.04550315 |
| rs35161099 | 3 | 46026015 | chr3:46026015:T:G | T | G | -0.3625 | 0.0774 | 2.83E-06 | IL6_LPS100ng_PBMC_24h | 0.200405424 | 0.045516949 |
| rs12610495 | 19 | 4717660 | chr19:4717660:A:G | A | G | -0.2534 | 0.0556 | 5.20E-06 | IL6_Cryptococcus_PBMC_24h | 0.397549471 | 0.045538991 |
| rs35539222 | 3 | 46112780 | chr3:46112780:T:C | T | C | -0.3556 | 0.0779 | 4.96E-06 | IL22_A.fumigatusconidia_PBMC_7days | -0.393805611 | 0.04554897 |
| rs35110864 | 3 | 46112965 | chr3:46112965:G:A | A | G | 0.3555 | 0.0779 | 4.99E-06 | IL22_A.fumigatusconidia_PBMC_7days | 0.393805611 | 0.04554897 |
| rs35930050 | 3 | 46024271 | chr3:46024271:G:A | A | G | 0.3681 | 0.0768 | 1.67E-06 | IL6_LPS100ng_PBMC_24h | -0.200285459 | 0.045574956 |
| rs34584867 | 3 | 46024471 | chr3:46024471:G:A | A | G | 0.3681 | 0.0768 | 1.67E-06 | IL6_LPS100ng_PBMC_24h | -0.200285459 | 0.045574956 |
| rs35334665 | 3 | 46019726 | chr3:46019726:T:C | T | C | -0.3632 | 0.0774 | 2.70E-06 | IL6_LPS100ng_PBMC_24h | 0.200436677 | 0.045618593 |
| rs13097556 | 3 | 46020505 | chr3:46020505:T:C | T | C | -0.3679 | 0.0768 | 1.69E-06 | IL6_LPS100ng_PBMC_24h | 0.200330227 | 0.045622742 |
| rs13074382 | 3 | 46020348 | chr3:46020348:A:G | A | G | -0.3679 | 0.0768 | 1.69E-06 | IL6_LPS100ng_PBMC_24h | 0.200347045 | 0.045626563 |
| rs876668 | 3 | 46022261 | chr3:46022261:T:C | T | C | -0.368 | 0.0768 | 1.68E-06 | IL6_LPS100ng_PBMC_24h | 0.200157367 | 0.045637464 |
| rs36040135 | 3 | 46019812 | chr3:46019812:A:G | A | G | -0.3679 | 0.0768 | 1.69E-06 | IL6_LPS100ng_PBMC_24h | 0.200399038 | 0.045640771 |
| rs35454877 | 3 | 46017992 | chr3:46017992:T:C | T | C | -0.3679 | 0.0768 | 1.69E-06 | IL6_LPS100ng_PBMC_24h | 0.200541863 | 0.045685019 |
| rs71327006 | 3 | 46017416 | chr3:46017416:G:A | A | G | 0.3628 | 0.0774 | 2.77E-06 | IL6_LPS100ng_PBMC_24h | -0.200571663 | 0.045693047 |
| rs71327007 | 3 | 46017757 | chr3:46017757:C:T | T | C | 0.3643 | 0.0774 | 2.51E-06 | IL6_LPS100ng_PBMC_24h | -0.200541782 | 0.045698042 |
| rs2230322 | 3 | 46021837 | chr3:46021837:T:C | T | C | -0.368 | 0.0768 | 1.68E-06 | IL6_LPS100ng_PBMC_24h | 0.198686686 | 0.045764183 |
| rs7152677 | 14 | 31953540 | chr14:31953540:G:C | C | G | 0.2637 | 0.0548 | 1.52E-06 | TNFA_MSUC16_PBMC_24h | 0.164936297 | 0.045782522 |
| rs807597 | 2 | 15653780 | chr2:15653780:T:C | T | C | 0.3087 | 0.0675 | 4.80E-06 | IFNy_C.albicanshyphae_PBMC_7days | 0.329120208 | 0.045950871 |
| rs2302929 | 2 | 15595524 | chr2:15595524:T:C | T | C | 0.2976 | 0.0672 | 9.54E-06 | IL22_C.albicanshyphae_PBMC_7days | 0.304290928 | 0.046006789 |
| rs10156036 | 7 | 158224085 | chr7:158224085:A:C | A | C | -0.2322 | 0.0509 | 4.96E-06 | IFNy_C.albicanshyphae_PBMC_7days | -0.247182302 | 0.046043182 |
| rs71327006 | 3 | 46017416 | chr3:46017416:G:A | A | G | 0.3628 | 0.0774 | 2.77E-06 | IL1b_MSUC16_PBMC_24h | -0.318142137 | 0.046116423 |
| rs71327025 | 3 | 46106012 | chr3:46106012:A:G | A | G | -0.3556 | 0.0779 | 4.96E-06 | IL6_LPS100ng_PBMC_24h | 0.199930578 | 0.04627564 |
| rs12619108 | 2 | 15586369 | chr2:15586369:G:A | A | G | -0.315 | 0.0658 | 1.68E-06 | IFNy_Cryptococcus_PBMC_7days | 0.248554644 | 0.046295829 |
| rs12619108 | 2 | 15586369 | chr2:15586369:G:A | A | G | -0.315 | 0.0658 | 1.68E-06 | IL1b_MTB_PBMC_24h | 0.247726091 | 0.046304565 |
| rs2088692 | 3 | 46035840 | chr3:46035840:A:G | A | G | -0.3654 | 0.0768 | 1.96E-06 | IL22_A.fumigatusconidia_PBMC_7days | -0.392114514 | 0.046397954 |
| rs2080811 | 2 | 15584186 | chr2:15584186:G:A | A | G | -0.329 | 0.0676 | 1.12E-06 | IL1b_MTB_PBMC_24h | 0.247950026 | 0.046410938 |
| rs71327006 | 3 | 46017416 | chr3:46017416:G:A | A | G | 0.3628 | 0.0774 | 2.77E-06 | IL6_A.fumigatusconidiaSerum_PBMC_24h | -0.331174826 | 0.046425427 |
| rs71327007 | 3 | 46017757 | chr3:46017757:C:T | T | C | 0.3643 | 0.0774 | 2.51E-06 | IL1b_MSUC16_PBMC_24h | -0.317636212 | 0.046437885 |
| rs34460587 | 3 | 46100972 | chr3:46100972:C:T | T | C | 0.3557 | 0.0779 | 4.93E-06 | IL22_A.fumigatusconidia_PBMC_7days | 0.390666189 | 0.046456149 |
| rs2080811 | 2 | 15584186 | chr2:15584186:G:A | A | G | -0.329 | 0.0676 | 1.12E-06 | IFNy_Cryptococcus_PBMC_7days | 0.248688435 | 0.046482528 |
| rs35454877 | 3 | 46017992 | chr3:46017992:T:C | T | C | -0.3679 | 0.0768 | 1.69E-06 | IL1b_MSUC16_PBMC_24h | 0.317351412 | 0.046622243 |
| rs36122610 | 3 | 45981341 | chr3:45981341:G:A | A | G | 0.3673 | 0.0775 | 2.18E-06 | IL22_B.burgdorferi_PBMC_7days | -0.326166181 | 0.046707191 |
| rs71327007 | 3 | 46017757 | chr3:46017757:C:T | T | C | 0.3643 | 0.0774 | 2.51E-06 | IL6_A.fumigatusconidiaSerum_PBMC_24h | -0.330634458 | 0.046757395 |
| rs34679077 | 3 | 46046500 | chr3:46046500:G:A | A | G | 0.3609 | 0.0774 | 3.13E-06 | IL1b_MSUC16_PBMC_24h | -0.317318888 | 0.046757536 |
| rs76860840 | 2 | 15618674 | chr2:15618674:C:G | C | G | 0.3536 | 0.0764 | 3.66E-06 | IL17_C.albicanshyphae_PBMC_7days | 0.310613483 | 0.046768602 |
| rs62120724 | 2 | 15593845 | chr2:15593845:G:A | A | G | -0.2986 | 0.0675 | 9.73E-06 | IL17_C.albicanshyphae_PBMC_7days | -0.299816626 | 0.046835388 |
| rs2032778 | 2 | 15596573 | chr2:15596573:G:A | A | G | -0.299 | 0.0675 | 9.48E-06 | IL17_C.albicanshyphae_PBMC_7days | -0.299816626 | 0.046835388 |
| rs13074382 | 3 | 46020348 | chr3:46020348:A:G | A | G | -0.3679 | 0.0768 | 1.69E-06 | IL22_A.fumigatusconidia_PBMC_7days | -0.390970935 | 0.046841088 |
| rs6791016 | 3 | 46033366 | chr3:46033366:G:A | A | G | 0.367 | 0.0769 | 1.80E-06 | IL22_A.fumigatusconidia_PBMC_7days | 0.391205685 | 0.04686305 |
| rs6780028 | 3 | 46033598 | chr3:46033598:A:C | A | C | -0.367 | 0.0769 | 1.80E-06 | IL22_A.fumigatusconidia_PBMC_7days | -0.391205685 | 0.04686305 |
| rs35454877 | 3 | 46017992 | chr3:46017992:T:C | T | C | -0.3679 | 0.0768 | 1.69E-06 | IL6_A.fumigatusconidiaSerum_PBMC_24h | 0.330314206 | 0.046958494 |
| rs6790866 | 3 | 46033219 | chr3:46033219:G:A | A | G | 0.367 | 0.0769 | 1.80E-06 | IL22_A.fumigatusconidia_PBMC_7days | 0.39093038 | 0.047004617 |
| rs807627 | 2 | 15639565 | chr2:15639565:T:C | T | C | 0.3105 | 0.0679 | 4.79E-06 | IL17_C.albicanshyphae_PBMC_7days | 0.299327656 | 0.047006367 |
| rs807628 | 2 | 15638176 | chr2:15638176:A:G | A | G | 0.3095 | 0.0678 | 5.04E-06 | IL17_C.albicanshyphae_PBMC_7days | 0.299308305 | 0.0470464 |
| rs35814488 | 3 | 46032431 | chr3:46032431:A:G | A | G | -0.3672 | 0.0769 | 1.77E-06 | IL22_A.fumigatusconidia_PBMC_7days | -0.390654132 | 0.047146811 |
| rs13097556 | 3 | 46020505 | chr3:46020505:T:C | T | C | -0.3679 | 0.0768 | 1.69E-06 | IL22_A.fumigatusconidia_PBMC_7days | -0.390377343 | 0.047155138 |
| rs4595081 | 7 | 158215231 | chr7:158215231:G:T | T | G | 0.2285 | 0.0507 | 6.62E-06 | IL6_B.burgdorferi_PBMC_24h | 0.176715647 | 0.047172556 |
| rs10784278 | 12 | 62042165 | chr12:62042165:T:A | A | T | -0.2836 | 0.06 | 2.32E-06 | IL6_MTB_PBMC_24h | -0.223468552 | 0.047237232 |
| rs34774687 | 3 | 46031822 | chr3:46031822:T:C | T | C | -0.367 | 0.0769 | 1.80E-06 | IL22_A.fumigatusconidia_PBMC_7days | -0.390376944 | 0.047289634 |
| rs34442130 | 3 | 45983037 | chr3:45983037:T:A | A | T | 0.3674 | 0.0775 | 2.16E-06 | IL22_B.burgdorferi_PBMC_7days | -0.325295026 | 0.047299832 |
| rs73833520 | 3 | 46016375 | chr3:46016375:A:G | A | G | -0.3679 | 0.0768 | 1.69E-06 | IFNy_A.fumigatusconidia_PBMC_7days | -0.35150761 | 0.047317806 |
| rs13075758 | 3 | 45983556 | chr3:45983556:G:A | A | G | 0.3672 | 0.0775 | 2.19E-06 | IL22_B.burgdorferi_PBMC_7days | -0.325209807 | 0.047345045 |
| rs4506155 | 7 | 158211289 | chr7:158211289:G:A | A | G | 0.2258 | 0.0508 | 8.62E-06 | IL6_B.burgdorferi_PBMC_24h | 0.176625268 | 0.047353818 |
| rs13060287 | 3 | 46030376 | chr3:46030376:A:G | A | G | -0.3671 | 0.0769 | 1.79E-06 | IL22_A.fumigatusconidia_PBMC_7days | -0.390141244 | 0.047398586 |
| rs34619093 | 3 | 46031027 | chr3:46031027:G:A | A | G | 0.3625 | 0.0775 | 2.89E-06 | IL22_A.fumigatusconidia_PBMC_7days | 0.390098818 | 0.047433087 |
| rs68087193 | 3 | 46031138 | chr3:46031138:C:T | T | C | 0.3673 | 0.0769 | 1.76E-06 | IL22_A.fumigatusconidia_PBMC_7days | 0.390098818 | 0.047433087 |
| rs2102055 | 3 | 46031212 | chr3:46031212:G:A | A | G | 0.3591 | 0.0767 | 2.83E-06 | IL22_A.fumigatusconidia_PBMC_7days | 0.390098818 | 0.047433087 |
| rs2102056 | 3 | 46031276 | chr3:46031276:G:A | A | G | 0.3671 | 0.0769 | 1.78E-06 | IL22_A.fumigatusconidia_PBMC_7days | 0.390098818 | 0.047433087 |
| rs2088690 | 3 | 46031347 | chr3:46031347:C:T | T | C | 0.3671 | 0.0769 | 1.78E-06 | IL22_A.fumigatusconidia_PBMC_7days | 0.390098818 | 0.047433087 |
| rs1491951 | 3 | 46100352 | chr3:46100352:G:A | A | G | 0.3485 | 0.0777 | 7.30E-06 | IL6_A.fumigatusconidiaSerum_PBMC_24h | -0.32508073 | 0.047459586 |
| rs13082697 | 3 | 46030271 | chr3:46030271:T:C | T | C | -0.3671 | 0.0769 | 1.78E-06 | IL22_A.fumigatusconidia_PBMC_7days | -0.389861195 | 0.047542869 |
| rs10784279 | 12 | 62044016 | chr12:62044016:C:T | T | C | -0.2778 | 0.0599 | 3.50E-06 | IL6_MTB_PBMC_24h | -0.22303281 | 0.047654754 |
| rs34562820 | 3 | 46029589 | chr3:46029589:G:A | A | G | 0.3625 | 0.0775 | 2.87E-06 | IL22_A.fumigatusconidia_PBMC_7days | 0.389580215 | 0.047687787 |
| rs13093179 | 3 | 46104822 | chr3:46104822:G:T | T | G | 0.3557 | 0.0779 | 4.91E-06 | IL6_LPS100ng_PBMC_24h | -0.200645955 | 0.047727063 |
| rs34438204 | 3 | 46039814 | chr3:46039814:T:C | T | C | -0.3609 | 0.0774 | 3.13E-06 | IL1b_MSUC16_PBMC_24h | 0.315759203 | 0.047751476 |
| rs59166269 | 3 | 46028845 | chr3:46028845:C:G | C | G | -0.3672 | 0.0769 | 1.77E-06 | IL22_A.fumigatusconidia_PBMC_7days | -0.389298307 | 0.047833344 |
| rs57437758 | 3 | 46028861 | chr3:46028861:A:C | A | C | -0.3672 | 0.0769 | 1.77E-06 | IL22_A.fumigatusconidia_PBMC_7days | -0.389298307 | 0.047833344 |
| rs13090194 | 3 | 46028081 | chr3:46028081:A:C | A | C | -0.3672 | 0.0769 | 1.77E-06 | IL22_A.fumigatusconidia_PBMC_7days | -0.389015473 | 0.04797954 |
| rs13263570 | 8 | 121845942 | chr8:121845942:G:A | A | G | -0.2298 | 0.0516 | 8.50E-06 | IL1b_Influenza_PBMC_24h | 0.346223167 | 0.04801318 |
| rs6948273 | 7 | 158222765 | chr7:158222765:T:C | T | C | -0.2286 | 0.0509 | 6.99E-06 | IFNy_C.albicanshyphae_PBMC_7days | -0.244783732 | 0.048103091 |
| rs34492478 | 3 | 46097350 | chr3:46097350:A:T | A | T | -0.3562 | 0.078 | 4.95E-06 | IL22_A.fumigatusconidia_PBMC_7days | -0.388384674 | 0.048127859 |
| rs71327010 | 3 | 46027343 | chr3:46027343:G:T | T | G | 0.3625 | 0.0774 | 2.83E-06 | IL22_A.fumigatusconidia_PBMC_7days | 0.388447038 | 0.048273861 |
| rs13381043 | 18 | 3724602 | chr18:3724602:G:A | A | G | -0.325 | 0.0613 | 1.13E-07 | IL1b_B.fragilis_PBMC_24h | -0.207820067 | 0.048315939 |
| rs35159820 | 3 | 46026981 | chr3:46026981:A:G | A | G | -0.3673 | 0.0769 | 1.76E-06 | IL22_A.fumigatusconidia_PBMC_7days | -0.388161442 | 0.04842199 |
| rs2088692 | 3 | 46035840 | chr3:46035840:A:G | A | G | -0.3654 | 0.0768 | 1.96E-06 | IL1b_MSUC16_PBMC_24h | 0.314663373 | 0.048445801 |
| rs911360 | 6 | 20314488 | chr6:20314488:C:T | T | C | 0.2673 | 0.058 | 4.09E-06 | IL1b_C.albicanshyphae_PBMC_24h | -0.179803991 | 0.048467408 |
| rs6930922 | 6 | 20314659 | chr6:20314659:C:T | T | C | 0.265 | 0.058 | 4.86E-06 | IL1b_C.albicanshyphae_PBMC_24h | -0.179504329 | 0.048561172 |
| rs35161099 | 3 | 46026015 | chr3:46026015:T:G | T | G | -0.3625 | 0.0774 | 2.83E-06 | IL22_A.fumigatusconidia_PBMC_7days | -0.387874931 | 0.048570768 |

| | | | | | | | | | | | |
|---|---|---|---|---|---|---|---|---|---|---|---|
| rs4595081 | 7 | 158215231 | chr7:158215231:G:T | T | G | 0.2285 | 0.0507 | 6.62E-06 | IFNy_C.albicanshyphae_PBMC_7days | 0.244292393 | 0.048635276 |
| rs35334665 | 3 | 46019726 | chr3:46019726:T:C | T | C | -0.3632 | 0.0774 | 2.70E-06 | IL1b_MSUC16_PBMC_24h | 0.31424049 | 0.048636907 |
| rs6948273 | 7 | 158222765 | chr7:158222765:T:C | T | C | -0.2286 | 0.0509 | 6.99E-06 | IL6_B.burgdorferi_PBMC_24h | -0.175475611 | 0.048672665 |
| rs36040135 | 3 | 46019812 | chr3:46019812:A:G | A | G | -0.3679 | 0.0768 | 1.69E-06 | IL1b_MSUC16_PBMC_24h | 0.314008247 | 0.048784928 |
| rs4506155 | 7 | 158211289 | chr7:158211289:G:A | A | G | 0.2258 | 0.0508 | 8.62E-06 | IFNy_C.albicanshyphae_PBMC_7days | 0.2441795 | 0.048808828 |
| rs6791016 | 3 | 46033366 | chr3:46033366:G:A | A | G | 0.367 | 0.0769 | 1.80E-06 | IL1b_MSUC16_PBMC_24h | -0.314018313 | 0.048862662 |
| rs6780028 | 3 | 46033598 | chr3:46033598:A:C | A | C | -0.367 | 0.0769 | 1.80E-06 | IL1b_MSUC16_PBMC_24h | 0.314018313 | 0.048862662 |
| rs6790866 | 3 | 46033219 | chr3:46033219:G:A | A | G | 0.367 | 0.0769 | 1.80E-06 | IL1b_MSUC16_PBMC_24h | -0.313856336 | 0.048965227 |
| rs35814488 | 3 | 46032431 | chr3:46032431:A:G | A | G | -0.3672 | 0.0769 | 1.77E-06 | IL1b_MSUC16_PBMC_24h | 0.313693586 | 0.049068253 |
| rs34774687 | 3 | 46031822 | chr3:46031822:T:C | T | C | -0.367 | 0.0769 | 1.80E-06 | IL1b_MSUC16_PBMC_24h | 0.313530066 | 0.049171741 |
| rs35930050 | 3 | 46024271 | chr3:46024271:G:A | A | G | 0.3681 | 0.0768 | 1.67E-06 | IL22_A.fumigatusconidia_PBMC_7days | 0.386719787 | 0.049172402 |
| rs34584867 | 3 | 46024471 | chr3:46024471:G:A | A | G | 0.3681 | 0.0768 | 1.67E-06 | IL22_A.fumigatusconidia_PBMC_7days | 0.386719787 | 0.049172402 |
| rs62120724 | 2 | 15593845 | chr2:15593845:G:A | A | G | -0.2986 | 0.0675 | 9.73E-06 | IFNy_Cryptococcus_PBMC_7days | 0.250628876 | 0.049236833 |
| rs2032778 | 2 | 15596573 | chr2:15596573:G:A | A | G | -0.299 | 0.0675 | 9.48E-06 | IFNy_Cryptococcus_PBMC_7days | 0.250628876 | 0.049236833 |
| rs34619093 | 3 | 46031027 | chr3:46031027:G:A | A | G | 0.3625 | 0.0775 | 2.89E-06 | IL1b_MSUC16_PBMC_24h | -0.313365776 | 0.049275693 |
| rs68087193 | 3 | 46031138 | chr3:46031138:C:T | T | C | 0.3673 | 0.0769 | 1.76E-06 | IL1b_MSUC16_PBMC_24h | -0.313365776 | 0.049275693 |
| rs2102055 | 3 | 46031212 | chr3:46031212:G:A | A | G | 0.3591 | 0.0767 | 2.83E-06 | IL1b_MSUC16_PBMC_24h | -0.313365776 | 0.049275693 |
| rs2102056 | 3 | 46031276 | chr3:46031276:G:A | A | G | 0.3671 | 0.0769 | 1.78E-06 | IL1b_MSUC16_PBMC_24h | -0.313365776 | 0.049275693 |
| rs2088690 | 3 | 46031347 | chr3:46031347:C:T | T | C | 0.3671 | 0.0769 | 1.78E-06 | IL1b_MSUC16_PBMC_24h | -0.313365776 | 0.049275693 |
| rs35334665 | 3 | 46019726 | chr3:46019726:T:C | T | C | -0.3632 | 0.0774 | 2.70E-06 | IL6_A.fumigatusconidiaSerum_PBMC_24h | 0.326589507 | 0.049321313 |
| rs13060287 | 3 | 46030376 | chr3:46030376:A:G | A | G | -0.3671 | 0.0769 | 1.79E-06 | IL1b_MSUC16_PBMC_24h | 0.313283572 | 0.049325028 |
| rs13082697 | 3 | 46030271 | chr3:46030271:T:C | T | C | -0.3671 | 0.0769 | 1.78E-06 | IL1b_MSUC16_PBMC_24h | 0.313117732 | 0.049429818 |
| rs13074382 | 3 | 46020348 | chr3:46020348:A:G | A | G | -0.3679 | 0.0768 | 1.69E-06 | IL1b_MSUC16_PBMC_24h | 0.312990026 | 0.049450412 |
| rs36040135 | 3 | 46019812 | chr3:46019812:A:G | A | G | -0.3679 | 0.0768 | 1.69E-06 | IL6_A.fumigatusconidiaSerum_PBMC_24h | 0.326357893 | 0.049463788 |
| rs34562820 | 3 | 46029589 | chr3:46029589:G:A | A | G | 0.3625 | 0.0775 | 2.87E-06 | IL1b_MSUC16_PBMC_24h | -0.312951127 | 0.049535074 |
| rs10156036 | 7 | 158224085 | chr7:158224085:A:C | A | C | -0.2322 | 0.0509 | 4.96E-06 | IL6_B.burgdorferi_PBMC_24h | -0.174881534 | 0.049549226 |
| rs2191031 | 3 | 45869378 | chr3:45869378:G:A | A | G | 0.2758 | 0.0613 | 6.80E-06 | IL6_Cryptococcus_PBMC_24h | -0.468162385 | 0.049570312 |
| rs59166269 | 3 | 46028845 | chr3:46028845:C:G | C | G | -0.3672 | 0.0769 | 1.77E-06 | IL1b_MSUC16_PBMC_24h | 0.31278376 | 0.049640799 |
| rs57437758 | 3 | 46028861 | chr3:46028861:A:C | A | C | -0.3672 | 0.0769 | 1.77E-06 | IL1b_MSUC16_PBMC_24h | 0.31278376 | 0.049640799 |
| rs35669129 | 3 | 46013758 | chr3:46013758:G:A | A | G | 0.3746 | 0.0768 | 1.08E-06 | IL22_A.fumigatusconidia_PBMC_7days | 0.395207763 | 0.049650748 |
| rs13097556 | 3 | 46020505 | chr3:46020505:T:C | T | C | -0.3679 | 0.0768 | 1.69E-06 | IL1b_MSUC16_PBMC_24h | 0.312680104 | 0.049653952 |
| rs911360 | 6 | 20314488 | chr6:20314488:C:T | T | C | 0.2673 | 0.058 | 4.09E-06 | TNFA_MSUC16_PBMC_24h | -0.154729659 | 0.04971196 |
| rs13090194 | 3 | 46028081 | chr3:46028081:A:C | A | C | -0.3672 | 0.0769 | 1.77E-06 | IL1b_MSUC16_PBMC_24h | 0.312615631 | 0.049746992 |
| rs876668 | 3 | 46022261 | chr3:46022261:T:C | T | C | -0.368 | 0.0768 | 1.68E-06 | IL22_A.fumigatusconidia_PBMC_7days | -0.385550213 | 0.049784575 |
| rs35751180 | 3 | 46014681 | chr3:46014681:C:T | T | C | 0.3731 | 0.0774 | 1.42E-06 | IL22_A.fumigatusconidia_PBMC_7days | 0.394439646 | 0.049914823 |
| rs71327014 | 3 | 46059974 | chr3:46059974:C:T | T | C | 0.3556 | 0.0773 | 4.25E-06 | IL6_A.fumigatusconidiaSerum_PBMC_24h | -0.32627508 | 0.049936897 |
| rs71327015 | 3 | 46060681 | chr3:46060681:G:C | C | G | 0.3585 | 0.0773 | 3.57E-06 | IL6_A.fumigatusconidiaSerum_PBMC_24h | -0.32627508 | 0.049936897 |
| rs34047915 | 3 | 46058999 | chr3:46058999:C:T | T | C | 0.3555 | 0.0773 | 4.29E-06 | IL6_A.fumigatusconidiaSerum_PBMC_24h | -0.326254094 | 0.049955725 |
| rs71327010 | 3 | 46027343 | chr3:46027343:G:T | T | G | 0.3625 | 0.0774 | 2.83E-06 | IL1b_MSUC16_PBMC_24h | -0.312277096 | 0.049960792 |
| rs35420565 | 3 | 46058646 | chr3:46058646:C:G | C | G | -0.3556 | 0.0773 | 4.26E-06 | IL6_A.fumigatusconidiaSerum_PBMC_24h | 0.326232256 | 0.049974859 |
| rs12610495 | 19 | 4717660 | chr19:4717660:A:G | A | G | -0.2534 | 0.0556 | 5.20E-06 | IL6_S.typhimurium_macroPG_24h | -0.186452329 | 0.005410048 |
| rs41264195 | 2 | 15606096 | chr2:15606096:C:T | T | C | -0.3359 | 0.0754 | 8.49E-06 | IL6_MTB_macroPG_24h | -0.35796968 | 0.011443609 |
| rs79808866 | 2 | 15604860 | chr2:15604860:T:G | T | G | 0.3357 | 0.0754 | 8.59E-06 | IL6_MTB_macroPG_24h | 0.357886923 | 0.011461963 |
| rs3770462 | 2 | 15609200 | chr2:15609200:T:G | T | G | 0.3409 | 0.0757 | 6.74E-06 | IL6_MTB_macroPG_24h | 0.357831548 | 0.011468203 |
| rs4668941 | 2 | 15610007 | chr2:15610007:T:G | T | G | 0.3359 | 0.0754 | 8.51E-06 | IL6_MTB_macroPG_24h | 0.357831548 | 0.011468203 |
| rs76860840 | 2 | 15618674 | chr2:15618674:C:G | C | G | 0.3536 | 0.0764 | 3.66E-06 | IL6_MTB_macroPG_24h | 0.360873523 | 0.011777271 |
| rs807635 | 2 | 15629483 | chr2:15629483:C:T | T | C | -0.3579 | 0.0765 | 2.85E-06 | IL6_MTB_macroPG_24h | -0.34791002 | 0.015269133 |
| rs807636 | 2 | 15627828 | chr2:15627828:C:T | T | C | -0.365 | 0.0772 | 2.25E-06 | IL6_MTB_macroPG_24h | -0.348205972 | 0.015309964 |
| rs2302934 | 2 | 15613155 | chr2:15613155:C:G | C | G | 0.3393 | 0.076 | 8.11E-06 | IL6_MTB_macroPG_24h | 0.348048027 | 0.015616342 |
| rs12616567 | 2 | 15615498 | chr2:15615498:A:C | A | C | 0.3427 | 0.0762 | 6.85E-06 | IL6_MTB_macroPG_24h | 0.348048027 | 0.015616342 |
| rs76880770 | 2 | 15618488 | chr2:15618488:G:A | A | G | -0.3455 | 0.0767 | 6.62E-06 | IL6_MTB_macroPG_24h | -0.362962283 | 0.016405157 |
| rs12619874 | 2 | 15586997 | chr2:15586997:G:C | C | G | -0.3204 | 0.0665 | 1.46E-06 | IL6_MTB_macroPG_24h | -0.306595473 | 0.021715748 |
| rs9739956 | 12 | 62065731 | chr12:62065731:T:C | T | C | 0.2934 | 0.0598 | 9.45E-07 | IL6_LPS_macroPG_24h | 0.219224817 | 0.022127953 |
| rs10877786 | 12 | 62070198 | chr12:62070198:T:G | T | G | 0.2964 | 0.0597 | 6.89E-07 | IL6_LPS_macroPG_24h | 0.217752146 | 0.023050147 |
| rs2302929 | 2 | 15595524 | chr2:15595524:T:C | T | C | 0.2976 | 0.0672 | 9.54E-06 | IL6_MTB_macroPG_24h | 0.302600688 | 0.026459528 |
| rs2080811 | 2 | 15584186 | chr2:15584186:G:A | A | G | -0.329 | 0.0676 | 1.12E-06 | IL6_MTB_macroPG_24h | -0.299777242 | 0.027257401 |
| rs12619108 | 2 | 15586369 | chr2:15586369:G:A | A | G | -0.315 | 0.0658 | 1.68E-06 | IL6_MTB_macroPG_24h | -0.297628452 | 0.028164786 |
| rs2191031 | 3 | 45869378 | chr3:45869378:G:A | A | G | 0.2758 | 0.0613 | 6.80E-06 | IL6_MTB_macroPG_24h | -0.289815557 | 0.029972299 |
| rs10877787 | 12 | 62072981 | chr12:62072981:A:G | A | G | 0.29 | 0.0604 | 1.58E-06 | IL6_LPS_macroPG_24h | 0.205914173 | 0.032188583 |
| rs11174294 | 12 | 62072129 | chr12:62072129:C:T | T | C | -0.29 | 0.0604 | 1.59E-06 | IL6_LPS_macroPG_24h | -0.20585868 | 0.032225353 |
| rs10506436 | 12 | 62028786 | chr12:62028786:T:A | A | T | -0.2811 | 0.06 | 2.84E-06 | IL6_LPS_macroPG_24h | -0.20630647 | 0.032251266 |
| rs9706516 | 12 | 62074403 | chr12:62074403:C:A | A | C | -0.2923 | 0.0604 | 1.30E-06 | IL6_LPS_macroPG_24h | -0.205312246 | 0.032743256 |
| rs9739464 | 12 | 62037992 | chr12:62037992:A:T | A | T | 0.2846 | 0.06 | 2.10E-06 | IL6_LPS_macroPG_24h | 0.205739893 | 0.032842858 |
| rs62122279 | 2 | 15621842 | chr2:15621842:T:C | T | C | 0.3047 | 0.0678 | 6.91E-06 | IL6_MTB_macroPG_24h | 0.295593392 | 0.032969364 |
| rs4668946 | 2 | 15624494 | chr2:15624494:A:G | A | G | 0.3109 | 0.0679 | 4.72E-06 | IL6_MTB_macroPG_24h | 0.295537372 | 0.032979894 |
| rs12614308 | 2 | 15624914 | chr2:15624914:G:A | A | G | -0.3109 | 0.0679 | 4.72E-06 | IL6_MTB_macroPG_24h | -0.295537372 | 0.032979894 |
| rs10747951 | 12 | 62075677 | chr12:62075677:C:T | T | C | -0.29 | 0.0604 | 1.58E-06 | IL6_LPS_macroPG_24h | -0.20456723 | 0.033433149 |
| rs807628 | 2 | 15638176 | chr2:15638176:A:G | A | G | 0.3095 | 0.0678 | 5.04E-06 | IL6_MTB_macroPG_24h | 0.294022147 | 0.033442995 |
| rs807627 | 2 | 15639565 | chr2:15639565:T:C | T | C | 0.3105 | 0.0679 | 4.79E-06 | IL6_MTB_macroPG_24h | 0.29395409 | 0.033463088 |
| rs10784278 | 12 | 62042165 | chr12:62042165:T:A | A | T | -0.2836 | 0.06 | 2.32E-06 | IL6_LPS_macroPG_24h | -0.204538929 | 0.033661966 |
| rs10784279 | 12 | 62044016 | chr12:62044016:C:T | T | C | -0.2778 | 0.0599 | 3.50E-06 | IL6_LPS_macroPG_24h | -0.204138101 | 0.033994483 |
| rs10506432 | 12 | 62077116 | chr12:62077116:A:C | A | C | 0.29 | 0.0604 | 1.58E-06 | IL6_LPS_macroPG_24h | 0.203815772 | 0.034139878 |
| rs11174296 | 12 | 62077677 | chr12:62077677:C:G | C | G | 0.288 | 0.0602 | 1.75E-06 | IL6_LPS_macroPG_24h | 0.203492579 | 0.034437609 |
| rs807623 | 2 | 15642467 | chr2:15642467:C:T | T | C | -0.312 | 0.0679 | 4.30E-06 | IL6_MTB_macroPG_24h | -0.290244612 | 0.034611447 |
| rs807622 | 2 | 15642631 | chr2:15642631:A:G | A | G | 0.3099 | 0.0675 | 4.43E-06 | IL6_MTB_macroPG_24h | 0.290244612 | 0.034611447 |
| rs807620 | 2 | 15643012 | chr2:15643012:C:T | T | C | -0.312 | 0.0679 | 4.30E-06 | IL6_MTB_macroPG_24h | -0.290244612 | 0.034611447 |
| rs807618 | 2 | 15646936 | chr2:15646936:G:C | C | G | -0.3046 | 0.0673 | 6.02E-06 | IL6_MTB_macroPG_24h | -0.289234524 | 0.035247577 |
| rs62120724 | 2 | 15593845 | chr2:15593845:G:A | A | G | -0.2986 | 0.0675 | 9.73E-06 | IL6_MTB_macroPG_24h | -0.291358611 | 0.035348594 |
| rs2032778 | 2 | 15596573 | chr2:15596573:G:A | A | G | -0.299 | 0.0675 | 9.48E-06 | IL6_MTB_macroPG_24h | -0.291358611 | 0.035348594 |
| rs9738190 | 12 | 62078790 | chr12:62078790:G:A | A | G | -0.2898 | 0.0602 | 1.51E-06 | IL6_LPS_macroPG_24h | -0.201559949 | 0.0362895 |
| rs3774641 | 3 | 45896341 | chr3:45896341:G:T | T | G | 0.2741 | 0.0605 | 5.95E-06 | IL6_MTB_macroPG_24h | -0.261536666 | 0.046150976 |
| rs807615 | 2 | 15648378 | chr2:15648378:A:G | A | G | 0.309 | 0.068 | 5.51E-06 | IL6_MTB_macroPG_24h | 0.277347295 | 0.046337504 |
| rs807613 | 2 | 15649443 | chr2:15649443:C:T | T | C | -0.3086 | 0.0679 | 5.60E-06 | IL6_MTB_macroPG_24h | -0.277198107 | 0.046513753 |
| rs807611 | 2 | 15649857 | chr2:15649857:G:T | T | G | -0.3084 | 0.0672 | 4.51E-06 | IL6_MTB_macroPG_24h | -0.277088408 | 0.046622827 |
| rs12185539 | 2 | 15650052 | chr2:15650052:A:G | A | G | 0.3079 | 0.067 | 4.31E-06 | IL6_MTB_macroPG_24h | 0.276873301 | 0.046834962 |
| rs12185540 | 2 | 15650056 | chr2:15650056:A:G | A | G | 0.3058 | 0.0677 | 6.19E-06 | IL6_MTB_macroPG_24h | 0.276873301 | 0.046834962 |
| rs807610 | 2 | 15650440 | chr2:15650440:C:G | C | G | 0.3082 | 0.0673 | 4.64E-06 | IL6_MTB_macroPG_24h | 0.27647322 | 0.047212679 |
| rs807607 | 2 | 15651223 | chr2:15651223:G:C | C | G | -0.3087 | 0.0679 | 5.52E-06 | IL6_MTB_macroPG_24h | -0.275141209 | 0.048352656 |
| rs807606 | 2 | 15651602 | chr2:15651602:C:A | A | C | -0.3042 | 0.0679 | 7.55E-06 | IL6_MTB_macroPG_24h | -0.274566287 | 0.04876936 |
| rs807602 | 2 | 15652485 | chr2:15652485:C:A | A | C | -0.3021 | 0.0679 | 8.60E-06 | IL6_MTB_macroPG_24h | -0.274327863 | 0.048935606 |
| rs807605 | 2 | 15652128 | chr2:15652128:G:A | A | G | -0.3007 | 0.0679 | 9.38E-06 | IL6_MTB_macroPG_24h | -0.274090719 | 0.04912291 |
| rs807604 | 2 | 15652384 | chr2:15652384:T:A | A | T | -0.3037 | 0.0679 | 7.64E-06 | IL6_MTB_macroPG_24h | -0.274047058 | 0.049155604 |
| rs7152677 | 14 | 31953540 | chr14:31953540:G:C | C | G | 0.2637 | 0.0548 | 1.52E-06 | IL6_LPS_WB_48h | -0.141736232 | 0.001778059 |
| rs1455664 | 4 | 179425171 | chr4:179425171:T:C | T | C | -0.3158 | 0.0714 | 9.77E-06 | TNFA_LPS_WB_48h | 0.271197145 | 0.002048745 |
| rs1455662 | 4 | 179423719 | chr4:179423719:A:G | A | G | -0.3156 | 0.0713 | 9.60E-06 | TNFA_LPS_WB_48h | 0.269592911 | 0.002151734 |
| rs1455663 | 4 | 179425076 | chr4:179425076:A:G | A | G | -0.3156 | 0.0714 | 9.87E-06 | TNFA_LPS_WB_48h | 0.26705588 | 0.002471713 |
| rs7152677 | 14 | 31953540 | chr14:31953540:G:C | C | G | 0.2637 | 0.0548 | 1.52E-06 | IFNy_S.aureus_WB_48h | -0.403829284 | 0.004761632 |
| rs7152677 | 14 | 31953540 | chr14:31953540:G:C | C | G | 0.2637 | 0.0548 | 1.52E-06 | IFNy_LPS_WB_48h | -0.335163222 | 0.007448562 |
| rs1491950 | 3 | 46096615 | chr3:46096615:G:A | A | G | 0.3544 | 0.078 | 5.55E-06 | IFNy_C.conidiaHK_WB_48h | -0.496249452 | 0.007539546 |
| rs13093179 | 3 | 46104822 | chr3:46104822:G:T | T | G | 0.3557 | 0.0779 | 4.91E-06 | IFNy_C.conidiaHK_WB_48h | -0.494393844 | 0.008790006 |
| rs34452002 | 3 | 46101695 | chr3:46101695:C:T | T | C | 0.3556 | 0.0779 | 4.95E-06 | IFNy_C.conidiaHK_WB_48h | -0.485579185 | 0.009131036 |
| rs34093271 | 3 | 46103489 | chr3:46103489:G:T | T | G | 0.3558 | 0.0779 | 4.89E-06 | IFNy_C.conidiaHK_WB_48h | -0.485678431 | 0.009150791 |
| rs34460587 | 3 | 46100972 | chr3:46100972:C:T | T | C | 0.3557 | 0.0779 | 4.93E-06 | IFNy_C.conidiaHK_WB_48h | -0.483617058 | 0.009386987 |
| rs1455664 | 4 | 179425171 | chr4:179425171:T:C | T | C | -0.3158 | 0.0714 | 9.77E-06 | IL1b_LPS_WB_48h | 0.163049965 | 0.009423371 |
| rs13091868 | 3 | 46081551 | chr3:46081551:A:G | A | G | -0.3538 | 0.0773 | 4.72E-06 | IFNy_C.conidiaHK_WB_48h | 0.481454657 | 0.009664521 |
| rs13092030 | 3 | 46081563 | chr3:46081563:C:T | T | C | 0.354 | 0.0773 | 4.66E-06 | IFNy_C.conidiaHK_WB_48h | -0.481454657 | 0.009664521 |
| rs17215981 | 3 | 46082926 | chr3:46082926:A:G | A | G | -0.3547 | 0.0773 | 4.47E-06 | IFNy_C.conidiaHK_WB_48h | 0.481454657 | 0.009664521 |
| rs13089554 | 3 | 46086065 | chr3:46086065:G:A | A | G | 0.3547 | 0.0774 | 4.61E-06 | IFNy_C.conidiaHK_WB_48h | -0.481454657 | 0.009664521 |
| rs13095717 | 3 | 46087050 | chr3:46087050:G:A | A | G | 0.3548 | 0.0774 | 4.58E-06 | IFNy_C.conidiaHK_WB_48h | -0.481454657 | 0.009664521 |
| rs13075528 | 3 | 46087217 | chr3:46087217:T:C | T | C | -0.3549 | 0.0775 | 4.63E-06 | IFNy_C.conidiaHK_WB_48h | 0.481454657 | 0.009664521 |
| rs13095602 | 3 | 46087221 | chr3:46087221:A:G | A | G | -0.3547 | 0.0774 | 4.60E-06 | IFNy_C.conidiaHK_WB_48h | 0.481454657 | 0.009664521 |
| rs71327023 | 3 | 46089733 | chr3:46089733:C:G | C | G | -0.3539 | 0.0777 | 5.19E-06 | IFNy_C.conidiaHK_WB_48h | 0.481454657 | 0.009664521 |
| rs13063527 | 3 | 46094198 | chr3:46094198:G:A | A | G | 0.3561 | 0.0777 | 4.51E-06 | IFNy_C.conidiaHK_WB_48h | -0.481454657 | 0.009664521 |
| rs13068570 | 3 | 46094946 | chr3:46094946:A:G | A | G | -0.3561 | 0.0777 | 4.61E-06 | IFNy_C.conidiaHK_WB_48h | 0.481454657 | 0.009664521 |
| rs4373103 | 3 | 46096963 | chr3:46096963:G:A | A | G | 0.3547 | 0.078 | 5.42E-06 | IFNy_C.conidiaHK_WB_48h | -0.481454657 | 0.009664521 |
| rs71327024 | 3 | 46098581 | chr3:46098581:G:T | T | G | 0.3547 | 0.078 | 5.37E-06 | IFNy_C.conidiaHK_WB_48h | -0.481454657 | 0.009664521 |
| rs71327017 | 3 | 46069721 | chr3:46069721:T:C | T | C | -0.3576 | 0.0774 | 3.80E-06 | IFNy_C.conidiaHK_WB_48h | 0.481391078 | 0.009675192 |
| rs13072267 | 3 | 46074518 | chr3:46074518:T:A | A | T | 0.3543 | 0.0773 | 4.57E-06 | IFNy_C.conidiaHK_WB_48h | -0.481391078 | 0.009675192 |
| rs36057789 | 3 | 46075604 | chr3:46075604:A:G | A | G | -0.3551 | 0.0773 | 4.36E-06 | IFNy_C.conidiaHK_WB_48h | 0.481391078 | 0.009675192 |
| rs76647202 | 3 | 46076468 | chr3:46076468:A:G | A | G | -0.3542 | 0.0773 | 4.60E-06 | IFNy_C.conidiaHK_WB_48h | 0.481391078 | 0.009675192 |
| rs71327021 | 3 | 46079025 | chr3:46079025:A:G | A | G | -0.3542 | 0.0773 | 4.61E-06 | IFNy_C.conidiaHK_WB_48h | 0.481391078 | 0.009675192 |
| rs71327025 | 3 | 46106012 | chr3:46106012:A:G | A | G | -0.3556 | 0.0779 | 4.96E-06 | IFNy_C.conidiaHK_WB_48h | 0.483197748 | 0.009714058 |
| rs1455662 | 4 | 179423719 | chr4:179423719:A:G | A | G | -0.3156 | 0.0713 | 9.60E-06 | IL1b_LPS_WB_48h | 0.162020227 | 0.009795172 |
| rs35539222 | 3 | 46112780 | chr3:46112780:T:C | T | C | -0.3556 | 0.0779 | 4.96E-06 | IFNy_C.conidiaHK_WB_48h | 0.482576474 | 0.009846329 |
| rs35110864 | 3 | 46112965 | chr3:46112965:G:A | A | G | 0.3555 | 0.0779 | 4.99E-06 | IFNy_C.conidiaHK_WB_48h | -0.482576474 | 0.009846329 |
| rs34492478 | 3 | 46097350 | chr3:46097350:A:T | A | T | -0.3562 | 0.078 | 4.95E-06 | IFNy_C.conidiaHK_WB_48h | 0.480277697 | 0.010023573 |
| rs1455663 | 4 | 179425076 | chr4:179425076:A:G | A | G | -0.3156 | 0.0714 | 9.87E-06 | IL1b_LPS_WB_48h | 0.162154588 | 0.010036304 |
| rs12610495 | 19 | 4717660 | chr19:4717660:A:G | A | G | -0.2534 | 0.0556 | 5.20E-06 | IFNy_S.aureus_WB_48h | -0.324056859 | 0.010487349 |
| rs11627388 | 14 | 31953140 | chr14:31953140:A:C | A | C | -0.2482 | 0.0533 | 3.19E-06 | IFNy_S.aureus_WB_48h | 0.342763061 | 0.011741732 |
| rs71327014 | 3 | 46059974 | chr3:46059974:C:T | T | C | 0.3556 | 0.0773 | 4.25E-06 | IFNy_C.conidiaHK_WB_48h | -0.471717127 | 0.011749086 |
| rs71327015 | 3 | 46060681 | chr3:46060681:G:C | C | G | 0.3585 | 0.0773 | 3.57E-06 | IFNy_C.conidiaHK_WB_48h | -0.471717127 | 0.011749086 |
| rs35516580 | 3 | 46066006 | chr3:46066006:A:G | A | G | -0.3562 | 0.0773 | 4.13E-06 | IFNy_C.conidiaHK_WB_48h | 0.471642303 | 0.011760739 |
| rs34766614 | 3 | 46066109 | chr3:46066109:A:G | A | G | -0.3484 | 0.0775 | 6.99E-06 | IFNy_C.conidiaHK_WB_48h | 0.471642303 | 0.011760739 |
| rs34127208 | 3 | 46062188 | chr3:46062188:G:T | T | G | 0.3554 | 0.0773 | 4.31E-06 | IFNy_C.conidiaHK_WB_48h | -0.47157349 | 0.011773915 |
| rs34047915 | 3 | 46058999 | chr3:46058999:C:T | T | C | 0.3555 | 0.0773 | 4.29E-06 | IFNy_C.conidiaHK_WB_48h | -0.471387125 | 0.011808996 |
| rs35420565 | 3 | 46058646 | chr3:46058646:C:G | C | G | -0.3556 | 0.0773 | 4.26E-06 | IFNy_C.conidiaHK_WB_48h | 0.471055876 | 0.011869289 |
| rs13096741 | 3 | 46057662 | chr3:46057662:C:T | T | C | 0.3556 | 0.0773 | 4.27E-06 | IFNy_C.conidiaHK_WB_48h | -0.469718457 | 0.012114335 |
| rs7152677 | 14 | 31953540 | chr14:31953540:G:C | C | G | 0.2637 | 0.0548 | 1.52E-06 | TNFA_LPS_WB_48h | -0.176204975 | 0.012143742 |
| rs13381043 | 18 | 3724602 | chr18:3724602:G:A | A | G | -0.325 | 0.0613 | 1.13E-07 | TNFA_PHA_WB_48h | 0.204485402 | 0.012147652 |
| rs34718164 | 3 | 46057034 | chr3:46057034:C:G | C | G | -0.3556 | 0.0773 | 4.25E-06 | IFNy_C.conidiaHK_WB_48h | 0.469042331 | 0.012239216 |
| rs77399277 | 3 | 46055306 | chr3:46055306:T:C | T | C | -0.3556 | 0.0773 | 4.25E-06 | IFNy_C.conidiaHK_WB_48h | 0.46663733 | 0.012689 |
| rs34340587 | 3 | 46054551 | chr3:46054551:G:A | A | G | 0.3542 | 0.0773 | 4.63E-06 | IFNy_C.conidiaHK_WB_48h | -0.466012773 | 0.012808771 |
| rs35772789 | 3 | 46053612 | chr3:46053612:A:G | A | G | -0.3544 | 0.0773 | 4.52E-06 | IFNy_C.conidiaHK_WB_48h | 0.465024022 | 0.012995844 |
| rs34863575 | 3 | 46052571 | chr3:46052571:A:G | A | G | -0.3543 | 0.0773 | 4.53E-06 | IFNy_C.conidiaHK_WB_48h | 0.463599066 | 0.013270024 |
| rs74586549 | 3 | 45884551 | chr3:45884551:C:T | T | C | 0.3443 | 0.0768 | 7.34E-06 | IFNy_C.conidiaHK_WB_48h | -0.440371317 | 0.013325811 |
| rs3774641 | 3 | 45896341 | chr3:45896341:G:T | T | G | 0.2741 | 0.0605 | 5.95E-06 | IL1b_C.conidiaHK_WB_48h | -0.225637005 | 0.014161258 |
| rs34679077 | 3 | 46046500 | chr3:46046500:G:A | A | G | 0.3609 | 0.0774 | 3.13E-06 | IFNy_C.conidiaHK_WB_48h | -0.456955307 | 0.014593042 |
| rs34438204 | 3 | 46039814 | chr3:46039814:T:C | T | C | -0.3609 | 0.0774 | 3.13E-06 | IFNy_C.conidiaHK_WB_48h | 0.452786281 | 0.015463645 |
| rs13381043 | 18 | 3724602 | chr18:3724602:G:A | A | G | -0.325 | 0.0613 | 1.13E-07 | IL1b_PHA_WB_48h | 0.191312782 | 0.015886429 |
| rs11627388 | 14 | 31953140 | chr14:31953140:A:C | A | C | -0.2482 | 0.0533 | 3.19E-06 | IFNy_PHA_WB_48h | 0.363449677 | 0.016002645 |
| rs2088692 | 3 | 46035840 | chr3:46035840:A:G | A | G | -0.3654 | 0.0768 | 1.96E-06 | IFNy_C.conidiaHK_WB_48h | 0.450018511 | 0.016058812 |
| rs6791016 | 3 | 46033366 | chr3:46033366:G:A | A | G | 0.367 | 0.0769 | 1.80E-06 | IFNy_C.conidiaHK_WB_48h | -0.448881996 | 0.016304713 |
| rs6780028 | 3 | 46033598 | chr3:46033598:A:C | A | C | -0.367 | 0.0769 | 1.80E-06 | IFNy_C.conidiaHK_WB_48h | 0.448881996 | 0.016304713 |
| rs34836513 | 3 | 46004967 | chr3:46004967:G:A | A | G | 0.3674 | 0.0772 | 1.96E-06 | IL1b_C.conidiaHK_WB_48h | -0.294462614 | 0.016379284 |
| rs6790866 | 3 | 46033219 | chr3:46033219:G:A | A | G | 0.367 | 0.0769 | 1.80E-06 | IFNy_C.conidiaHK_WB_48h | -0.44847913 | 0.01639342 |
| rs35814488 | 3 | 46032431 | chr3:46032431:A:G | A | G | -0.3672 | 0.0769 | 1.77E-06 | IFNy_C.conidiaHK_WB_48h | 0.448075214 | 0.016482677 |
| rs34774687 | 3 | 46031822 | chr3:46031822:T:C | T | C | -0.367 | 0.0769 | 1.80E-06 | IFNy_C.conidiaHK_WB_48h | 0.447670252 | 0.016572486 |
| rs34619093 | 3 | 46031027 | chr3:46031027:G:A | A | G | 0.3625 | 0.0775 | 2.89E-06 | IFNy_C.conidiaHK_WB_48h | -0.447264247 | 0.016662853 |
| rs68087193 | 3 | 46031138 | chr3:46031138:C:T | T | C | 0.3673 | 0.0769 | 1.76E-06 | IFNy_C.conidiaHK_WB_48h | -0.447264247 | 0.016662853 |
| rs2102055 | 3 | 46031212 | chr3:46031212:G:A | A | G | 0.3591 | 0.0767 | 2.83E-06 | IFNy_C.conidiaHK_WB_48h | -0.447264247 | 0.016662853 |
| rs2102056 | 3 | 46031276 | chr3:46031276:G:A | A | G | 0.3671 | 0.0769 | 1.78E-06 | IFNy_C.conidiaHK_WB_48h | -0.447264247 | 0.016662853 |
| rs2088690 | 3 | 46031347 | chr3:46031347:C:T | T | C | 0.3671 | 0.0769 | 1.78E-06 | IFNy_C.conidiaHK_WB_48h | -0.447264247 | 0.016662853 |
| rs13060287 | 3 | 46030376 | chr3:46030376:A:G | A | G | -0.3671 | 0.0769 | 1.79E-06 | IFNy_C.conidiaHK_WB_48h | 0.446927675 | 0.016738401 |
| rs13082697 | 3 | 46030271 | chr3:46030271:T:C | T | C | -0.3671 | 0.0769 | 1.78E-06 | IFNy_C.conidiaHK_WB_48h | 0.446519578 | 0.016829817 |
| rs34562820 | 3 | 46029589 | chr3:46029589:G:A | A | G | 0.3625 | 0.0775 | 2.87E-06 | IFNy_C.conidiaHK_WB_48h | -0.44611045 | 0.016921799 |
| rs35751180 | 3 | 46014681 | chr3:46014681:C:T | T | C | 0.3731 | 0.0774 | 1.42E-06 | IL1b_C.conidiaHK_WB_48h | -0.292511181 | 0.016939929 |
| rs35669129 | 3 | 46013758 | chr3:46013758:G:A | A | G | 0.3746 | 0.0768 | 1.08E-06 | IL1b_C.conidiaHK_WB_48h | -0.29272534 | 0.016946961 |
| rs59166269 | 3 | 46028845 | chr3:46028845:C:G | C | G | -0.3672 | 0.0769 | 1.77E-06 | IFNy_C.conidiaHK_WB_48h | 0.445700293 | 0.017014349 |
| rs57437758 | 3 | 46028861 | chr3:46028861:A:C | A | C | -0.3672 | 0.0769 | 1.77E-06 | IFNy_C.conidiaHK_WB_48h | 0.445700293 | 0.017014349 |
| rs13081151 | 3 | 46014224 | chr3:46014224:G:A | A | G | 0.3751 | 0.0777 | 1.38E-06 | IL1b_C.conidiaHK_WB_48h | -0.292336751 | 0.017074541 |
| rs13090194 | 3 | 46028081 | chr3:46028081:A:C | A | C | -0.3672 | 0.0769 | 1.77E-06 | IFNy_C.conidiaHK_WB_48h | 0.445289112 | 0.017107471 |
| rs13081213 | 3 | 46014417 | chr3:46014417:C:T | T | C | 0.3746 | 0.0768 | 1.08E-06 | IL1b_C.conidiaHK_WB_48h | -0.29218472 | 0.017130475 |
| rs34168660 | 3 | 46014670 | chr3:46014670:G:A | A | G | 0.375 | 0.0768 | 1.06E-06 | IL1b_C.conidiaHK_WB_48h | -0.292061967 | 0.017163978 |
| rs4362758 | 3 | 46015419 | chr3:46015419:T:A | A | T | 0.3681 | 0.0768 | 1.67E-06 | IL1b_C.conidiaHK_WB_48h | -0.291923856 | 0.017191782 |
| rs71327010 | 3 | 46027343 | chr3:46027343:G:T | T | G | 0.3625 | 0.0774 | 2.83E-06 | IFNy_C.conidiaHK_WB_48h | -0.444463693 | 0.017295441 |
| rs35159820 | 3 | 46026981 | chr3:46026981:A:G | A | G | -0.3673 | 0.0769 | 1.76E-06 | IFNy_C.conidiaHK_WB_48h | 0.444049462 | 0.017390295 |
| rs35161099 | 3 | 46026015 | chr3:46026015:T:G | T | G | -0.3625 | 0.0774 | 2.83E-06 | IFNy_C.conidiaHK_WB_48h | 0.443634222 | 0.017485734 |
| rs34754340 | 3 | 46000345 | chr3:46000345:C:T | T | C | 0.3673 | 0.0772 | 1.94E-06 | IL1b_C.conidiaHK_WB_48h | -0.291608563 | 0.017662277 |
| rs35930050 | 3 | 46024271 | chr3:46024271:G:A | A | G | 0.3681 | 0.0768 | 1.67E-06 | IFNy_C.conidiaHK_WB_48h | -0.441963249 | 0.017873394 |
| rs34584867 | 3 | 46024471 | chr3:46024471:G:A | A | G | 0.3681 | 0.0768 | 1.67E-06 | IFNy_C.conidiaHK_WB_48h | -0.441963249 | 0.017873394 |
| rs71615438 | 3 | 45998376 | chr3:45998376:A:G | A | G | -0.3666 | 0.0772 | 2.04E-06 | IL1b_C.conidiaHK_WB_48h | 0.290385696 | 0.018217781 |
| rs876668 | 3 | 46022261 | chr3:46022261:T:C | T | C | -0.368 | 0.0768 | 1.68E-06 | IFNy_C.conidiaHK_WB_48h | 0.440276439 | 0.018270658 |
| rs36023124 | 3 | 45997568 | chr3:45997568:G:C | C | G | 0.3665 | 0.0771 | 2.01E-06 | IL1b_C.conidiaHK_WB_48h | -0.289730701 | 0.018509994 |
| rs13097556 | 3 | 46020505 | chr3:46020505:T:C | T | C | -0.3679 | 0.0768 | 1.69E-06 | IFNy_C.conidiaHK_WB_48h | 0.439489965 | 0.018569874 |
| rs13074382 | 3 | 46020348 | chr3:46020348:A:G | A | G | -0.3679 | 0.0768 | 1.69E-06 | IFNy_C.conidiaHK_WB_48h | 0.439472466 | 0.018586259 |
| rs2191031 | 3 | 45869378 | chr3:45869378:G:A | A | G | 0.2758 | 0.0613 | 6.80E-06 | IL1b_C.conidiaHK_WB_48h | -0.220625792 | 0.018670509 |
| rs36040135 | 3 | 46019812 | chr3:46019812:A:G | A | G | -0.3679 | 0.0768 | 1.69E-06 | IFNy_C.conidiaHK_WB_48h | 0.438961559 | 0.018762646 |
| rs35334665 | 3 | 46019726 | chr3:46019726:T:C | T | C | -0.3632 | 0.0774 | 2.70E-06 | IFNy_C.conidiaHK_WB_48h | 0.438880361 | 0.018795265 |
| rs1491951 | 3 | 46100352 | chr3:46100352:G:A | A | G | 0.3485 | 0.0777 | 7.30E-06 | IFNy_C.conidiaHK_WB_48h | -0.432156481 | 0.018946295 |
| rs35454877 | 3 | 46017992 | chr3:46017992:T:C | T | C | -0.3679 | 0.0768 | 1.69E-06 | IFNy_C.conidiaHK_WB_48h | 0.437660562 | 0.019225591 |
| rs71327007 | 3 | 46017757 | chr3:46017757:C:T | T | C | 0.3643 | 0.0774 | 2.51E-06 | IFNy_C.conidiaHK_WB_48h | -0.437605038 | 0.019247772 |
| rs71327006 | 3 | 46017416 | chr3:46017416:G:A | A | G | 0.3628 | 0.0774 | 2.77E-06 | IFNy_C.conidiaHK_WB_48h | -0.437450665 | 0.019305828 |
| rs17330872 | 3 | 45993605 | chr3:45993605:A:G | A | G | -0.3702 | 0.0774 | 1.74E-06 | IL1b_C.conidiaHK_WB_48h | 0.286803946 | 0.019821308 |
| rs4797120 | 18 | 3725189 | chr18:3725189:G:C | C | G | -0.2956 | 0.0621 | 1.91E-06 | TNFA_PHA_WB_48h | 0.202030473 | 0.020428665 |
| rs73833520 | 3 | 46016375 | chr3:46016375:A:G | A | G | -0.3679 | 0.0768 | 1.69E-06 | IL1b_C.conidiaHK_WB_48h | 0.280397279 | 0.020478572 |
| rs35751180 | 3 | 46014681 | chr3:46014681:C:T | T | C | 0.3731 | 0.0774 | 1.42E-06 | TNFA_C.conidiaHK_WB_48h | -0.304730751 | 0.021070308 |
| rs4362758 | 3 | 46015419 | chr3:46015419:T:A | A | T | 0.3681 | 0.0768 | 1.67E-06 | TNFA_C.conidiaHK_WB_48h | -0.304448422 | 0.021223144 |
| rs2080811 | 2 | 15584186 | chr2:15584186:G:A | A | G | -0.329 | 0.0676 | 1.12E-06 | IFNy_C.conidiaHK_WB_48h | 0.342988387 | 0.021315284 |
| rs34168660 | 3 | 46014670 | chr3:46014670:G:A | A | G | 0.375 | 0.0768 | 1.06E-06 | TNFA_C.conidiaHK_WB_48h | -0.30400467 | 0.02144309 |
| rs12619108 | 2 | 15586369 | chr2:15586369:G:A | A | G | -0.315 | 0.0658 | 1.68E-06 | IFNy_C.conidiaHK_WB_48h | 0.342151873 | 0.021453177 |

| | | | | | | | | | | | |
|---|---|---|---|---|---|---|---|---|---|---|---|
| rs13081213 | 3 | 46014417 | chr3:46014417:C:T | T | C | 0.3746 | 0.0768 | 1.08E-06 | TNFA_C.conidiaHK_WB_48h | -0.303985622 | 0.021467133 |
| rs35669129 | 3 | 46013758 | chr3:46013758:G:A | A | G | 0.3746 | 0.0768 | 1.08E-06 | TNFA_C.conidiaHK_WB_48h | -0.304012332 | 0.021481851 |
| rs13081151 | 3 | 46014224 | chr3:46014224:G:A | A | G | 0.3751 | 0.0777 | 1.38E-06 | TNFA_C.conidiaHK_WB_48h | -0.303922293 | 0.021496994 |
| rs73833520 | 3 | 46016375 | chr3:46016375:A:G | A | G | -0.3679 | 0.0768 | 1.69E-06 | IFNy_C.conidiaHK_WB_48h | 0.431157301 | 0.022155515 |
| rs2230322 | 3 | 46021837 | chr3:46021837:T:C | T | C | -0.368 | 0.0768 | 1.68E-06 | IFNy_C.conidiaHK_WB_48h | 0.421744662 | 0.022767282 |
| rs13063033 | 3 | 46040989 | chr3:46040989:G:A | A | G | 0.3555 | 0.0772 | 4.12E-06 | IFNy_C.conidiaHK_WB_48h | -0.40457718 | 0.022768665 |
| rs71327006 | 3 | 46017416 | chr3:46017416:G:A | A | G | 0.3628 | 0.0774 | 2.77E-06 | IL1b_C.conidiaHK_WB_48h | -0.273267457 | 0.022850761 |
| rs71327007 | 3 | 46017757 | chr3:46017757:C:T | T | C | 0.3643 | 0.0774 | 2.51E-06 | IL1b_C.conidiaHK_WB_48h | -0.273029831 | 0.022953129 |
| rs35454877 | 3 | 46017992 | chr3:46017992:T:C | T | C | -0.3679 | 0.0768 | 1.69E-06 | IL1b_C.conidiaHK_WB_48h | 0.272864301 | 0.023028959 |
| rs35334665 | 3 | 46019726 | chr3:46019726:T:C | T | C | -0.3632 | 0.0774 | 2.70E-06 | IL1b_C.conidiaHK_WB_48h | 0.271265095 | 0.023737813 |
| rs36040135 | 3 | 46019812 | chr3:46019812:A:G | A | G | -0.3679 | 0.0768 | 1.69E-06 | IL1b_C.conidiaHK_WB_48h | 0.271182792 | 0.023768124 |
| rs13074382 | 3 | 46020348 | chr3:46020348:A:G | A | G | -0.3679 | 0.0768 | 1.69E-06 | IL1b_C.conidiaHK_WB_48h | 0.270735797 | 0.023955978 |
| rs13097556 | 3 | 46020505 | chr3:46020505:T:C | T | C | -0.3679 | 0.0768 | 1.69E-06 | IL1b_C.conidiaHK_WB_48h | 0.270547744 | 0.024040586 |
| rs34836513 | 3 | 46004967 | chr3:46004967:G:A | A | G | 0.3674 | 0.0772 | 1.96E-06 | TNFA_C.conidiaHK_WB_48h | -0.298417374 | 0.024123833 |
| rs876668 | 3 | 46022261 | chr3:46022261:T:C | T | C | -0.368 | 0.0768 | 1.68E-06 | IL1b_C.conidiaHK_WB_48h | 0.269201265 | 0.024647522 |
| rs35930050 | 3 | 46024271 | chr3:46024271:G:A | A | G | 0.3681 | 0.0768 | 1.67E-06 | IL1b_C.conidiaHK_WB_48h | -0.269231527 | 0.02468955 |
| rs34584867 | 3 | 46024471 | chr3:46024471:G:A | A | G | 0.3681 | 0.0768 | 1.67E-06 | IL1b_C.conidiaHK_WB_48h | -0.269231527 | 0.02468955 |
| rs35161099 | 3 | 46026015 | chr3:46026015:T:G | T | G | -0.3625 | 0.0774 | 2.83E-06 | IL1b_C.conidiaHK_WB_48h | 0.269250575 | 0.024734681 |
| rs35159820 | 3 | 46026981 | chr3:46026981:A:G | A | G | -0.3673 | 0.0769 | 1.76E-06 | IL1b_C.conidiaHK_WB_48h | 0.269253581 | 0.024746451 |
| rs71327010 | 3 | 46027343 | chr3:46027343:G:T | T | G | 0.3625 | 0.0774 | 2.83E-06 | IL1b_C.conidiaHK_WB_48h | -0.269255884 | 0.024758416 |
| rs13090194 | 3 | 46028081 | chr3:46028081:A:C | A | C | -0.3672 | 0.0769 | 1.77E-06 | IL1b_C.conidiaHK_WB_48h | 0.269258379 | 0.024782934 |
| rs59166269 | 3 | 46028845 | chr3:46028845:C:G | C | G | -0.3672 | 0.0769 | 1.77E-06 | IL1b_C.conidiaHK_WB_48h | 0.26925857 | 0.024795487 |
| rs57437758 | 3 | 46028861 | chr3:46028861:A:C | A | C | -0.3672 | 0.0769 | 1.77E-06 | IL1b_C.conidiaHK_WB_48h | 0.26925857 | 0.024795487 |
| rs34562820 | 3 | 46029589 | chr3:46029589:G:A | A | G | 0.3625 | 0.0775 | 2.87E-06 | IL1b_C.conidiaHK_WB_48h | -0.269258056 | 0.024808236 |
| rs13082697 | 3 | 46030271 | chr3:46030271:T:C | T | C | -0.3671 | 0.0769 | 1.78E-06 | IL1b_C.conidiaHK_WB_48h | 0.269256838 | 0.024821183 |
| rs13060287 | 3 | 46030376 | chr3:46030376:A:G | A | G | -0.3671 | 0.0769 | 1.79E-06 | IL1b_C.conidiaHK_WB_48h | 0.269254913 | 0.024834327 |
| rs34619093 | 3 | 46031027 | chr3:46031027:G:A | A | G | 0.3625 | 0.0775 | 2.89E-06 | IL1b_C.conidiaHK_WB_48h | -0.269142199 | 0.024904561 |
| rs68087193 | 3 | 46031138 | chr3:46031138:C:T | T | C | 0.3673 | 0.0769 | 1.76E-06 | IL1b_C.conidiaHK_WB_48h | -0.269142199 | 0.024904561 |
| rs2102055 | 3 | 46031212 | chr3:46031212:G:A | A | G | 0.3591 | 0.0767 | 2.83E-06 | IL1b_C.conidiaHK_WB_48h | -0.269142199 | 0.024904561 |
| rs2102056 | 3 | 46031276 | chr3:46031276:G:A | A | G | 0.3671 | 0.0769 | 1.78E-06 | IL1b_C.conidiaHK_WB_48h | -0.269142199 | 0.024904561 |
| rs2088690 | 3 | 46031347 | chr3:46031347:C:T | T | C | 0.3671 | 0.0769 | 1.78E-06 | IL1b_C.conidiaHK_WB_48h | -0.269142199 | 0.024904561 |
| rs34774687 | 3 | 46031822 | chr3:46031822:T:C | T | C | -0.367 | 0.0769 | 1.80E-06 | IL1b_C.conidiaHK_WB_48h | 0.26913885 | 0.02491813 |
| rs35814488 | 3 | 46032431 | chr3:46032431:A:G | A | G | -0.3672 | 0.0769 | 1.77E-06 | IL1b_C.conidiaHK_WB_48h | 0.269134795 | 0.024931898 |
| rs6790866 | 3 | 46033219 | chr3:46033219:G:A | A | G | 0.367 | 0.0769 | 1.80E-06 | IL1b_C.conidiaHK_WB_48h | -0.269130034 | 0.024945864 |
| rs6791016 | 3 | 46033366 | chr3:46033366:G:A | A | G | 0.367 | 0.0769 | 1.80E-06 | IL1b_C.conidiaHK_WB_48h | -0.269124566 | 0.024960029 |
| rs6780028 | 3 | 46033598 | chr3:46033598:A:C | A | C | -0.367 | 0.0769 | 1.80E-06 | IL1b_C.conidiaHK_WB_48h | 0.269124566 | 0.024960029 |
| rs2088692 | 3 | 46035840 | chr3:46035840:A:G | A | G | -0.3654 | 0.0768 | 1.96E-06 | IL1b_C.conidiaHK_WB_48h | 0.269155263 | 0.024977765 |
| rs34438204 | 3 | 46039814 | chr3:46039814:T:C | T | C | -0.3609 | 0.0774 | 3.13E-06 | IL1b_C.conidiaHK_WB_48h | 0.269082296 | 0.02508665 |
| rs34679077 | 3 | 46046500 | chr3:46046500:G:A | A | G | 0.3609 | 0.0774 | 3.13E-06 | IL1b_C.conidiaHK_WB_48h | -0.268786386 | 0.025335841 |
| rs34863575 | 3 | 46052571 | chr3:46052571:A:G | A | G | -0.3543 | 0.0773 | 4.53E-06 | IL1b_C.conidiaHK_WB_48h | 0.26829837 | 0.025703535 |
| rs35772789 | 3 | 46053612 | chr3:46053612:A:G | A | G | -0.3544 | 0.0773 | 4.52E-06 | IL1b_C.conidiaHK_WB_48h | 0.268158697 | 0.025794558 |
| rs34340587 | 3 | 46054551 | chr3:46054551:G:A | A | G | 0.3542 | 0.0773 | 4.63E-06 | IL1b_C.conidiaHK_WB_48h | -0.268096953 | 0.025838607 |
| rs13063033 | 3 | 46040989 | chr3:46040989:G:A | A | G | 0.3555 | 0.0772 | 4.12E-06 | IL1b_C.conidiaHK_WB_48h | -0.254080897 | 0.025930265 |
| rs77399277 | 3 | 46055306 | chr3:46055306:T:C | T | C | -0.3556 | 0.0773 | 4.25E-06 | IL1b_C.conidiaHK_WB_48h | 0.267906873 | 0.025946056 |
| rs13063033 | 3 | 46040989 | chr3:46040989:G:A | A | G | 0.3555 | 0.0772 | 4.12E-06 | TNFA_C.conidiaHK_WB_48h | -0.273755771 | 0.026041052 |
| rs34718164 | 3 | 46057034 | chr3:46057034:C:G | C | G | -0.3556 | 0.0773 | 4.25E-06 | IL1b_C.conidiaHK_WB_48h | 0.267610407 | 0.02612145 |
| rs34754340 | 3 | 46000345 | chr3:46000345:C:T | T | C | 0.3673 | 0.0772 | 1.94E-06 | TNFA_C.conidiaHK_WB_48h | -0.29486867 | 0.026139055 |
| rs13096741 | 3 | 46057662 | chr3:46057662:C:T | T | C | 0.3556 | 0.0773 | 4.27E-06 | IL1b_C.conidiaHK_WB_48h | -0.267519353 | 0.026173538 |
| rs35420565 | 3 | 46058646 | chr3:46058646:C:G | C | G | -0.3556 | 0.0773 | 4.26E-06 | IL1b_C.conidiaHK_WB_48h | 0.267328794 | 0.026280369 |
| rs34047915 | 3 | 46058999 | chr3:46058999:C:T | T | C | 0.3555 | 0.0773 | 4.29E-06 | IL1b_C.conidiaHK_WB_48h | -0.267279396 | 0.026307633 |
| rs71327014 | 3 | 46059974 | chr3:46059974:C:T | T | C | 0.3556 | 0.0773 | 4.25E-06 | IL1b_C.conidiaHK_WB_48h | -0.267229296 | 0.02633512 |
| rs71327015 | 3 | 46060681 | chr3:46060681:G:C | C | G | 0.3585 | 0.0773 | 3.57E-06 | IL1b_C.conidiaHK_WB_48h | -0.267229296 | 0.02633512 |
| rs34127208 | 3 | 46062188 | chr3:46062188:G:T | T | G | 0.3554 | 0.0773 | 4.31E-06 | IL1b_C.conidiaHK_WB_48h | -0.267166557 | 0.026368869 |
| rs35516580 | 3 | 46066006 | chr3:46066006:A:G | A | G | -0.3562 | 0.0773 | 4.13E-06 | IL1b_C.conidiaHK_WB_48h | 0.267116973 | 0.026395564 |
| rs34766614 | 3 | 46066109 | chr3:46066109:A:G | A | G | -0.3484 | 0.0775 | 6.99E-06 | IL1b_C.conidiaHK_WB_48h | 0.267116973 | 0.026395564 |
| rs1491950 | 3 | 46096615 | chr3:46096615:G:A | A | G | 0.3544 | 0.078 | 5.55E-06 | IL1b_C.conidiaHK_WB_48h | -0.265039012 | 0.026437873 |
| rs71615438 | 3 | 45998376 | chr3:45998376:A:G | A | G | -0.3666 | 0.0772 | 2.04E-06 | TNFA_C.conidiaHK_WB_48h | 0.293370043 | 0.027006979 |
| rs36023124 | 3 | 45997568 | chr3:45997568:G:C | C | G | 0.3665 | 0.0771 | 2.01E-06 | TNFA_C.conidiaHK_WB_48h | -0.292586907 | 0.027456268 |
| rs12619874 | 2 | 15586997 | chr2:15586997:G:C | C | G | -0.3204 | 0.0665 | 1.46E-06 | IFNy_C.conidiaHK_WB_48h | 0.322625539 | 0.027729735 |
| rs34452002 | 3 | 46101695 | chr3:46101695:C:T | T | C | 0.3556 | 0.0779 | 4.95E-06 | IL1b_C.conidiaHK_WB_48h | -0.26254225 | 0.02831918 |
| rs34093271 | 3 | 46103489 | chr3:46103489:G:T | T | G | 0.3558 | 0.0779 | 4.89E-06 | IL1b_C.conidiaHK_WB_48h | -0.262519291 | 0.028410436 |
| rs73833520 | 3 | 46016375 | chr3:46016375:A:G | A | G | -0.3679 | 0.0768 | 1.69E-06 | TNFA_C.conidiaHK_WB_48h | 0.285552825 | 0.028666097 |
| rs2302934 | 2 | 15613155 | chr2:15613155:C:G | C | G | 0.3393 | 0.076 | 8.11E-06 | IFNy_C.conidiaHK_WB_48h | -0.345379352 | 0.028859641 |
| rs12616567 | 2 | 15615498 | chr2:15615498:A:C | A | C | 0.3427 | 0.0762 | 6.85E-06 | IFNy_C.conidiaHK_WB_48h | -0.345379352 | 0.028859641 |
| rs807635 | 2 | 15629483 | chr2:15629483:C:T | T | C | -0.3579 | 0.0765 | 2.85E-06 | IFNy_C.conidiaHK_WB_48h | 0.343211712 | 0.02931218 |
| rs807636 | 2 | 15627828 | chr2:15627828:C:T | T | C | -0.365 | 0.0772 | 2.25E-06 | IFNy_C.conidiaHK_WB_48h | 0.343577533 | 0.029339827 |
| rs17330872 | 3 | 45993605 | chr3:45993605:A:G | A | G | -0.3702 | 0.0774 | 1.74E-06 | TNFA_C.conidiaHK_WB_48h | 0.289060162 | 0.029499172 |
| rs34460587 | 3 | 46100972 | chr3:46100972:C:T | T | C | 0.3557 | 0.0779 | 4.93E-06 | IL1b_C.conidiaHK_WB_48h | -0.260487176 | 0.029515945 |
| rs876668 | 3 | 46022261 | chr3:46022261:T:C | T | C | -0.368 | 0.0768 | 1.68E-06 | TNFA_C.conidiaHK_WB_48h | 0.280918865 | 0.02969526 |
| rs35930050 | 3 | 46024271 | chr3:46024271:G:A | A | G | 0.3681 | 0.0768 | 1.67E-06 | TNFA_C.conidiaHK_WB_48h | -0.280679257 | 0.029902021 |
| rs34584867 | 3 | 46024471 | chr3:46024471:G:A | A | G | 0.3681 | 0.0768 | 1.67E-06 | TNFA_C.conidiaHK_WB_48h | -0.280679257 | 0.029902021 |
| rs13097556 | 3 | 46020505 | chr3:46020505:T:C | T | C | -0.3679 | 0.0768 | 1.69E-06 | TNFA_C.conidiaHK_WB_48h | 0.280693791 | 0.029951351 |
| rs13074382 | 3 | 46020348 | chr3:46020348:A:G | A | G | -0.3679 | 0.0768 | 1.69E-06 | TNFA_C.conidiaHK_WB_48h | 0.280677444 | 0.029977245 |
| rs35161099 | 3 | 46026015 | chr3:46026015:T:G | T | G | -0.3625 | 0.0774 | 2.83E-06 | TNFA_C.conidiaHK_WB_48h | 0.28042753 | 0.030113621 |
| rs35751180 | 3 | 46014681 | chr3:46014681:C:T | T | C | 0.3731 | 0.0774 | 1.42E-06 | IFNy_C.conidiaHK_WB_48h | -0.413919633 | 0.030142942 |
| rs35159820 | 3 | 46026981 | chr3:46026981:A:G | A | G | -0.3673 | 0.0769 | 1.76E-06 | TNFA_C.conidiaHK_WB_48h | 0.280362706 | 0.030167285 |
| rs4362758 | 3 | 46015419 | chr3:46015419:T:A | A | T | 0.3681 | 0.0768 | 1.67E-06 | IFNy_C.conidiaHK_WB_48h | -0.413952425 | 0.03017128 |
| rs71327010 | 3 | 46027343 | chr3:46027343:G:T | T | G | 0.3625 | 0.0774 | 2.83E-06 | TNFA_C.conidiaHK_WB_48h | -0.280297124 | 0.030221257 |
| rs36040135 | 3 | 46019812 | chr3:46019812:A:G | A | G | -0.3679 | 0.0768 | 1.69E-06 | TNFA_C.conidiaHK_WB_48h | 0.280344605 | 0.030226953 |
| rs35334665 | 3 | 46019726 | chr3:46019726:T:C | T | C | -0.3632 | 0.0774 | 2.70E-06 | TNFA_C.conidiaHK_WB_48h | 0.280329563 | 0.030250545 |
| rs34492478 | 3 | 46097350 | chr3:46097350:A:T | A | T | -0.3562 | 0.078 | 4.95E-06 | IL1b_C.conidiaHK_WB_48h | 0.259779634 | 0.030251773 |
| rs13090194 | 3 | 46028081 | chr3:46028081:A:C | A | C | -0.3672 | 0.0769 | 1.77E-06 | TNFA_C.conidiaHK_WB_48h | 0.280163689 | 0.030330127 |
| rs59166269 | 3 | 46028845 | chr3:46028845:C:G | C | G | -0.3672 | 0.0769 | 1.77E-06 | TNFA_C.conidiaHK_WB_48h | 0.280095836 | 0.030385028 |
| rs57437758 | 3 | 46028861 | chr3:46028861:A:C | A | C | -0.3672 | 0.0769 | 1.77E-06 | TNFA_C.conidiaHK_WB_48h | 0.280095836 | 0.030385028 |
| rs34562820 | 3 | 46029589 | chr3:46029589:G:A | A | G | 0.3625 | 0.0775 | 2.87E-06 | TNFA_C.conidiaHK_WB_48h | -0.280027227 | 0.030440241 |
| rs13082697 | 3 | 46030271 | chr3:46030271:T:C | T | C | -0.3671 | 0.0769 | 1.78E-06 | TNFA_C.conidiaHK_WB_48h | 0.279957861 | 0.030495767 |
| rs13060287 | 3 | 46030376 | chr3:46030376:A:G | A | G | -0.3671 | 0.0769 | 1.79E-06 | TNFA_C.conidiaHK_WB_48h | 0.279887739 | 0.030551607 |
| rs13381043 | 18 | 3724602 | chr18:3724602:G:A | A | G | -0.325 | 0.0613 | 1.13E-07 | IL1b_LPS_WB_48h | 0.102290939 | 0.030581167 |
| rs34168660 | 3 | 46014670 | chr3:46014670:G:A | A | G | 0.375 | 0.0768 | 1.06E-06 | IFNy_C.conidiaHK_WB_48h | -0.41297388 | 0.030603419 |
| rs34619093 | 3 | 46031027 | chr3:46031027:G:A | A | G | 0.3625 | 0.0775 | 2.89E-06 | TNFA_C.conidiaHK_WB_48h | -0.279814069 | 0.030606449 |
| rs68087193 | 3 | 46031138 | chr3:46031138:C:T | T | C | 0.3673 | 0.0769 | 1.76E-06 | TNFA_C.conidiaHK_WB_48h | -0.279814069 | 0.030606449 |
| rs2102055 | 3 | 46031212 | chr3:46031212:G:A | A | G | 0.3591 | 0.0767 | 2.83E-06 | TNFA_C.conidiaHK_WB_48h | -0.279814069 | 0.030606449 |
| rs2102056 | 3 | 46031276 | chr3:46031276:G:A | A | G | 0.3671 | 0.0769 | 1.78E-06 | TNFA_C.conidiaHK_WB_48h | -0.279814069 | 0.030606449 |
| rs2088690 | 3 | 46031347 | chr3:46031347:C:T | T | C | 0.3671 | 0.0769 | 1.78E-06 | TNFA_C.conidiaHK_WB_48h | -0.279814069 | 0.030606449 |
| rs34774687 | 3 | 46031822 | chr3:46031822:T:C | T | C | -0.367 | 0.0769 | 1.80E-06 | TNFA_C.conidiaHK_WB_48h | 0.279742442 | 0.030662915 |
| rs13081213 | 3 | 46014417 | chr3:46014417:C:T | T | C | 0.3746 | 0.0768 | 1.08E-06 | IFNy_C.conidiaHK_WB_48h | -0.412779924 | 0.03070245 |
| rs35814488 | 3 | 46032431 | chr3:46032431:A:G | A | G | -0.3672 | 0.0769 | 1.77E-06 | TNFA_C.conidiaHK_WB_48h | 0.279670059 | 0.030719699 |
| rs13081151 | 3 | 46014224 | chr3:46014224:G:A | A | G | 0.3751 | 0.0777 | 1.38E-06 | IFNy_C.conidiaHK_WB_48h | -0.412686823 | 0.030743549 |
| rs71327017 | 3 | 46069721 | chr3:46069721:T:C | T | C | -0.3576 | 0.0774 | 3.80E-06 | IL1b_C.conidiaHK_WB_48h | 0.258377497 | 0.030759255 |
| rs13072267 | 3 | 46074518 | chr3:46074518:T:A | A | T | 0.3543 | 0.0773 | 4.57E-06 | IL1b_C.conidiaHK_WB_48h | -0.258377497 | 0.030759255 |
| rs36057789 | 3 | 46075604 | chr3:46075604:A:G | A | G | -0.3551 | 0.0773 | 4.36E-06 | IL1b_C.conidiaHK_WB_48h | 0.258377497 | 0.030759255 |
| rs76647202 | 3 | 46076468 | chr3:46076468:A:G | A | G | -0.3542 | 0.0773 | 4.60E-06 | IL1b_C.conidiaHK_WB_48h | 0.258377497 | 0.030759255 |
| rs71327021 | 3 | 46079025 | chr3:46079025:A:G | A | G | -0.3542 | 0.0773 | 4.61E-06 | IL1b_C.conidiaHK_WB_48h | 0.258377497 | 0.030759255 |
| rs6790866 | 3 | 46033219 | chr3:46033219:G:A | A | G | 0.367 | 0.0769 | 1.80E-06 | TNFA_C.conidiaHK_WB_48h | -0.279596921 | 0.030776801 |
| rs13091868 | 3 | 46081551 | chr3:46081551:A:G | A | G | -0.3538 | 0.0773 | 4.72E-06 | IL1b_C.conidiaHK_WB_48h | 0.258326391 | 0.030790228 |
| rs13092030 | 3 | 46081563 | chr3:46081563:C:T | T | C | 0.354 | 0.0773 | 4.66E-06 | IL1b_C.conidiaHK_WB_48h | -0.258326391 | 0.030790228 |
| rs17215981 | 3 | 46082926 | chr3:46082926:A:G | A | G | -0.3547 | 0.0773 | 4.47E-06 | IL1b_C.conidiaHK_WB_48h | 0.258326391 | 0.030790228 |
| rs13089554 | 3 | 46086065 | chr3:46086065:G:A | A | G | 0.3547 | 0.0774 | 4.61E-06 | IL1b_C.conidiaHK_WB_48h | -0.258326391 | 0.030790228 |
| rs13095717 | 3 | 46087050 | chr3:46087050:G:A | A | G | 0.3548 | 0.0774 | 4.58E-06 | IL1b_C.conidiaHK_WB_48h | -0.258326391 | 0.030790228 |
| rs13075528 | 3 | 46087217 | chr3:46087217:T:C | T | C | -0.3549 | 0.0775 | 4.63E-06 | IL1b_C.conidiaHK_WB_48h | 0.258326391 | 0.030790228 |
| rs13095602 | 3 | 46087221 | chr3:46087221:A:G | A | G | -0.3547 | 0.0774 | 4.60E-06 | IL1b_C.conidiaHK_WB_48h | 0.258326391 | 0.030790228 |
| rs71327023 | 3 | 46089733 | chr3:46089733:C:G | C | G | -0.3539 | 0.0777 | 5.19E-06 | IL1b_C.conidiaHK_WB_48h | 0.258326391 | 0.030790228 |
| rs13063527 | 3 | 46094198 | chr3:46094198:G:A | A | G | 0.3561 | 0.0777 | 4.51E-06 | IL1b_C.conidiaHK_WB_48h | -0.258326391 | 0.030790228 |
| rs13068570 | 3 | 46094946 | chr3:46094946:A:G | A | G | -0.3561 | 0.0777 | 4.61E-06 | IL1b_C.conidiaHK_WB_48h | 0.258326391 | 0.030790228 |
| rs4373103 | 3 | 46096963 | chr3:46096963:G:A | A | G | 0.3547 | 0.078 | 5.42E-06 | IL1b_C.conidiaHK_WB_48h | -0.258326391 | 0.030790228 |
| rs71327024 | 3 | 46098581 | chr3:46098581:G:T | T | G | 0.3547 | 0.078 | 5.37E-06 | IL1b_C.conidiaHK_WB_48h | -0.258326391 | 0.030790228 |
| rs6791016 | 3 | 46033366 | chr3:46033366:G:A | A | G | 0.367 | 0.0769 | 1.80E-06 | TNFA_C.conidiaHK_WB_48h | -0.279523027 | 0.030834222 |
| rs6780028 | 3 | 46033598 | chr3:46033598:A:C | A | C | -0.367 | 0.0769 | 1.80E-06 | TNFA_C.conidiaHK_WB_48h | 0.279523027 | 0.030834222 |
| rs35669129 | 3 | 46013758 | chr3:46013758:G:A | A | G | 0.3746 | 0.0768 | 1.08E-06 | IFNy_C.conidiaHK_WB_48h | -0.412532209 | 0.030837033 |
| rs613534 | 9 | 133267707 | chr9:133267707:G:A | A | G | -0.2801 | 0.0517 | 6.16E-08 | IL6_PHA_WB_48h | 0.101453668 | 0.030840862 |
| rs35454877 | 3 | 46017992 | chr3:46017992:T:C | T | C | -0.3679 | 0.0768 | 1.69E-06 | TNFA_C.conidiaHK_WB_48h | 0.279492105 | 0.030883826 |
| rs71327007 | 3 | 46017757 | chr3:46017757:C:T | T | C | 0.3643 | 0.0774 | 2.51E-06 | TNFA_C.conidiaHK_WB_48h | -0.279451456 | 0.030917765 |
| rs71327006 | 3 | 46017416 | chr3:46017416:G:A | A | G | 0.3628 | 0.0774 | 2.77E-06 | TNFA_C.conidiaHK_WB_48h | -0.279384557 | 0.030979215 |
| rs2088692 | 3 | 46035840 | chr3:46035840:A:G | A | G | -0.3654 | 0.0768 | 1.96E-06 | TNFA_C.conidiaHK_WB_48h | 0.279262203 | 0.03103155 |
| rs34438204 | 3 | 46039814 | chr3:46039814:T:C | T | C | -0.3609 | 0.0774 | 3.13E-06 | TNFA_C.conidiaHK_WB_48h | 0.278707886 | 0.031449675 |
| rs71327025 | 3 | 46106012 | chr3:46106012:A:G | A | G | -0.3556 | 0.0779 | 4.96E-06 | IL1b_C.conidiaHK_WB_48h | 0.25838508 | 0.031478794 |
| rs13075758 | 3 | 45983556 | chr3:45983556:G:A | A | G | 0.3672 | 0.0775 | 2.19E-06 | IL1b_C.conidiaHK_WB_48h | -0.264361637 | 0.031618541 |
| rs34442130 | 3 | 45983037 | chr3:45983037:T:A | A | T | 0.3674 | 0.0775 | 2.16E-06 | IL1b_C.conidiaHK_WB_48h | -0.264350242 | 0.031638163 |
| rs13093179 | 3 | 46104822 | chr3:46104822:G:T | T | G | 0.3557 | 0.0779 | 4.91E-06 | IL1b_C.conidiaHK_WB_48h | -0.260449791 | 0.031802895 |
| rs34679077 | 3 | 46046500 | chr3:46046500:G:A | A | G | 0.3609 | 0.0774 | 3.13E-06 | TNFA_C.conidiaHK_WB_48h | -0.277759097 | 0.032138506 |
| rs36122610 | 3 | 45981341 | chr3:45981341:G:A | A | G | 0.3673 | 0.0775 | 2.18E-06 | IL1b_C.conidiaHK_WB_48h | -0.263507462 | 0.032199852 |
| rs35539222 | 3 | 46112780 | chr3:46112780:T:C | T | C | -0.3556 | 0.0779 | 4.96E-06 | IL1b_C.conidiaHK_WB_48h | 0.257209144 | 0.032351833 |
| rs35110864 | 3 | 46112965 | chr3:46112965:G:A | A | G | 0.3555 | 0.0779 | 4.99E-06 | IL1b_C.conidiaHK_WB_48h | -0.257209144 | 0.032351833 |
| rs657152 | 9 | 133263862 | chr9:133263862:A:C | A | C | 0.2814 | 0.0516 | 4.95E-08 | IL6_PHA_WB_48h | -0.092794515 | 0.03308046 |
| rs2230322 | 3 | 46021837 | chr3:46021837:T:C | T | C | -0.368 | 0.0768 | 1.68E-06 | IL1b_C.conidiaHK_WB_48h | 0.253122918 | 0.033337883 |
| rs34863575 | 3 | 46052571 | chr3:46052571:A:G | A | G | -0.3543 | 0.0773 | 4.53E-06 | TNFA_C.conidiaHK_WB_48h | 0.276017436 | 0.033358138 |
| rs34442130 | 3 | 45983037 | chr3:45983037:T:A | A | T | 0.3674 | 0.0775 | 2.16E-06 | IFNy_C.conidiaHK_WB_48h | -0.407440032 | 0.033470162 |
| rs13075758 | 3 | 45983556 | chr3:45983556:G:A | A | G | 0.3672 | 0.0775 | 2.19E-06 | IFNy_C.conidiaHK_WB_48h | -0.40703136 | 0.033636251 |
| rs35772789 | 3 | 46053612 | chr3:46053612:A:G | A | G | -0.3544 | 0.0773 | 4.52E-06 | TNFA_C.conidiaHK_WB_48h | 0.275597859 | 0.03364501 |
| rs36122610 | 3 | 45981341 | chr3:45981341:G:A | A | G | 0.3673 | 0.0775 | 2.18E-06 | IFNy_C.conidiaHK_WB_48h | -0.40685908 | 0.033736384 |
| rs11627388 | 14 | 31953140 | chr14:31953140:A:C | A | C | -0.2482 | 0.0533 | 3.19E-06 | IFNy_LPS_WB_48h | 0.252834388 | 0.033854498 |
| rs34340587 | 3 | 46054551 | chr3:46054551:G:A | A | G | 0.3542 | 0.0773 | 4.63E-06 | TNFA_C.conidiaHK_WB_48h | -0.275239533 | 0.033889431 |
| rs77399277 | 3 | 46055306 | chr3:46055306:T:C | T | C | -0.3556 | 0.0773 | 4.25E-06 | TNFA_C.conidiaHK_WB_48h | 0.275017115 | 0.034036318 |
| rs17330872 | 3 | 45993605 | chr3:45993605:A:G | A | G | -0.3702 | 0.0774 | 1.74E-06 | IFNy_C.conidiaHK_WB_48h | 0.406267617 | 0.034212109 |
| rs647800 | 9 | 133272585 | chr9:133272585:G:A | A | G | -0.2683 | 0.052 | 2.41E-07 | IL6_S.aureus_WB_48h | 0.15209724 | 0.034216106 |
| rs34718164 | 3 | 46057034 | chr3:46057034:C:G | C | G | -0.3556 | 0.0773 | 4.25E-06 | TNFA_C.conidiaHK_WB_48h | 0.274229117 | 0.034566223 |
| rs36023124 | 3 | 45997568 | chr3:45997568:G:C | C | G | 0.3665 | 0.0771 | 2.01E-06 | IFNy_C.conidiaHK_WB_48h | -0.405043061 | 0.034636521 |
| rs13096741 | 3 | 46057662 | chr3:46057662:C:T | T | C | 0.3556 | 0.0773 | 4.27E-06 | TNFA_C.conidiaHK_WB_48h | -0.273997449 | 0.034721079 |
| rs71615438 | 3 | 45998376 | chr3:45998376:A:G | A | G | -0.3666 | 0.0772 | 2.04E-06 | IFNy_C.conidiaHK_WB_48h | 0.404736032 | 0.034732858 |
| rs35420565 | 3 | 46058646 | chr3:46058646:C:G | C | G | -0.3556 | 0.0773 | 4.26E-06 | TNFA_C.conidiaHK_WB_48h | 0.273525458 | 0.035035462 |
| rs34754340 | 3 | 46000345 | chr3:46000345:C:T | T | C | 0.3673 | 0.0772 | 1.94E-06 | IFNy_C.conidiaHK_WB_48h | -0.403785013 | 0.035052131 |
| rs34047915 | 3 | 46058999 | chr3:46058999:C:T | T | C | 0.3555 | 0.0773 | 4.29E-06 | TNFA_C.conidiaHK_WB_48h | -0.273405663 | 0.035115039 |
| rs35516580 | 3 | 46066006 | chr3:46066006:A:G | A | G | -0.3562 | 0.0773 | 4.13E-06 | TNFA_C.conidiaHK_WB_48h | 0.273280776 | 0.035194056 |
| rs34766614 | 3 | 46066109 | chr3:46066109:A:G | A | G | -0.3484 | 0.0775 | 6.99E-06 | TNFA_C.conidiaHK_WB_48h | 0.273280776 | 0.035194056 |
| rs71327014 | 3 | 46059974 | chr3:46059974:C:T | T | C | 0.3556 | 0.0773 | 4.25E-06 | TNFA_C.conidiaHK_WB_48h | -0.27328515 | 0.03519501 |
| rs71327015 | 3 | 46060681 | chr3:46060681:G:C | C | G | 0.3585 | 0.0773 | 3.57E-06 | TNFA_C.conidiaHK_WB_48h | -0.27328515 | 0.03519501 |
| rs34127208 | 3 | 46062188 | chr3:46062188:G:T | T | G | 0.3554 | 0.0773 | 4.31E-06 | TNFA_C.conidiaHK_WB_48h | -0.27324392 | 0.03522059 |
| rs3770462 | 2 | 15609200 | chr2:15609200:T:G | T | G | 0.3409 | 0.0757 | 6.74E-06 | IFNy_C.conidiaHK_WB_48h | -0.32711716 | 0.035359406 |
| rs4668941 | 2 | 15610007 | chr2:15610007:T:G | T | G | 0.3359 | 0.0754 | 8.51E-06 | IFNy_C.conidiaHK_WB_48h | -0.32711716 | 0.035359406 |
| rs657152 | 9 | 133263862 | chr9:133263862:A:C | A | C | 0.2814 | 0.0516 | 4.95E-08 | IL6_S.aureus_WB_48h | -0.155807154 | 0.035360926 |
| rs41264195 | 2 | 15606096 | chr2:15606096:C:T | T | C | -0.3359 | 0.0754 | 8.49E-06 | IFNy_C.conidiaHK_WB_48h | 0.327091227 | 0.035390609 |
| rs79808866 | 2 | 15604860 | chr2:15604860:T:G | T | G | 0.3357 | 0.0754 | 8.59E-06 | IFNy_C.conidiaHK_WB_48h | -0.327031283 | 0.035422838 |
| rs34836513 | 3 | 46004967 | chr3:46004967:G:A | A | G | 0.3674 | 0.0772 | 1.96E-06 | IFNy_C.conidiaHK_WB_48h | -0.401770265 | 0.03564304 |
| rs543968 | 9 | 133267708 | chr9:133267708:C:T | T | C | -0.2815 | 0.0517 | 5.32E-08 | IL6_PHA_WB_48h | 0.092662655 | 0.036220984 |
| rs11627388 | 14 | 31953140 | chr14:31953140:A:C | A | C | -0.2482 | 0.0533 | 3.19E-06 | IL6_LPS_WB_48h | 0.0897389 | 0.037895095 |
| rs1491950 | 3 | 46096615 | chr3:46096615:G:A | A | G | 0.3544 | 0.078 | 5.55E-06 | TNFA_C.conidiaHK_WB_48h | -0.267038204 | 0.038136017 |
| rs13093179 | 3 | 46104822 | chr3:46104822:G:T | T | G | 0.3557 | 0.0779 | 4.91E-06 | TNFA_C.conidiaHK_WB_48h | -0.271260729 | 0.038138593 |
| rs1491951 | 3 | 46100352 | chr3:46100352:G:A | A | G | 0.3485 | 0.0777 | 7.30E-06 | IL1b_C.conidiaHK_WB_48h | -0.245002231 | 0.038403757 |
| rs613534 | 9 | 133267707 | chr9:133267707:G:A | A | G | -0.2801 | 0.0517 | 6.16E-08 | IFNy_S.aureus_WB_48h | 0.268745289 | 0.038497709 |
| rs807636 | 2 | 15627828 | chr2:15627828:C:T | T | C | -0.365 | 0.0772 | 2.25E-06 | IFNy_LPS_WB_48h | 0.29726349 | 0.038518085 |
| rs34093271 | 3 | 46103489 | chr3:46103489:G:T | T | G | 0.3558 | 0.0779 | 4.89E-06 | TNFA_C.conidiaHK_WB_48h | -0.267107973 | 0.038695178 |
| rs35539222 | 3 | 46112780 | chr3:46112780:T:C | T | C | -0.3556 | 0.0779 | 4.96E-06 | TNFA_C.conidiaHK_WB_48h | 0.267514171 | 0.039029521 |
| rs35110864 | 3 | 46112965 | chr3:46112965:G:A | A | G | 0.3555 | 0.0779 | 4.99E-06 | TNFA_C.conidiaHK_WB_48h | -0.267514171 | 0.039029521 |
| rs71327025 | 3 | 46106012 | chr3:46106012:A:G | A | G | -0.3556 | 0.0779 | 4.96E-06 | TNFA_C.conidiaHK_WB_48h | 0.267324753 | 0.039069669 |
| rs34452002 | 3 | 46101695 | chr3:46101695:C:T | T | C | 0.3556 | 0.0779 | 4.95E-06 | TNFA_C.conidiaHK_WB_48h | -0.266350437 | 0.039157615 |
| rs494242 | 9 | 133269706 | chr9:133269706:T:C | T | C | 0.2814 | 0.0517 | 5.39E-08 | IL6_PHA_WB_48h | -0.089920234 | 0.03917595 |
| rs644234 | 9 | 133266804 | chr9:133266804:G:T | T | G | -0.2815 | 0.0517 | 5.29E-08 | IL6_PHA_WB_48h | 0.089886624 | 0.039247098 |
| rs643434 | 9 | 133266942 | chr9:133266942:A:G | A | G | 0.2815 | 0.0517 | 5.33E-08 | IL6_PHA_WB_48h | -0.089886624 | 0.039247098 |
| rs544873 | 9 | 133267800 | chr9:133267800:A:G | A | G | 0.2815 | 0.0517 | 5.29E-08 | IL6_PHA_WB_48h | -0.089886624 | 0.039247098 |
| rs807635 | 2 | 15629483 | chr2:15629483:C:T | T | C | -0.3579 | 0.0765 | 2.85E-06 | IFNy_LPS_WB_48h | 0.295376472 | 0.039527972 |
| rs2302934 | 2 | 15613155 | chr2:15613155:C:G | C | G | 0.3393 | 0.076 | 8.11E-06 | IFNy_LPS_WB_48h | -0.295913442 | 0.039863062 |
| rs12616567 | 2 | 15615498 | chr2:15615498:A:C | A | C | 0.3427 | 0.0762 | 6.85E-06 | IFNy_LPS_WB_48h | -0.295913442 | 0.039863062 |
| rs34460587 | 3 | 46100972 | chr3:46100972:C:T | T | C | 0.3557 | 0.0779 | 4.93E-06 | TNFA_C.conidiaHK_WB_48h | -0.264710593 | 0.040299893 |
| rs76860840 | 2 | 15618674 | chr2:15618674:C:G | C | G | 0.3536 | 0.0764 | 3.66E-06 | IFNy_C.conidiaHK_WB_48h | -0.322354634 | 0.040609831 |
| rs657152 | 9 | 133263862 | chr9:133263862:A:C | A | C | 0.2814 | 0.0516 | 4.95E-08 | IL1b_S.aureus_WB_48h | -0.104114728 | 0.041461646 |
| rs13091868 | 3 | 46081551 | chr3:46081551:A:G | A | G | -0.3538 | 0.0773 | 4.72E-06 | TNFA_C.conidiaHK_WB_48h | 0.262962353 | 0.041512414 |
| rs13092030 | 3 | 46081563 | chr3:46081563:C:T | T | C | 0.354 | 0.0773 | 4.66E-06 | TNFA_C.conidiaHK_WB_48h | -0.262962353 | 0.041512414 |
| rs17215981 | 3 | 46082926 | chr3:46082926:A:G | A | G | -0.3547 | 0.0773 | 4.47E-06 | TNFA_C.conidiaHK_WB_48h | 0.262962353 | 0.041512414 |
| rs13089554 | 3 | 46086065 | chr3:46086065:G:A | A | G | 0.3547 | 0.0774 | 4.61E-06 | TNFA_C.conidiaHK_WB_48h | -0.262962353 | 0.041512414 |
| rs13095717 | 3 | 46087050 | chr3:46087050:G:A | A | G | 0.3548 | 0.0774 | 4.58E-06 | TNFA_C.conidiaHK_WB_48h | -0.262962353 | 0.041512414 |
| rs13075528 | 3 | 46087217 | chr3:46087217:T:C | T | C | -0.3549 | 0.0775 | 4.63E-06 | TNFA_C.conidiaHK_WB_48h | 0.262962353 | 0.041512414 |
| rs13095602 | 3 | 46087221 | chr3:46087221:A:G | A | G | -0.3547 | 0.0774 | 4.60E-06 | TNFA_C.conidiaHK_WB_48h | 0.262962353 | 0.041512414 |
| rs71327023 | 3 | 46089733 | chr3:46089733:C:G | C | G | -0.3539 | 0.0777 | 5.19E-06 | TNFA_C.conidiaHK_WB_48h | 0.262962353 | 0.041512414 |
| rs13063527 | 3 | 46094198 | chr3:46094198:G:A | A | G | 0.3561 | 0.0777 | 4.51E-06 | TNFA_C.conidiaHK_WB_48h | -0.262962353 | 0.041512414 |
| rs13068570 | 3 | 46094946 | chr3:46094946:A:G | A | G | -0.3561 | 0.0777 | 4.61E-06 | TNFA_C.conidiaHK_WB_48h | 0.262962353 | 0.041512414 |
| rs4373103 | 3 | 46096963 | chr3:46096963:G:A | A | G | 0.3547 | 0.078 | 5.42E-06 | TNFA_C.conidiaHK_WB_48h | -0.262962353 | 0.041512414 |
| rs71327024 | 3 | 46098581 | chr3:46098581:G:T | T | G | 0.3547 | 0.078 | 5.37E-06 | TNFA_C.conidiaHK_WB_48h | -0.262962353 | 0.041512414 |
| rs71327017 | 3 | 46069721 | chr3:46069721:T:C | T | C | -0.3576 | 0.0774 | 3.80E-06 | TNFA_C.conidiaHK_WB_48h | 0.262928056 | 0.041541923 |
| rs13072267 | 3 | 46074518 | chr3:46074518:T:A | A | T | 0.3543 | 0.0773 | 4.57E-06 | TNFA_C.conidiaHK_WB_48h | -0.262928056 | 0.041541923 |
| rs36057789 | 3 | 46075604 | chr3:46075604:A:G | A | G | -0.3551 | 0.0773 | 4.36E-06 | TNFA_C.conidiaHK_WB_48h | 0.262928056 | 0.041541923 |
| rs76647202 | 3 | 46076468 | chr3:46076468:A:G | A | G | -0.3542 | 0.0773 | 4.60E-06 | TNFA_C.conidiaHK_WB_48h | 0.262928056 | 0.041541923 |
| rs71327021 | 3 | 46079025 | chr3:46079025:A:G | A | G | -0.3542 | 0.0773 | 4.61E-06 | TNFA_C.conidiaHK_WB_48h | 0.262928056 | 0.041541923 |
| rs2230322 | 3 | 46021837 | chr3:46021837:T:C | T | C | -0.368 | 0.0768 | 1.68E-06 | TNFA_C.conidiaHK_WB_48h | 0.261359655 | 0.041616076 |
| rs13381043 | 18 | 3724602 | chr18:3724602:G:A | A | G | -0.325 | 0.0613 | 1.13E-07 | IL6_PHA_WB_48h | 0.099699557 | 0.041789917 |
| rs494242 | 9 | 133269706 | chr9:133269706:T:C | T | C | 0.2814 | 0.0517 | 5.39E-08 | IL6_S.aureus_WB_48h | -0.150833851 | 0.041920252 |
| rs644234 | 9 | 133266804 | chr9:133266804:G:T | T | G | -0.2815 | 0.0517 | 5.29E-08 | IL6_S.aureus_WB_48h | 0.150775935 | 0.041996673 |
| rs643434 | 9 | 133266942 | chr9:133266942:A:G | A | G | 0.2815 | 0.0517 | 5.33E-08 | IL6_S.aureus_WB_48h | -0.150775935 | 0.041996673 |
| rs544873 | 9 | 133267800 | chr9:133267800:A:G | A | G | 0.2815 | 0.0517 | 5.29E-08 | IL6_S.aureus_WB_48h | -0.150775935 | 0.041996673 |
| rs543968 | 9 | 133267708 | chr9:133267708:C:T | T | C | -0.2815 | 0.0517 | 5.32E-08 | IL6_S.aureus_WB_48h | 0.152630788 | 0.042506966 |
| rs34492478 | 3 | 46097350 | chr3:46097350:A:T | A | T | -0.3562 | 0.078 | 4.95E-06 | TNFA_C.conidiaHK_WB_48h | 0.26201657 | 0.042751209 |
| rs4797120 | 18 | 3725189 | chr18:3725189:G:C | C | G | -0.2956 | 0.0621 | 1.91E-06 | IL1b_C.conidiaHK_WB_48h | 0.178757786 | 0.044987981 |
| rs494242 | 9 | 133269706 | chr9:133269706:T:C | T | C | 0.2814 | 0.0517 | 5.39E-08 | IL1b_S.aureus_WB_48h | -0.101376558 | 0.047402761 |
| rs644234 | 9 | 133266804 | chr9:133266804:G:T | T | G | -0.2815 | 0.0517 | 5.29E-08 | IL1b_S.aureus_WB_48h | 0.101343751 | 0.047471895 |
| rs643434 | 9 | 133266942 | chr9:133266942:A:G | A | G | 0.2815 | 0.0517 | 5.33E-08 | IL1b_S.aureus_WB_48h | -0.101343751 | 0.047471895 |
| rs544873 | 9 | 133267800 | chr9:133267800:A:G | A | G | 0.2815 | 0.0517 | 5.29E-08 | IL1b_S.aureus_WB_48h | -0.101343751 | 0.047471895 |
| rs543968 | 9 | 133267708 | chr9:133267708:C:T | T | C | -0.2815 | 0.0517 | 5.32E-08 | IL1b_S.aureus_WB_48h | 0.102628326 | 0.047940412 |
| rs1491951 | 3 | 46100352 | chr3:46100352:G:A | A | G | 0.3485 | 0.0777 | 7.30E-06 | TNFA_C.conidiaHK_WB_48h | -0.251560422 | 0.048700477 |
| rs13075758 | 3 | 45983556 | chr3:45983556:G:A | A | G | 0.3672 | 0.0775 | 2.19E-06 | TNFA_C.conidiaHK_WB_48h | -0.261302811 | 0.048918743 |
| rs76860840 | 2 | 15618674 | chr2:15618674:C:G | C | G | 0.3536 | 0.0764 | 3.66E-06 | IFNy_LPS_WB_48h | -0.282429091 | 0.048920042 |
| rs34442130 | 3 | 45983037 | chr3:45983037:T:A | A | T | 0.3674 | 0.0775 | 2.16E-06 | TNFA_C.conidiaHK_WB_48h | -0.261217465 | 0.049009203 |
| rs807581 | 2 | 15662227 | chr2:15662227:T:G | T | G | 0.3698 | 0.0806 | 4.51E-06 | IFNy_LPS_WB_48h | -0.28567455 | 0.049138684 |
| rs76880770 | 2 | 15618488 | chr2:15618488:G:A | A | G | -0.3455 | 0.0767 | 6.62E-06 | IFNy_C.conidiaHK_WB_48h | 0.32637433 | 0.049498387 |
| rs613534 | 9 | 133267707 | chr9:133267707:G:A | A | G | -0.2801 | 0.0517 | 6.16E-08 | IL1b_S.aureus_WB_48h | 0.108041406 | 0.049983041 |

* GWAS loci were selected from The Severe Covid-19 GWAS Group N Engl J Med (2020), doi:10.1056/NEJMoa2020283, all SNP with assoication p value <1e-5 have been tested.

** all normal significant cQTLs have been listed

Table S6 circulating mediators QTLs of COVID-19 GWAS loci from the 500 FG cohort study

| SNP* | chr | pos | variant_id | effect_allele | alternative_a | beta-GWAS | standard_err | p_value-GW. | circulating mediators | beta-QTL | pvalue-QTL** |
|---|---|---|---|---|---|---|---|---|---|---|---|
| rs74586549 | 3 | 45884551 | chr3:4588455 | T | C | 0.3443 | 0.0768 | 7.34E-06 | IL18bpx | 0.1899971 | 3.66E-05 |
| rs13063033 | 3 | 46040989 | chr3:4604098 | A | G | 0.3555 | 0.0772 | 4.12E-06 | IL18bpx | 0.16754042 | 0.000247784 |
| rs34836513 | 3 | 46004967 | chr3:4600496 | A | G | 0.3674 | 0.0772 | 1.96E-06 | IL18bpx | 0.17324257 | 0.000432857 |
| rs34754340 | 3 | 46000345 | chr3:4600034 | T | C | 0.3673 | 0.0772 | 1.94E-06 | IL18bpx | 0.17217991 | 0.00047999 |
| rs71615438 | 3 | 45998376 | chr3:4599837 | A | G | -0.3666 | 0.0772 | 2.04E-06 | IL18bpx | -0.17163046 | 0.000504163 |
| rs36023124 | 3 | 45997568 | chr3:4599756 | C | G | 0.3665 | 0.0771 | 2.01E-06 | IL18bpx | 0.1713777 | 0.000515316 |
| rs17330872 | 3 | 45993605 | chr3:4599360 | A | G | -0.3702 | 0.0774 | 1.74E-06 | IL18bpx | -0.17015104 | 0.000569718 |
| rs35669129 | 3 | 46013758 | chr3:4601375 | A | G | 0.3746 | 0.0768 | 1.08E-06 | IL18bpx | 0.16804016 | 0.000639439 |
| rs13081151 | 3 | 46014224 | chr3:4601422 | A | G | 0.3751 | 0.0777 | 1.38E-06 | IL18bpx | 0.16772623 | 0.000653294 |
| rs13081213 | 3 | 46014417 | chr3:4601441 | T | C | 0.3746 | 0.0768 | 1.08E-06 | IL18bpx | 0.16756082 | 0.00066131 |
| rs35751180 | 3 | 46014681 | chr3:4601468 | T | C | 0.3731 | 0.0774 | 1.42E-06 | IL18bpx | 0.16737087 | 0.000666127 |
| rs34168660 | 3 | 46014670 | chr3:4601467 | A | G | 0.375 | 0.0768 | 1.06E-06 | IL18bpx | 0.16736066 | 0.000670351 |
| rs4362758 | 3 | 46015419 | chr3:4601541 | A | T | 0.3681 | 0.0768 | 1.67E-06 | IL18bpx | 0.16692218 | 0.00069104 |
| rs1491950 | 3 | 46096615 | chr3:4609661 | A | G | 0.3544 | 0.078 | 5.55E-06 | IL18bpx | 0.16232256 | 0.000694358 |
| rs36122610 | 3 | 45981341 | chr3:4598134 | A | G | 0.3673 | 0.0775 | 2.18E-06 | IL18bpx | 0.16422193 | 0.000859403 |
| rs34442130 | 3 | 45983037 | chr3:4598303 | A | T | 0.3674 | 0.0775 | 2.16E-06 | IL18bpx | 0.16403179 | 0.000871926 |
| rs13075758 | 3 | 45983556 | chr3:4598355 | A | G | 0.3672 | 0.0775 | 2.19E-06 | IL18bpx | 0.1639533 | 0.000876453 |
| rs71327014 | 3 | 46059974 | chr3:4605997 | T | C | 0.3556 | 0.0773 | 4.25E-06 | IL18bpx | 0.15596944 | 0.001207194 |
| rs71327015 | 3 | 46060681 | chr3:4606068 | C | G | 0.3585 | 0.0773 | 3.57E-06 | IL18bpx | 0.15596944 | 0.001207194 |
| rs34047915 | 3 | 46058999 | chr3:4605899 | T | C | 0.3555 | 0.0773 | 4.29E-06 | IL18bpx | 0.1559177 | 0.001212128 |
| rs34127208 | 3 | 46062188 | chr3:4606218 | T | G | 0.3554 | 0.0773 | 4.31E-06 | IL18bpx | 0.15588972 | 0.001214257 |
| rs35516580 | 3 | 46066006 | chr3:4606600 | A | G | -0.3562 | 0.0773 | 4.13E-06 | IL18bpx | -0.15587666 | 0.001215134 |
| rs34766614 | 3 | 46066109 | chr3:4606610 | A | G | -0.3484 | 0.0775 | 6.99E-06 | IL18bpx | -0.15587666 | 0.001215134 |
| rs35420565 | 3 | 46058646 | chr3:4605864 | C | G | -0.3556 | 0.0773 | 4.26E-06 | IL18bpx | -0.15586557 | 0.001217099 |
| rs13096741 | 3 | 46057662 | chr3:4605766 | T | C | 0.3556 | 0.0773 | 4.27E-06 | IL18bpx | 0.15565306 | 0.001237349 |
| rs34718164 | 3 | 46057034 | chr3:4605703 | C | G | -0.3556 | 0.0773 | 4.25E-06 | IL18bpx | -0.15554443 | 0.001247699 |
| rs71327006 | 3 | 46017416 | chr3:4601741 | A | G | 0.3628 | 0.0774 | 2.77E-06 | IL18bpx | 0.15531744 | 0.001263879 |
| rs77399277 | 3 | 46055306 | chr3:4605530 | T | C | -0.3556 | 0.0773 | 4.25E-06 | IL18bpx | -0.15515182 | 0.001285138 |
| rs71327007 | 3 | 46017757 | chr3:4601775 | T | C | 0.3643 | 0.0774 | 2.51E-06 | IL18bpx | 0.15506248 | 0.001285925 |
| rs34340587 | 3 | 46054551 | chr3:4605455 | A | G | 0.3542 | 0.0773 | 4.63E-06 | IL18bpx | 0.15512853 | 0.001287782 |
| rs35454877 | 3 | 46017992 | chr3:4601799 | T | C | -0.3679 | 0.0768 | 1.69E-06 | IL18bpx | -0.1548939 | 0.001301062 |
| rs35772789 | 3 | 46053612 | chr3:4605361 | A | G | -0.3544 | 0.0773 | 4.52E-06 | IL18bpx | -0.15493856 | 0.001305521 |
| rs34863575 | 3 | 46052571 | chr3:4605257 | A | G | -0.3543 | 0.0773 | 4.53E-06 | IL18bpx | -0.15469777 | 0.001328492 |
| rs73833520 | 3 | 46016375 | chr3:4601637 | A | G | -0.3679 | 0.0768 | 1.69E-06 | IL18bpx | -0.15510175 | 0.001397584 |
| rs34679077 | 3 | 46046500 | chr3:4604650 | A | G | 0.3609 | 0.0774 | 3.13E-06 | IL18bpx | 0.15353781 | 0.001440524 |
| rs34452002 | 3 | 46101695 | chr3:4610169 | T | C | 0.3556 | 0.0779 | 4.95E-06 | IL18bpx | 0.1529439 | 0.001459479 |
| rs34492478 | 3 | 46097350 | chr3:4609735 | A | T | -0.3562 | 0.078 | 4.95E-06 | IL18bpx | -0.15308226 | 0.001463484 |
| rs34093271 | 3 | 46103489 | chr3:4610348 | T | G | 0.3558 | 0.0779 | 4.89E-06 | IL18bpx | 0.1529112 | 0.001469955 |
| rs35334665 | 3 | 46019726 | chr3:4601972 | T | C | -0.3632 | 0.0774 | 2.70E-06 | IL18bpx | -0.15285902 | 0.001495841 |
| rs13093179 | 3 | 46104822 | chr3:4610482 | T | G | 0.3557 | 0.0779 | 4.91E-06 | IL18bpx | 0.15477734 | 0.001496667 |
| rs36040135 | 3 | 46019812 | chr3:4601981 | A | G | -0.3679 | 0.0768 | 1.69E-06 | IL18bpx | -0.15276741 | 0.001504381 |
| rs34438204 | 3 | 46039814 | chr3:4603981 | T | C | -0.3609 | 0.0774 | 3.13E-06 | IL18bpx | -0.1528602 | 0.00150683 |
| rs2088692 | 3 | 46035840 | chr3:4603584 | A | G | -0.3654 | 0.0768 | 1.96E-06 | IL18bpx | -0.15234698 | 0.001558118 |
| rs34460587 | 3 | 46100972 | chr3:4610097 | T | C | 0.3557 | 0.0779 | 4.93E-06 | IL18bpx | 0.15188848 | 0.001567443 |
| rs13074382 | 3 | 46020348 | chr3:4602034 | A | G | -0.3679 | 0.0768 | 1.69E-06 | IL18bpx | -0.15213165 | 0.00156983 |
| rs6791016 | 3 | 46033366 | chr3:4603336 | A | G | 0.367 | 0.0769 | 1.80E-06 | IL18bpx | 0.15210772 | 0.001582058 |
| rs6780028 | 3 | 46033598 | chr3:4603359 | A | C | -0.367 | 0.0769 | 1.80E-06 | IL18bpx | -0.15210772 | 0.001582058 |
| rs13097556 | 3 | 46020505 | chr3:4602050 | T | C | -0.3679 | 0.0768 | 1.69E-06 | IL18bpx | -0.15195123 | 0.00158869 |
| rs6790866 | 3 | 46033219 | chr3:4603321 | A | G | 0.367 | 0.0769 | 1.80E-06 | IL18bpx | 0.15203184 | 0.001589767 |
| rs35814488 | 3 | 46032431 | chr3:4603243 | A | G | -0.3672 | 0.0769 | 1.77E-06 | IL18bpx | -0.15195558 | 0.001597531 |
| rs34774687 | 3 | 46031822 | chr3:4603182 | T | C | -0.367 | 0.0769 | 1.80E-06 | IL18bpx | -0.15187896 | 0.001605352 |
| rs13060287 | 3 | 46030376 | chr3:4603037 | A | G | -0.3671 | 0.0769 | 1.79E-06 | IL18bpx | -0.15181595 | 0.001610897 |
| rs34619093 | 3 | 46031027 | chr3:4603102 | A | G | 0.3625 | 0.0775 | 2.89E-06 | IL18bpx | 0.15180198 | 0.001613228 |
| rs68087193 | 3 | 46031138 | chr3:4603113 | T | C | 0.3673 | 0.0769 | 1.76E-06 | IL18bpx | 0.15180198 | 0.001613228 |
| rs2102055 | 3 | 46031212 | chr3:4603121 | A | G | 0.3591 | 0.0767 | 2.83E-06 | IL18bpx | 0.15180198 | 0.001613228 |
| rs2102056 | 3 | 46031276 | chr3:4603127 | A | G | 0.3671 | 0.0769 | 1.78E-06 | IL18bpx | 0.15180198 | 0.001613228 |
| rs2088690 | 3 | 46031347 | chr3:4603134 | T | C | 0.3671 | 0.0769 | 1.78E-06 | IL18bpx | 0.15180198 | 0.001613228 |
| rs13082697 | 3 | 46030271 | chr3:4603027 | T | C | -0.3671 | 0.0769 | 1.78E-06 | IL18bpx | -0.15173823 | 0.001618841 |
| rs34562820 | 3 | 46029589 | chr3:4602958 | A | G | 0.3625 | 0.0775 | 2.87E-06 | IL18bpx | 0.15166015 | 0.001626843 |
| rs59166269 | 3 | 46028845 | chr3:4602884 | C | G | -0.3672 | 0.0769 | 1.77E-06 | IL18bpx | -0.15158171 | 0.001634902 |
| rs57437758 | 3 | 46028861 | chr3:4602886 | A | C | -0.3672 | 0.0769 | 1.77E-06 | IL18bpx | -0.15158171 | 0.001634902 |
| rs13090194 | 3 | 46028081 | chr3:4602808 | A | C | -0.3672 | 0.0769 | 1.77E-06 | IL18bpx | -0.15150291 | 0.001643019 |
| rs71327010 | 3 | 46027343 | chr3:4602734 | T | G | 0.3625 | 0.0774 | 2.83E-06 | IL18bpx | 0.15134423 | 0.001659429 |
| rs35159820 | 3 | 46026981 | chr3:4602698 | A | G | -0.3673 | 0.0769 | 1.76E-06 | IL18bpx | -0.15126435 | 0.001667723 |
| rs35161099 | 3 | 46026015 | chr3:4602601 | T | G | -0.3625 | 0.0774 | 2.83E-06 | IL18bpx | -0.15118411 | 0.001676076 |
| rs71327017 | 3 | 46069721 | chr3:4606972 | T | C | -0.3576 | 0.0774 | 3.80E-06 | IL18bpx | -0.15078704 | 0.00168509 |
| rs13072267 | 3 | 46074518 | chr3:4607451 | A | T | 0.3543 | 0.0773 | 4.57E-06 | IL18bpx | 0.15078704 | 0.00168509 |
| rs36057789 | 3 | 46075604 | chr3:4607560 | A | G | -0.3551 | 0.0773 | 4.36E-06 | IL18bpx | -0.15078704 | 0.00168509 |
| rs76647202 | 3 | 46076468 | chr3:4607646 | A | G | -0.3542 | 0.0773 | 4.60E-06 | IL18bpx | -0.15078704 | 0.00168509 |
| rs71327021 | 3 | 46079025 | chr3:4607902 | A | G | -0.3542 | 0.0773 | 4.61E-06 | IL18bpx | -0.15078704 | 0.00168509 |
| rs13091868 | 3 | 46081551 | chr3:4608155 | A | G | -0.3538 | 0.0773 | 4.72E-06 | IL18bpx | -0.15077334 | 0.001686314 |
| rs13092030 | 3 | 46081563 | chr3:4608156 | T | C | 0.354 | 0.0773 | 4.66E-06 | IL18bpx | 0.15077334 | 0.001686314 |
| rs17215981 | 3 | 46082926 | chr3:4608292 | A | G | -0.3547 | 0.0773 | 4.47E-06 | IL18bpx | -0.15077334 | 0.001686314 |
| rs13089554 | 3 | 46086065 | chr3:4608606 | A | G | 0.3547 | 0.0774 | 4.61E-06 | IL18bpx | 0.15077334 | 0.001686314 |
| rs13095717 | 3 | 46087050 | chr3:4608705 | A | G | 0.3548 | 0.0774 | 4.58E-06 | IL18bpx | 0.15077334 | 0.001686314 |
| rs13075528 | 3 | 46087217 | chr3:4608721 | T | C | -0.3549 | 0.0775 | 4.63E-06 | IL18bpx | -0.15077334 | 0.001686314 |
| rs13095602 | 3 | 46087221 | chr3:4608722 | A | G | -0.3547 | 0.0774 | 4.60E-06 | IL18bpx | -0.15077334 | 0.001686314 |
| rs71327023 | 3 | 46089733 | chr3:4608973 | C | G | -0.3539 | 0.0777 | 5.19E-06 | IL18bpx | -0.15077334 | 0.001686314 |
| rs13063527 | 3 | 46094198 | chr3:4609419 | A | G | 0.3561 | 0.0777 | 4.51E-06 | IL18bpx | 0.15077334 | 0.001686314 |
| rs13068570 | 3 | 46094946 | chr3:4609494 | A | G | -0.3561 | 0.0777 | 4.61E-06 | IL18bpx | -0.15077334 | 0.001686314 |
| rs4373103 | 3 | 46096963 | chr3:4609696 | A | G | 0.3547 | 0.078 | 5.42E-06 | IL18bpx | 0.15077334 | 0.001686314 |
| rs71327024 | 3 | 46098581 | chr3:4609858 | T | G | 0.3547 | 0.078 | 5.37E-06 | IL18bpx | 0.15077334 | 0.001686314 |
| rs71327025 | 3 | 46106012 | chr3:4610601 | A | G | -0.3556 | 0.0779 | 4.96E-06 | IL18bpx | -0.15130608 | 0.001700891 |
| rs35930050 | 3 | 46024271 | chr3:4602427 | A | G | 0.3681 | 0.0768 | 1.67E-06 | IL18bpx | 0.15085961 | 0.001710092 |
| rs34584867 | 3 | 46024471 | chr3:4602447 | A | G | 0.3681 | 0.0768 | 1.67E-06 | IL18bpx | 0.15085961 | 0.001710092 |
| rs876668 | 3 | 46022261 | chr3:4602226 | T | C | -0.368 | 0.0768 | 1.68E-06 | IL18bpx | -0.15052946 | 0.001745095 |
| rs35539222 | 3 | 46112780 | chr3:4611278 | T | C | -0.3556 | 0.0779 | 4.96E-06 | IL18bpx | -0.15070589 | 0.00178474 |
| rs35110864 | 3 | 46112965 | chr3:4611296 | A | G | 0.3555 | 0.0779 | 4.99E-06 | IL18bpx | 0.15070589 | 0.00178474 |
| rs76880770 | 2 | 15618488 | chr2:1561848 | A | G | -0.3455 | 0.0767 | 6.62E-06 | AAT | 0.27981077 | 0.002302198 |
| rs2230322 | 3 | 46021837 | chr3:4602183 | T | C | -0.368 | 0.0768 | 1.68E-06 | IL18bpx | -0.14449079 | 0.002531372 |
| rs1491951 | 3 | 46100352 | chr3:4610035 | A | G | 0.3485 | 0.0777 | 7.30E-06 | IL18bpx | 0.13960947 | 0.003417858 |
| rs76860840 | 2 | 15618674 | chr2:1561867 | C | G | 0.3536 | 0.0764 | 3.66E-06 | AAT | -0.23072285 | 0.007962754 |
| rs807636 | 2 | 15627828 | chr2:1562782 | T | C | -0.365 | 0.0772 | 2.25E-06 | AAT | 0.22957614 | 0.008413778 |
| rs807635 | 2 | 15629483 | chr2:1562948 | T | C | -0.3579 | 0.0765 | 2.85E-06 | AAT | 0.22876688 | 0.008565765 |
| rs41264195 | 2 | 15606096 | chr2:1560609 | T | C | -0.3359 | 0.0754 | 8.49E-06 | AAT | 0.2215509 | 0.009931056 |
| rs3770462 | 2 | 15609200 | chr2:1560920 | T | G | 0.3409 | 0.0757 | 6.74E-06 | AAT | -0.22147388 | 0.009950404 |
| rs4668941 | 2 | 15610007 | chr2:1561000 | T | G | 0.3359 | 0.0754 | 8.51E-06 | AAT | -0.22147388 | 0.009950404 |
| rs79808866 | 2 | 15604860 | chr2:1560486 | T | G | 0.3357 | 0.0754 | 8.59E-06 | AAT | -0.22138295 | 0.009987049 |

* GWAS loci were selected from The Severe Covid-19 GWAS Group N Engl J Med (2020), doi:10.1056/NEJMoa2020283, all SNP with assoication p value <1e-5 have been tested.

** all circulating medators QTLs(P value <0.01) have been listed

Table S7 metabolites QTLs of COVID-19 GWAS loci from the 500 FG cohort study

| SNP* | chr | pos | variant_id | effect_allele | alternative_ | beta-GWAS | standard_error-GWAS | p_value-GWAS | metabolite_ | pvalue-QTL* | beta-QTL | annotation |
|---|---|---|---|---|---|---|---|---|---|---|---|---|
| rs11174294 | 12 | 62072129 | chr12:620721 | T | C | -0.29 | 0.0604 | 1.59E-06 | Tyr | 0.00077507 | -0.28459127 | Tyrosine(mmol/l) |
| rs10877787 | 12 | 62072981 | chr12:620729 | A | G | 0.29 | 0.0604 | 1.58E-06 | Tyr | 0.00081522 | 0.2833966 | Tyrosine(mmol/l) |
| rs9706516 | 12 | 62074403 | chr12:620744 | A | C | -0.2923 | 0.0604 | 1.30E-06 | Tyr | 0.00081556 | -0.28340664 | Tyrosine(mmol/l) |
| rs10747951 | 12 | 62075677 | chr12:620756 | T | C | -0.29 | 0.0604 | 1.58E-06 | Tyr | 0.00081595 | -0.28341422 | Tyrosine(mmol/l) |
| rs10506432 | 12 | 62077116 | chr12:620771 | A | C | 0.29 | 0.0604 | 1.58E-06 | Tyr | 0.00081675 | 0.28341487 | Tyrosine(mmol/l) |
| rs11174296 | 12 | 62077677 | chr12:620776 | C | G | 0.288 | 0.0602 | 1.75E-06 | Tyr | 0.00081675 | 0.28341487 | Tyrosine(mmol/l) |
| rs9738190 | 12 | 62078790 | chr12:620787 | A | G | -0.2898 | 0.0602 | 1.51E-06 | Tyr | 0.0008243 | -0.2832527 | Tyrosine(mmol/l) |
| rs12610495 | 19 | 4717660 | chr19:471766 | A | G | -0.2534 | 0.0556 | 5.20E-06 | XS_VLDL_FC | 0.00104814 | -0.25543589 | Free cholesterol in very small VLDL(mmol/l) |
| rs12610495 | 19 | 4717660 | chr19:471766 | A | G | -0.2534 | 0.0556 | 5.20E-06 | FreeC | 0.00113189 | -0.24482735 | Free cholesterol(mmol/l) |
| rs12610495 | 19 | 4717660 | chr19:471766 | A | G | -0.2534 | 0.0556 | 5.20E-06 | L_LDL_PL_pe | 0.0013018 | 0.24568176 | Phospholipids to total lipds ratio in large LDL(%) |
| rs543968 | 9 | 133267708 | chr9:1332677 | T | C | -0.2815 | 0.0517 | 5.32E-08 | XXL_VLDL_PI | 0.00132333 | 0.22949061 | Phospholipids to total lipds ratio in chylomicrons and extremely large VLDL(%) |
| rs657152 | 9 | 133263862 | chr9:1332638 | A | C | 0.2814 | 0.0516 | 4.95E-08 | XXL_VLDL_PI | 0.00137206 | -0.22500066 | Phospholipids to total lipds ratio in chylomicrons and extremely large VLDL(%) |
| rs644234 | 9 | 133266804 | chr9:1332668 | T | G | -0.2815 | 0.0517 | 5.29E-08 | XXL_VLDL_PI | 0.00137206 | 0.22500066 | Phospholipids to total lipds ratio in chylomicrons and extremely large VLDL(%) |
| rs643434 | 9 | 133266942 | chr9:1332669 | A | G | 0.2815 | 0.0517 | 5.33E-08 | XXL_VLDL_PI | 0.00137206 | -0.22500066 | Phospholipids to total lipds ratio in chylomicrons and extremely large VLDL(%) |
| rs544873 | 9 | 133267800 | chr9:1332678 | A | G | 0.2815 | 0.0517 | 5.29E-08 | XXL_VLDL_PI | 0.00137206 | -0.22500066 | Phospholipids to total lipds ratio in chylomicrons and extremely large VLDL(%) |
| rs494242 | 9 | 133269706 | chr9:1332697 | T | C | 0.2814 | 0.0517 | 5.39E-08 | XXL_VLDL_PI | 0.00137206 | -0.22500066 | Phospholipids to total lipds ratio in chylomicrons and extremely large VLDL(%) |
| rs12610495 | 19 | 4717660 | chr19:471766 | A | G | -0.2534 | 0.0556 | 5.20E-06 | XS_VLDL_C | 0.00137763 | -0.25386109 | Total cholesterol in very small VLDL(mmol/l) |
| rs613534 | 9 | 133267707 | chr9:1332677 | A | G | -0.2801 | 0.0517 | 6.16E-08 | XXL_VLDL_PI | 0.0016994 | 0.2398102 | Phospholipids to total lipds ratio in chylomicrons and extremely large VLDL(%) |
| rs11627388 | 14 | 31953140 | chr14:319531 | A | C | -0.2482 | 0.0533 | 3.19E-06 | L_HDL_CE_pe | 0.0017326 | -0.24323013 | Cholesterol esters to total lipids ratio in large HDL(%) |
| rs10506436 | 12 | 62028786 | chr12:620287 | A | T | -0.2811 | 0.06 | 2.84E-06 | Gln | 0.00179994 | -0.24665616 | Glutamine(mmol/l) |
| rs9739464 | 12 | 62037992 | chr12:620379 | A | T | 0.2846 | 0.06 | 2.10E-06 | Gln | 0.00181212 | 0.24649242 | Glutamine(mmol/l) |
| rs12610495 | 19 | 4717660 | chr19:471766 | A | G | -0.2534 | 0.0556 | 5.20E-06 | XS_VLDL_L | 0.00181916 | -0.24019062 | Total lipids in very small VLDL(mmol/l) |
| rs10784278 | 12 | 62042165 | chr12:620421 | A | T | -0.2836 | 0.06 | 2.32E-06 | Gln | 0.00182029 | -0.24597573 | Glutamine(mmol/l) |
| rs10784279 | 12 | 62044016 | chr12:620440 | T | C | -0.2778 | 0.0599 | 3.50E-06 | Gln | 0.00183772 | -0.24566899 | Glutamine(mmol/l) |
| rs12610495 | 19 | 4717660 | chr19:471766 | A | G | -0.2534 | 0.0556 | 5.20E-06 | XS_VLDL_CE | 0.00206037 | -0.24605735 | Cholesterol esters in very small VLDL(mmol/l) |
| rs12610495 | 19 | 4717660 | chr19:471766 | A | G | -0.2534 | 0.0556 | 5.20E-06 | IDL_P | 0.00209362 | -0.23454865 | Concentration of IDL particles(mol/l) |
| rs687621 | 9 | 133261662 | chr9:1332616 | A | G | -0.2837 | 0.0528 | 7.63E-08 | XXL_VLDL_PI | 0.00211881 | 0.21723448 | Phospholipids to total lipds ratio in chylomicrons and extremely large VLDL(%) |
| rs687289 | 9 | 133261703 | chr9:1332617 | A | G | 0.2826 | 0.0527 | 8.14E-08 | XXL_VLDL_PI | 0.00211881 | -0.21723448 | Phospholipids to total lipds ratio in chylomicrons and extremely large VLDL(%) |
| rs545971 | 9 | 133267960 | chr9:1332679 | T | C | 0.2794 | 0.0524 | 9.58E-08 | XXL_VLDL_PI | 0.00211881 | -0.21723448 | Phospholipids to total lipds ratio in chylomicrons and extremely large VLDL(%) |
| rs612169 | 9 | 133268030 | chr9:1332680 | A | G | -0.2797 | 0.0527 | 1.08E-07 | XXL_VLDL_PI | 0.00211881 | 0.21723448 | Phospholipids to total lipds ratio in chylomicrons and extremely large VLDL(%) |
| rs597988 | 9 | 133268872 | chr9:1332688 | A | T | 0.2796 | 0.0526 | 1.07E-07 | XXL_VLDL_PI | 0.00211881 | -0.21723448 | Phospholipids to total lipds ratio in chylomicrons and extremely large VLDL(%) |
| rs597974 | 9 | 133268885 | chr9:1332688 | A | G | -0.2796 | 0.0526 | 1.07E-07 | XXL_VLDL_PI | 0.00211881 | 0.21723448 | Phospholipids to total lipds ratio in chylomicrons and extremely large VLDL(%) |
| rs8176663 | 9 | 133269015 | chr9:1332690 | T | C | -0.2798 | 0.0526 | 1.05E-07 | XXL_VLDL_PI | 0.00211881 | 0.21723448 | Phospholipids to total lipds ratio in chylomicrons and extremely large VLDL(%) |
| rs491626 | 9 | 133269461 | chr9:1332694 | T | C | 0.2797 | 0.0526 | 1.05E-07 | XXL_VLDL_PI | 0.00211881 | -0.21723448 | Phospholipids to total lipds ratio in chylomicrons and extremely large VLDL(%) |
| rs492488 | 9 | 133269548 | chr9:1332695 | A | G | 0.2798 | 0.0526 | 1.02E-07 | XXL_VLDL_PI | 0.00211881 | -0.21723448 | Phospholipids to total lipds ratio in chylomicrons and extremely large VLDL(%) |
| rs493246 | 9 | 133269582 | chr9:1332695 | A | G | 0.2799 | 0.0526 | 1.01E-07 | XXL_VLDL_PI | 0.00211881 | -0.21723448 | Phospholipids to total lipds ratio in chylomicrons and extremely large VLDL(%) |
| rs495203 | 9 | 133269828 | chr9:1332698 | T | C | 0.2816 | 0.0526 | 8.52E-08 | XXL_VLDL_PI | 0.00211881 | -0.21723448 | Phospholipids to total lipds ratio in chylomicrons and extremely large VLDL(%) |
| rs582118 | 9 | 133270061 | chr9:1332700 | A | G | -0.2798 | 0.0526 | 1.02E-07 | XXL_VLDL_PI | 0.00211881 | 0.21723448 | Phospholipids to total lipds ratio in chylomicrons and extremely large VLDL(%) |
| rs582094 | 9 | 133270074 | chr9:1332700 | A | T | -0.2796 | 0.0526 | 1.04E-07 | XXL_VLDL_PI | 0.00211881 | 0.21723448 | Phospholipids to total lipds ratio in chylomicrons and extremely large VLDL(%) |
| rs2769071 | 9 | 133270565 | chr9:1332705 | A | G | -0.2785 | 0.0526 | 1.17E-07 | XXL_VLDL_PI | 0.00211881 | 0.21723448 | Phospholipids to total lipds ratio in chylomicrons and extremely large VLDL(%) |
| rs677355 | 9 | 133270615 | chr9:1332706 | A | G | 0.2789 | 0.0526 | 1.12E-07 | XXL_VLDL_PI | 0.00211881 | -0.21723448 | Phospholipids to total lipds ratio in chylomicrons and extremely large VLDL(%) |
| rs676996 | 9 | 133270647 | chr9:1332706 | T | G | -0.2785 | 0.0526 | 1.17E-07 | XXL_VLDL_PI | 0.00211881 | 0.21723448 | Phospholipids to total lipds ratio in chylomicrons and extremely large VLDL(%) |
| rs676457 | 9 | 133270797 | chr9:1332707 | A | T | -0.2783 | 0.0526 | 1.21E-07 | XXL_VLDL_PI | 0.00211881 | 0.21723448 | Phospholipids to total lipds ratio in chylomicrons and extremely large VLDL(%) |
| rs674302 | 9 | 133271249 | chr9:1332712 | A | T | 0.278 | 0.0526 | 1.26E-07 | XXL_VLDL_PI | 0.00211881 | -0.21723448 | Phospholipids to total lipds ratio in chylomicrons and extremely large VLDL(%) |
| rs554833 | 9 | 133271745 | chr9:1332717 | T | C | 0.2788 | 0.0526 | 1.13E-07 | XXL_VLDL_PI | 0.00211881 | -0.21723448 | Phospholipids to total lipds ratio in chylomicrons and extremely large VLDL(%) |
| rs505922 | 9 | 133273813 | chr9:1332738 | T | C | -0.2687 | 0.0526 | 3.21E-07 | XXL_VLDL_PI | 0.00211881 | 0.21723448 | Phospholipids to total lipds ratio in chylomicrons and extremely large VLDL(%) |
| rs514659 | 9 | 133266790 | chr9:1332667 | A | C | -0.2811 | 0.0526 | 9.08E-08 | XXL_VLDL_PI | 0.00211881 | 0.21723448 | Phospholipids to total lipds ratio in chylomicrons and extremely large VLDL(%) |
| rs10088118 | 8 | 121806381 | chr8:1218063 | C | G | -0.2329 | 0.0517 | 6.79E-06 | S_HDL_P | 0.00212094 | 0.21874102 | Concentration of small HDL particles(mol/l) |
| rs529565 | 9 | 133274084 | chr9:1332740 | T | C | -0.2717 | 0.0526 | 2.44E-07 | XXL_VLDL_PI | 0.00212809 | 0.21716031 | Phospholipids to total lipds ratio in chylomicrons and extremely large VLDL(%) |
| rs10088118 | 8 | 121806381 | chr8:1218063 | C | G | -0.2329 | 0.0517 | 6.79E-06 | S_HDL_L | 0.00220717 | 0.21790333 | Total lipids in small HDL(mmol/l) |
| rs12610495 | 19 | 4717660 | chr19:471766 | A | G | -0.2534 | 0.0556 | 5.20E-06 | S_LDL_L | 0.00223668 | -0.23555325 | Total lipids in small LDL(mmol/l) |
| rs12610495 | 19 | 4717660 | chr19:471766 | A | G | -0.2534 | 0.0556 | 5.20E-06 | S_LDL_C | 0.0022564 | -0.23951062 | Total cholesterol in small LDL(mmol/l) |
| rs12610495 | 19 | 4717660 | chr19:471766 | A | G | -0.2534 | 0.0556 | 5.20E-06 | IDL_L | 0.00226183 | -0.23414115 | Total lipids in IDL(mmol/l) |
| rs12610495 | 19 | 4717660 | chr19:471766 | A | G | -0.2534 | 0.0556 | 5.20E-06 | XS_VLDL_P | 0.00226957 | -0.23416529 | Concentration of very small VLDL particles(mol/l) |
| rs12610495 | 19 | 4717660 | chr19:471766 | A | G | -0.2534 | 0.0556 | 5.20E-06 | S_LDL_P | 0.00228494 | -0.23375057 | Concentration of small LDL particles(mol/l) |
| rs12610495 | 19 | 4717660 | chr19:471766 | A | G | -0.2534 | 0.0556 | 5.20E-06 | M_LDL_PL_p | 0.00237082 | 0.2484 | Phospholipids to total lipds ratio in medium LDL(%) |
| rs2302929 | 2 | 15595524 | chr2:1559552 | T | C | 0.2976 | 0.0672 | 9.54E-06 | M_HDL_CE | 0.00239135 | 0.27679654 | Cholesterol esters in medium HDL(mmol/l) |
| rs1455664 | 4 | 179425171 | chr4:1794251 | T | C | -0.3158 | 0.0714 | 9.77E-06 | XL_VLDL_CE | 0.00242694 | -0.33614538 | Cholesterol esters in very large VLDL(mmol/l) |
| rs12610495 | 19 | 4717660 | chr19:471766 | A | G | -0.2534 | 0.0556 | 5.20E-06 | S_LDL_PL_pe | 0.00246805 | 0.24089845 | Phospholipids to total lipds ratio in small LDL(%) |
| rs1455662 | 4 | 179423719 | chr4:1794237 | A | G | -0.3156 | 0.0713 | 9.60E-06 | XL_VLDL_CE | 0.00248159 | -0.33514763 | Cholesterol esters in very large VLDL(mmol/l) |
| rs1455664 | 4 | 179425171 | chr4:1794251 | T | C | -0.3158 | 0.0714 | 9.77E-06 | XL_VLDL_C | 0.00260126 | -0.33489917 | Total cholesterol in very large VLDL(mmol/l) |
| rs1455662 | 4 | 179423719 | chr4:1794237 | A | G | -0.3156 | 0.0713 | 9.60E-06 | XL_VLDL_C | 0.00265632 | -0.33393953 | Total cholesterol in very large VLDL(mmol/l) |
| rs1455663 | 4 | 179425076 | chr4:1794250 | A | G | -0.3156 | 0.0714 | 9.87E-06 | XL_VLDL_CE | 0.00266452 | -0.33426232 | Cholesterol esters in very large VLDL(mmol/l) |
| rs9739956 | 12 | 62065731 | chr12:620657 | T | C | 0.2934 | 0.0598 | 9.45E-07 | Tyr | 0.00269877 | 0.25355473 | Tyrosine(mmol/l) |
| rs10877786 | 12 | 62070198 | chr12:620701 | T | G | 0.2964 | 0.0597 | 6.89E-07 | Tyr | 0.00270028 | 0.25341486 | Tyrosine(mmol/l) |
| rs12610495 | 19 | 4717660 | chr19:471766 | A | G | -0.2534 | 0.0556 | 5.20E-06 | IDL_C | 0.00273164 | -0.23350236 | Total cholesterol in IDL(mmol/l) |
| rs12610495 | 19 | 4717660 | chr19:471766 | A | G | -0.2534 | 0.0556 | 5.20E-06 | M_LDL_P | 0.00275333 | -0.22717906 | Concentration of medium LDL particles(mol/l) |
| rs1455663 | 4 | 179425076 | chr4:1794250 | A | G | -0.3156 | 0.0714 | 9.87E-06 | XL_VLDL_C | 0.00285405 | -0.33300409 | Total cholesterol in very large VLDL(mmol/l) |
| rs12610495 | 19 | 4717660 | chr19:471766 | A | G | -0.2534 | 0.0556 | 5.20E-06 | S_LDL_FC | 0.00286212 | -0.23691754 | Free cholesterol in small LDL(mmol/l) |
| rs12610495 | 19 | 4717660 | chr19:471766 | A | G | -0.2534 | 0.0556 | 5.20E-06 | M_LDL_L | 0.00288057 | -0.22726546 | Total lipids in medium LDL(mmol/l) |
| rs4389947 | 8 | 121816361 | chr8:1218163 | A | G | -0.2364 | 0.0517 | 4.91E-06 | S_HDL_P | 0.00289219 | 0.2126477 | Concentration of small HDL particles(mol/l) |
| rs12610495 | 19 | 4717660 | chr19:471766 | A | G | -0.2534 | 0.0556 | 5.20E-06 | M_LDL_C | 0.00292999 | -0.23037104 | Total cholesterol in medium LDL(mmol/l) |
| rs2191031 | 3 | 45869378 | chr3:4586937 | A | G | 0.2758 | 0.0613 | 6.80E-06 | XL_HDL_CE | 0.00293241 | -0.29984707 | Cholesterol esters in very large HDL(mmol/l) |
| rs7152677 | 14 | 31953540 | chr14:319535 | C | G | 0.2637 | 0.0548 | 1.52E-06 | XS_VLDL_FC | 0.00294202 | -0.24979916 | Free cholesterol in very small VLDL(mmol/l) |
| rs12610495 | 19 | 4717660 | chr19:471766 | A | G | -0.2534 | 0.0556 | 5.20E-06 | IDL_PL | 0.00298407 | -0.22445532 | Phospholipids in IDL(mmol/l) |
| rs12610495 | 19 | 4717660 | chr19:471766 | A | G | -0.2534 | 0.0556 | 5.20E-06 | EstC | 0.00301243 | -0.2271389 | Esterified cholesterol(mmol/l) |
| rs4389947 | 8 | 121816361 | chr8:1218163 | A | G | -0.2364 | 0.0517 | 4.91E-06 | S_HDL_L | 0.00301322 | 0.21175978 | Total lipids in small HDL(mmol/l) |
| rs12610495 | 19 | 4717660 | chr19:471766 | A | G | -0.2534 | 0.0556 | 5.20E-06 | L_LDL_P | 0.00303833 | -0.22288012 | Concentration of large LDL particles(mol/l) |
| rs12610495 | 19 | 4717660 | chr19:471766 | A | G | -0.2534 | 0.0556 | 5.20E-06 | M_LDL_FC_p | 0.00308085 | 0.21862327 | Free cholesterol to total lipids ratio in medium LDL(%) |
| rs12610495 | 19 | 4717660 | chr19:471766 | A | G | -0.2534 | 0.0556 | 5.20E-06 | IDL_CE | 0.00309913 | -0.23100645 | Cholesterol esters in IDL(mmol/l) |
| rs12610495 | 19 | 4717660 | chr19:471766 | A | G | -0.2534 | 0.0556 | 5.20E-06 | M_LDL_CE_p | 0.00310702 | -0.23959055 | Cholesterol esters to total lipids ratio in medium LDL(%) |
| rs12610495 | 19 | 4717660 | chr19:471766 | A | G | -0.2534 | 0.0556 | 5.20E-06 | L_LDL_L | 0.00314084 | -0.223198 | Total lipids in large LDL(mmol/l) |
| rs12610495 | 19 | 4717660 | chr19:471766 | A | G | -0.2534 | 0.0556 | 5.20E-06 | Serum_C | 0.00320237 | -0.22649553 | Serum total cholesterol(mmol/l) |
| rs7152677 | 14 | 31953540 | chr14:319535 | C | G | 0.2637 | 0.0548 | 1.52E-06 | L_VLDL_C_pe | 0.00323394 | -0.24769607 | Total cholesterol to total lipids ratio in large VLDL(%) |
| rs12610495 | 19 | 4717660 | chr19:471766 | A | G | -0.2534 | 0.0556 | 5.20E-06 | L_LDL_CE | 0.00334093 | -0.22241531 | Cholesterol esters in large LDL(mmol/l) |
| rs633862 | 9 | 133279871 | chr9:1332798 | T | C | 0.2534 | 0.0515 | 8.79E-07 | L_VLDL_CE_p | 0.00346971 | 0.21409471 | Cholesterol esters to total lipids ratio in large VLDL(%) |
| rs12610495 | 19 | 4717660 | chr19:471766 | A | G | -0.2534 | 0.0556 | 5.20E-06 | L_LDL_C | 0.00349195 | -0.22218014 | Total cholesterol in large LDL(mmol/l) |
| rs12610495 | 19 | 4717660 | chr19:471766 | A | G | -0.2534 | 0.0556 | 5.20E-06 | IDL_FC | 0.0035062 | -0.22747391 | Free cholesterol in IDL(mmol/l) |
| rs660340 | 9 | 133272138 | chr9:1332721 | A | G | -0.255 | 0.0517 | 7.97E-07 | L_VLDL_PL_p | 0.00360335 | 0.20655516 | Phospholipids to total lipds ratio in large VLDL(%) |
| rs581107 | 9 | 133272287 | chr9:1332722 | T | C | 0.2553 | 0.0517 | 7.87E-07 | L_VLDL_PL_p | 0.00360335 | -0.20655516 | Phospholipids to total lipds ratio in large VLDL(%) |
| rs659104 | 9 | 133272408 | chr9:1332724 | T | G | -0.255 | 0.0517 | 7.97E-07 | L_VLDL_PL_p | 0.00360335 | 0.20655516 | Phospholipids to total lipds ratio in large VLDL(%) |
| rs473533 | 9 | 133272620 | chr9:1332726 | T | C | -0.2551 | 0.0517 | 7.89E-07 | L_VLDL_PL_p | 0.00360335 | 0.20655516 | Phospholipids to total lipds ratio in large VLDL(%) |
| rs475419 | 9 | 133272816 | chr9:1332728 | T | C | 0.2576 | 0.0517 | 6.32E-07 | L_VLDL_PL_p | 0.00360335 | -0.20655516 | Phospholipids to total lipds ratio in large VLDL(%) |
| rs476410 | 9 | 133272953 | chr9:1332729 | C | G | 0.2538 | 0.0517 | 8.96E-07 | L_VLDL_PL_p | 0.00360335 | -0.20655516 | Phospholipids to total lipds ratio in large VLDL(%) |
| rs645982 | 9 | 133272994 | chr9:1332729 | A | G | -0.2536 | 0.0516 | 9.02E-07 | L_VLDL_PL_p | 0.00360335 | 0.20655516 | Phospholipids to total lipds ratio in large VLDL(%) |
| rs500498 | 9 | 133273232 | chr9:1332732 | T | C | -0.2535 | 0.0516 | 9.06E-07 | L_VLDL_PL_p | 0.00361089 | 0.20649887 | Phospholipids to total lipds ratio in large VLDL(%) |
| rs500499 | 9 | 133273233 | chr9:1332732 | C | G | 0.2535 | 0.0516 | 9.06E-07 | L_VLDL_PL_p | 0.00361089 | -0.20649887 | Phospholipids to total lipds ratio in large VLDL(%) |
| rs550057 | 9 | 133271182 | chr9:1332711 | T | C | 0.2681 | 0.0562 | 1.86E-06 | XXL_VLDL_PI | 0.00368756 | -0.21092559 | Phospholipids to total lipds ratio in chylomicrons and extremely large VLDL(%) |
| rs7152677 | 14 | 31953540 | chr14:319535 | C | G | 0.2637 | 0.0548 | 1.52E-06 | L_HDL_CE_pe | 0.00371273 | 0.2352002 | Cholesterol esters to total lipids ratio in large HDL(%) |
| rs12610495 | 19 | 4717660 | chr19:471766 | A | G | -0.2534 | 0.0556 | 5.20E-06 | LDL_C | 0.00382002 | -0.22010788 | Total cholesterol in LDL(mmol/l) |
| rs12610495 | 19 | 4717660 | chr19:471766 | A | G | -0.2534 | 0.0556 | 5.20E-06 | FAw6 | 0.00395104 | -0.22086414 | Omega_6 fatty acids(mmol/l) |
| rs11627388 | 14 | 31953140 | chr14:319531 | A | C | -0.2482 | 0.0533 | 3.19E-06 | L_HDL_C_per | 0.00400307 | -0.21889842 | Total cholesterol to total lipids ratio in large HDL(%) |
| rs1455664 | 4 | 179425171 | chr4:1794251 | T | C | -0.3158 | 0.0714 | 9.77E-06 | XL_VLDL_FC | 0.00405327 | -0.32620829 | Free cholesterol in very large VLDL(mmol/l) |
| rs12610495 | 19 | 4717660 | chr19:471766 | A | G | -0.2534 | 0.0556 | 5.20E-06 | XS_VLDL_PL | 0.00407691 | -0.21804983 | Phospholipids in very small VLDL(mmol/l) |
| rs3774641 | 3 | 45896341 | chr3:4589634 | T | G | 0.2741 | 0.0605 | 5.95E-06 | XL_HDL_CE | 0.00410549 | -0.28664436 | Cholesterol esters in very large HDL(mmol/l) |
| rs7152677 | 14 | 31953540 | chr14:319535 | C | G | 0.2637 | 0.0548 | 1.52E-06 | XS_VLDL_P | 0.00413794 | -0.23692959 | Concentration of very small VLDL particles(mol/l) |
| rs1455662 | 4 | 179423719 | chr4:1794237 | A | G | -0.3156 | 0.0713 | 9.60E-06 | XL_VLDL_FC | 0.00413795 | -0.32522105 | Free cholesterol in very large VLDL(mmol/l) |
| rs2302929 | 2 | 15595524 | chr2:1559552 | T | C | 0.2976 | 0.0672 | 9.54E-06 | M_HDL_C | 0.00414425 | 0.2593767 | Total cholesterol in medium HDL(mmol/l) |
| rs74586549 | 3 | 45884551 | chr3:4588455 | T | C | 0.3443 | 0.0768 | 7.34E-06 | XL_HDL_FC | 0.00427982 | -0.34370029 | Free cholesterol in very large HDL(mmol/l) |
| rs2191031 | 3 | 45869378 | chr3:4586937 | A | G | 0.2758 | 0.0613 | 6.80E-06 | XL_HDL_C | 0.00432702 | -0.28710737 | Total cholesterol in very large HDL(mmol/l) |
| rs1455663 | 4 | 179425076 | chr4:1794250 | A | G | -0.3156 | 0.0714 | 9.87E-06 | XL_VLDL_FC | 0.00443012 | -0.32422229 | Free cholesterol in very large VLDL(mmol/l) |
| rs10091098 | 8 | 121836590 | chr8:1218365 | A | G | -0.233 | 0.0508 | 4.55E-06 | S_HDL_P | 0.00445851 | 0.20346138 | Concentration of small HDL particles(mol/l) |
| rs10087789 | 8 | 121836324 | chr8:1218363 | C | G | 0.2327 | 0.0518 | 7.00E-06 | S_HDL_P | 0.00446244 | -0.2034432 | Concentration of small HDL particles(mol/l) |
| rs7152677 | 14 | 31953540 | chr14:319535 | C | G | 0.2637 | 0.0548 | 1.52E-06 | XS_VLDL_PL | 0.00451221 | -0.23206481 | Phospholipids in very small VLDL(mmol/l) |
| rs7152677 | 14 | 31953540 | chr14:319535 | C | G | 0.2637 | 0.0548 | 1.52E-06 | XS_VLDL_L | 0.00452452 | -0.2356465 | Total lipids in very small VLDL(mmol/l) |
| rs7152677 | 14 | 31953540 | chr14:319535 | C | G | 0.2637 | 0.0548 | 1.52E-06 | L_HDL_C_per | 0.00455382 | 0.22507636 | Total cholesterol to total lipids ratio in large HDL(%) |
| rs647800 | 9 | 133272585 | chr9:1332725 | A | G | -0.2683 | 0.052 | 2.41E-07 | L_VLDL_PL_p | 0.0045607 | 0.20301486 | Phospholipids to total lipds ratio in large VLDL(%) |
| rs4797120 | 18 | 3725189 | chr18:372518 | C | G | -0.2956 | 0.0621 | 1.91E-06 | IDL_TG | 0.00457544 | -0.22349762 | Triglycerides in IDL(mmol/l) |
| rs10091098 | 8 | 121836590 | chr8:1218365 | A | G | -0.233 | 0.0508 | 4.55E-06 | S_HDL_L | 0.00464653 | 0.20252774 | Total lipids in small HDL(mmol/l) |
| rs10087789 | 8 | 121836324 | chr8:1218363 | C | G | 0.2327 | 0.0518 | 7.00E-06 | S_HDL_L | 0.00465256 | -0.20250009 | Total lipids in small HDL(mmol/l) |
| rs7152677 | 14 | 31953540 | chr14:319535 | C | G | 0.2637 | 0.0548 | 1.52E-06 | ApoB_ApoA1 | 0.00466119 | -0.23726672 | Ratio of apolipoprotein B to apolipoprotein A_I |
| rs7152677 | 14 | 31953540 | chr14:319535 | C | G | 0.2637 | 0.0548 | 1.52E-06 | ApoB | 0.00470032 | -0.23714693 | Apolipoprotein B(g/l) |
| rs12610495 | 19 | 4717660 | chr19:471766 | A | G | -0.2534 | 0.0556 | 5.20E-06 | L_LDL_PL | 0.004703 | -0.21339408 | Phospholipids in large LDL(mmol/l) |
| rs12610495 | 19 | 4717660 | chr19:471766 | A | G | -0.2534 | 0.0556 | 5.20E-06 | PUFA | 0.00495539 | -0.21416154 | Polyunsaturated fatty acids(mmol/l) |
| rs12610495 | 19 | 4717660 | chr19:471766 | A | G | -0.2534 | 0.0556 | 5.20E-06 | L_LDL_FC | 0.0051545 | -0.21569757 | Free cholesterol in large LDL(mmol/l) |
| rs12610495 | 19 | 4717660 | chr19:471766 | A | G | -0.2534 | 0.0556 | 5.20E-06 | S_LDL_CE | 0.00517861 | -0.22112605 | Cholesterol esters in small LDL(mmol/l) |
| rs10091098 | 8 | 121836590 | chr8:1218365 | A | G | -0.233 | 0.0508 | 4.55E-06 | XL_VLDL_PL_ | 0.00527577 | -0.20823162 | Phospholipids to total lipds ratio in very large VLDL(%) |
| rs10087789 | 8 | 121836324 | chr8:1218363 | C | G | 0.2327 | 0.0518 | 7.00E-06 | XL_VLDL_PL_ | 0.00527733 | 0.20822624 | Phospholipids to total lipds ratio in very large VLDL(%) |
| rs7152677 | 14 | 31953540 | chr14:319535 | C | G | 0.2637 | 0.0548 | 1.52E-06 | M_LDL_PL | 0.00537968 | -0.22692881 | Phospholipids in medium LDL(mmol/l) |

| | | | | | | | | | | | | |
|---|---|---|---|---|---|---|---|---|---|---|---|---|
| rs12610495 | 19 | 4717660 | chr19:471766 | A | G | -0.2534 | 0.0556 | 5.20E-06 | M_LDL_FC | 0.00543386 | -0.2155532 | Free cholesterol in medium LDL(mmol/l) |
| rs4389947 | 8 | 121816361 | chr8:1218163 | A | G | -0.2364 | 0.0517 | 4.91E-06 | XL_VLDL_PL_ | 0.00546509 | -0.20703819 | Phospholipids to total lipds ratio in very large VLDL(%) |
| rs3770462 | 2 | 15609200 | chr2:1560920 | T | G | 0.3409 | 0.0757 | 6.74E-06 | M_HDL_CE | 0.00547839 | 0.26023939 | Cholesterol esters in medium HDL(mmol/l) |
| rs4668941 | 2 | 15610007 | chr2:1561000 | T | G | 0.3359 | 0.0754 | 8.51E-06 | M_HDL_CE | 0.00547839 | 0.26023939 | Cholesterol esters in medium HDL(mmol/l) |
| rs41264195 | 2 | 15606096 | chr2:1560609 | T | C | -0.3359 | 0.0754 | 8.49E-06 | M_HDL_CE | 0.00547989 | -0.26025835 | Cholesterol esters in medium HDL(mmol/l) |
| rs79808866 | 2 | 15604860 | chr2:1560486 | T | G | 0.3357 | 0.0754 | 8.59E-06 | M_HDL_CE | 0.00547989 | 0.26025835 | Cholesterol esters in medium HDL(mmol/l) |
| rs10088118 | 8 | 121806381 | chr8:1218063 | C | G | -0.2329 | 0.0517 | 6.79E-06 | XL_VLDL_PL_ | 0.00555289 | -0.20617958 | Phospholipids to total lipds ratio in very large VLDL(%) |
| rs647800 | 9 | 133272585 | chr9:1332725 | A | G | -0.2683 | 0.052 | 2.41E-07 | L_VLDL_CE_p | 0.00579634 | -0.20783708 | Cholesterol esters to total lipids ratio in large VLDL(%) |
| rs62120724 | 2 | 15593845 | chr2:1559384 | A | G | -0.2986 | 0.0675 | 9.73E-06 | M_HDL_CE | 0.00591027 | -0.25299782 | Cholesterol esters in medium HDL(mmol/l) |
| rs2032778 | 2 | 15596573 | chr2:1559657 | A | G | -0.299 | 0.0675 | 9.48E-06 | M_HDL_CE | 0.00591027 | -0.25299782 | Cholesterol esters in medium HDL(mmol/l) |
| rs9739464 | 12 | 62037992 | chr12:620379 | A | T | 0.2846 | 0.06 | 2.10E-06 | Ala | 0.0061787 | 0.25694382 | Alanine(mmol/l) |
| rs10506436 | 12 | 62028786 | chr12:620287 | A | T | -0.2811 | 0.06 | 2.84E-06 | Ala | 0.00628487 | -0.25643313 | Alanine(mmol/l) |
| rs10784279 | 12 | 62044016 | chr12:620440 | T | C | -0.2778 | 0.0599 | 3.50E-06 | Ala | 0.00628505 | -0.25590038 | Alanine(mmol/l) |
| rs3774641 | 3 | 45896341 | chr3:4589634 | T | G | 0.2741 | 0.0605 | 5.95E-06 | XL_HDL_C | 0.00629746 | -0.27239671 | Total cholesterol in very large HDL(mmol/l) |
| rs10784278 | 12 | 62042165 | chr12:620421 | A | T | -0.2836 | 0.06 | 2.32E-06 | Ala | 0.00630497 | -0.25589584 | Alanine(mmol/l) |
| rs12610495 | 19 | 4717660 | chr19:471766 | A | G | -0.2534 | 0.0556 | 5.20E-06 | HDL3_C | 0.00631842 | -0.21514916 | Total cholesterol in HDL3(mmol/l) |
| rs12610495 | 19 | 4717660 | chr19:471766 | A | G | -0.2534 | 0.0556 | 5.20E-06 | S_LDL_PL | 0.00633543 | -0.21029361 | Phospholipids in small LDL(mmol/l) |
| rs74586549 | 3 | 45884551 | chr3:4588455 | T | C | 0.3443 | 0.0768 | 7.34E-06 | XL_HDL_C | 0.00635431 | -0.33444665 | Total cholesterol in very large HDL(mmol/l) |
| rs7152677 | 14 | 31953540 | chr14:319535 | C | G | 0.2637 | 0.0548 | 1.52E-06 | Cit | 0.00636988 | -0.24993016 | Citrate(mmol/l) |
| rs12610495 | 19 | 4717660 | chr19:471766 | A | G | -0.2534 | 0.0556 | 5.20E-06 | M_LDL_CE | 0.00643303 | -0.21476372 | Cholesterol esters in medium LDL(mmol/l) |
| rs12610495 | 19 | 4717660 | chr19:471766 | A | G | -0.2534 | 0.0556 | 5.20E-06 | Remnant_C | 0.00658158 | -0.21427864 | Remnant cholesterol (non_HDL, non_LDL _cholesterol)(mmol/l) |
| rs74586549 | 3 | 45884551 | chr3:4588455 | T | C | 0.3443 | 0.0768 | 7.34E-06 | XL_HDL_CE | 0.0067847 | -0.33257391 | Cholesterol esters in very large HDL(mmol/l) |
| rs1455664 | 4 | 179425171 | chr4:1794251 | T | C | -0.3158 | 0.0714 | 9.77E-06 | XL_VLDL_L | 0.0067881 | -0.30367894 | Total lipids in very large VLDL(mmol/l) |
| rs11627388 | 14 | 31953140 | chr14:319531 | A | C | -0.2482 | 0.0533 | 3.19E-06 | LA | 0.0068049 | 0.21862997 | 18:2, linoleic acid(mmol/l) |
| rs4797120 | 18 | 3725189 | chr18:372518 | C | G | -0.2956 | 0.0621 | 1.91E-06 | XS_VLDL_TG | 0.00686448 | -0.23190156 | Triglycerides in very small VLDL(mmol/l) |
| rs1455662 | 4 | 179423719 | chr4:1794237 | A | G | -0.3156 | 0.0713 | 9.60E-06 | XL_VLDL_L | 0.00694287 | -0.30261086 | Total lipids in very large VLDL(mmol/l) |
| rs62122279 | 2 | 15621842 | chr2:1562184 | T | C | 0.3047 | 0.0678 | 6.91E-06 | M_HDL_CE | 0.00707673 | 0.24707476 | Cholesterol esters in medium HDL(mmol/l) |
| rs4668946 | 2 | 15624494 | chr2:1562449 | A | G | 0.3109 | 0.0679 | 4.72E-06 | M_HDL_CE | 0.00709577 | 0.24695856 | Cholesterol esters in medium HDL(mmol/l) |
| rs12614308 | 2 | 15624914 | chr2:1562491 | A | G | -0.3109 | 0.0679 | 4.72E-06 | M_HDL_CE | 0.00709577 | -0.24695856 | Cholesterol esters in medium HDL(mmol/l) |
| rs13263570 | 8 | 121845942 | chr8:1218459 | A | G | -0.2298 | 0.0516 | 8.50E-06 | S_HDL_L | 0.00709808 | 0.19013198 | Total lipids in small HDL(mmol/l) |
| rs10784279 | 12 | 62044016 | chr12:620440 | T | C | -0.2778 | 0.0599 | 3.50E-06 | Tyr | 0.00713545 | -0.235132 | Tyrosine(mmol/l) |
| rs7152677 | 14 | 31953540 | chr14:319535 | C | G | 0.2637 | 0.0548 | 1.52E-06 | S_VLDL_C | 0.0071646 | -0.22870734 | Total cholesterol in small VLDL(mmol/l) |
| rs10506436 | 12 | 62028786 | chr12:620287 | A | T | -0.2811 | 0.06 | 2.84E-06 | Tyr | 0.00717255 | -0.23547082 | Tyrosine(mmol/l) |
| rs7152677 | 14 | 31953540 | chr14:319535 | C | G | 0.2637 | 0.0548 | 1.52E-06 | LA | 0.00722191 | -0.22635074 | 18:2, linoleic acid(mmol/l) |
| rs10784278 | 12 | 62042165 | chr12:620421 | A | T | -0.2836 | 0.06 | 2.32E-06 | Tyr | 0.00722814 | -0.23484399 | Tyrosine(mmol/l) |
| rs74586549 | 3 | 45884551 | chr3:4588455 | T | C | 0.3443 | 0.0768 | 7.34E-06 | MUFA_FA_ra | 0.00727254 | 0.3274299 | Ratio of monounsaturated fatty acids to total fatty acids(percent) |
| rs13263570 | 8 | 121845942 | chr8:1218459 | A | G | -0.2298 | 0.0516 | 8.50E-06 | S_HDL_P | 0.00728441 | 0.18952921 | Concentration of small HDL particles(mol/l) |
| rs2302929 | 2 | 15595524 | chr2:1559552 | T | C | 0.2976 | 0.0672 | 9.54E-06 | M_HDL_P | 0.00729757 | 0.23452437 | Concentration of medium HDL particles(mol/l) |
| rs13263570 | 8 | 121845942 | chr8:1218459 | A | G | -0.2298 | 0.0516 | 8.50E-06 | XL_VLDL_PL_ | 0.00737033 | -0.19739989 | Phospholipids to total lipds ratio in very large VLDL(%) |
| rs1455663 | 4 | 179425076 | chr4:1794250 | A | G | -0.3156 | 0.0714 | 9.87E-06 | XL_HDL_TG | 0.00738377 | -0.32583591 | Triglycerides in very large HDL(mmol/l) |
| rs1455663 | 4 | 179425076 | chr4:1794250 | A | G | -0.3156 | 0.0714 | 9.87E-06 | XL_VLDL_L | 0.00741469 | -0.30150451 | Total lipids in very large VLDL(mmol/l) |
| rs633862 | 9 | 133279871 | chr9:1332798 | T | C | 0.2534 | 0.0515 | 8.79E-07 | L_VLDL_PL_p | 0.00745929 | -0.18647738 | Phospholipids to total lipds ratio in large VLDL(%) |
| rs9739464 | 12 | 62037992 | chr12:620379 | A | T | 0.2846 | 0.06 | 2.10E-06 | Tyr | 0.00745941 | 0.23432593 | Tyrosine(mmol/l) |
| rs7152677 | 14 | 31953540 | chr14:319535 | C | G | 0.2637 | 0.0548 | 1.52E-06 | S_LDL_PL | 0.00750762 | -0.2216885 | Phospholipids in small LDL(mmol/l) |
| rs13063033 | 3 | 46040989 | chr3:4604098 | A | G | 0.3555 | 0.0772 | 4.12E-06 | Crea | 0.00751903 | 0.29539179 | Creatinine(mmol/l) |
| rs2302929 | 2 | 15595524 | chr2:1559552 | T | C | 0.2976 | 0.0672 | 9.54E-06 | M_HDL_L | 0.00757866 | 0.23398538 | Total lipids in medium HDL(mmol/l) |
| rs647800 | 9 | 133272585 | chr9:1332725 | A | G | -0.2683 | 0.052 | 2.41E-07 | XXL_VLDL_PI | 0.00784915 | 0.18699641 | Phospholipids to total lipds ratio in chylomicrons and extremely large VLDL(%) |
| rs1455664 | 4 | 179425171 | chr4:1794251 | T | C | -0.3158 | 0.0714 | 9.77E-06 | XL_VLDL_P | 0.00795017 | -0.29772538 | Concentration of very large VLDL particles(mol/l) |
| rs1455664 | 4 | 179425171 | chr4:1794251 | T | C | -0.3158 | 0.0714 | 9.77E-06 | XL_HDL_TG | 0.00797759 | -0.3215385 | Triglycerides in very large HDL(mmol/l) |
| rs12619874 | 2 | 15586997 | chr2:1558699 | C | G | -0.3204 | 0.0665 | 1.46E-06 | M_HDL_CE | 0.00801452 | -0.23424983 | Cholesterol esters in medium HDL(mmol/l) |
| rs12610495 | 19 | 4717660 | chr19:471766 | A | G | -0.2534 | 0.0556 | 5.20E-06 | M_LDL_PL | 0.00806456 | -0.20082141 | Phospholipids in medium LDL(mmol/l) |
| rs1455662 | 4 | 179423719 | chr4:1794237 | A | G | -0.3156 | 0.0713 | 9.60E-06 | XL_HDL_TG | 0.00812655 | -0.32053811 | Triglycerides in very large HDL(mmol/l) |
| rs1455662 | 4 | 179423719 | chr4:1794237 | A | G | -0.3156 | 0.0713 | 9.60E-06 | XL_VLDL_P | 0.00813033 | -0.29665312 | Concentration of very large VLDL particles(mol/l) |
| rs76860840 | 2 | 15618674 | chr2:1561867 | C | G | 0.3536 | 0.0764 | 3.66E-06 | M_HDL_CE | 0.00819282 | 0.24933358 | Cholesterol esters in medium HDL(mmol/l) |
| rs807628 | 2 | 15638176 | chr2:1563817 | A | G | 0.3095 | 0.0678 | 5.04E-06 | M_HDL_CE | 0.00821799 | 0.2417959 | Cholesterol esters in medium HDL(mmol/l) |
| rs807627 | 2 | 15639565 | chr2:1563956 | T | C | 0.3105 | 0.0679 | 4.79E-06 | M_HDL_CE | 0.00821857 | 0.24175783 | Cholesterol esters in medium HDL(mmol/l) |
| rs2191031 | 3 | 45869378 | chr3:4586937 | A | G | 0.2758 | 0.0613 | 6.80E-06 | XL_HDL_PL | 0.00833889 | -0.24773733 | Phospholipids in very large HDL(mmol/l) |
| rs11627388 | 14 | 31953140 | chr14:319531 | A | C | -0.2482 | 0.0533 | 3.19E-06 | S_VLDL_C | 0.0083431 | 0.21517358 | Total cholesterol in small VLDL(mmol/l) |
| rs7152677 | 14 | 31953540 | chr14:319535 | C | G | 0.2637 | 0.0548 | 1.52E-06 | IDL_TG | 0.00838304 | -0.19894533 | Triglycerides in IDL(mmol/l) |
| rs7152677 | 14 | 31953540 | chr14:319535 | C | G | 0.2637 | 0.0548 | 1.52E-06 | M_LDL_P | 0.00839515 | -0.21556545 | Concentration of medium LDL particles(mol/l) |
| rs4797120 | 18 | 3725189 | chr18:372518 | C | G | -0.2956 | 0.0621 | 1.91E-06 | L_HDL_CE_pe | 0.00853905 | 0.2231054 | Cholesterol esters to total lipids ratio in large HDL(%) |
| rs4797120 | 18 | 3725189 | chr18:372518 | C | G | -0.2956 | 0.0621 | 1.91E-06 | M_LDL_TG | 0.00858505 | -0.19993147 | Triglycerides in medium LDL(mmol/l) |
| rs7152677 | 14 | 31953540 | chr14:319535 | C | G | 0.2637 | 0.0548 | 1.52E-06 | S_LDL_P | 0.00860105 | -0.21708085 | Concentration of small LDL particles(mol/l) |
| rs1455663 | 4 | 179425076 | chr4:1794250 | A | G | -0.3156 | 0.0714 | 9.87E-06 | XL_VLDL_P | 0.00867598 | -0.29550911 | Concentration of very large VLDL particles(mol/l) |
| rs7152677 | 14 | 31953540 | chr14:319535 | C | G | 0.2637 | 0.0548 | 1.52E-06 | M_LDL_L | 0.00868138 | -0.21570623 | Total lipids in medium LDL(mmol/l) |
| rs7152677 | 14 | 31953540 | chr14:319535 | C | G | 0.2637 | 0.0548 | 1.52E-06 | S_VLDL_FC | 0.00868142 | -0.22382293 | Free cholesterol in small VLDL(mmol/l) |
| rs62120724 | 2 | 15593845 | chr2:1559384 | A | G | -0.2986 | 0.0675 | 9.73E-06 | M_HDL_C | 0.00878923 | -0.23896936 | Total cholesterol in medium HDL(mmol/l) |
| rs2032778 | 2 | 15596573 | chr2:1559657 | A | G | -0.299 | 0.0675 | 9.48E-06 | M_HDL_C | 0.00878923 | -0.23896936 | Total cholesterol in medium HDL(mmol/l) |
| rs7152677 | 14 | 31953540 | chr14:319535 | C | G | 0.2637 | 0.0548 | 1.52E-06 | M_LDL_FC | 0.00903186 | -0.21800453 | Free cholesterol in medium LDL(mmol/l) |
| rs3770462 | 2 | 15609200 | chr2:1560920 | T | G | 0.3409 | 0.0757 | 6.74E-06 | M_HDL_C | 0.0091295 | 0.24245005 | Total cholesterol in medium HDL(mmol/l) |
| rs4668941 | 2 | 15610007 | chr2:1561000 | T | G | 0.3359 | 0.0754 | 8.51E-06 | M_HDL_C | 0.0091295 | 0.24245005 | Total cholesterol in medium HDL(mmol/l) |
| rs79808866 | 2 | 15604860 | chr2:1560486 | T | G | 0.3357 | 0.0754 | 8.59E-06 | M_HDL_C | 0.00913272 | 0.24246427 | Total cholesterol in medium HDL(mmol/l) |
| rs41264195 | 2 | 15606096 | chr2:1560609 | T | C | -0.3359 | 0.0754 | 8.49E-06 | M_HDL_C | 0.00913272 | -0.24246427 | Total cholesterol in medium HDL(mmol/l) |
| rs7152677 | 14 | 31953540 | chr14:319535 | C | G | 0.2637 | 0.0548 | 1.52E-06 | S_LDL_L | 0.0092357 | -0.21630426 | Total lipids in small LDL(mmol/l) |
| rs11174294 | 12 | 62072129 | chr12:620721 | T | C | -0.29 | 0.0604 | 1.59E-06 | Ala | 0.009297 | -0.23681782 | Alanine(mmol/l) |
| rs12610495 | 19 | 4717660 | chr19:471766 | A | G | -0.2534 | 0.0556 | 5.20E-06 | S_LDL_FC_pe | 0.00932556 | 0.18935203 | Free cholesterol to total lipids ratio in small LDL(%) |
| rs10747951 | 12 | 62075677 | chr12:620756 | T | C | -0.29 | 0.0604 | 1.58E-06 | Ala | 0.0093424 | -0.23667244 | Alanine(mmol/l) |
| rs10506432 | 12 | 62077116 | chr12:620771 | A | C | 0.29 | 0.0604 | 1.58E-06 | Ala | 0.00934759 | 0.23667462 | Alanine(mmol/l) |
| rs11174296 | 12 | 62077677 | chr12:620776 | C | G | 0.288 | 0.0602 | 1.75E-06 | Ala | 0.00934759 | 0.23667462 | Alanine(mmol/l) |
| rs9706516 | 12 | 62074403 | chr12:620744 | A | C | -0.2923 | 0.0604 | 1.30E-06 | Ala | 0.00934994 | -0.23663202 | Alanine(mmol/l) |
| rs10877787 | 12 | 62072981 | chr12:620729 | A | G | 0.29 | 0.0604 | 1.58E-06 | Ala | 0.0093578 | 0.23658954 | Alanine(mmol/l) |
| rs13381043 | 18 | 3724602 | chr18:372460 | A | G | -0.325 | 0.0613 | 1.13E-07 | Pyr | 0.00935894 | -0.23212282 | Pyruvate(mmol/l) |
| rs9738190 | 12 | 62078790 | chr12:620787 | A | G | -0.2898 | 0.0602 | 1.51E-06 | Ala | 0.00939768 | -0.23655116 | Alanine(mmol/l) |
| rs7152677 | 14 | 31953540 | chr14:319535 | C | G | 0.2637 | 0.0548 | 1.52E-06 | S_VLDL_CE | 0.00940354 | -0.22087514 | Cholesterol esters in small VLDL(mmol/l) |
| rs12610495 | 19 | 4717660 | chr19:471766 | A | G | -0.2534 | 0.0556 | 5.20E-06 | LA | 0.00940803 | -0.20338609 | 18:2, linoleic acid(mmol/l) |
| rs12610495 | 19 | 4717660 | chr19:471766 | A | G | -0.2534 | 0.0556 | 5.20E-06 | M_LDL_TG | 0.00945663 | -0.17550652 | Triglycerides in medium LDL(mmol/l) |
| rs514659 | 9 | 133266790 | chr9:1332667 | A | C | -0.2811 | 0.0526 | 9.08E-08 | Cit | 0.00945707 | 0.20350258 | Citrate(mmol/l) |
| rs687621 | 9 | 133261662 | chr9:1332616 | A | G | -0.2837 | 0.0528 | 7.63E-08 | Cit | 0.00945707 | 0.20350258 | Citrate(mmol/l) |
| rs687289 | 9 | 133261703 | chr9:1332617 | A | G | 0.2826 | 0.0527 | 8.14E-08 | Cit | 0.00945707 | -0.20350258 | Citrate(mmol/l) |
| rs545971 | 9 | 133267960 | chr9:1332679 | T | C | 0.2794 | 0.0524 | 9.58E-08 | Cit | 0.00945707 | -0.20350258 | Citrate(mmol/l) |
| rs612169 | 9 | 133268030 | chr9:1332680 | A | G | -0.2797 | 0.0527 | 1.08E-07 | Cit | 0.00945707 | 0.20350258 | Citrate(mmol/l) |
| rs597988 | 9 | 133268872 | chr9:1332688 | A | T | 0.2796 | 0.0526 | 1.07E-07 | Cit | 0.00945707 | -0.20350258 | Citrate(mmol/l) |
| rs597974 | 9 | 133268885 | chr9:1332688 | A | G | -0.2796 | 0.0526 | 1.07E-07 | Cit | 0.00945707 | 0.20350258 | Citrate(mmol/l) |
| rs8176663 | 9 | 133269015 | chr9:1332690 | T | C | -0.2798 | 0.0526 | 1.05E-07 | Cit | 0.00945707 | 0.20350258 | Citrate(mmol/l) |
| rs491626 | 9 | 133269461 | chr9:1332694 | T | C | 0.2797 | 0.0526 | 1.05E-07 | Cit | 0.00945707 | -0.20350258 | Citrate(mmol/l) |
| rs492488 | 9 | 133269548 | chr9:1332695 | A | G | 0.2798 | 0.0526 | 1.02E-07 | Cit | 0.00945707 | -0.20350258 | Citrate(mmol/l) |
| rs493246 | 9 | 133269582 | chr9:1332695 | A | G | 0.2799 | 0.0526 | 1.01E-07 | Cit | 0.00945707 | -0.20350258 | Citrate(mmol/l) |
| rs495203 | 9 | 133269828 | chr9:1332698 | T | C | 0.2816 | 0.0526 | 8.52E-08 | Cit | 0.00945707 | -0.20350258 | Citrate(mmol/l) |
| rs582118 | 9 | 133270061 | chr9:1332700 | A | G | -0.2798 | 0.0526 | 1.02E-07 | Cit | 0.00945707 | 0.20350258 | Citrate(mmol/l) |
| rs582094 | 9 | 133270074 | chr9:1332700 | A | T | -0.2796 | 0.0526 | 1.04E-07 | Cit | 0.00945707 | 0.20350258 | Citrate(mmol/l) |
| rs2769071 | 9 | 133270565 | chr9:1332705 | A | G | -0.2785 | 0.0526 | 1.17E-07 | Cit | 0.00945707 | 0.20350258 | Citrate(mmol/l) |
| rs677355 | 9 | 133270615 | chr9:1332706 | A | G | 0.2789 | 0.0526 | 1.12E-07 | Cit | 0.00945707 | -0.20350258 | Citrate(mmol/l) |
| rs676996 | 9 | 133270647 | chr9:1332706 | T | G | -0.2785 | 0.0526 | 1.17E-07 | Cit | 0.00945707 | 0.20350258 | Citrate(mmol/l) |
| rs676457 | 9 | 133270797 | chr9:1332707 | A | T | -0.2783 | 0.0526 | 1.21E-07 | Cit | 0.00945707 | 0.20350258 | Citrate(mmol/l) |
| rs674302 | 9 | 133271249 | chr9:1332712 | A | T | 0.278 | 0.0526 | 1.26E-07 | Cit | 0.00945707 | -0.20350258 | Citrate(mmol/l) |
| rs554833 | 9 | 133271745 | chr9:1332717 | T | C | 0.2788 | 0.0526 | 1.13E-07 | Cit | 0.00945707 | -0.20350258 | Citrate(mmol/l) |
| rs505922 | 9 | 133273813 | chr9:1332738 | T | C | -0.2687 | 0.0526 | 3.21E-07 | Cit | 0.00945707 | 0.20350258 | Citrate(mmol/l) |
| rs529565 | 9 | 133274084 | chr9:1332740 | T | C | -0.2717 | 0.0526 | 2.44E-07 | Cit | 0.0094942 | 0.20341372 | Citrate(mmol/l) |
| rs807576 | 2 | 15664332 | chr2:1566433 | T | C | 0.3185 | 0.0704 | 6.15E-06 | XL_VLDL_TG_ | 0.00950893 | 0.22953423 | Triglycerides to total lipids ratio in very large VLDL(%) |
| rs807577 | 2 | 15664270 | chr2:1566427 | C | G | 0.3181 | 0.0704 | 6.28E-06 | XL_VLDL_TG_ | 0.00952488 | 0.22951499 | Triglycerides to total lipids ratio in very large VLDL(%) |
| rs12610495 | 19 | 4717660 | chr19:471766 | A | G | -0.2534 | 0.0556 | 5.20E-06 | S_LDL_CE_pe | 0.00952772 | -0.21125299 | Cholesterol esters to total lipids ratio in small LDL(%) |
| rs807587 | 2 | 15659856 | chr2:1565985 | C | G | 0.3246 | 0.0702 | 3.76E-06 | XL_VLDL_TG_ | 0.00963525 | 0.22931425 | Triglycerides to total lipids ratio in very large VLDL(%) |
| rs807586 | 2 | 15660004 | chr2:1566000 | T | C | 0.3125 | 0.0703 | 8.76E-06 | XL_VLDL_TG_ | 0.00963525 | 0.22931425 | Triglycerides to total lipids ratio in very large VLDL(%) |
| rs807583 | 2 | 15660597 | chr2:1566059 | A | G | -0.3121 | 0.0703 | 9.01E-06 | XL_VLDL_TG_ | 0.00963525 | -0.22931425 | Triglycerides to total lipids ratio in very large VLDL(%) |
| rs807579 | 2 | 15663460 | chr2:1566346 | T | C | 0.3124 | 0.0703 | 8.88E-06 | XL_VLDL_TG_ | 0.00963525 | 0.22931425 | Triglycerides to total lipids ratio in very large VLDL(%) |
| rs807578 | 2 | 15663516 | chr2:1566351 | C | G | -0.3124 | 0.0703 | 8.81E-06 | XL_VLDL_TG_ | 0.00963525 | -0.22931425 | Triglycerides to total lipids ratio in very large VLDL(%) |
| rs807589 | 2 | 15658661 | chr2:1565866 | A | C | -0.326 | 0.0702 | 3.48E-06 | XL_VLDL_TG_ | 0.00968067 | -0.22934124 | Triglycerides to total lipids ratio in very large VLDL(%) |
| rs7152677 | 14 | 31953540 | chr14:319535 | C | G | 0.2637 | 0.0548 | 1.52E-06 | Remnant_C | 0.00972547 | -0.2195319 | Remnant cholesterol (non_HDL, non_LDL _cholesterol)(mmol/l) |
| rs1455663 | 4 | 179425076 | chr4:1794250 | A | G | -0.3156 | 0.0714 | 9.87E-06 | S_LDL_C_per | 0.00981951 | 0.3228713 | Total cholesterol to total lipids ratio in small LDL(%) |
| rs500498 | 9 | 133273232 | chr9:1332732 | T | C | -0.2535 | 0.0516 | 9.06E-07 | L_VLDL_CE_p | 0.00982675 | -0.19301329 | Cholesterol esters to total lipids ratio in large VLDL(%) |
| rs500499 | 9 | 133273233 | chr9:1332732 | C | G | 0.2535 | 0.0516 | 9.06E-07 | L_VLDL_CE_p | 0.00982675 | 0.19301329 | Cholesterol esters to total lipids ratio in large VLDL(%) |
| rs660340 | 9 | 133272138 | chr9:1332721 | A | G | -0.255 | 0.0517 | 7.97E-07 | L_VLDL_CE_p | 0.00984043 | -0.1929876 | Cholesterol esters to total lipids ratio in large VLDL(%) |
| rs581107 | 9 | 133272287 | chr9:1332722 | T | C | 0.2553 | 0.0517 | 7.87E-07 | L_VLDL_CE_p | 0.00984043 | 0.1929876 | Cholesterol esters to total lipids ratio in large VLDL(%) |
| rs659104 | 9 | 133272408 | chr9:1332724 | T | G | -0.255 | 0.0517 | 7.97E-07 | L_VLDL_CE_p | 0.00984043 | -0.1929876 | Cholesterol esters to total lipids ratio in large VLDL(%) |
| rs473533 | 9 | 133272620 | chr9:1332726 | T | C | -0.2551 | 0.0517 | 7.89E-07 | L_VLDL_CE_p | 0.00984043 | -0.1929876 | Cholesterol esters to total lipids ratio in large VLDL(%) |
| rs475419 | 9 | 133272816 | chr9:1332728 | T | C | 0.2576 | 0.0517 | 6.32E-07 | L_VLDL_CE_p | 0.00984043 | 0.1929876 | Cholesterol esters to total lipids ratio in large VLDL(%) |
| rs476410 | 9 | 133272953 | chr9:1332729 | C | G | 0.2538 | 0.0517 | 8.96E-07 | L_VLDL_CE_p | 0.00984043 | 0.1929876 | Cholesterol esters to total lipids ratio in large VLDL(%) |
| rs645982 | 9 | 133272994 | chr9:1332729 | A | G | -0.2536 | 0.0516 | 9.02E-07 | L_VLDL_CE_p | 0.00984043 | -0.1929876 | Cholesterol esters to total lipids ratio in large VLDL(%) |
| rs11174294 | 12 | 62072129 | chr12:620721 | T | C | -0.29 | 0.0604 | 1.59E-06 | Gln | 0.00990609 | -0.1980978 | Glutamine(mmol/l) |

* GWAS loci were selected from The Severe Covid-19 GWAS Group N Engl J Med (2020), doi:10.1056/NEJMoa2020283, all SNP with assoication p value <1e-5 have been tested.

** all metalite QTLs(P value <0.01) have been listed

Table S8 metabolites QTLs of COVID-19 GWAS loci from the 500 FG cohort study

| SNP* | chr | pos | effect_allele | alternative_ | beta-GWAS | standard_err | p_value-GWAS | metabolite_id | p_value-QTL** | beta-QTL | annotation |
|---|---|---|---|---|---|---|---|---|---|---|---|
| rs2109069 | 19 | 4719443 | A | G | 0.3056227 | 0.0440513 | 3.97936E-12 | FreeC | 0.000919209 | 0.2424425 | Free cholesterol(mmol/l) |
| rs2323476 | 13 | 37966084 | A | G | -0.1870708 | 0.0422955 | 9.73636E-06 | M_VLDL_TG_percent | 0.000930628 | -0.22839517 | Triglycerides to total lipids ratio in medium VLDL(%) |
| rs7958379 | 12 | 113363284 | A | G | -0.2213876 | 0.04553 | 1.15938E-06 | L_LDL_TG_percent | 0.001004478 | 0.24059418 | Triglycerides to total lipids ratio in large LDL(%) |
| rs4076440 | 1 | 9690476 | C | T | 0.301866 | 0.0682837 | 9.83419E-06 | S_VLDL_FC_percent | 0.001430507 | -0.34868294 | Free cholesterol to total lipids ratio in small VLDL(%) |
| rs12622794 | 2 | 53809057 | G | A | 0.1937065 | 0.0429963 | 6.62852E-06 | Lac | 0.001462855 | -0.24331837 | Lactate(mmol/l) |
| rs1846615 | 3 | 46032388 | G | C | 0.2347796 | 0.0439973 | 9.48679E-08 | XL_HDL_TG_percent | 0.00151714 | 0.2475966 | Triglycerides to total lipids ratio in very large HDL(%) |
| rs2109069 | 19 | 4719443 | A | G | 0.3056227 | 0.0440513 | 3.97936E-12 | L_LDL_PL_percent | 0.001537711 | -0.23553251 | Phospholipids to total lipds ratio in large LDL(%) |
| rs1873001 | 3 | 46032847 | C | T | 0.2324521 | 0.0439644 | 1.24209E-07 | XL_HDL_TG_percent | 0.001558219 | 0.24685874 | Triglycerides to total lipids ratio in very large HDL(%) |
| rs2133660 | 3 | 46031957 | C | T | 0.234661 | 0.0439958 | 9.62254E-08 | XL_HDL_TG_percent | 0.001571914 | 0.24671909 | Triglycerides to total lipids ratio in very large HDL(%) |
| rs1500003 | 3 | 46033819 | A | G | 0.2328325 | 0.0439728 | 1.19113E-07 | XL_HDL_TG_percent | 0.001612322 | 0.24614626 | Triglycerides to total lipids ratio in very large HDL(%) |
| rs2323476 | 13 | 37966084 | A | G | -0.1870708 | 0.0422955 | 9.73636E-06 | L_VLDL_C_percent | 0.001615688 | 0.22192542 | Total cholesterol to total lipids ratio in large VLDL(%) |
| rs2109069 | 19 | 4719443 | A | G | 0.3056227 | 0.0440513 | 3.97936E-12 | XS_VLDL_FC | 0.001711165 | 0.23801502 | Free cholesterol in very small VLDL(mmol/l) |
| rs2109069 | 19 | 4719443 | A | G | 0.3056227 | 0.0440513 | 3.97936E-12 | XS_VLDL_C | 0.001882851 | 0.24007795 | Total cholesterol in very small VLDL(mmol/l) |
| rs3851347 | 3 | 46053778 | G | A | 0.2605395 | 0.0443679 | 4.2992E-09 | AcAce | 0.001971567 | 0.25507103 | Acetoacetate(mmol/l) |
| rs6764042 | 3 | 46059139 | G | T | 0.2550607 | 0.0444658 | 9.68312E-09 | AcAce | 0.002035641 | 0.25555924 | Acetoacetate(mmol/l) |
| rs7623460 | 3 | 46068764 | C | A | 0.2603006 | 0.0444995 | 4.93E-09 | AcAce | 0.002090413 | 0.25504175 | Acetoacetate(mmol/l) |
| rs1392290 | 3 | 46069210 | A | G | 0.2486631 | 0.0443137 | 2.00599E-08 | AcAce | 0.002092919 | 0.25501605 | Acetoacetate(mmol/l) |
| rs6805094 | 3 | 46071463 | G | C | 0.25647 | 0.044452 | 7.94929E-09 | AcAce | 0.002105729 | 0.25488198 | Acetoacetate(mmol/l) |
| rs13060901 | 3 | 46072006 | A | G | 0.2564932 | 0.0444529 | 7.92797E-09 | AcAce | 0.002108348 | 0.25485406 | Acetoacetate(mmol/l) |
| rs2109069 | 19 | 4719443 | A | G | 0.3056227 | 0.0440513 | 3.97936E-12 | S_LDL_PL_percent | 0.002110959 | -0.23798128 | Phospholipids to total lipds ratio in small LDL(%) |
| rs2109069 | 19 | 4719443 | A | G | 0.3056227 | 0.0440513 | 3.97936E-12 | IDL_P | 0.002114773 | 0.22800191 | Concentration of IDL particles(mol/l) |
| rs4411920 | 3 | 46077525 | G | A | 0.2565551 | 0.0444575 | 7.88908E-09 | AcAce | 0.002142718 | 0.25447451 | Acetoacetate(mmol/l) |
| rs2109069 | 19 | 4719443 | A | G | 0.3056227 | 0.0440513 | 3.97936E-12 | XS_VLDL_L | 0.002203076 | 0.22950986 | Total lipids in very small VLDL(mmol/l) |
| rs2109069 | 19 | 4719443 | A | G | 0.3056227 | 0.0440513 | 3.97936E-12 | IDL_L | 0.002260816 | 0.22783453 | Total lipids in IDL(mmol/l) |
| rs1846616 | 3 | 46032441 | A | T | 0.2351907 | 0.0439068 | 8.48375E-08 | M_HDL_PL | 0.002384303 | -0.27788385 | Phospholipids in medium HDL(mmol/l) |
| rs2109069 | 19 | 4719443 | A | G | 0.3056227 | 0.0440513 | 3.97936E-12 | M_LDL_PL_percent | 0.002400217 | -0.24140762 | Phospholipids to total lipds ratio in medium LDL(%) |
| rs9849015 | 3 | 46056120 | C | T | 0.2579159 | 0.0444721 | 6.6533E-09 | AcAce | 0.002498035 | 0.25083784 | Acetoacetate(mmol/l) |
| rs2109069 | 19 | 4719443 | A | G | 0.3056227 | 0.0440513 | 3.97936E-12 | S_LDL_C | 0.002527356 | 0.23047761 | Total cholesterol in small LDL(mmol/l) |
| rs7623190 | 3 | 46068325 | C | T | 0.2556107 | 0.0444523 | 8.91342E-09 | AcAce | 0.002531997 | 0.2500626 | Acetoacetate(mmol/l) |
| rs10087754 | 8 | 122832148 | T | A | 0.1935335 | 0.0432237 | 7.55487E-06 | XL_VLDL_PL_percent | 0.00254317 | 0.22662026 | Phospholipids to total lipds ratio in very large VLDL(%) |
| rs2109069 | 19 | 4719443 | A | G | 0.3056227 | 0.0440513 | 3.97936E-12 | XS_VLDL_CE | 0.002617722 | 0.23390354 | Cholesterol esters in very small VLDL(mmol/l) |
| rs2109069 | 19 | 4719443 | A | G | 0.3056227 | 0.0440513 | 3.97936E-12 | XS_VLDL_P | 0.002710708 | 0.2238946 | Concentration of very small VLDL particles(mol/l) |
| rs2109069 | 19 | 4719443 | A | G | 0.3056227 | 0.0440513 | 3.97936E-12 | IDL_C | 0.002742562 | 0.22711211 | Total cholesterol in IDL(mmol/l) |
| rs2109069 | 19 | 4719443 | A | G | 0.3056227 | 0.0440513 | 3.97936E-12 | IDL_PL | 0.002785954 | 0.21992685 | Phospholipids in IDL(mmol/l) |
| rs4076440 | 1 | 9690476 | C | T | 0.301866 | 0.0682837 | 9.83419E-06 | S_VLDL_C_percent | 0.002799554 | -0.34845109 | Total cholesterol to total lipids ratio in small VLDL(%) |
| rs2109069 | 19 | 4719443 | A | G | 0.3056227 | 0.0440513 | 3.97936E-12 | S_LDL_L | 0.002843929 | 0.22382003 | Total lipids in small LDL(mmol/l) |
| rs4076440 | 1 | 9690476 | C | T | 0.301866 | 0.0682837 | 9.83419E-06 | IDL_FC_percent | 0.002925579 | -0.36500231 | Free cholesterol to total lipids ratio in IDL(%) |
| rs2109069 | 19 | 4719443 | A | G | 0.3056227 | 0.0440513 | 3.97936E-12 | S_LDL_P | 0.002939289 | 0.22182328 | Concentration of small LDL particles(mol/l) |
| rs2109069 | 19 | 4719443 | A | G | 0.3056227 | 0.0440513 | 3.97936E-12 | Serum_C | 0.003027022 | 0.22166635 | Serum total cholesterol(mmol/l) |
| rs2109069 | 19 | 4719443 | A | G | 0.3056227 | 0.0440513 | 3.97936E-12 | XS_VLDL_PL | 0.003027473 | 0.21893085 | Phospholipids in very small VLDL(mmol/l) |
| rs2109069 | 19 | 4719443 | A | G | 0.3056227 | 0.0440513 | 3.97936E-12 | L_LDL_P | 0.003027809 | 0.21694444 | Concentration of large LDL particles(mol/l) |
| rs2109069 | 19 | 4719443 | A | G | 0.3056227 | 0.0440513 | 3.97936E-12 | M_LDL_FC_percent | 0.003068784 | -0.2128111 | Free cholesterol to total lipids ratio in medium LDL(%) |
| rs2109069 | 19 | 4719443 | A | G | 0.3056227 | 0.0440513 | 3.97936E-12 | IDL_FC | 0.003099146 | 0.22419877 | Free cholesterol in IDL(mmol/l) |
| rs10087754 | 8 | 122832148 | T | A | 0.1935335 | 0.0432237 | 7.55487E-06 | S_HDL_P | 0.003111907 | -0.21288861 | Concentration of small HDL particles(mol/l) |
| rs2109069 | 19 | 4719443 | A | G | 0.3056227 | 0.0440513 | 3.97936E-12 | M_LDL_P | 0.00311501 | 0.21829278 | Concentration of medium LDL particles(mol/l) |
| rs2109069 | 19 | 4719443 | A | G | 0.3056227 | 0.0440513 | 3.97936E-12 | L_LDL_L | 0.003115778 | 0.21735599 | Total lipids in large LDL(mmol/l) |
| rs4366152 | 9 | 117564875 | T | C | -0.2109346 | 0.0464797 | 5.6731E-06 | S_VLDL_FC_percent | 0.003169773 | 0.20429961 | Free cholesterol to total lipids ratio in small VLDL(%) |
| rs2109069 | 19 | 4719443 | A | G | 0.3056227 | 0.0440513 | 3.97936E-12 | M_LDL_C | 0.003179822 | 0.22228353 | Total cholesterol in medium LDL(mmol/l) |
| rs2109069 | 19 | 4719443 | A | G | 0.3056227 | 0.0440513 | 3.97936E-12 | IDL_CE | 0.003234932 | 0.22378123 | Cholesterol esters in IDL(mmol/l) |
| rs2109069 | 19 | 4719443 | A | G | 0.3056227 | 0.0440513 | 3.97936E-12 | L_LDL_CE | 0.003264233 | 0.2169391 | Cholesterol esters in large LDL(mmol/l) |
| rs2109069 | 19 | 4719443 | A | G | 0.3056227 | 0.0440513 | 3.97936E-12 | M_LDL_L | 0.003287235 | 0.21815568 | Total lipids in medium LDL(mmol/l) |
| rs2109069 | 19 | 4719443 | A | G | 0.3056227 | 0.0440513 | 3.97936E-12 | EstC | 0.003300097 | 0.21894117 | Esterified cholesterol(mmol/l) |
| rs10087754 | 8 | 122832148 | T | A | 0.1935335 | 0.0432237 | 7.55487E-06 | S_HDL_L | 0.003324928 | -0.2114317 | Total lipids in small HDL(mmol/l) |
| rs2109069 | 19 | 4719443 | A | G | 0.3056227 | 0.0440513 | 3.97936E-12 | L_LDL_C | 0.003460084 | 0.21639487 | Total cholesterol in large LDL(mmol/l) |
| rs4683167 | 3 | 46092288 | T | A | 0.2173025 | 0.0437008 | 6.60953E-07 | AcAce | 0.00361894 | 0.23886279 | Acetoacetate(mmol/l) |
| rs2109069 | 19 | 4719443 | A | G | 0.3056227 | 0.0440513 | 3.97936E-12 | M_LDL_CE_percent | 0.003697195 | 0.22892174 | Cholesterol esters to total lipids ratio in medium LDL(%) |
| rs4256368 | 5 | 162772648 | G | A | 0.2609633 | 0.0581836 | 7.28458E-06 | XS_VLDL_PL_percent | 0.003768214 | 0.28467992 | Phospholipids to total lipds ratio in very small VLDL(%) |
| rs4299751 | 5 | 162772358 | A | G | 0.2573595 | 0.0581598 | 9.64344E-06 | XS_VLDL_PL_percent | 0.003783592 | 0.28453785 | Phospholipids to total lipds ratio in very small VLDL(%) |
| rs4767023 | 12 | 113352159 | T | C | -0.2215523 | 0.0462147 | 1.63515E-06 | Crea | 0.003838418 | 0.20376846 | Creatinine(mmol/l) |
| rs2109069 | 19 | 4719443 | A | G | 0.3056227 | 0.0440513 | 3.97936E-12 | S_LDL_FC | 0.003840053 | 0.22357082 | Free cholesterol in small LDL(mmol/l) |
| rs2057778 | 12 | 113350796 | G | T | -0.2102767 | 0.0461802 | 5.27842E-06 | Crea | 0.003853367 | 0.20366812 | Creatinine(mmol/l) |
| rs2109069 | 19 | 4719443 | A | G | 0.3056227 | 0.0440513 | 3.97936E-12 | LDL_C | 0.003892828 | 0.21373606 | Total cholesterol in LDL(mmol/l) |
| rs302784 | 1 | 99628037 | G | A | 0.2558895 | 0.0568914 | 6.86364E-06 | IDL_CE_percent | 0.003958545 | -0.27623496 | Cholesterol esters to total lipids ratio in IDL(%) |
| rs302785 | 1 | 99628110 | A | G | 0.2566557 | 0.0568882 | 6.43538E-06 | IDL_CE_percent | 0.003958545 | -0.27623496 | Cholesterol esters to total lipids ratio in IDL(%) |
| rs302786 | 1 | 99628220 | A | G | 0.2558895 | 0.0568927 | 6.86677E-06 | IDL_CE_percent | 0.003958545 | -0.27623496 | Cholesterol esters to total lipids ratio in IDL(%) |
| rs4908307 | 1 | 99631619 | T | C | 0.2577073 | 0.056613 | 5.31198E-06 | IDL_CE_percent | 0.003958545 | -0.27623496 | Cholesterol esters to total lipids ratio in IDL(%) |
| rs4907993 | 1 | 99631626 | T | G | 0.2577382 | 0.056614 | 5.3007E-06 | IDL_CE_percent | 0.003958545 | -0.27623496 | Cholesterol esters to total lipids ratio in IDL(%) |
| rs302808 | 1 | 99641652 | T | C | 0.2578464 | 0.0564942 | 5.01538E-06 | IDL_CE_percent | 0.003958545 | -0.27623496 | Cholesterol esters to total lipids ratio in IDL(%) |
| rs302784 | 1 | 99628037 | G | A | 0.2558895 | 0.0568914 | 6.86364E-06 | XS_VLDL_CE_percent | 0.003962018 | -0.28921031 | Cholesterol esters to total lipids ratio in very small VLDL(%) |
| rs302785 | 1 | 99628110 | A | G | 0.2566557 | 0.0568882 | 6.43538E-06 | XS_VLDL_CE_percent | 0.003962018 | -0.28921031 | Cholesterol esters to total lipids ratio in very small VLDL(%) |
| rs302786 | 1 | 99628220 | A | G | 0.2558895 | 0.0568927 | 6.86677E-06 | XS_VLDL_CE_percent | 0.003962018 | -0.28921031 | Cholesterol esters to total lipids ratio in very small VLDL(%) |
| rs4908307 | 1 | 99631619 | T | C | 0.2577073 | 0.056613 | 5.31198E-06 | XS_VLDL_CE_percent | 0.003962018 | -0.28921031 | Cholesterol esters to total lipids ratio in very small VLDL(%) |
| rs4907993 | 1 | 99631626 | T | G | 0.2577382 | 0.056614 | 5.3007E-06 | XS_VLDL_CE_percent | 0.003962018 | -0.28921031 | Cholesterol esters to total lipids ratio in very small VLDL(%) |
| rs302808 | 1 | 99641652 | T | C | 0.2578464 | 0.0564942 | 5.01538E-06 | XS_VLDL_CE_percent | 0.003962018 | -0.28921031 | Cholesterol esters to total lipids ratio in very small VLDL(%) |
| rs302807 | 1 | 99639835 | A | G | 0.2572976 | 0.056468 | 5.2004E-06 | IDL_CE_percent | 0.00398217 | -0.27677213 | Cholesterol esters to total lipids ratio in IDL(%) |
| rs302807 | 1 | 99639835 | A | G | 0.2572976 | 0.056468 | 5.2004E-06 | XS_VLDL_CE_percent | 0.00398378 | -0.28978749 | Cholesterol esters to total lipids ratio in very small VLDL(%) |
| rs2860743 | 5 | 162768043 | T | A | 0.2611404 | 0.0582178 | 7.27344E-06 | XS_VLDL_PL_percent | 0.004185511 | 0.28084251 | Phospholipids to total lipds ratio in very small VLDL(%) |
| rs11085727 | 19 | 10466123 | T | C | 0.2430246 | 0.0460589 | 1.31748E-07 | XXL_VLDL_C_percent | 0.004310017 | 0.24133163 | Total cholesterol to total lipids ratio in chylomicrons and extremely large VLDL(%) |
| rs2109069 | 19 | 4719443 | A | G | 0.3056227 | 0.0440513 | 3.97936E-12 | L_LDL_PL | 0.004420701 | 0.2090713 | Phospholipids in large LDL(mmol/l) |
| rs2109069 | 19 | 4719443 | A | G | 0.3056227 | 0.0440513 | 3.97936E-12 | FAw6 | 0.004612525 | 0.21128796 | Omega_6 fatty acids(mmol/l) |
| rs2544971 | 5 | 162766078 | T | C | 0.2605241 | 0.058244 | 7.71508E-06 | XS_VLDL_PL_percent | 0.004663921 | 0.27770118 | Phospholipids to total lipds ratio in very small VLDL(%) |
| rs302784 | 1 | 99628037 | G | A | 0.2558895 | 0.0568914 | 6.86364E-06 | IDL_C_percent | 0.004788094 | -0.25906453 | Total cholesterol to total lipids ratio in IDL(%) |
| rs302785 | 1 | 99628110 | A | G | 0.2566557 | 0.0568882 | 6.43538E-06 | IDL_C_percent | 0.004788094 | -0.25906453 | Total cholesterol to total lipids ratio in IDL(%) |
| rs302786 | 1 | 99628220 | A | G | 0.2558895 | 0.0568927 | 6.86677E-06 | IDL_C_percent | 0.004788094 | -0.25906453 | Total cholesterol to total lipids ratio in IDL(%) |
| rs4908307 | 1 | 99631619 | T | C | 0.2577073 | 0.056613 | 5.31198E-06 | IDL_C_percent | 0.004788094 | -0.25906453 | Total cholesterol to total lipids ratio in IDL(%) |
| rs4907993 | 1 | 99631626 | T | G | 0.2577382 | 0.056614 | 5.3007E-06 | IDL_C_percent | 0.004788094 | -0.25906453 | Total cholesterol to total lipids ratio in IDL(%) |
| rs302808 | 1 | 99641652 | T | C | 0.2578464 | 0.0564942 | 5.01538E-06 | IDL_C_percent | 0.004788094 | -0.25906453 | Total cholesterol to total lipids ratio in IDL(%) |
| rs12657692 | 5 | 162765185 | T | G | 0.2606628 | 0.0582457 | 7.63065E-06 | XS_VLDL_PL_percent | 0.004790811 | 0.27690133 | Phospholipids to total lipds ratio in very small VLDL(%) |
| rs302807 | 1 | 99639835 | A | G | 0.2572976 | 0.056468 | 5.2004E-06 | IDL_C_percent | 0.004831169 | -0.25947381 | Total cholesterol to total lipids ratio in IDL(%) |
| rs4234452 | 3 | 46047767 | C | G | -0.1997213 | 0.0440866 | 5.89248E-06 | Cit | 0.004996638 | -0.21580778 | Citrate(mmol/l) |
| rs10057923 | 5 | 162762696 | C | G | 0.2599151 | 0.058263 | 8.15725E-06 | XS_VLDL_PL_percent | 0.005102708 | 0.2749613 | Phospholipids to total lipds ratio in very small VLDL(%) |
| rs12495073 | 3 | 46053882 | G | T | -0.2055529 | 0.0440441 | 3.05638E-06 | Cit | 0.005164794 | -0.21643782 | Citrate(mmol/l) |
| rs28808964 | 3 | 46045182 | T | C | -0.1992586 | 0.0440795 | 6.17134E-06 | Cit | 0.005165524 | -0.21502996 | Citrate(mmol/l) |
| rs2109069 | 19 | 4719443 | A | G | 0.3056227 | 0.0440513 | 3.97936E-12 | L_LDL_FC | 0.005275202 | 0.20932358 | Free cholesterol in large LDL(mmol/l) |
| rs2109069 | 19 | 4719443 | A | G | 0.3056227 | 0.0440513 | 3.97936E-12 | PUFA | 0.005308699 | 0.20675444 | Polyunsaturated fatty acids(mmol/l) |
| rs1846616 | 3 | 46032441 | A | T | 0.2351907 | 0.0439068 | 8.48375E-08 | L_HDL_PL | 0.005377656 | -0.27321713 | Phospholipids in large HDL(mmol/l) |
| rs4076440 | 1 | 9690476 | C | T | 0.301866 | 0.0682837 | 9.83419E-06 | S_VLDL_TG_percent | 0.005403031 | 0.33010496 | Triglycerides to total lipids ratio in small VLDL(%) |
| rs2109069 | 19 | 4719443 | A | G | 0.3056227 | 0.0440513 | 3.97936E-12 | HDL3_C | 0.005412825 | 0.2131784 | Total cholesterol in HDL3(mmol/l) |
| rs2323476 | 13 | 37966084 | A | G | -0.1870708 | 0.0422955 | 9.73636E-06 | M_VLDL_C_percent | 0.005449388 | 0.18995867 | Total cholesterol to total lipids ratio in medium VLDL(%) |
| rs4299751 | 5 | 162772358 | A | G | 0.2573595 | 0.0581598 | 9.64344E-06 | XL_VLDL_FC_percent | 0.005563947 | 0.26187926 | Free cholesterol to total lipids ratio in very large VLDL(%) |
| rs4256368 | 5 | 162772648 | G | A | 0.2609633 | 0.0581836 | 7.28458E-06 | XL_VLDL_FC_percent | 0.005571513 | 0.26185461 | Free cholesterol to total lipids ratio in very large VLDL(%) |
| rs2860743 | 5 | 162768043 | T | A | 0.2611404 | 0.0582178 | 7.27344E-06 | XL_VLDL_FC_percent | 0.005601971 | 0.26110795 | Free cholesterol to total lipids ratio in very large VLDL(%) |
| rs1846616 | 3 | 46032441 | A | T | 0.2351907 | 0.0439068 | 8.48375E-08 | M_HDL_P | 0.005634375 | -0.26026929 | Concentration of medium HDL particles(mol/l) |
| rs7135577 | 12 | 113358106 | A | G | -0.2223601 | 0.0462273 | 1.50815E-06 | Crea | 0.00568755 | 0.19413715 | Creatinine(mmol/l) |
| rs4767026 | 12 | 113359132 | A | G | -0.2192374 | 0.0461869 | 2.06725E-06 | Crea | 0.005720606 | 0.19399437 | Creatinine(mmol/l) |
| rs1846616 | 3 | 46032441 | A | T | 0.2351907 | 0.0439068 | 8.48375E-08 | M_HDL_L | 0.005763747 | -0.26019935 | Total lipids in medium HDL(mmol/l) |
| rs2109069 | 19 | 4719443 | A | G | 0.3056227 | 0.0440513 | 3.97936E-12 | S_LDL_CE | 0.00583926 | 0.21219168 | Cholesterol esters in small LDL(mmol/l) |
| rs10850097 | 12 | 113361117 | C | T | -0.2258898 | 0.0461035 | 9.60255E-07 | Crea | 0.005840366 | 0.19293571 | Creatinine(mmol/l) |
| rs2544971 | 5 | 162766078 | T | C | 0.2605241 | 0.058244 | 7.71508E-06 | XL_VLDL_FC_percent | 0.005932605 | 0.25954641 | Free cholesterol to total lipids ratio in very large VLDL(%) |
| rs6766332 | 3 | 46101203 | A | G | 0.1980883 | 0.043773 | 6.02874E-06 | Cit | 0.006068554 | 0.21115607 | Citrate(mmol/l) |
| rs2323476 | 13 | 37966084 | A | G | -0.1870708 | 0.0422955 | 9.73636E-06 | IDL_C_percent | 0.006075625 | 0.17247737 | Total cholesterol to total lipids ratio in IDL(%) |
| rs12657692 | 5 | 162765185 | T | G | 0.2606628 | 0.0582457 | 7.63065E-06 | XL_VLDL_FC_percent | 0.006081133 | 0.25882212 | Free cholesterol to total lipids ratio in very large VLDL(%) |
| rs1392295 | 3 | 46101104 | T | A | 0.1980473 | 0.0437725 | 6.05667E-06 | Cit | 0.006084505 | 0.21106396 | Citrate(mmol/l) |
| rs1392294 | 3 | 46100999 | A | G | 0.1980637 | 0.0437718 | 6.0436E-06 | Cit | 0.006100608 | 0.21097092 | Citrate(mmol/l) |
| rs2173640 | 3 | 46100496 | A | G | 0.2021569 | 0.0437283 | 3.78101E-06 | Cit | 0.006184575 | 0.2105506 | Citrate(mmol/l) |
| rs2133663 | 3 | 46100294 | G | C | 0.1979898 | 0.0437715 | 6.08948E-06 | Cit | 0.006199142 | 0.21048963 | Citrate(mmol/l) |
| rs10057923 | 5 | 162762696 | C | G | 0.2599151 | 0.058263 | 8.15725E-06 | XL_VLDL_FC_percent | 0.006369416 | 0.25742697 | Free cholesterol to total lipids ratio in very large VLDL(%) |
| rs299284 | 5 | 162896650 | T | C | 0.2919903 | 0.0632056 | 3.84234E-06 | M_VLDL_PL_percent | 0.006390485 | 0.29567512 | Phospholipids to total lipds ratio in medium VLDL(%) |
| rs4767028 | 12 | 113359188 | A | G | -0.2208262 | 0.0461964 | 1.75143E-06 | Crea | 0.006474938 | 0.19070166 | Creatinine(mmol/l) |
| rs6489865 | 12 | 113360302 | A | G | -0.2222252 | 0.046229 | 1.53177E-06 | Crea | 0.006474938 | 0.19070166 | Creatinine(mmol/l) |
| rs4076440 | 1 | 9690476 | C | T | 0.301866 | 0.0682837 | 9.83419E-06 | S_VLDL_CE_percent | 0.006485678 | -0.32065085 | Cholesterol esters to total lipids ratio in small VLDL(%) |
| rs2109069 | 19 | 4719443 | A | G | 0.3056227 | 0.0440513 | 3.97936E-12 | M_LDL_FC | 0.006486885 | 0.20541094 | Free cholesterol in medium LDL(mmol/l) |
| rs4767027 | 12 | 113359157 | T | C | -0.2215948 | 0.0461449 | 1.56962E-06 | Crea | 0.006527519 | 0.19081481 | Creatinine(mmol/l) |
| rs4767024 | 12 | 113358791 | T | C | -0.2216634 | 0.0461518 | 1.56362E-06 | Crea | 0.006540446 | 0.19077244 | Creatinine(mmol/l) |
| rs4767025 | 12 | 113358794 | C | T | -0.2198041 | 0.0461418 | 1.90095E-06 | Crea | 0.006540446 | 0.19077244 | Creatinine(mmol/l) |
| rs10774671 | 12 | 113357193 | G | A | -0.2281498 | 0.0460801 | 7.37785E-07 | Crea | 0.006552078 | 0.19063648 | Creatinine(mmol/l) |
| rs2323476 | 13 | 37966084 | A | G | -0.1870708 | 0.0422955 | 9.73636E-06 | M_VLDL_CE_percent | 0.006552964 | 0.18965221 | Cholesterol esters to total lipids ratio in medium VLDL(%) |
| rs4299751 | 5 | 162772358 | A | G | 0.2573595 | 0.0581598 | 9.64344E-06 | M_LDL_C | 0.006929772 | 0.26304875 | Total cholesterol in medium LDL(mmol/l) |
| rs4256368 | 5 | 162772648 | G | A | 0.2609633 | 0.0581836 | 7.28458E-06 | M_LDL_C | 0.006936895 | 0.26303254 | Total cholesterol in medium LDL(mmol/l) |
| rs4299751 | 5 | 162772358 | A | G | 0.2573595 | 0.0581598 | 9.64344E-06 | M_LDL_L | 0.007001814 | 0.25872812 | Total lipids in medium LDL(mmol/l) |

| | | | | | | | | | | | |
|---|---|---|---|---|---|---|---|---|---|---|---|
| rs4256368 | 5 | 162772648 | G | A | 0.2609633 | 0.0581836 | 7.28458E-06 | M_LDL_L | 0.007010662 | 0.25870466 | Total lipids in medium LDL(mmol/l) |
| rs934191 | 15 | 54395262 | G | T | 0.2177128 | 0.0480675 | 5.9157E-06 | S_HDL_FC_percent | 0.007016881 | 0.19990247 | Free cholesterol to total lipids ratio in small HDL(%) |
| rs12622794 | 2 | 53809057 | G | A | 0.1937065 | 0.0429963 | 6.62852E-06 | L_VLDL_PL_percent | 0.007023881 | -0.19383835 | Phospholipids to total lipds ratio in large VLDL(%) |
| rs4299751 | 5 | 162772358 | A | G | 0.2573595 | 0.0581598 | 9.64344E-06 | M_LDL_P | 0.007176559 | 0.25667701 | Concentration of medium LDL particles(mol/l) |
| rs4256368 | 5 | 162772648 | G | A | 0.2609633 | 0.0581836 | 7.28458E-06 | M_LDL_P | 0.00718616 | 0.25665119 | Concentration of medium LDL particles(mol/l) |
| rs2109069 | 19 | 4719443 | A | G | 0.3056227 | 0.0440513 | 3.97936E-12 | M_LDL_CE | 0.0072513 | 0.20595354 | Cholesterol esters in medium LDL(mmol/l) |
| rs2860743 | 5 | 162768043 | T | A | 0.2611404 | 0.0582178 | 7.27344E-06 | M_LDL_C | 0.007289801 | 0.26085746 | Total cholesterol in medium LDL(mmol/l) |
| rs7961861 | 12 | 113385918 | C | T | -0.1897668 | 0.0429529 | 9.96094E-06 | L_LDL_TG_percent | 0.007326301 | 0.16981004 | Triglycerides to total lipids ratio in large LDL(%) |
| rs6489864 | 12 | 113360025 | A | G | -0.2199885 | 0.0461481 | 1.86979E-06 | Crea | 0.0073279 | 0.18771701 | Creatinine(mmol/l) |
| rs7961128 | 12 | 113385246 | C | T | -0.1898055 | 0.0429659 | 9.9805E-06 | L_LDL_TG_percent | 0.007328873 | 0.16982467 | Triglycerides to total lipids ratio in large LDL(%) |
| rs934191 | 15 | 54395262 | G | T | 0.2177128 | 0.0480675 | 5.9157E-06 | S_HDL_CE | 0.007334446 | -0.21759619 | Cholesterol esters in small HDL(mmol/l) |
| rs2860743 | 5 | 162768043 | T | A | 0.2611404 | 0.0582178 | 7.27344E-06 | M_LDL_L | 0.007335745 | 0.2566977 | Total lipids in medium LDL(mmol/l) |
| rs4767029 | 12 | 113359318 | G | A | -0.2212266 | 0.046164 | 1.64972E-06 | Crea | 0.007345423 | 0.18765453 | Creatinine(mmol/l) |
| rs10850094 | 12 | 113360563 | T | C | -0.2216309 | 0.0461566 | 1.57317E-06 | Crea | 0.0073612 | 0.18762146 | Creatinine(mmol/l) |
| rs10850095 | 12 | 113360575 | T | C | -0.2216022 | 0.0461562 | 1.57767E-06 | Crea | 0.0073612 | 0.18762146 | Creatinine(mmol/l) |
| rs10774672 | 12 | 113360737 | G | T | -0.2212503 | 0.0461548 | 1.6378E-06 | Crea | 0.0073612 | 0.18762146 | Creatinine(mmol/l) |
| rs10850096 | 12 | 113360786 | T | C | -0.2217882 | 0.04616 | 1.54919E-06 | Crea | 0.0073612 | 0.18762146 | Creatinine(mmol/l) |
| rs7953402 | 12 | 113390748 | C | T | -0.1924446 | 0.042964 | 7.49271E-06 | L_LDL_TG_percent | 0.007475866 | 0.16921617 | Triglycerides to total lipids ratio in large LDL(%) |
| rs13084541 | 3 | 46038447 | G | A | 0.2226714 | 0.0436187 | 3.30734E-07 | XL_HDL_TG_percent | 0.007484137 | 0.20338488 | Triglycerides to total lipids ratio in very large HDL(%) |
| rs7310791 | 12 | 113392065 | T | G | -0.1922046 | 0.0429663 | 7.69924E-06 | L_LDL_TG_percent | 0.007487784 | 0.16920157 | Triglycerides to total lipids ratio in large LDL(%) |
| rs13064616 | 3 | 46038644 | T | C | 0.2227595 | 0.0436193 | 3.27357E-07 | XL_HDL_TG_percent | 0.007504104 | 0.20331184 | Triglycerides to total lipids ratio in very large HDL(%) |
| rs2860743 | 5 | 162768043 | T | A | 0.2611404 | 0.0582178 | 7.27344E-06 | M_LDL_P | 0.007517327 | 0.25466071 | Concentration of medium LDL particles(mol/l) |
| rs13063033 | 3 | 46082481 | A | G | 0.4971019 | 0.0616938 | 7.78443E-16 | Crea | 0.007519028 | 0.29539179 | Creatinine(mmol/l) |
| rs2107418 | 12 | 113398139 | C | A | -0.1928569 | 0.0429973 | 7.28058E-06 | L_LDL_TG_percent | 0.007529172 | 0.1691525 | Triglycerides to total lipids ratio in large LDL(%) |
| rs10850093 | 12 | 113360468 | C | T | -0.2216022 | 0.0461559 | 1.57751E-06 | Crea | 0.00755474 | 0.18695992 | Creatinine(mmol/l) |
| rs4767030 | 12 | 113359577 | C | T | -0.219423 | 0.046142 | 1.98073E-06 | Crea | 0.007555806 | 0.18694798 | Creatinine(mmol/l) |
| rs10850092 | 12 | 113359703 | C | G | -0.220714 | 0.0461682 | 1.74721E-06 | Crea | 0.007555806 | 0.18694798 | Creatinine(mmol/l) |
| rs57542549 | 3 | 46039152 | T | C | 0.2227595 | 0.0436216 | 3.2783E-07 | XL_HDL_TG_percent | 0.007556237 | 0.2031041 | Triglycerides to total lipids ratio in very large HDL(%) |
| rs2860743 | 5 | 162768043 | T | A | 0.2611404 | 0.0582178 | 7.27344E-06 | M_VLDL_C_percent | 0.007595332 | 0.25393766 | Total cholesterol to total lipids ratio in medium VLDL(%) |
| rs1131476 | 12 | 113357209 | G | A | -0.2569822 | 0.0462203 | 2.69863E-08 | Crea | 0.007599205 | 0.18681335 | Creatinine(mmol/l) |
| rs2660 | 12 | 113357442 | G | A | -0.2570559 | 0.0462403 | 2.71149E-08 | Crea | 0.007599205 | 0.18681335 | Creatinine(mmol/l) |
| rs7612402 | 3 | 46039599 | T | C | 0.2228795 | 0.0436286 | 3.24747E-07 | XL_HDL_TG_percent | 0.007613 | 0.20288898 | Triglycerides to total lipids ratio in very large HDL(%) |
| rs2891413 | 12 | 113397125 | C | T | -0.1918048 | 0.0429836 | 8.10899E-06 | L_LDL_TG_percent | 0.007643163 | 0.16911401 | Triglycerides to total lipids ratio in large LDL(%) |
| rs2109069 | 19 | 4719443 | A | G | 0.3056227 | 0.0440513 | 3.97936E-12 | Remnant_C | 0.007715223 | 0.20447434 | Remnant cholesterol (non_HDL, non_LDL _cholesterol)(mmol/l) |
| rs1392294 | 3 | 46100999 | A | G | 0.1980637 | 0.0437718 | 6.0436E-06 | LDL_D | 0.007716977 | 0.19822173 | Mean diameter for LDL particles(nm) |
| rs2129881 | 3 | 46040678 | A | G | 0.2234635 | 0.0436404 | 3.04534E-07 | XL_HDL_TG_percent | 0.007725108 | 0.20242433 | Triglycerides to total lipids ratio in very large HDL(%) |
| rs1392295 | 3 | 46101104 | T | A | 0.1980473 | 0.0437725 | 6.05667E-06 | LDL_D | 0.007730896 | 0.1982025 | Mean diameter for LDL particles(nm) |
| rs6766332 | 3 | 46101203 | A | G | 0.1980883 | 0.043773 | 6.02874E-06 | LDL_D | 0.007744983 | 0.19818236 | Mean diameter for LDL particles(nm) |
| rs2173640 | 3 | 46100496 | A | G | 0.2021569 | 0.0437283 | 3.78101E-06 | LDL_D | 0.007756793 | 0.19802033 | Mean diameter for LDL particles(nm) |
| rs2133663 | 3 | 46100294 | G | C | 0.1979898 | 0.0437715 | 6.08948E-06 | LDL_D | 0.007776661 | 0.19795497 | Mean diameter for LDL particles(nm) |
| rs12485445 | 3 | 46041582 | C | T | 0.2230155 | 0.0436267 | 3.18956E-07 | XL_HDL_TG_percent | 0.007838946 | 0.20198807 | Triglycerides to total lipids ratio in very large HDL(%) |
| rs2860743 | 5 | 162768043 | T | A | 0.2611404 | 0.0582178 | 7.27344E-06 | M_VLDL_TG_percent | 0.007855117 | -0.25586112 | Triglycerides to total lipids ratio in medium VLDL(%) |
| rs2373089 | 3 | 46041909 | C | T | 0.2230875 | 0.0436272 | 3.16116E-07 | XL_HDL_TG_percent | 0.007877431 | 0.20184133 | Triglycerides to total lipids ratio in very large HDL(%) |
| rs2431752 | 5 | 162882702 | A | G | 0.2882119 | 0.0632158 | 5.13516E-06 | M_VLDL_PL_percent | 0.007921466 | 0.28748241 | Phospholipids to total lipds ratio in medium VLDL(%) |
| rs1846616 | 3 | 46032441 | A | T | 0.2351907 | 0.0439068 | 8.48375E-08 | XXL_VLDL_C_percent | 0.007939286 | -0.27779092 | Total cholesterol to total lipids ratio in chylomicrons and extremely large VLDL(%) |
| rs35283688 | 3 | 46042413 | A | T | 0.2229835 | 0.0436289 | 3.20477E-07 | XL_HDL_TG_percent | 0.00795522 | 0.20154585 | Triglycerides to total lipids ratio in very large HDL(%) |
| rs2109069 | 19 | 4719443 | A | G | 0.3056227 | 0.0440513 | 3.97936E-12 | M_LDL_TG | 0.008100379 | 0.17420731 | Triglycerides in medium LDL(mmol/l) |
| rs2544971 | 5 | 162766078 | T | C | 0.2605241 | 0.058244 | 7.71508E-06 | M_VLDL_C_percent | 0.008126898 | 0.25197045 | Total cholesterol to total lipids ratio in medium VLDL(%) |
| rs4299751 | 5 | 162772358 | A | G | 0.2573595 | 0.0581598 | 9.64344E-06 | M_VLDL_C_percent | 0.008132705 | 0.25231391 | Total cholesterol to total lipids ratio in medium VLDL(%) |
| rs4256368 | 5 | 162772648 | G | A | 0.2609633 | 0.0581836 | 7.28458E-06 | M_VLDL_C_percent | 0.008166037 | 0.25219944 | Total cholesterol to total lipids ratio in medium VLDL(%) |
| rs11085727 | 19 | 10466123 | T | C | 0.2430246 | 0.0460589 | 1.31748E-07 | S_VLDL_TG | 0.008182205 | -0.22371465 | Triglycerides in small VLDL(mmol/l) |
| rs4299751 | 5 | 162772358 | A | G | 0.2573595 | 0.0581598 | 9.64344E-06 | S_LDL_C | 0.008198154 | 0.26098046 | Total cholesterol in small LDL(mmol/l) |
| rs12705891 | 7 | 113317708 | C | T | 0.2753944 | 0.0434621 | 2.35272E-10 | TG_PG_ratio | 0.008202821 | -0.20527225 | Ratio of triglycerides to phosphoglycerides |
| rs4256368 | 5 | 162772648 | G | A | 0.2609633 | 0.0581836 | 7.28458E-06 | S_LDL_C | 0.008210087 | 0.26094896 | Total cholesterol in small LDL(mmol/l) |
| rs2544971 | 5 | 162766078 | T | C | 0.2605241 | 0.058244 | 7.71508E-06 | M_LDL_L | 0.00826562 | 0.25307071 | Total lipids in medium LDL(mmol/l) |
| rs12657692 | 5 | 162765185 | T | G | 0.2606628 | 0.0582457 | 7.63065E-06 | M_VLDL_C_percent | 0.008295421 | 0.25134688 | Total cholesterol to total lipids ratio in medium VLDL(%) |
| rs2544971 | 5 | 162766078 | T | C | 0.2605241 | 0.058244 | 7.71508E-06 | M_LDL_C | 0.00829987 | 0.2568376 | Total cholesterol in medium LDL(mmol/l) |
| rs2236757 | 21 | 34624917 | A | G | 0.2510622 | 0.0460548 | 4.99845E-08 | M_LDL_FC_percent | 0.008304611 | -0.21022024 | Free cholesterol to total lipids ratio in medium LDL(%) |
| rs2544971 | 5 | 162766078 | T | C | 0.2605241 | 0.058244 | 7.71508E-06 | M_VLDL_TG_percent | 0.008381445 | -0.25395219 | Triglycerides to total lipids ratio in medium VLDL(%) |
| rs6766332 | 3 | 46101203 | A | G | 0.1980883 | 0.043773 | 6.02874E-06 | XL_HDL_TG_percent | 0.008396811 | 0.19896372 | Triglycerides to total lipids ratio in very large HDL(%) |
| rs1392295 | 3 | 46101104 | T | A | 0.1980473 | 0.0437725 | 6.05667E-06 | XL_HDL_TG_percent | 0.00843218 | 0.19883214 | Triglycerides to total lipids ratio in very large HDL(%) |
| rs4299751 | 5 | 162772358 | A | G | 0.2573595 | 0.0581598 | 9.64344E-06 | M_VLDL_TG_percent | 0.008450607 | -0.25405507 | Triglycerides to total lipids ratio in medium VLDL(%) |
| rs1392294 | 3 | 46100999 | A | G | 0.1980637 | 0.0437718 | 6.0436E-06 | XL_HDL_TG_percent | 0.008467825 | 0.19869971 | Triglycerides to total lipids ratio in very large HDL(%) |
| rs2544971 | 5 | 162766078 | T | C | 0.2605241 | 0.058244 | 7.71508E-06 | M_LDL_P | 0.00847318 | 0.25102979 | Concentration of medium LDL particles(mol/l) |
| rs4256368 | 5 | 162772648 | G | A | 0.2609633 | 0.0581836 | 7.28458E-06 | M_VLDL_TG_percent | 0.008488914 | -0.25392458 | Triglycerides to total lipids ratio in medium VLDL(%) |
| rs12657692 | 5 | 162765185 | T | G | 0.2606628 | 0.0582457 | 7.63065E-06 | M_LDL_L | 0.008512286 | 0.25215573 | Total lipids in medium LDL(mmol/l) |
| rs2860743 | 5 | 162768043 | T | A | 0.2611404 | 0.0582178 | 7.27344E-06 | S_LDL_C | 0.008523019 | 0.25913301 | Total cholesterol in small LDL(mmol/l) |
| rs2129879 | 3 | 46046769 | T | C | 0.2217426 | 0.0436362 | 3.74065E-07 | XL_HDL_TG_percent | 0.008531419 | 0.19940493 | Triglycerides to total lipids ratio in very large HDL(%) |
| rs2173640 | 3 | 46100496 | A | G | 0.2021569 | 0.0437283 | 3.78101E-06 | XL_HDL_TG_percent | 0.008539004 | 0.1984132 | Triglycerides to total lipids ratio in very large HDL(%) |
| rs12657692 | 5 | 162765185 | T | G | 0.2606628 | 0.0582457 | 7.63065E-06 | M_VLDL_TG_percent | 0.008546804 | -0.25335089 | Triglycerides to total lipids ratio in medium VLDL(%) |
| rs2133663 | 3 | 46100294 | G | C | 0.1979898 | 0.0437715 | 6.08948E-06 | XL_HDL_TG_percent | 0.008558558 | 0.19835313 | Triglycerides to total lipids ratio in very large HDL(%) |
| rs12657692 | 5 | 162765185 | T | G | 0.2606628 | 0.0582457 | 7.63065E-06 | M_LDL_C | 0.008562613 | 0.25585044 | Total cholesterol in medium LDL(mmol/l) |
| rs934191 | 15 | 54395262 | G | T | 0.2177128 | 0.0480675 | 5.9157E-06 | XXL_VLDL_PL_percent | 0.008570744 | 0.21187202 | Phospholipids to total lipds ratio in chylomicrons and extremely large VLDL(%) |
| rs10774673 | 12 | 113361158 | C | T | -0.2222415 | 0.0461681 | 1.48126E-06 | Crea | 0.008570876 | 0.18445311 | Creatinine(mmol/l) |
| rs2171529 | 3 | 46047032 | G | A | 0.2231436 | 0.043637 | 3.16039E-07 | XL_HDL_TG_percent | 0.008574773 | 0.19924724 | Triglycerides to total lipids ratio in very large HDL(%) |
| rs2171528 | 3 | 46047044 | T | C | 0.2217746 | 0.0436481 | 3.75381E-07 | XL_HDL_TG_percent | 0.008574773 | 0.19924724 | Triglycerides to total lipids ratio in very large HDL(%) |
| rs10057923 | 5 | 162762696 | C | G | 0.2599151 | 0.058263 | 8.15725E-06 | XXL_VLDL_TG_percent | 0.008651109 | -0.25544615 | Triglycerides to total lipids ratio in chylomicrons and extremely large VLDL(%) |
| rs4299751 | 5 | 162772358 | A | G | 0.2573595 | 0.0581598 | 9.64344E-06 | M_LDL_FC_percent | 0.008698904 | -0.24388291 | Free cholesterol to total lipids ratio in medium LDL(%) |
| rs10057923 | 5 | 162762696 | C | G | 0.2599151 | 0.058263 | 8.15725E-06 | M_VLDL_C_percent | 0.008703839 | 0.24983414 | Total cholesterol to total lipids ratio in medium VLDL(%) |
| rs4256368 | 5 | 162772648 | G | A | 0.2609633 | 0.0581836 | 7.28458E-06 | M_LDL_FC_percent | 0.008704255 | -0.24387915 | Free cholesterol to total lipids ratio in medium LDL(%) |
| rs12657692 | 5 | 162765185 | T | G | 0.2606628 | 0.0582457 | 7.63065E-06 | XXL_VLDL_TG_percent | 0.00871843 | -0.25516577 | Triglycerides to total lipids ratio in chylomicrons and extremely large VLDL(%) |
| rs12657692 | 5 | 162765185 | T | G | 0.2606628 | 0.0582457 | 7.63065E-06 | M_LDL_P | 0.008727414 | 0.25011159 | Concentration of medium LDL particles(mol/l) |
| rs7642701 | 3 | 46091058 | G | A | 0.2750678 | 0.045002 | 9.82172E-10 | AcAce | 0.0087345 | 0.2258102 | Acetoacetate(mmol/l) |
| rs2171529 | 3 | 46047032 | G | A | 0.2231436 | 0.043637 | 3.16039E-07 | Cit | 0.008767252 | 0.20261836 | Citrate(mmol/l) |
| rs2171528 | 3 | 46047044 | T | C | 0.2217746 | 0.0436481 | 3.75381E-07 | Cit | 0.008767252 | 0.20261836 | Citrate(mmol/l) |
| rs1846616 | 3 | 46032441 | A | T | 0.2351907 | 0.0439068 | 8.48375E-08 | M_HDL_FC | 0.008768367 | -0.25262076 | Free cholesterol in medium HDL(mmol/l) |
| rs2544971 | 5 | 162766078 | T | C | 0.2605241 | 0.058244 | 7.71508E-06 | XXL_VLDL_TG_percent | 0.008781854 | -0.25489663 | Triglycerides to total lipids ratio in chylomicrons and extremely large VLDL(%) |
| rs2834157 | 21 | 34616545 | A | G | 0.244678 | 0.0459929 | 1.03792E-07 | M_LDL_FC_percent | 0.008782543 | -0.20698635 | Free cholesterol to total lipids ratio in medium LDL(%) |
| rs2129879 | 3 | 46046769 | T | C | 0.2217426 | 0.0436362 | 3.74065E-07 | Cit | 0.008796154 | 0.2025614 | Citrate(mmol/l) |
| rs13098271 | 3 | 46093858 | A | G | 0.277071 | 0.0450291 | 7.59521E-10 | AcAce | 0.008845288 | 0.22539356 | Acetoacetate(mmol/l) |
| rs35657218 | 3 | 46094462 | G | A | 0.2755462 | 0.0450449 | 9.52199E-10 | AcAce | 0.008870659 | 0.22529716 | Acetoacetate(mmol/l) |
| rs11085727 | 19 | 10466123 | T | C | 0.2430246 | 0.0460589 | 1.31748E-07 | VLDL_TG | 0.008871613 | -0.2247202 | Triglycerides in VLDL(mmol/l) |
| rs34906043 | 3 | 46094623 | T | C | 0.2717585 | 0.0450788 | 1.655E-09 | AcAce | 0.008883447 | 0.22524844 | Acetoacetate(mmol/l) |
| rs2323476 | 13 | 37966084 | A | G | -0.1870708 | 0.0422955 | 9.73636E-06 | L_VLDL_TG_percent | 0.008899948 | -0.18709408 | Triglycerides to total lipids ratio in large VLDL(%) |
| rs10057923 | 5 | 162762696 | C | G | 0.2599151 | 0.058263 | 8.15725E-06 | M_VLDL_TG_percent | 0.008963811 | -0.25182881 | Triglycerides to total lipids ratio in medium VLDL(%) |
| rs2860743 | 5 | 162768043 | T | A | 0.2611404 | 0.0582178 | 7.27344E-06 | XXL_VLDL_TG_percent | 0.008978554 | -0.25398926 | Triglycerides to total lipids ratio in chylomicrons and extremely large VLDL(%) |
| rs4366152 | 9 | 117564875 | T | C | -0.2109346 | 0.0464797 | 5.6731E-06 | L_HDL_FC_percent | 0.009007619 | 0.18723025 | Free cholesterol to total lipids ratio in large HDL(%) |
| rs11066452 | 12 | 113361574 | G | T | -0.2080444 | 0.0458258 | 5.62847E-06 | Crea | 0.009092419 | 0.18300792 | Creatinine(mmol/l) |
| rs10057923 | 5 | 162762696 | C | G | 0.2599151 | 0.058263 | 8.15725E-06 | M_LDL_L | 0.009122358 | 0.24994231 | Total lipids in medium LDL(mmol/l) |
| rs11066451 | 12 | 113361443 | G | A | -0.2065126 | 0.0458278 | 6.59781E-06 | Crea | 0.009124241 | 0.18290741 | Creatinine(mmol/l) |
| rs10774675 | 12 | 113361237 | C | T | -0.2081601 | 0.045824 | 5.55675E-06 | Crea | 0.009179653 | 0.18277225 | Creatinine(mmol/l) |
| rs35283688 | 3 | 46042413 | A | T | 0.2229835 | 0.0436289 | 3.20477E-07 | Cit | 0.009223913 | 0.20168422 | Citrate(mmol/l) |
| rs10057923 | 5 | 162762696 | C | G | 0.2599151 | 0.058263 | 8.15725E-06 | M_LDL_C | 0.009224605 | 0.25342806 | Total cholesterol in medium LDL(mmol/l) |
| rs7958379 | 12 | 113363284 | A | G | -0.2213876 | 0.04553 | 1.15938E-06 | S_HDL_PL | 0.009235628 | 0.19946092 | Phospholipids in small HDL(mmol/l) |
| rs2373089 | 3 | 46041909 | C | T | 0.2230875 | 0.0436272 | 3.16116E-07 | Cit | 0.00928867 | 0.20154677 | Citrate(mmol/l) |
| rs2860743 | 5 | 162768043 | T | A | 0.2611404 | 0.0582178 | 7.27344E-06 | M_LDL_FC_percent | 0.009290061 | -0.24129403 | Free cholesterol to total lipids ratio in medium LDL(%) |
| rs12485445 | 3 | 46041582 | C | T | 0.2230155 | 0.0436267 | 3.18956E-07 | Cit | 0.009321402 | 0.20147691 | Citrate(mmol/l) |
| rs10057923 | 5 | 162762696 | C | G | 0.2599151 | 0.058263 | 8.15725E-06 | M_LDL_P | 0.009358047 | 0.247885 | Concentration of medium LDL particles(mol/l) |
| rs10774674 | 12 | 113361174 | T | C | -0.2223501 | 0.0461692 | 1.4647E-06 | Crea | 0.009412507 | 0.18229673 | Creatinine(mmol/l) |
| rs2129881 | 3 | 46040678 | A | G | 0.2234635 | 0.0436404 | 3.04534E-07 | Cit | 0.009421031 | 0.20126277 | Citrate(mmol/l) |
| rs2109069 | 19 | 4719443 | A | G | 0.3056227 | 0.0440513 | 3.97936E-12 | S_LDL_CE_percent | 0.009500363 | 0.20563154 | Cholesterol esters to total lipids ratio in small LDL(%) |
| rs2109069 | 19 | 4719443 | A | G | 0.3056227 | 0.0440513 | 3.97936E-12 | M_LDL_PL | 0.009563772 | 0.19116025 | Phospholipids in medium LDL(mmol/l) |
| rs7612402 | 3 | 46039599 | T | C | 0.2228795 | 0.0436286 | 3.24747E-07 | Cit | 0.009573655 | 0.20093321 | Citrate(mmol/l) |
| rs57542549 | 3 | 46039152 | T | C | 0.2227595 | 0.0436216 | 3.2783E-07 | Cit | 0.009590267 | 0.20091329 | Citrate(mmol/l) |
| rs2544971 | 5 | 162766078 | T | C | 0.2605241 | 0.058244 | 7.71508E-06 | S_LDL_C | 0.009608928 | 0.25531413 | Total cholesterol in small LDL(mmol/l) |
| rs2109069 | 19 | 4719443 | A | G | 0.3056227 | 0.0440513 | 3.97936E-12 | S_LDL_PL | 0.009641182 | 0.19412929 | Phospholipids in small LDL(mmol/l) |
| rs4299751 | 5 | 162772358 | A | G | 0.2573595 | 0.0581598 | 9.64344E-06 | S_LDL_L | 0.009656431 | 0.25108413 | Total lipids in small LDL(mmol/l) |
| rs13084541 | 3 | 46038447 | G | A | 0.2226714 | 0.0436187 | 3.30734E-07 | Cit | 0.009660365 | 0.20075918 | Citrate(mmol/l) |
| rs4256368 | 5 | 162772648 | G | A | 0.2609633 | 0.0581836 | 7.28458E-06 | S_LDL_L | 0.00967263 | 0.25104464 | Total lipids in small LDL(mmol/l) |
| rs13064616 | 3 | 46038644 | T | C | 0.2227595 | 0.0436193 | 3.27357E-07 | Cit | 0.009678471 | 0.20070388 | Citrate(mmol/l) |
| rs7958379 | 12 | 113363284 | A | G | -0.2213876 | 0.04553 | 1.15938E-06 | IDL_TG_percent | 0.009749947 | 0.2003153 | Triglycerides to total lipids ratio in IDL(%) |
| rs2384074 | 12 | 113382977 | C | T | -0.2574206 | 0.0464183 | 2.92819E-08 | Crea | 0.009755071 | 0.17999934 | Creatinine(mmol/l) |
| rs4299751 | 5 | 162772358 | A | G | 0.2573595 | 0.0581598 | 9.64344E-06 | S_LDL_P | 0.009755709 | 0.24934155 | Concentration of small LDL particles(mol/l) |
| rs4256368 | 5 | 162772648 | G | A | 0.2609633 | 0.0581836 | 7.28458E-06 | S_LDL_P | 0.009771498 | 0.24930415 | Concentration of small LDL particles(mol/l) |
| rs4299751 | 5 | 162772358 | A | G | 0.2573595 | 0.0581598 | 9.64344E-06 | XXL_VLDL_TG_percent | 0.009861773 | -0.25143056 | Triglycerides to total lipids ratio in chylomicrons and extremely large VLDL(%) |
| rs4256368 | 5 | 162772648 | G | A | 0.2609633 | 0.0581836 | 7.28458E-06 | XXL_VLDL_TG_percent | 0.009867111 | -0.25142853 | Triglycerides to total lipids ratio in chylomicrons and extremely large VLDL(%) |
| rs12657692 | 5 | 162765185 | T | G | 0.2606628 | 0.0582457 | 7.63065E-06 | S_LDL_C | 0.009892013 | 0.25436833 | Total cholesterol in small LDL(mmol/l) |
| rs12622794 | 2 | 53809057 | G | A | 0.1937065 | 0.0429963 | 6.62852E-06 | L_VLDL_CE_percent | 0.0099095 | 0.19520498 | Cholesterol esters to total lipids ratio in large VLDL(%) |
| rs4493469 | 3 | 46177992 | C | T | 0.4298715 | 0.0661781 | 8.26615E-11 | DHA_FA_ratio | 0.009939651 | -0.32456597 | Ratio of 22:6 docosahexaenoic acid to total fatty acids(percent) |

* GWAS loci were selected from E. Pairo-Castineira et.al Nature 2020 ,  https://doi.org/10.1038/s41586-020-03065-y, all SNP with assoication p value <1e-5 have been tested.

** all  metalite QTLs(P value <0.01) have been listed

**Table S9: Comparsion polygeneic risk score between male and female in 500FG (N=451) and 300BCG (N=313)**

| Fraction | Risk group | Male -500FG | Female -500FG | OR (95% CI) | Pvalue-500FG* | Male -300BCG | Female -300BCG | OR (95% CI) | Pvalue-300BCG | Meta-Pvalue** |
|---|---|---|---|---|---|---|---|---|---|---|
| 10% | high risk | 26 | 19 | | | 15 | 17 | | | |
| 10% | low risk | 17 | 28 | 1.496 (0.980 - 2.283) | 0.045 | 9 | 22 | 1.434 (0.894 - 2.300) | 0.115 | 0.011 |
| 15% | high risk | 32 | 35 | | | 25 | 21 | | | |
| 15% | low risk | 25 | 42 | 1.235 (0.883 - 1.727) | 0.147 | 12 | 34 | 1.770 (1.182 - 2.650) | 0.005 | 0.004 |
| 20% | high risk | 41 | 49 | | | 33 | 29 | | | |
| 20% | low risk | 34 | 56 | 1.171 (0.876 - 1.566) | 0.182 | 20 | 42 | 1.524 (1.075 - 2.163) | 0.014 | 0.010 |
| 25% | high risk | 53 | 59 | | | 41 | 37 | | | |
| 25% | low risk | 44 | 68 | 1.176 (0.907 - 1.526) | 0.140 | 28 | 50 | 1.397 (1.022 - 1.910) | 0.026 | 0.011 |
| 30% | high risk | 61 | 73 | | | 45 | 48 | | | |
| 30% | low risk | 52 | 83 | 1.154 (0.909 - 1.465) | 0.149 | 38 | 55 | 1.163 (0.874 - 1.549) | 0.188 | 0.051 |

* the p value is obtained using the fisher extact test

** the p value is obtained using the meta analyzed z score approach

**Table S10: Comparsion polygeneic risk score between male and female in 500FG (N=451) and 300BCG (N=313) (age and gender corrected model)**

| Fraction | Risk group | Male -500FG | Female -500FG | OR (95% CI) | Pvalue-500FG* | Male -300BCG | Female -300BCG | OR (95% CI) | Pvalue-300FG* | Meta-Pvalue** |
|---|---|---|---|---|---|---|---|---|---|---|
| 10% | high risk | 23 | 22 | | | 16 | 15 | | | |
| 10% | low risk | 14 | 31 | 1.498 (0.997 - 2.249) | 0.043 | 10 | 21 | 1.477 (0.903 - 2.415) | 0.099 | 0.030 |
| 15% | high risk | 32 | 35 | | | 25 | 21 | | | |
| 15% | low risk | 20 | 47 | 1.442 (1.036 - 2.006) | 0.025 | 12 | 34 | 1.770 (1.182 - 2.650) | 0.005 | 0.005 |
| 20% | high risk | 43 | 46 | | | 31 | 31 | | | |
| 20% | low risk | 32 | 58 | 1.296 (0.969 - 1.734) | 0.057 | 19 | 43 | 1.48 (1.048 - 2.090) | 0.022 | 0.008 |
| 25% | high risk | 53 | 58 | | | 43 | 35 | | | |
| 25% | low risk | 42 | 70 | 1.231 (0.948 - 1.599) | 0.079 | 30 | 48 | 1.397 (1.018 - 1.917) | 0.027 | 0.039 |
| 30% | high risk | 58 | 76 | | | 47 | 46 | | | |
| 30% | low risk | 54 | 81 | 1.070 (0.841 - 1.361) | 0.336 | 37 | 56 | 1.241 (0.932 - 1.652) | 0.092 | 0.214 |

* the p value is obtained using the fisher extact test

** the p value is obtained using the meta analyzed z score approach

**Table S11 cytokine QTLs of COVID-19 GWAS loci from the 500 FG cohort study**

| SNP | Chr | BP | Effect.Allele | Non.Effect.Allele | Beta-GWAS | Pval-GWAS | Cytokine | beta-cQTL | pvalue-CQTL* |
|---|---|---|---|---|---|---|---|---|---|
| rs36122610 | 3 | 46022833 | A | G | 0.5300752 | 4.70E-18 | IFNy_A.fumigatusconidia_PBMC_7days | 0.39099837 | 0.029802731 |
| rs13075758 | 3 | 46025048 | A | G | 0.5301634 | 4.64E-18 | IFNy_A.fumigatusconidia_PBMC_7days | 0.38959212 | 0.030390113 |
| rs34442130 | 3 | 46024529 | A | T | 0.5301222 | 4.67E-18 | IFNy_A.fumigatusconidia_PBMC_7days | 0.38961078 | 0.030391925 |
| rs17330872 | 3 | 46035097 | G | A | 0.527665 | 7.34E-18 | IFNy_A.fumigatusconidia_PBMC_7days | 0.37847035 | 0.035863852 |
| rs71615438 | 3 | 46039868 | G | A | 0.5258581 | 9.97E-18 | IFNy_A.fumigatusconidia_PBMC_7days | 0.3762894 | 0.036792968 |
| rs34754340 | 3 | 46041837 | T | C | 0.5256571 | 1.04E-17 | IFNy_A.fumigatusconidia_PBMC_7days | 0.37486826 | 0.037404892 |
| rs34836513 | 3 | 46046459 | A | G | 0.5263958 | 9.51E-18 | IFNy_A.fumigatusconidia_PBMC_7days | 0.37196481 | 0.038577233 |
| rs73833520 | 3 | 46057867 | G | A | 0.5115713 | 8.68E-17 | IFNy_A.fumigatusconidia_PBMC_7days | 0.35150761 | 0.047317806 |
| rs4493469 | 3 | 46177992 | C | T | 0.4298715 | 8.27E-11 | IFNy_C.albicansconidia_PBMC_7days | -0.41274765 | 0.009937227 |
| rs4443214 | 3 | 46177864 | C | T | 0.4456917 | 5.27E-12 | IFNy_C.albicansconidia_PBMC_7days | -0.38926029 | 0.013326893 |
| rs13063033 | 3 | 46082481 | A | G | 0.4971019 | 7.78E-16 | IFNy_C.albicansconidia_PBMC_7days | -0.33218082 | 0.026815875 |
| rs4493469 | 3 | 46177992 | C | T | 0.4298715 | 8.27E-11 | IFNy_C.conidiaHK_WB_48h | -0.55681586 | 0.003192706 |
| rs4443214 | 3 | 46177864 | C | T | 0.4456917 | 5.27E-12 | IFNy_C.conidiaHK_WB_48h | -0.53435659 | 0.003971248 |
| rs34460587 | 3 | 46142464 | T | C | 0.5045379 | 4.48E-16 | IFNy_C.conidiaHK_WB_48h | -0.48361706 | 0.009386987 |
| rs13091868 | 3 | 46123043 | G | A | 0.509687 | 2.63E-16 | IFNy_C.conidiaHK_WB_48h | -0.48145466 | 0.009664521 |
| rs13092030 | 3 | 46123055 | T | C | 0.5103595 | 2.38E-16 | IFNy_C.conidiaHK_WB_48h | -0.48145466 | 0.009664521 |
| rs17215981 | 3 | 46124418 | G | A | 0.4971931 | 1.04E-15 | IFNy_C.conidiaHK_WB_48h | -0.48145466 | 0.009664521 |
| rs13089554 | 3 | 46127557 | A | G | 0.4966395 | 1.13E-15 | IFNy_C.conidiaHK_WB_48h | -0.48145466 | 0.009664521 |
| rs13095717 | 3 | 46128542 | A | G | 0.4967673 | 1.11E-15 | IFNy_C.conidiaHK_WB_48h | -0.48145466 | 0.009664521 |
| rs13075528 | 3 | 46128709 | C | T | 0.5001448 | 7.62E-16 | IFNy_C.conidiaHK_WB_48h | -0.48145466 | 0.009664521 |
| rs13095602 | 3 | 46128713 | G | A | 0.4997202 | 8.04E-16 | IFNy_C.conidiaHK_WB_48h | -0.48145466 | 0.009664521 |
| rs71327023 | 3 | 46131225 | G | C | 0.49419 | 1.69E-15 | IFNy_C.conidiaHK_WB_48h | -0.48145466 | 0.009664521 |
| rs13063527 | 3 | 46135690 | A | G | 0.4983724 | 9.69E-16 | IFNy_C.conidiaHK_WB_48h | -0.48145466 | 0.009664521 |
| rs4373103 | 3 | 46138455 | A | G | 0.4974729 | 1.14E-15 | IFNy_C.conidiaHK_WB_48h | -0.48145466 | 0.009664521 |
| rs71327024 | 3 | 46140073 | T | G | 0.5013388 | 6.45E-16 | IFNy_C.conidiaHK_WB_48h | -0.48145466 | 0.009664521 |
| rs71327017 | 3 | 46111213 | C | T | 0.4985668 | 8.96E-16 | IFNy_C.conidiaHK_WB_48h | -0.48139108 | 0.009675192 |
| rs9845382 | 3 | 46111759 | A | G | 0.5066369 | 2.03E-16 | IFNy_C.conidiaHK_WB_48h | -0.48139108 | 0.009675192 |
| rs13069742 | 3 | 46114216 | G | A | 0.5064079 | 2.08E-16 | IFNy_C.conidiaHK_WB_48h | -0.48139108 | 0.009675192 |
| rs13072267 | 3 | 46116010 | A | T | 0.4979167 | 9.53E-16 | IFNy_C.conidiaHK_WB_48h | -0.48139108 | 0.009675192 |
| rs36057789 | 3 | 46117096 | G | A | 0.4975033 | 1.01E-15 | IFNy_C.conidiaHK_WB_48h | -0.48139108 | 0.009675192 |
| rs76647202 | 3 | 46117960 | G | A | 0.4978073 | 9.57E-16 | IFNy_C.conidiaHK_WB_48h | -0.48139108 | 0.009675192 |
| rs71327021 | 3 | 46120517 | G | A | 0.4978316 | 9.43E-16 | IFNy_C.conidiaHK_WB_48h | -0.48139108 | 0.009675192 |
| rs9824651 | 3 | 46108285 | T | C | 0.5120269 | 9.50E-17 | IFNy_C.conidiaHK_WB_48h | -0.48132626 | 0.009685968 |
| rs9825081 | 3 | 46108400 | A | G | 0.5118771 | 9.67E-17 | IFNy_C.conidiaHK_WB_48h | -0.48132626 | 0.009685968 |
| rs71327014 | 3 | 46101466 | T | C | 0.5076245 | 2.27E-16 | IFNy_C.conidiaHK_WB_48h | -0.47171713 | 0.011749086 |
| rs71327015 | 3 | 46102173 | C | G | 0.5043145 | 4.35E-16 | IFNy_C.conidiaHK_WB_48h | -0.47171713 | 0.011749086 |
| rs34766614 | 3 | 46107601 | G | A | 0.500351 | 7.64E-16 | IFNy_C.conidiaHK_WB_48h | -0.4716423 | 0.011760739 |
| rs34127208 | 3 | 46103680 | T | G | 0.5047371 | 4.15E-16 | IFNy_C.conidiaHK_WB_48h | -0.47157349 | 0.011773915 |
| rs34047915 | 3 | 46100491 | T | C | 0.5072994 | 2.36E-16 | IFNy_C.conidiaHK_WB_48h | -0.47138713 | 0.011808996 |
| rs35420565 | 3 | 46100138 | G | C | 0.5074198 | 2.32E-16 | IFNy_C.conidiaHK_WB_48h | -0.47105588 | 0.011869289 |
| rs13096741 | 3 | 46099154 | T | C | 0.5072512 | 2.37E-16 | IFNy_C.conidiaHK_WB_48h | -0.46971846 | 0.012114335 |
| rs34718164 | 3 | 46098526 | G | C | 0.5071188 | 2.42E-16 | IFNy_C.conidiaHK_WB_48h | -0.46904233 | 0.012239216 |
| rs77399277 | 3 | 46096798 | C | T | 0.509008 | 1.80E-16 | IFNy_C.conidiaHK_WB_48h | -0.46663733 | 0.012689 |
| rs35772789 | 3 | 46095104 | G | A | 0.5079254 | 2.08E-16 | IFNy_C.conidiaHK_WB_48h | -0.46502402 | 0.012995844 |
| rs34863575 | 3 | 46094063 | G | A | 0.5076787 | 2.15E-16 | IFNy_C.conidiaHK_WB_48h | -0.46359907 | 0.013270024 |
| rs34679077 | 3 | 46087992 | A | G | 0.5051656 | 2.90E-16 | IFNy_C.conidiaHK_WB_48h | -0.45695531 | 0.014593042 |
| rs72901034 | 3 | 46085321 | G | A | 0.4961586 | 1.34E-17 | IFNy_C.conidiaHK_WB_48h | -0.45511623 | 0.014973728 |
| rs9877748 | 3 | 46111081 | G | A | 0.5070104 | 1.95E-16 | IFNy_C.conidiaHK_WB_48h | -0.46184539 | 0.014994632 |
| rs34438204 | 3 | 46081306 | C | T | 0.504556 | 3.11E-16 | IFNy_C.conidiaHK_WB_48h | -0.45278628 | 0.015463645 |
| rs2088692 | 3 | 46077332 | G | A | 0.5114274 | 8.77E-17 | IFNy_C.conidiaHK_WB_48h | -0.45001851 | 0.016058812 |
| rs6791016 | 3 | 46074858 | A | G | 0.5114394 | 8.76E-17 | IFNy_C.conidiaHK_WB_48h | -0.448882 | 0.016304713 |
| rs6780028 | 3 | 46075090 | C | A | 0.5114934 | 8.69E-17 | IFNy_C.conidiaHK_WB_48h | -0.448882 | 0.016304713 |
| rs6790866 | 3 | 46074711 | A | G | 0.5114514 | 8.74E-17 | IFNy_C.conidiaHK_WB_48h | -0.44847913 | 0.01639342 |
| rs35814488 | 3 | 46073923 | G | A | 0.5114394 | 8.75E-17 | IFNy_C.conidiaHK_WB_48h | -0.44807521 | 0.016482677 |
| rs34774687 | 3 | 46073314 | C | T | 0.5114574 | 8.73E-17 | IFNy_C.conidiaHK_WB_48h | -0.44767025 | 0.016572486 |
| rs34619093 | 3 | 46072519 | A | G | 0.5041695 | 3.25E-16 | IFNy_C.conidiaHK_WB_48h | -0.44726425 | 0.016662853 |
| rs68087193 | 3 | 46072630 | T | C | 0.5113135 | 8.97E-17 | IFNy_C.conidiaHK_WB_48h | -0.44726425 | 0.016662853 |
| rs2102055 | 3 | 46072704 | A | G | 0.5140265 | 4.75E-17 | IFNy_C.conidiaHK_WB_48h | -0.44726425 | 0.016662853 |
| rs2102056 | 3 | 46072768 | A | G | 0.5114095 | 8.83E-17 | IFNy_C.conidiaHK_WB_48h | -0.44726425 | 0.016662853 |
| rs2088690 | 3 | 46072839 | T | C | 0.5114095 | 8.83E-17 | IFNy_C.conidiaHK_WB_48h | -0.44726425 | 0.016662853 |
| rs13060287 | 3 | 46071868 | G | A | 0.5111396 | 9.18E-17 | IFNy_C.conidiaHK_WB_48h | -0.44692768 | 0.016738401 |
| rs13082697 | 3 | 46071763 | C | T | 0.5096029 | 1.21E-16 | IFNy_C.conidiaHK_WB_48h | -0.44651958 | 0.016829817 |
| rs34562820 | 3 | 46071081 | A | G | 0.5041333 | 3.27E-16 | IFNy_C.conidiaHK_WB_48h | -0.44611045 | 0.016921799 |
| rs59166269 | 3 | 46070337 | G | C | 0.5113075 | 8.94E-17 | IFNy_C.conidiaHK_WB_48h | -0.44570029 | 0.017014349 |
| rs13090194 | 3 | 46069573 | C | A | 0.5115833 | 8.64E-17 | IFNy_C.conidiaHK_WB_48h | -0.44528911 | 0.017107471 |
| rs71327010 | 3 | 46068835 | T | G | 0.5039037 | 3.39E-16 | IFNy_C.conidiaHK_WB_48h | -0.44446369 | 0.017295441 |
| rs35159820 | 3 | 46068473 | G | A | 0.5112475 | 9.02E-17 | IFNy_C.conidiaHK_WB_48h | -0.44404946 | 0.017390295 |
| rs35161099 | 3 | 46067507 | G | T | 0.5040789 | 3.29E-16 | IFNy_C.conidiaHK_WB_48h | -0.44363422 | 0.017485734 |
| rs35930050 | 3 | 46065763 | A | G | 0.5113015 | 8.93E-17 | IFNy_C.conidiaHK_WB_48h | -0.44196325 | 0.017873394 |
| rs34584867 | 3 | 46065963 | A | G | 0.5112955 | 8.94E-17 | IFNy_C.conidiaHK_WB_48h | -0.44196325 | 0.017873394 |
| rs876668 | 3 | 46063753 | C | T | 0.5112415 | 9.01E-17 | IFNy_C.conidiaHK_WB_48h | -0.44027644 | 0.018270658 |
| rs13097556 | 3 | 46061997 | C | T | 0.5096389 | 1.23E-16 | IFNy_C.conidiaHK_WB_48h | -0.43948996 | 0.018569874 |
| rs36040135 | 3 | 46061304 | G | A | 0.5113495 | 8.88E-17 | IFNy_C.conidiaHK_WB_48h | -0.43896156 | 0.018762646 |
| rs35334665 | 3 | 46061218 | C | T | 0.5042903 | 3.19E-16 | IFNy_C.conidiaHK_WB_48h | -0.43888036 | 0.018795265 |
| rs1491951 | 3 | 46141844 | A | G | 0.5083165 | 2.07E-16 | IFNy_C.conidiaHK_WB_48h | -0.43215648 | 0.018946295 |
| rs35454877 | 3 | 46059484 | C | T | 0.5121288 | 7.99E-17 | IFNy_C.conidiaHK_WB_48h | -0.43766056 | 0.019225591 |
| rs71327007 | 3 | 46059249 | T | C | 0.5044473 | 3.14E-16 | IFNy_C.conidiaHK_WB_48h | -0.43760504 | 0.019247772 |
| rs71327006 | 3 | 46058908 | A | G | 0.5044715 | 3.14E-16 | IFNy_C.conidiaHK_WB_48h | -0.43745066 | 0.019305828 |
| rs73833520 | 3 | 46057867 | G | A | 0.5115713 | 8.68E-17 | IFNy_C.conidiaHK_WB_48h | -0.4311573 | 0.022155515 |
| rs2230322 | 3 | 46063329 | C | T | 0.5110856 | 9.20E-17 | IFNy_C.conidiaHK_WB_48h | -0.42174466 | 0.022767282 |
| rs13063033 | 3 | 46082481 | A | G | 0.4971019 | 7.78E-16 | IFNy_C.conidiaHK_WB_48h | -0.40457718 | 0.022768665 |
| rs35751180 | 3 | 46056173 | T | C | 0.5342276 | 4.95E-18 | IFNy_C.conidiaHK_WB_48h | -0.41391963 | 0.030142942 |
| rs4362758 | 3 | 46056911 | A | T | 0.5157296 | 5.39E-17 | IFNy_C.conidiaHK_WB_48h | -0.41395242 | 0.03017128 |
| rs34168660 | 3 | 46056162 | A | G | 0.5435793 | 1.04E-18 | IFNy_C.conidiaHK_WB_48h | -0.41297388 | 0.030603419 |
| rs13081213 | 3 | 46055909 | T | C | 0.5281664 | 6.72E-18 | IFNy_C.conidiaHK_WB_48h | -0.41277992 | 0.03070245 |
| rs35669129 | 3 | 46055250 | A | G | 0.5281192 | 6.78E-18 | IFNy_C.conidiaHK_WB_48h | -0.41253221 | 0.030837033 |
| rs34442130 | 3 | 46024529 | A | T | 0.5301222 | 4.67E-18 | IFNy_C.conidiaHK_WB_48h | -0.40744003 | 0.033470162 |
| rs13075758 | 3 | 46025048 | A | G | 0.5301634 | 4.64E-18 | IFNy_C.conidiaHK_WB_48h | -0.40703136 | 0.033636251 |
| rs36122610 | 3 | 46022833 | A | G | 0.5300752 | 4.70E-18 | IFNy_C.conidiaHK_WB_48h | -0.40685908 | 0.033736384 |
| rs17330872 | 3 | 46035097 | G | A | 0.527665 | 7.34E-18 | IFNy_C.conidiaHK_WB_48h | -0.40626762 | 0.034212109 |
| rs71615438 | 3 | 46039868 | G | A | 0.5258581 | 9.97E-18 | IFNy_C.conidiaHK_WB_48h | -0.40473603 | 0.034732858 |
| rs34754340 | 3 | 46041837 | T | C | 0.5256571 | 1.04E-17 | IFNy_C.conidiaHK_WB_48h | -0.40378501 | 0.035052131 |
| rs34836513 | 3 | 46046459 | A | G | 0.5263958 | 9.51E-18 | IFNy_C.conidiaHK_WB_48h | -0.40177027 | 0.03564304 |
| rs72856706 | 6 | 31120729 | T | C | 0.6148018 | 9.15E-18 | IFNy_Cryptococcus_PBMC_7days | 0.30023853 | 0.049256492 |
| rs111837807 | 6 | 31121232 | C | T | 0.6149802 | 8.95E-18 | IFNy_Cryptococcus_PBMC_7days | 0.30023853 | 0.049256492 |
| rs112785163 | 6 | 31121381 | C | G | 0.6149316 | 9.02E-18 | IFNy_Cryptococcus_PBMC_7days | 0.30023853 | 0.049256492 |
| rs143334143 | 6 | 31121426 | A | G | 0.6150829 | 8.82E-18 | IFNy_Cryptococcus_PBMC_7days | 0.30023853 | 0.049256492 |
| rs72856713 | 6 | 31121958 | G | C | 0.5882588 | 2.84E-16 | IFNy_Cryptococcus_PBMC_7days | 0.30023853 | 0.049256492 |
| rs2109069 | 19 | 4719443 | A | G | 0.3056227 | 3.98E-12 | IFNy_S.aureus_PBMC_7days | 0.31320494 | 0.006636505 |
| rs72856706 | 6 | 31120729 | T | C | 0.6148018 | 9.15E-18 | IFNy_S.aureus_WB_48h | 0.55667481 | 0.003251067 |
| rs111837807 | 6 | 31121232 | C | T | 0.6149802 | 8.95E-18 | IFNy_S.aureus_WB_48h | 0.55667481 | 0.003251067 |
| rs112785163 | 6 | 31121381 | C | G | 0.6149316 | 9.02E-18 | IFNy_S.aureus_WB_48h | 0.55667481 | 0.003251067 |
| rs143334143 | 6 | 31121426 | A | G | 0.6150829 | 8.82E-18 | IFNy_S.aureus_WB_48h | 0.55667481 | 0.003251067 |
| rs72856713 | 6 | 31121958 | G | C | 0.5882588 | 2.84E-16 | IFNy_S.aureus_WB_48h | 0.55667481 | 0.003251067 |
| rs2109069 | 19 | 4719443 | A | G | 0.3056227 | 3.98E-12 | IFNy_S.aureus_WB_48h | 0.26127968 | 0.034291507 |
| rs35951367 | 3 | 46118439 | C | T | 0.3240092 | 5.94E-09 | IL.10_Pam3Cys_PBMC_24h | -0.10436546 | 0.034697306 |
| rs9845542 | 3 | 46111869 | A | G | 0.3328729 | 1.75E-09 | IL.10_Pam3Cys_PBMC_24h | -0.10444674 | 0.034611518 |
| rs13060287 | 3 | 46071868 | G | A | 0.5111396 | 9.18E-17 | IL.1Ra_C.albicans.yeast_PBMC_24h | -0.27194779 | 0.0181121 |
| rs13063527 | 3 | 46135690 | A | G | 0.4983724 | 9.69E-16 | IL.1Ra_C.albicans.yeast_PBMC_24h | -0.25888106 | 0.020579792 |
| rs13069742 | 3 | 46114216 | G | A | 0.5064079 | 2.08E-16 | IL.1Ra_C.albicans.yeast_PBMC_24h | -0.25888106 | 0.020579792 |
| rs13072267 | 3 | 46116010 | A | T | 0.4979167 | 9.53E-16 | IL.1Ra_C.albicans.yeast_PBMC_24h | -0.25888106 | 0.020579792 |
| rs13075528 | 3 | 46128709 | C | T | 0.5001448 | 7.62E-16 | IL.1Ra_C.albicans.yeast_PBMC_24h | -0.25888106 | 0.020579792 |
| rs13075758 | 3 | 46025048 | A | G | 0.5301634 | 4.64E-18 | IL.1Ra_C.albicans.yeast_PBMC_24h | -0.25629322 | 0.034237811 |
| rs13082697 | 3 | 46071763 | C | T | 0.5096029 | 1.21E-16 | IL.1Ra_C.albicans.yeast_PBMC_24h | -0.27194779 | 0.0181121 |
| rs13089554 | 3 | 46127557 | A | G | 0.4966395 | 1.13E-15 | IL.1Ra_C.albicans.yeast_PBMC_24h | -0.25888106 | 0.020579792 |
| rs13090194 | 3 | 46069573 | C | A | 0.5115833 | 8.64E-17 | IL.1Ra_C.albicans.yeast_PBMC_24h | -0.27194779 | 0.0181121 |
| rs13091868 | 3 | 46123043 | G | A | 0.509687 | 2.63E-16 | IL.1Ra_C.albicans.yeast_PBMC_24h | -0.25888106 | 0.020579792 |
| rs13092030 | 3 | 46123055 | T | C | 0.5103595 | 2.38E-16 | IL.1Ra_C.albicans.yeast_PBMC_24h | -0.25888106 | 0.020579792 |
| rs13095602 | 3 | 46128713 | G | A | 0.4997202 | 8.04E-16 | IL.1Ra_C.albicans.yeast_PBMC_24h | -0.25888106 | 0.020579792 |
| rs13095717 | 3 | 46128542 | A | G | 0.4967673 | 1.11E-15 | IL.1Ra_C.albicans.yeast_PBMC_24h | -0.25888106 | 0.020579792 |
| rs13096741 | 3 | 46099154 | T | C | 0.5072512 | 2.37E-16 | IL.1Ra_C.albicans.yeast_PBMC_24h | -0.27194779 | 0.0181121 |
| rs13097556 | 3 | 46061997 | C | T | 0.5096389 | 1.23E-16 | IL.1Ra_C.albicans.yeast_PBMC_24h | -0.27194779 | 0.0181121 |
| rs1491951 | 3 | 46141844 | A | G | 0.5083165 | 2.07E-16 | IL.1Ra_C.albicans.yeast_PBMC_24h | -0.25888106 | 0.020579792 |
| rs17215981 | 3 | 46124418 | G | A | 0.4971931 | 1.04E-15 | IL.1Ra_C.albicans.yeast_PBMC_24h | -0.25888106 | 0.020579792 |
| rs17330872 | 3 | 46035097 | G | A | 0.527665 | 7.34E-18 | IL.1Ra_C.albicans.yeast_PBMC_24h | -0.23586863 | 0.049682435 |
| rs2088690 | 3 | 46072839 | T | C | 0.5114095 | 8.83E-17 | IL.1Ra_C.albicans.yeast_PBMC_24h | -0.27194779 | 0.0181121 |
| rs2088692 | 3 | 46077332 | G | A | 0.5114274 | 8.77E-17 | IL.1Ra_C.albicans.yeast_PBMC_24h | -0.27194779 | 0.0181121 |
| rs2102055 | 3 | 46072704 | A | G | 0.5140265 | 4.75E-17 | IL.1Ra_C.albicans.yeast_PBMC_24h | -0.27194779 | 0.0181121 |
| rs2102056 | 3 | 46072768 | A | G | 0.5114095 | 8.83E-17 | IL.1Ra_C.albicans.yeast_PBMC_24h | -0.27194779 | 0.0181121 |
| rs2230322 | 3 | 46063329 | C | T | 0.5110856 | 9.20E-17 | IL.1Ra_C.albicans.yeast_PBMC_24h | -0.27194779 | 0.0181121 |
| rs34047915 | 3 | 46100491 | T | C | 0.5072994 | 2.36E-16 | IL.1Ra_C.albicans.yeast_PBMC_24h | -0.27194779 | 0.0181121 |
| rs34127208 | 3 | 46103680 | T | G | 0.5047371 | 4.15E-16 | IL.1Ra_C.albicans.yeast_PBMC_24h | -0.27194779 | 0.0181121 |
| rs34438204 | 3 | 46081306 | C | T | 0.504556 | 3.11E-16 | IL.1Ra_C.albicans.yeast_PBMC_24h | -0.27194779 | 0.0181121 |
| rs34442130 | 3 | 46024529 | A | T | 0.5301222 | 4.67E-18 | IL.1Ra_C.albicans.yeast_PBMC_24h | -0.25629322 | 0.034237811 |
| rs34460587 | 3 | 46142464 | T | C | 0.5045379 | 4.48E-16 | IL.1Ra_C.albicans.yeast_PBMC_24h | -0.2580695 | 0.021079213 |
| rs34562820 | 3 | 46071081 | A | G | 0.5041333 | 3.27E-16 | IL.1Ra_C.albicans.yeast_PBMC_24h | -0.27194779 | 0.0181121 |
| rs34584867 | 3 | 46065963 | A | G | 0.5112955 | 8.94E-17 | IL.1Ra_C.albicans.yeast_PBMC_24h | -0.27194779 | 0.0181121 |
| rs34619093 | 3 | 46072519 | A | G | 0.5041695 | 3.25E-16 | IL.1Ra_C.albicans.yeast_PBMC_24h | -0.27194779 | 0.0181121 |
| rs34679077 | 3 | 46087992 | A | G | 0.5051656 | 2.90E-16 | IL.1Ra_C.albicans.yeast_PBMC_24h | -0.27194779 | 0.0181121 |
| rs34718164 | 3 | 46098526 | G | C | 0.5071188 | 2.42E-16 | IL.1Ra_C.albicans.yeast_PBMC_24h | -0.27194779 | 0.0181121 |
| rs34766614 | 3 | 46107601 | G | A | 0.500351 | 7.64E-16 | IL.1Ra_C.albicans.yeast_PBMC_24h | -0.27194779 | 0.0181121 |
| rs34774687 | 3 | 46073314 | C | T | 0.5114574 | 8.73E-17 | IL.1Ra_C.albicans.yeast_PBMC_24h | -0.27194779 | 0.0181121 |
| rs34863575 | 3 | 46094063 | G | A | 0.5076787 | 2.15E-16 | IL.1Ra_C.albicans.yeast_PBMC_24h | -0.27194779 | 0.0181121 |
| rs35159820 | 3 | 46068473 | G | A | 0.5112475 | 9.02E-17 | IL.1Ra_C.albicans.yeast_PBMC_24h | -0.27194779 | 0.0181121 |
| rs35161099 | 3 | 46067507 | G | T | 0.5040789 | 3.29E-16 | IL.1Ra_C.albicans.yeast_PBMC_24h | -0.27194779 | 0.0181121 |
| rs35334665 | 3 | 46061218 | C | T | 0.5042903 | 3.19E-16 | IL.1Ra_C.albicans.yeast_PBMC_24h | -0.27194779 | 0.0181121 |
| rs35420565 | 3 | 46100138 | G | C | 0.5074198 | 2.32E-16 | IL.1Ra_C.albicans.yeast_PBMC_24h | -0.27194779 | 0.0181121 |
| rs35454877 | 3 | 46059484 | C | T | 0.5121288 | 7.99E-17 | IL.1Ra_C.albicans.yeast_PBMC_24h | -0.27194779 | 0.0181121 |
| rs35772789 | 3 | 46095104 | G | A | 0.5079254 | 2.08E-16 | IL.1Ra_C.albicans.yeast_PBMC_24h | -0.27194779 | 0.0181121 |
| rs35814488 | 3 | 46073923 | G | A | 0.5114394 | 8.75E-17 | IL.1Ra_C.albicans.yeast_PBMC_24h | -0.27194779 | 0.0181121 |
| rs35930050 | 3 | 46065763 | A | G | 0.5113015 | 8.93E-17 | IL.1Ra_C.albicans.yeast_PBMC_24h | -0.27194779 | 0.0181121 |
| rs35951367 | 3 | 46118439 | C | T | 0.3240092 | 5.94E-09 | IL.1Ra_C.albicans.yeast_PBMC_24h | -0.2320228 | 0.02438599 |
| rs36040135 | 3 | 46061304 | G | A | 0.5113495 | 8.88E-17 | IL.1Ra_C.albicans.yeast_PBMC_24h | -0.27194779 | 0.0181121 |
| rs36057789 | 3 | 46117096 | G | A | 0.4975033 | 1.01E-15 | IL.1Ra_C.albicans.yeast_PBMC_24h | -0.25888106 | 0.020579792 |
| rs36122610 | 3 | 46022833 | A | G | 0.5300752 | 4.70E-18 | IL.1Ra_C.albicans.yeast_PBMC_24h | -0.25629322 | 0.034237811 |
| rs4373103 | 3 | 46138455 | A | G | 0.4974729 | 1.14E-15 | IL.1Ra_C.albicans.yeast_PBMC_24h | -0.25888106 | 0.020579792 |
| rs59166269 | 3 | 46070337 | G | C | 0.5113075 | 8.94E-17 | IL.1Ra_C.albicans.yeast_PBMC_24h | -0.27194779 | 0.0181121 |
| rs6780028 | 3 | 46075090 | C | A | 0.5114934 | 8.69E-17 | IL.1Ra_C.albicans.yeast_PBMC_24h | -0.27194779 | 0.0181121 |
| rs6790866 | 3 | 46074711 | A | G | 0.5114514 | 8.74E-17 | IL.1Ra_C.albicans.yeast_PBMC_24h | -0.27194779 | 0.0181121 |
| rs6791016 | 3 | 46074858 | A | G | 0.5114394 | 8.76E-17 | IL.1Ra_C.albicans.yeast_PBMC_24h | -0.27194779 | 0.0181121 |
| rs68087193 | 3 | 46072630 | T | C | 0.5113135 | 8.97E-17 | IL.1Ra_C.albicans.yeast_PBMC_24h | -0.27194779 | 0.0181121 |
| rs71327006 | 3 | 46058908 | A | G | 0.5044715 | 3.14E-16 | IL.1Ra_C.albicans.yeast_PBMC_24h | -0.27194779 | 0.0181121 |
| rs71327007 | 3 | 46059249 | T | C | 0.5044473 | 3.14E-16 | IL.1Ra_C.albicans.yeast_PBMC_24h | -0.27194779 | 0.0181121 |
| rs71327010 | 3 | 46068835 | T | G | 0.5039037 | 3.39E-16 | IL.1Ra_C.albicans.yeast_PBMC_24h | -0.27194779 | 0.0181121 |
| rs71327014 | 3 | 46101466 | T | C | 0.5076245 | 2.27E-16 | IL.1Ra_C.albicans.yeast_PBMC_24h | -0.27194779 | 0.0181121 |
| rs71327015 | 3 | 46102173 | C | G | 0.5043145 | 4.35E-16 | IL.1Ra_C.albicans.yeast_PBMC_24h | -0.27194779 | 0.0181121 |
| rs71327017 | 3 | 46111213 | C | T | 0.4985668 | 8.96E-16 | IL.1Ra_C.albicans.yeast_PBMC_24h | -0.25888106 | 0.020579792 |
| rs71327021 | 3 | 46120517 | G | A | 0.4978316 | 9.43E-16 | IL.1Ra_C.albicans.yeast_PBMC_24h | -0.25888106 | 0.020579792 |
| rs71327023 | 3 | 46131225 | G | C | 0.49419 | 1.69E-15 | IL.1Ra_C.albicans.yeast_PBMC_24h | -0.25888106 | 0.020579792 |
| rs71327024 | 3 | 46140073 | T | G | 0.5013388 | 6.45E-16 | IL.1Ra_C.albicans.yeast_PBMC_24h | -0.25888106 | 0.020579792 |
| rs72901034 | 3 | 46085321 | G | A | 0.4961586 | 1.34E-17 | IL.1Ra_C.albicans.yeast_PBMC_24h | -0.27194779 | 0.0181121 |
| rs73833520 | 3 | 46057867 | G | A | 0.5115713 | 8.68E-17 | IL.1Ra_C.albicans.yeast_PBMC_24h | -0.27194779 | 0.0181121 |
| rs76647202 | 3 | 46117960 | G | A | 0.4978073 | 9.57E-16 | IL.1Ra_C.albicans.yeast_PBMC_24h | -0.25888106 | 0.020579792 |
| rs77399277 | 3 | 46096798 | C | T | 0.509008 | 1.80E-16 | IL.1Ra_C.albicans.yeast_PBMC_24h | -0.27194779 | 0.0181121 |
| rs876668 | 3 | 46063753 | C | T | 0.5112415 | 9.01E-17 | IL.1Ra_C.albicans.yeast_PBMC_24h | -0.27194779 | 0.0181121 |
| rs9824651 | 3 | 46108285 | T | C | 0.5120269 | 9.50E-17 | IL.1Ra_C.albicans.yeast_PBMC_24h | -0.25888106 | 0.020579792 |
| rs9825081 | 3 | 46108400 | A | G | 0.5118771 | 9.67E-17 | IL.1Ra_C.albicans.yeast_PBMC_24h | -0.25888106 | 0.020579792 |
| rs9838450 | 3 | 46123333 | C | A | 0.3461396 | 7.25E-10 | IL.1Ra_C.albicans.yeast_PBMC_24h | -0.30817684 | 0.003925949 |
| rs9845382 | 3 | 46111759 | A | G | 0.5066369 | 2.03E-16 | IL.1Ra_C.albicans.yeast_PBMC_24h | -0.25888106 | 0.020579792 |
| rs9845542 | 3 | 46111869 | A | G | 0.3328729 | 1.75E-09 | IL.1Ra_C.albicans.yeast_PBMC_24h | -0.2320228 | 0.02438599 |
| rs7623190 | 3 | 46068325 | C | T | 0.2556107 | 8.91E-09 | IL.1Ra_LPS.100ng_PBMC_24h | 0.09526084 | 0.04950986 |
| rs9849015 | 3 | 46056120 | C | T | 0.2579159 | 6.65E-09 | IL.1Ra_LPS.100ng_PBMC_24h | 0.09488469 | 0.049852063 |
| rs13060901 | 3 | 46072006 | A | G | 0.2564932 | 7.93E-09 | IL.8_Pam3Cys_PBMC_24h | 0.04079613 | 0.033786104 |
| rs1392290 | 3 | 46069210 | A | G | 0.2486631 | 2.01E-08 | IL.8_Pam3Cys_PBMC_24h | 0.04081239 | 0.033688988 |
| rs4411920 | 3 | 46077525 | G | A | 0.2565551 | 7.89E-09 | IL.8_Pam3Cys_PBMC_24h | 0.04075256 | 0.034018189 |
| rs6764042 | 3 | 46059139 | G | T | 0.2550607 | 9.68E-09 | IL.8_Pam3Cys_PBMC_24h | 0.04084761 | 0.0333915 |
| rs6805094 | 3 | 46071463 | G | C | 0.25647 | 7.95E-09 | IL.8_Pam3Cys_PBMC_24h | 0.04079897 | 0.033769608 |
| rs7623190 | 3 | 46068325 | C | T | 0.2556107 | 8.91E-09 | IL.8_Pam3Cys_PBMC_24h | 0.04475807 | 0.020464932 |
| rs7623460 | 3 | 46068764 | C | A | 0.2603006 | 4.93E-09 | IL.8_Pam3Cys_PBMC_24h | 0.04081491 | 0.033673234 |
| rs7642701 | 3 | 46091058 | G | A | 0.2750678 | 9.82E-10 | IL.8_Pam3Cys_PBMC_24h | 0.03895841 | 0.049890552 |
| rs9849015 | 3 | 46056120 | C | T | 0.2579159 | 6.65E-09 | IL.8_Pam3Cys_PBMC_24h | 0.04412251 | 0.02254586 |
| rs111837807 | 6 | 31121232 | C | T | 0.6149802 | 8.95E-18 | IL.8_S.aureus_PBMC_24h | -0.10869608 | 0.032969688 |
| rs112785163 | 6 | 31121381 | C | G | 0.6149316 | 9.02E-18 | IL.8_S.aureus_PBMC_24h | -0.10869608 | 0.032969688 |
| rs143334143 | 6 | 31121426 | A | G | 0.6150829 | 8.82E-18 | IL.8_S.aureus_PBMC_24h | -0.10869608 | 0.032969688 |
| rs72856706 | 6 | 31120729 | T | C | 0.6148018 | 9.15E-18 | IL.8_S.aureus_PBMC_24h | -0.10869608 | 0.032969688 |
| rs72856713 | 6 | 31121958 | G | C | 0.5882588 | 2.84E-16 | IL.8_S.aureus_PBMC_24h | -0.10869608 | 0.032969688 |
| rs3851347 | 3 | 46053778 | G | A | 0.2605395 | 4.30E-09 | IL17_MTB_PBMC_7days | 0.19920249 | 0.046475334 |

| | | | | | | | | | |
|---|---|---|---|---|---|---|---|---|---|
| rs9849015 | 3 | 46056120 | C | T | 0.2579159 | 6.65E-09 | IL17_MTB_PBMC_7days | 0.19994132 | 0.046817327 |
| rs72856706 | 6 | 31120729 | T | C | 0.6148018 | 9.15E-18 | IL1b_C.albicansconidia_PBMC_24h | -0.15502224 | 0.003711668 |
| rs111837807 | 6 | 31121232 | C | T | 0.6149802 | 8.95E-18 | IL1b_C.albicansconidia_PBMC_24h | -0.15502224 | 0.003711668 |
| rs112785163 | 6 | 31121381 | C | G | 0.6149316 | 9.02E-18 | IL1b_C.albicansconidia_PBMC_24h | -0.15502224 | 0.003711668 |
| rs143334143 | 6 | 31121426 | A | G | 0.6150829 | 8.82E-18 | IL1b_C.albicansconidia_PBMC_24h | -0.15502224 | 0.003711668 |
| rs72856713 | 6 | 31121958 | G | C | 0.5882588 | 2.84E-16 | IL1b_C.albicansconidia_PBMC_24h | -0.15502224 | 0.003711668 |
| rs34836513 | 3 | 46046459 | A | G | 0.5263958 | 9.51E-18 | IL1b_C.conidiaHK_WB_48h | -0.29446261 | 0.016379284 |
| rs35751180 | 3 | 46056173 | T | C | 0.5342276 | 4.95E-18 | IL1b_C.conidiaHK_WB_48h | -0.29251118 | 0.016939929 |
| rs35669129 | 3 | 46055250 | A | G | 0.5281192 | 6.78E-18 | IL1b_C.conidiaHK_WB_48h | -0.29272534 | 0.016946961 |
| rs13081213 | 3 | 46055909 | T | C | 0.5281664 | 6.72E-18 | IL1b_C.conidiaHK_WB_48h | -0.29218472 | 0.017130475 |
| rs34168660 | 3 | 46056162 | A | G | 0.5435793 | 1.04E-18 | IL1b_C.conidiaHK_WB_48h | -0.29206197 | 0.017163978 |
| rs4362758 | 3 | 46056911 | A | T | 0.5157296 | 5.39E-17 | IL1b_C.conidiaHK_WB_48h | -0.29192386 | 0.017191782 |
| rs34754340 | 3 | 46041837 | T | C | 0.5256571 | 1.04E-17 | IL1b_C.conidiaHK_WB_48h | -0.29160856 | 0.017662277 |
| rs71615438 | 3 | 46039868 | G | A | 0.5258581 | 9.97E-18 | IL1b_C.conidiaHK_WB_48h | -0.2903857 | 0.018217781 |
| rs17330872 | 3 | 46035097 | G | A | 0.527665 | 7.34E-18 | IL1b_C.conidiaHK_WB_48h | -0.28680395 | 0.019821308 |
| rs73833520 | 3 | 46057867 | G | A | 0.5115713 | 8.68E-17 | IL1b_C.conidiaHK_WB_48h | -0.28039728 | 0.020478572 |
| rs71327006 | 3 | 46058908 | A | G | 0.5044715 | 3.14E-16 | IL1b_C.conidiaHK_WB_48h | -0.27326746 | 0.022850761 |
| rs71327007 | 3 | 46059249 | T | C | 0.5044473 | 3.14E-16 | IL1b_C.conidiaHK_WB_48h | -0.27302983 | 0.022953129 |
| rs35454877 | 3 | 46059484 | C | T | 0.5121288 | 7.99E-17 | IL1b_C.conidiaHK_WB_48h | -0.2728643 | 0.023028959 |
| rs35334665 | 3 | 46061218 | C | T | 0.5042903 | 3.19E-16 | IL1b_C.conidiaHK_WB_48h | -0.27126509 | 0.023737813 |
| rs36040135 | 3 | 46061304 | G | A | 0.5113495 | 8.88E-17 | IL1b_C.conidiaHK_WB_48h | -0.27118279 | 0.023768124 |
| rs9877748 | 3 | 46111081 | G | A | 0.5070104 | 1.95E-16 | IL1b_C.conidiaHK_WB_48h | -0.27552231 | 0.023876624 |
| rs13097556 | 3 | 46061997 | C | T | 0.5096389 | 1.23E-16 | IL1b_C.conidiaHK_WB_48h | -0.27054774 | 0.024040586 |
| rs876668 | 3 | 46063753 | C | T | 0.5112415 | 9.01E-17 | IL1b_C.conidiaHK_WB_48h | -0.26920126 | 0.024647522 |
| rs35930050 | 3 | 46065763 | A | G | 0.5113015 | 8.93E-17 | IL1b_C.conidiaHK_WB_48h | -0.26923153 | 0.02468955 |
| rs34584867 | 3 | 46065963 | A | G | 0.5112955 | 8.94E-17 | IL1b_C.conidiaHK_WB_48h | -0.26923153 | 0.02468955 |
| rs35161099 | 3 | 46067507 | G | T | 0.5040789 | 3.29E-16 | IL1b_C.conidiaHK_WB_48h | -0.26925057 | 0.024734681 |
| rs35159820 | 3 | 46068473 | G | A | 0.5112475 | 9.02E-17 | IL1b_C.conidiaHK_WB_48h | -0.26925358 | 0.024746451 |
| rs71327010 | 3 | 46068835 | T | G | 0.5039037 | 3.39E-16 | IL1b_C.conidiaHK_WB_48h | -0.26925588 | 0.024758416 |
| rs13090194 | 3 | 46069573 | C | A | 0.5115833 | 8.64E-17 | IL1b_C.conidiaHK_WB_48h | -0.26925838 | 0.024782934 |
| rs59166269 | 3 | 46070337 | G | C | 0.5113075 | 8.94E-17 | IL1b_C.conidiaHK_WB_48h | -0.26925857 | 0.024795487 |
| rs34562820 | 3 | 46071081 | A | G | 0.5041333 | 3.27E-16 | IL1b_C.conidiaHK_WB_48h | -0.26925806 | 0.024808236 |
| rs13082697 | 3 | 46071763 | C | T | 0.5096029 | 1.21E-16 | IL1b_C.conidiaHK_WB_48h | -0.26925684 | 0.024821183 |
| rs13060287 | 3 | 46071868 | G | A | 0.5111396 | 9.18E-17 | IL1b_C.conidiaHK_WB_48h | -0.26925491 | 0.024834327 |
| rs34619093 | 3 | 46072519 | A | G | 0.5041695 | 3.25E-16 | IL1b_C.conidiaHK_WB_48h | -0.2691422 | 0.024904561 |
| rs68087193 | 3 | 46072630 | T | C | 0.5113135 | 8.97E-17 | IL1b_C.conidiaHK_WB_48h | -0.2691422 | 0.024904561 |
| rs2102055 | 3 | 46072704 | A | G | 0.5140265 | 4.75E-17 | IL1b_C.conidiaHK_WB_48h | -0.2691422 | 0.024904561 |
| rs2102056 | 3 | 46072768 | A | G | 0.5114095 | 8.83E-17 | IL1b_C.conidiaHK_WB_48h | -0.2691422 | 0.024904561 |
| rs2088690 | 3 | 46072839 | T | C | 0.5114095 | 8.83E-17 | IL1b_C.conidiaHK_WB_48h | -0.2691422 | 0.024904561 |
| rs34774687 | 3 | 46073314 | C | T | 0.5114574 | 8.73E-17 | IL1b_C.conidiaHK_WB_48h | -0.26913885 | 0.02491813 |
| rs35814488 | 3 | 46073923 | G | A | 0.5114394 | 8.75E-17 | IL1b_C.conidiaHK_WB_48h | -0.2691348 | 0.024931898 |
| rs6790866 | 3 | 46074711 | A | G | 0.5114514 | 8.74E-17 | IL1b_C.conidiaHK_WB_48h | -0.26913003 | 0.024945864 |
| rs6791016 | 3 | 46074858 | A | G | 0.5114394 | 8.76E-17 | IL1b_C.conidiaHK_WB_48h | -0.26912457 | 0.024960029 |
| rs6780028 | 3 | 46075090 | C | A | 0.5114934 | 8.69E-17 | IL1b_C.conidiaHK_WB_48h | -0.26912457 | 0.024960029 |
| rs2088692 | 3 | 46077332 | G | A | 0.5114274 | 8.77E-17 | IL1b_C.conidiaHK_WB_48h | -0.26915526 | 0.024977765 |
| rs34438204 | 3 | 46081306 | C | T | 0.504556 | 3.11E-16 | IL1b_C.conidiaHK_WB_48h | -0.2690823 | 0.02508665 |
| rs72901034 | 3 | 46085321 | G | A | 0.4961586 | 1.34E-17 | IL1b_C.conidiaHK_WB_48h | -0.26899209 | 0.025187874 |
| rs34679077 | 3 | 46087992 | A | G | 0.5051656 | 2.90E-16 | IL1b_C.conidiaHK_WB_48h | -0.26878639 | 0.025335841 |
| rs4683162 | 3 | 46083941 | T | G | 0.4971505 | 1.12E-17 | IL1b_C.conidiaHK_WB_48h | -0.25728283 | 0.025426191 |
| rs34863575 | 3 | 46094063 | G | A | 0.5076787 | 2.15E-16 | IL1b_C.conidiaHK_WB_48h | -0.26829837 | 0.025703535 |
| rs35772789 | 3 | 46095104 | G | A | 0.5079254 | 2.08E-16 | IL1b_C.conidiaHK_WB_48h | -0.2681587 | 0.025794558 |
| rs13063033 | 3 | 46082481 | A | G | 0.4971019 | 7.78E-16 | IL1b_C.conidiaHK_WB_48h | -0.2540809 | 0.025930265 |
| rs77399277 | 3 | 46096798 | C | T | 0.509008 | 1.80E-16 | IL1b_C.conidiaHK_WB_48h | -0.26790687 | 0.025946056 |
| rs34718164 | 3 | 46098526 | G | C | 0.5071188 | 2.42E-16 | IL1b_C.conidiaHK_WB_48h | -0.26761041 | 0.02612145 |
| rs13096741 | 3 | 46099154 | T | C | 0.5072512 | 2.37E-16 | IL1b_C.conidiaHK_WB_48h | -0.26751935 | 0.026173538 |
| rs35420565 | 3 | 46100138 | G | C | 0.5074198 | 2.32E-16 | IL1b_C.conidiaHK_WB_48h | -0.26732879 | 0.026280369 |
| rs34047915 | 3 | 46100491 | T | C | 0.5072994 | 2.36E-16 | IL1b_C.conidiaHK_WB_48h | -0.2672794 | 0.026307633 |
| rs71327014 | 3 | 46101466 | T | C | 0.5076245 | 2.27E-16 | IL1b_C.conidiaHK_WB_48h | -0.2672293 | 0.02633512 |
| rs71327015 | 3 | 46102173 | C | G | 0.5043145 | 4.35E-16 | IL1b_C.conidiaHK_WB_48h | -0.2672293 | 0.02633512 |
| rs34127208 | 3 | 46103680 | T | G | 0.5047371 | 4.15E-16 | IL1b_C.conidiaHK_WB_48h | -0.26716656 | 0.026368869 |
| rs34766614 | 3 | 46107601 | G | A | 0.500351 | 7.64E-16 | IL1b_C.conidiaHK_WB_48h | -0.26711697 | 0.026395564 |
| rs2088693 | 3 | 46077390 | C | T | 0.4958297 | 1.42E-17 | IL1b_C.conidiaHK_WB_48h | -0.25424276 | 0.02741309 |
| rs58552652 | 3 | 46078292 | T | C | 0.4959028 | 1.40E-17 | IL1b_C.conidiaHK_WB_48h | -0.25415252 | 0.027469088 |
| rs13087776 | 3 | 46079871 | G | A | 0.4931951 | 2.14E-17 | IL1b_C.conidiaHK_WB_48h | -0.25407408 | 0.027516245 |
| rs13068410 | 3 | 46080176 | A | T | 0.4959272 | 1.38E-17 | IL1b_C.conidiaHK_WB_48h | -0.25407408 | 0.027516245 |
| rs13094262 | 3 | 46080912 | T | C | 0.4960064 | 1.37E-17 | IL1b_C.conidiaHK_WB_48h | -0.25407408 | 0.027516245 |
| rs34805664 | 3 | 46081108 | T | A | 0.5056844 | 2.69E-18 | IL1b_C.conidiaHK_WB_48h | -0.25398264 | 0.027572795 |
| rs35028116 | 3 | 46081333 | C | T | 0.4961586 | 1.35E-17 | IL1b_C.conidiaHK_WB_48h | -0.25398264 | 0.027572795 |
| rs34610609 | 3 | 46081707 | T | C | 0.4963534 | 1.31E-17 | IL1b_C.conidiaHK_WB_48h | -0.25398264 | 0.027572795 |
| rs34254987 | 3 | 46083634 | A | G | 0.4959698 | 1.38E-17 | IL1b_C.conidiaHK_WB_48h | -0.2538906 | 0.02762963 |
| rs4683163 | 3 | 46084372 | C | A | 0.495982 | 1.37E-17 | IL1b_C.conidiaHK_WB_48h | -0.2538906 | 0.02762963 |
| rs4683164 | 3 | 46084461 | G | A | 0.4945133 | 1.71E-17 | IL1b_C.conidiaHK_WB_48h | -0.25379795 | 0.027686752 |
| rs34155121 | 3 | 46084742 | A | G | 0.4955069 | 1.50E-17 | IL1b_C.conidiaHK_WB_48h | -0.25379795 | 0.027686752 |
| rs4234453 | 3 | 46084949 | A | C | 0.4954216 | 1.50E-17 | IL1b_C.conidiaHK_WB_48h | -0.25379795 | 0.027686752 |
| rs12108042 | 3 | 46086083 | G | A | 0.4961038 | 1.36E-17 | IL1b_C.conidiaHK_WB_48h | -0.25379795 | 0.027686752 |
| rs34867672 | 3 | 46088282 | C | T | 0.4970593 | 1.20E-17 | IL1b_C.conidiaHK_WB_48h | -0.25361086 | 0.027801858 |
| rs34924300 | 3 | 46088336 | T | G | 0.4970776 | 1.20E-17 | IL1b_C.conidiaHK_WB_48h | -0.25361086 | 0.027801858 |
| rs13088143 | 3 | 46080024 | T | A | 0.4959637 | 1.38E-17 | IL1b_C.conidiaHK_WB_48h | -0.25481601 | 0.027802929 |
| rs7652478 | 3 | 46090477 | A | G | 0.4976614 | 1.11E-17 | IL1b_C.conidiaHK_WB_48h | -0.25333754 | 0.027967614 |
| rs7642320 | 3 | 46090622 | G | A | 0.4978316 | 1.08E-17 | IL1b_C.conidiaHK_WB_48h | -0.25333754 | 0.027967614 |
| rs7642229 | 3 | 46090557 | G | A | 0.4959576 | 1.47E-17 | IL1b_C.conidiaHK_WB_48h | -0.25163696 | 0.028937417 |
| rs34460587 | 3 | 46142464 | T | C | 0.5045379 | 4.48E-16 | IL1b_C.conidiaHK_WB_48h | -0.26048718 | 0.029515945 |
| rs9824651 | 3 | 46108285 | T | C | 0.5120269 | 9.50E-17 | IL1b_C.conidiaHK_WB_48h | -0.25842794 | 0.030728524 |
| rs9825081 | 3 | 46108400 | A | G | 0.5118771 | 9.67E-17 | IL1b_C.conidiaHK_WB_48h | -0.25842794 | 0.030728524 |
| rs71327017 | 3 | 46111213 | C | T | 0.4985668 | 8.96E-16 | IL1b_C.conidiaHK_WB_48h | -0.2583775 | 0.030759255 |
| rs9845382 | 3 | 46111759 | A | G | 0.5066369 | 2.03E-16 | IL1b_C.conidiaHK_WB_48h | -0.2583775 | 0.030759255 |
| rs13069742 | 3 | 46114216 | G | A | 0.5064079 | 2.08E-16 | IL1b_C.conidiaHK_WB_48h | -0.2583775 | 0.030759255 |
| rs13072267 | 3 | 46116010 | A | T | 0.4979167 | 9.53E-16 | IL1b_C.conidiaHK_WB_48h | -0.2583775 | 0.030759255 |
| rs36057789 | 3 | 46117096 | G | A | 0.4975033 | 1.01E-15 | IL1b_C.conidiaHK_WB_48h | -0.2583775 | 0.030759255 |
| rs76647202 | 3 | 46117960 | G | A | 0.4978073 | 9.57E-16 | IL1b_C.conidiaHK_WB_48h | -0.2583775 | 0.030759255 |
| rs71327021 | 3 | 46120517 | G | A | 0.4978316 | 9.43E-16 | IL1b_C.conidiaHK_WB_48h | -0.2583775 | 0.030759255 |
| rs13091868 | 3 | 46123043 | G | A | 0.509687 | 2.63E-16 | IL1b_C.conidiaHK_WB_48h | -0.25832639 | 0.030790228 |
| rs13092030 | 3 | 46123055 | T | C | 0.5103595 | 2.38E-16 | IL1b_C.conidiaHK_WB_48h | -0.25832639 | 0.030790228 |
| rs17215981 | 3 | 46124418 | G | A | 0.4971931 | 1.04E-15 | IL1b_C.conidiaHK_WB_48h | -0.25832639 | 0.030790228 |
| rs13089554 | 3 | 46127557 | A | G | 0.4966395 | 1.13E-15 | IL1b_C.conidiaHK_WB_48h | -0.25832639 | 0.030790228 |
| rs13095717 | 3 | 46128542 | A | G | 0.4967673 | 1.11E-15 | IL1b_C.conidiaHK_WB_48h | -0.25832639 | 0.030790228 |
| rs13075528 | 3 | 46128709 | C | T | 0.5001448 | 7.62E-16 | IL1b_C.conidiaHK_WB_48h | -0.25832639 | 0.030790228 |
| rs13095602 | 3 | 46128713 | G | A | 0.4997202 | 8.04E-16 | IL1b_C.conidiaHK_WB_48h | -0.25832639 | 0.030790228 |
| rs71327023 | 3 | 46131225 | G | C | 0.49419 | 1.69E-15 | IL1b_C.conidiaHK_WB_48h | -0.25832639 | 0.030790228 |
| rs13063527 | 3 | 46135690 | A | G | 0.4983724 | 9.69E-16 | IL1b_C.conidiaHK_WB_48h | -0.25832639 | 0.030790228 |
| rs4373103 | 3 | 46138455 | A | G | 0.4974729 | 1.14E-15 | IL1b_C.conidiaHK_WB_48h | -0.25832639 | 0.030790228 |
| rs71327024 | 3 | 46140073 | T | G | 0.5013388 | 6.45E-16 | IL1b_C.conidiaHK_WB_48h | -0.25832639 | 0.030790228 |
| rs13075758 | 3 | 46025048 | A | G | 0.5301634 | 4.64E-18 | IL1b_C.conidiaHK_WB_48h | -0.26436164 | 0.031618541 |
| rs34442130 | 3 | 46024529 | A | T | 0.5301222 | 4.67E-18 | IL1b_C.conidiaHK_WB_48h | -0.26435024 | 0.031638163 |
| rs36122610 | 3 | 46022833 | A | G | 0.5300752 | 4.70E-18 | IL1b_C.conidiaHK_WB_48h | -0.26350746 | 0.032199852 |
| rs2230322 | 3 | 46063329 | C | T | 0.5110856 | 9.20E-17 | IL1b_C.conidiaHK_WB_48h | -0.25312292 | 0.033337883 |
| rs4443214 | 3 | 46177864 | C | T | 0.4456917 | 5.27E-12 | IL1b_C.conidiaHK_WB_48h | -0.25240523 | 0.034500207 |
| rs4493469 | 3 | 46177992 | C | T | 0.4298715 | 8.27E-11 | IL1b_C.conidiaHK_WB_48h | -0.25311322 | 0.037340701 |
| rs1491951 | 3 | 46141844 | A | G | 0.5083165 | 2.07E-16 | IL1b_C.conidiaHK_WB_48h | -0.24500223 | 0.038403757 |
| rs2109069 | 19 | 4719443 | A | G | 0.3056227 | 3.98E-12 | IL1b_Cryptococcus_PBMC_24h | -0.40381904 | 0.015635439 |
| rs2109069 | 19 | 4719443 | A | G | 0.3056227 | 3.98E-12 | IL1b_Influenza_PBMC_24h | -0.35139464 | 0.035664727 |
| rs34836513 | 3 | 46046459 | A | G | 0.5263958 | 9.51E-18 | IL1b_MSUC16_PBMC_24h | -0.36401342 | 0.025583958 |
| rs35669129 | 3 | 46055250 | A | G | 0.5281192 | 6.78E-18 | IL1b_MSUC16_PBMC_24h | -0.355231 | 0.029274541 |
| rs13081213 | 3 | 46055909 | T | C | 0.5281664 | 6.72E-18 | IL1b_MSUC16_PBMC_24h | -0.35436196 | 0.029642076 |
| rs34168660 | 3 | 46056162 | A | G | 0.5435793 | 1.04E-18 | IL1b_MSUC16_PBMC_24h | -0.35400458 | 0.029787175 |
| rs35751180 | 3 | 46056173 | T | C | 0.5342276 | 4.95E-18 | IL1b_MSUC16_PBMC_24h | -0.35381802 | 0.029796447 |
| rs4362758 | 3 | 46056911 | A | T | 0.5157296 | 5.39E-17 | IL1b_MSUC16_PBMC_24h | -0.35302927 | 0.030208025 |
| rs34754340 | 3 | 46041837 | T | C | 0.5256571 | 1.04E-17 | IL1b_MSUC16_PBMC_24h | -0.35074504 | 0.031799565 |
| rs71615438 | 3 | 46039868 | G | A | 0.5258581 | 9.97E-18 | IL1b_MSUC16_PBMC_24h | -0.34467964 | 0.034998099 |
| rs4493469 | 3 | 46177992 | C | T | 0.4298715 | 8.27E-11 | IL1b_MSUC16_PBMC_24h | -0.33695905 | 0.03733804 |
| rs4443214 | 3 | 46177864 | C | T | 0.4456917 | 5.27E-12 | IL1b_MSUC16_PBMC_24h | -0.32678141 | 0.039792309 |
| rs4683162 | 3 | 46083941 | T | G | 0.4971505 | 1.12E-17 | IL1b_MSUC16_PBMC_24h | -0.31263918 | 0.040331525 |
| rs9877748 | 3 | 46111081 | G | A | 0.5070104 | 1.95E-16 | IL1b_MSUC16_PBMC_24h | -0.33185499 | 0.040587544 |
| rs34127208 | 3 | 46103680 | T | G | 0.5047371 | 4.15E-16 | IL1b_MSUC16_PBMC_24h | -0.32284118 | 0.043124608 |
| rs71327014 | 3 | 46101466 | T | C | 0.5076245 | 2.27E-16 | IL1b_MSUC16_PBMC_24h | -0.32276436 | 0.043175523 |
| rs71327015 | 3 | 46102173 | C | G | 0.5043145 | 4.35E-16 | IL1b_MSUC16_PBMC_24h | -0.32276436 | 0.043175523 |
| rs34766614 | 3 | 46107601 | G | A | 0.500351 | 7.64E-16 | IL1b_MSUC16_PBMC_24h | -0.32267851 | 0.043225751 |
| rs34047915 | 3 | 46100491 | T | C | 0.5072994 | 2.36E-16 | IL1b_MSUC16_PBMC_24h | -0.32265369 | 0.043251239 |
| rs35420565 | 3 | 46100138 | G | C | 0.5074198 | 2.32E-16 | IL1b_MSUC16_PBMC_24h | -0.32254218 | 0.043327344 |
| rs13096741 | 3 | 46099154 | T | C | 0.5072512 | 2.37E-16 | IL1b_MSUC16_PBMC_24h | -0.32208768 | 0.043635665 |
| rs34718164 | 3 | 46098526 | G | C | 0.5071188 | 2.42E-16 | IL1b_MSUC16_PBMC_24h | -0.32185536 | 0.043792183 |
| rs17330872 | 3 | 46035097 | G | A | 0.527665 | 7.34E-18 | IL1b_MSUC16_PBMC_24h | -0.32971565 | 0.043936927 |
| rs73833520 | 3 | 46057867 | G | A | 0.5115713 | 8.68E-17 | IL1b_MSUC16_PBMC_24h | -0.32342941 | 0.044240474 |
| rs77399277 | 3 | 46096798 | C | T | 0.509008 | 1.80E-16 | IL1b_MSUC16_PBMC_24h | -0.32101579 | 0.044352523 |
| rs35772789 | 3 | 46095104 | G | A | 0.5079254 | 2.08E-16 | IL1b_MSUC16_PBMC_24h | -0.3203125 | 0.044817964 |
| rs34863575 | 3 | 46094063 | G | A | 0.5076787 | 2.15E-16 | IL1b_MSUC16_PBMC_24h | -0.31979774 | 0.045155519 |
| rs13088143 | 3 | 46080024 | T | A | 0.4959637 | 1.38E-17 | IL1b_MSUC16_PBMC_24h | -0.30631759 | 0.045778763 |
| rs2088693 | 3 | 46077390 | C | T | 0.4958297 | 1.42E-17 | IL1b_MSUC16_PBMC_24h | -0.30478752 | 0.045857099 |
| rs13087776 | 3 | 46079871 | G | A | 0.4931951 | 2.14E-17 | IL1b_MSUC16_PBMC_24h | -0.30473934 | 0.045886277 |
| rs13068410 | 3 | 46080176 | A | T | 0.4959272 | 1.38E-17 | IL1b_MSUC16_PBMC_24h | -0.30473934 | 0.045886277 |
| rs13094262 | 3 | 46080912 | T | C | 0.4960064 | 1.37E-17 | IL1b_MSUC16_PBMC_24h | -0.30473934 | 0.045886277 |
| rs58552652 | 3 | 46078292 | T | C | 0.4959028 | 1.40E-17 | IL1b_MSUC16_PBMC_24h | -0.3047239 | 0.045902296 |
| rs34805664 | 3 | 46081108 | T | A | 0.5056844 | 2.69E-18 | IL1b_MSUC16_PBMC_24h | -0.3046743 | 0.045932004 |
| rs35028116 | 3 | 46081333 | C | T | 0.4961586 | 1.35E-17 | IL1b_MSUC16_PBMC_24h | -0.3046743 | 0.045932004 |
| rs34610609 | 3 | 46081707 | T | C | 0.4963534 | 1.31E-17 | IL1b_MSUC16_PBMC_24h | -0.3046743 | 0.045932004 |
| rs34254987 | 3 | 46083634 | A | G | 0.4959698 | 1.38E-17 | IL1b_MSUC16_PBMC_24h | -0.30460855 | 0.045978024 |
| rs4683163 | 3 | 46084372 | C | A | 0.495982 | 1.37E-17 | IL1b_MSUC16_PBMC_24h | -0.30460855 | 0.045978024 |
| rs4683164 | 3 | 46084461 | G | A | 0.4945133 | 1.71E-17 | IL1b_MSUC16_PBMC_24h | -0.30454209 | 0.046024336 |
| rs34155121 | 3 | 46084742 | A | G | 0.4955069 | 1.50E-17 | IL1b_MSUC16_PBMC_24h | -0.30454209 | 0.046024336 |
| rs4234453 | 3 | 46084949 | A | C | 0.4954216 | 1.50E-17 | IL1b_MSUC16_PBMC_24h | -0.30454209 | 0.046024336 |
| rs12108042 | 3 | 46086083 | G | A | 0.4961038 | 1.36E-17 | IL1b_MSUC16_PBMC_24h | -0.30454209 | 0.046024336 |
| rs71327006 | 3 | 46058908 | A | G | 0.5044715 | 3.14E-16 | IL1b_MSUC16_PBMC_24h | -0.31814214 | 0.046116423 |
| rs34867672 | 3 | 46088282 | C | T | 0.4970593 | 1.20E-17 | IL1b_MSUC16_PBMC_24h | -0.30440706 | 0.04611784 |
| rs34924300 | 3 | 46088336 | T | G | 0.4970776 | 1.20E-17 | IL1b_MSUC16_PBMC_24h | -0.30440706 | 0.04611784 |
| rs7652478 | 3 | 46090477 | A | G | 0.4976614 | 1.11E-17 | IL1b_MSUC16_PBMC_24h | -0.3042783 | 0.046198695 |
| rs7642320 | 3 | 46090622 | G | A | 0.4978316 | 1.08E-17 | IL1b_MSUC16_PBMC_24h | -0.3042783 | 0.046198695 |
| rs71327007 | 3 | 46059249 | T | C | 0.5044473 | 3.14E-16 | IL1b_MSUC16_PBMC_24h | -0.31763621 | 0.046437885 |
| rs35454877 | 3 | 46059484 | C | T | 0.5121288 | 7.99E-17 | IL1b_MSUC16_PBMC_24h | -0.31735141 | 0.046622243 |
| rs34679077 | 3 | 46087992 | A | G | 0.5051656 | 2.90E-16 | IL1b_MSUC16_PBMC_24h | -0.31731889 | 0.046757536 |
| rs72901034 | 3 | 46085321 | G | A | 0.4961586 | 1.34E-17 | IL1b_MSUC16_PBMC_24h | -0.31666772 | 0.0471738 |
| rs7642229 | 3 | 46090557 | G | A | 0.4959576 | 1.47E-17 | IL1b_MSUC16_PBMC_24h | -0.30251726 | 0.047344312 |
| rs34438204 | 3 | 46081306 | C | T | 0.504556 | 3.11E-16 | IL1b_MSUC16_PBMC_24h | -0.3157592 | 0.047751476 |
| rs2088692 | 3 | 46077332 | G | A | 0.5114274 | 8.77E-17 | IL1b_MSUC16_PBMC_24h | -0.31466337 | 0.048445801 |
| rs35334665 | 3 | 46061218 | C | T | 0.5042903 | 3.19E-16 | IL1b_MSUC16_PBMC_24h | -0.31424049 | 0.048636907 |
| rs36040135 | 3 | 46061304 | G | A | 0.5113495 | 8.88E-17 | IL1b_MSUC16_PBMC_24h | -0.31400825 | 0.048784928 |
| rs6791016 | 3 | 46074858 | A | G | 0.5114394 | 8.76E-17 | IL1b_MSUC16_PBMC_24h | -0.31401831 | 0.048862662 |
| rs6780028 | 3 | 46075090 | C | A | 0.5114934 | 8.69E-17 | IL1b_MSUC16_PBMC_24h | -0.31401831 | 0.048862662 |
| rs6790866 | 3 | 46074711 | A | G | 0.5114514 | 8.74E-17 | IL1b_MSUC16_PBMC_24h | -0.31385634 | 0.048965227 |
| rs35814488 | 3 | 46073923 | G | A | 0.5114394 | 8.75E-17 | IL1b_MSUC16_PBMC_24h | -0.31369359 | 0.049068253 |
| rs34774687 | 3 | 46073314 | C | T | 0.5114574 | 8.73E-17 | IL1b_MSUC16_PBMC_24h | -0.31353007 | 0.049171741 |
| rs34619093 | 3 | 46072519 | A | G | 0.5041695 | 3.25E-16 | IL1b_MSUC16_PBMC_24h | -0.31336578 | 0.049275693 |
| rs68087193 | 3 | 46072630 | T | C | 0.5113135 | 8.97E-17 | IL1b_MSUC16_PBMC_24h | -0.31336578 | 0.049275693 |
| rs2102055 | 3 | 46072704 | A | G | 0.5140265 | 4.75E-17 | IL1b_MSUC16_PBMC_24h | -0.31336578 | 0.049275693 |
| rs2102056 | 3 | 46072768 | A | G | 0.5114095 | 8.83E-17 | IL1b_MSUC16_PBMC_24h | -0.31336578 | 0.049275693 |
| rs2088690 | 3 | 46072839 | T | C | 0.5114095 | 8.83E-17 | IL1b_MSUC16_PBMC_24h | -0.31336578 | 0.049275693 |
| rs13060287 | 3 | 46071868 | G | A | 0.5111396 | 9.18E-17 | IL1b_MSUC16_PBMC_24h | -0.31328357 | 0.049325028 |
| rs13082697 | 3 | 46071763 | C | T | 0.5096029 | 1.21E-16 | IL1b_MSUC16_PBMC_24h | -0.31311773 | 0.049429818 |
| rs34562820 | 3 | 46071081 | A | G | 0.5041333 | 3.27E-16 | IL1b_MSUC16_PBMC_24h | -0.31295113 | 0.049535074 |
| rs59166269 | 3 | 46070337 | G | C | 0.5113075 | 8.94E-17 | IL1b_MSUC16_PBMC_24h | -0.31278376 | 0.049640799 |
| rs13097556 | 3 | 46061997 | C | T | 0.5096389 | 1.23E-16 | IL1b_MSUC16_PBMC_24h | -0.3126801 | 0.049653952 |
| rs13090194 | 3 | 46069573 | C | A | 0.5115833 | 8.64E-17 | IL1b_MSUC16_PBMC_24h | -0.31261563 | 0.049746992 |
| rs71327010 | 3 | 46068835 | T | G | 0.5039037 | 3.39E-16 | IL1b_MSUC16_PBMC_24h | -0.3122771 | 0.049960792 |
| rs72856706 | 6 | 31120729 | T | C | 0.6148018 | 9.15E-18 | IL1b_PHA_WB_48h | 0.22855868 | 0.040037638 |
| rs111837807 | 6 | 31121232 | C | T | 0.6149802 | 8.95E-18 | IL1b_PHA_WB_48h | 0.22855868 | 0.040037638 |
| rs112785163 | 6 | 31121381 | C | G | 0.6149316 | 9.02E-18 | IL1b_PHA_WB_48h | 0.22855868 | 0.040037638 |
| rs143334143 | 6 | 31121426 | A | G | 0.6150829 | 8.82E-18 | IL1b_PHA_WB_48h | 0.22855868 | 0.040037638 |
| rs72856713 | 6 | 31121958 | G | C | 0.5882588 | 2.84E-16 | IL1b_PHA_WB_48h | 0.22855868 | 0.040037638 |
| rs13063033 | 3 | 46082481 | A | G | 0.4971019 | 7.78E-16 | IL22_A.fumigatusconidia_PBMC_7days | 0.44924111 | 0.016222315 |
| rs7642229 | 3 | 46090557 | G | A | 0.4959576 | 1.47E-17 | IL22_A.fumigatusconidia_PBMC_7days | 0.41829834 | 0.026264205 |
| rs2088693 | 3 | 46077390 | C | T | 0.4958297 | 1.42E-17 | IL22_A.fumigatusconidia_PBMC_7days | 0.41772483 | 0.026582318 |
| rs13087776 | 3 | 46079871 | G | A | 0.4931951 | 2.14E-17 | IL22_A.fumigatusconidia_PBMC_7days | 0.41743696 | 0.026683708 |
| rs13068410 | 3 | 46080176 | A | T | 0.4959272 | 1.38E-17 | IL22_A.fumigatusconidia_PBMC_7days | 0.41743696 | 0.026683708 |
| rs13094262 | 3 | 46080912 | T | C | 0.4960064 | 1.37E-17 | IL22_A.fumigatusconidia_PBMC_7days | 0.41743696 | 0.026683708 |
| rs58552652 | 3 | 46078292 | T | C | 0.4959028 | 1.40E-17 | IL22_A.fumigatusconidia_PBMC_7days | 0.41744516 | 0.026684181 |
| rs34805664 | 3 | 46081108 | T | A | 0.5056844 | 2.69E-18 | IL22_A.fumigatusconidia_PBMC_7days | 0.41715536 | 0.026786258 |
| rs35028116 | 3 | 46081333 | C | T | 0.4961586 | 1.35E-17 | IL22_A.fumigatusconidia_PBMC_7days | 0.41715536 | 0.026786258 |
| rs34610609 | 3 | 46081707 | T | C | 0.4963534 | 1.31E-17 | IL22_A.fumigatusconidia_PBMC_7days | 0.41715536 | 0.026786258 |
| rs34254987 | 3 | 46083634 | A | G | 0.4959698 | 1.38E-17 | IL22_A.fumigatusconidia_PBMC_7days | 0.4168728 | 0.026889324 |
| rs4683163 | 3 | 46084372 | C | A | 0.495982 | 1.37E-17 | IL22_A.fumigatusconidia_PBMC_7days | 0.4168728 | 0.026889324 |
| rs4683164 | 3 | 46084461 | G | A | 0.4945133 | 1.71E-17 | IL22_A.fumigatusconidia_PBMC_7days | 0.41658927 | 0.026992909 |

| | | | | | | | | | |
|---|---|---|---|---|---|---|---|---|---|
| rs34155121 | 3 | 46084742 | A | G | 0.4955069 | 1.50E-17 | IL22_A.fumigatusconidia_PBMC_7days | 0.41658927 | 0.026992909 |
| rs4234453 | 3 | 46084949 | A | C | 0.4954216 | 1.50E-17 | IL22_A.fumigatusconidia_PBMC_7days | 0.41658927 | 0.026992909 |
| rs12108042 | 3 | 46086083 | G | A | 0.4961038 | 1.36E-17 | IL22_A.fumigatusconidia_PBMC_7days | 0.41658927 | 0.026992909 |
| rs34867672 | 3 | 46088282 | C | T | 0.4970593 | 1.20E-17 | IL22_A.fumigatusconidia_PBMC_7days | 0.41601932 | 0.027201645 |
| rs34924300 | 3 | 46088336 | T | G | 0.4970776 | 1.20E-17 | IL22_A.fumigatusconidia_PBMC_7days | 0.41601932 | 0.027201645 |
| rs4683162 | 3 | 46083941 | T | G | 0.4971505 | 1.12E-17 | IL22_A.fumigatusconidia_PBMC_7days | 0.41523409 | 0.027357882 |
| rs7652478 | 3 | 46090477 | A | G | 0.4976614 | 1.11E-17 | IL22_A.fumigatusconidia_PBMC_7days | 0.41542876 | 0.02741361 |
| rs7642320 | 3 | 46090622 | G | A | 0.4978316 | 1.08E-17 | IL22_A.fumigatusconidia_PBMC_7days | 0.41542876 | 0.02741361 |
| rs36122610 | 3 | 46022833 | A | G | 0.5300752 | 4.70E-18 | IL22_A.fumigatusconidia_PBMC_7days | 0.43926469 | 0.029324556 |
| rs13088143 | 3 | 46080024 | T | A | 0.4959637 | 1.38E-17 | IL22_A.fumigatusconidia_PBMC_7days | 0.41145909 | 0.029709065 |
| rs34442130 | 3 | 46024529 | A | T | 0.5301222 | 4.67E-18 | IL22_A.fumigatusconidia_PBMC_7days | 0.43683092 | 0.030238686 |
| rs13075758 | 3 | 46025048 | A | G | 0.5301634 | 4.64E-18 | IL22_A.fumigatusconidia_PBMC_7days | 0.43654335 | 0.030338485 |
| rs17330872 | 3 | 46035097 | G | A | 0.527665 | 7.34E-18 | IL22_A.fumigatusconidia_PBMC_7days | 0.42335303 | 0.036126825 |
| rs71615438 | 3 | 46039868 | G | A | 0.5258581 | 9.97E-18 | IL22_A.fumigatusconidia_PBMC_7days | 0.42230075 | 0.036437812 |
| rs73833520 | 3 | 46057867 | G | A | 0.5115713 | 8.68E-17 | IL22_A.fumigatusconidia_PBMC_7days | 0.41463984 | 0.036685905 |
| rs34754340 | 3 | 46041837 | T | C | 0.5256571 | 1.04E-17 | IL22_A.fumigatusconidia_PBMC_7days | 0.4213386 | 0.036760744 |
| rs34836513 | 3 | 46046459 | A | G | 0.5263958 | 9.51E-18 | IL22_A.fumigatusconidia_PBMC_7days | 0.41937273 | 0.037324622 |
| rs71327014 | 3 | 46101466 | T | C | 0.5076245 | 2.27E-16 | IL22_A.fumigatusconidia_PBMC_7days | 0.40607718 | 0.039349956 |
| rs71327015 | 3 | 46102173 | C | G | 0.5043145 | 4.35E-16 | IL22_A.fumigatusconidia_PBMC_7days | 0.40607718 | 0.039349956 |
| rs34047915 | 3 | 46100491 | T | C | 0.5072994 | 2.36E-16 | IL22_A.fumigatusconidia_PBMC_7days | 0.40586572 | 0.039456551 |
| rs34127208 | 3 | 46103680 | T | G | 0.5047371 | 4.15E-16 | IL22_A.fumigatusconidia_PBMC_7days | 0.40583807 | 0.039465661 |
| rs34766614 | 3 | 46107601 | G | A | 0.500351 | 7.64E-16 | IL22_A.fumigatusconidia_PBMC_7days | 0.40575097 | 0.039503801 |
| rs35420565 | 3 | 46100138 | G | C | 0.5074198 | 2.32E-16 | IL22_A.fumigatusconidia_PBMC_7days | 0.40565318 | 0.039563653 |
| rs13096741 | 3 | 46099154 | T | C | 0.5072512 | 2.37E-16 | IL22_A.fumigatusconidia_PBMC_7days | 0.40479237 | 0.039997157 |
| rs34718164 | 3 | 46098526 | G | C | 0.5071188 | 2.42E-16 | IL22_A.fumigatusconidia_PBMC_7days | 0.40435558 | 0.040216995 |
| rs71327006 | 3 | 46058908 | A | G | 0.5044715 | 3.14E-16 | IL22_A.fumigatusconidia_PBMC_7days | 0.40279455 | 0.04083317 |
| rs77399277 | 3 | 46096798 | C | T | 0.509008 | 1.80E-16 | IL22_A.fumigatusconidia_PBMC_7days | 0.40279353 | 0.041002875 |
| rs71327007 | 3 | 46059249 | T | C | 0.5044473 | 3.14E-16 | IL22_A.fumigatusconidia_PBMC_7days | 0.4019656 | 0.041226197 |
| rs35772789 | 3 | 46095104 | G | A | 0.5079254 | 2.08E-16 | IL22_A.fumigatusconidia_PBMC_7days | 0.40188335 | 0.041459757 |
| rs35454877 | 3 | 46059484 | C | T | 0.5121288 | 7.99E-17 | IL22_A.fumigatusconidia_PBMC_7days | 0.40140222 | 0.041500556 |
| rs4493469 | 3 | 46177992 | C | T | 0.4298715 | 8.27E-11 | IL22_A.fumigatusconidia_PBMC_7days | 0.40700338 | 0.041698439 |
| rs34863575 | 3 | 46094063 | G | A | 0.5076787 | 2.15E-16 | IL22_A.fumigatusconidia_PBMC_7days | 0.40094686 | 0.041930171 |
| rs34679077 | 3 | 46087992 | A | G | 0.5051656 | 2.90E-16 | IL22_A.fumigatusconidia_PBMC_7days | 0.39653025 | 0.044157411 |
| rs4443214 | 3 | 46177864 | C | T | 0.4456917 | 5.27E-12 | IL22_A.fumigatusconidia_PBMC_7days | 0.39456299 | 0.044410904 |
| rs72901034 | 3 | 46085321 | G | A | 0.4961586 | 1.34E-17 | IL22_A.fumigatusconidia_PBMC_7days | 0.39556915 | 0.044638806 |
| rs35334665 | 3 | 46061218 | C | T | 0.5042903 | 3.19E-16 | IL22_A.fumigatusconidia_PBMC_7days | 0.39389875 | 0.04529905 |
| rs36040135 | 3 | 46061304 | G | A | 0.5113495 | 8.88E-17 | IL22_A.fumigatusconidia_PBMC_7days | 0.39361981 | 0.045433926 |
| rs34438204 | 3 | 46081306 | C | T | 0.504556 | 3.11E-16 | IL22_A.fumigatusconidia_PBMC_7days | 0.39399505 | 0.045438016 |
| rs2088692 | 3 | 46077332 | G | A | 0.5114274 | 8.77E-17 | IL22_A.fumigatusconidia_PBMC_7days | 0.39211451 | 0.046397954 |
| rs34460587 | 3 | 46142464 | T | C | 0.5045379 | 4.48E-16 | IL22_A.fumigatusconidia_PBMC_7days | 0.39066619 | 0.046456149 |
| rs6791016 | 3 | 46074858 | A | G | 0.5114394 | 8.76E-17 | IL22_A.fumigatusconidia_PBMC_7days | 0.39120568 | 0.04686305 |
| rs6780028 | 3 | 46075090 | C | A | 0.5114934 | 8.69E-17 | IL22_A.fumigatusconidia_PBMC_7days | 0.39120568 | 0.04686305 |
| rs6790866 | 3 | 46074711 | A | G | 0.5114514 | 8.74E-17 | IL22_A.fumigatusconidia_PBMC_7days | 0.39093038 | 0.047004617 |
| rs35814488 | 3 | 46073923 | G | A | 0.5114394 | 8.75E-17 | IL22_A.fumigatusconidia_PBMC_7days | 0.39065413 | 0.047146811 |
| rs13097556 | 3 | 46061997 | C | T | 0.5096389 | 1.23E-16 | IL22_A.fumigatusconidia_PBMC_7days | 0.39037734 | 0.047155138 |
| rs34774687 | 3 | 46073314 | C | T | 0.5114574 | 8.73E-17 | IL22_A.fumigatusconidia_PBMC_7days | 0.39037694 | 0.047289634 |
| rs13060287 | 3 | 46071868 | G | A | 0.5111396 | 9.18E-17 | IL22_A.fumigatusconidia_PBMC_7days | 0.39014124 | 0.047398586 |
| rs34619093 | 3 | 46072519 | A | G | 0.5041695 | 3.25E-16 | IL22_A.fumigatusconidia_PBMC_7days | 0.39009882 | 0.047433087 |
| rs68087193 | 3 | 46072630 | T | C | 0.5113135 | 8.97E-17 | IL22_A.fumigatusconidia_PBMC_7days | 0.39009882 | 0.047433087 |
| rs2102055 | 3 | 46072704 | A | G | 0.5140265 | 4.75E-17 | IL22_A.fumigatusconidia_PBMC_7days | 0.39009882 | 0.047433087 |
| rs2102056 | 3 | 46072768 | A | G | 0.5114095 | 8.83E-17 | IL22_A.fumigatusconidia_PBMC_7days | 0.39009882 | 0.047433087 |
| rs2088690 | 3 | 46072839 | T | C | 0.5114095 | 8.83E-17 | IL22_A.fumigatusconidia_PBMC_7days | 0.39009882 | 0.047433087 |
| rs13082697 | 3 | 46071763 | C | T | 0.5096029 | 1.21E-16 | IL22_A.fumigatusconidia_PBMC_7days | 0.3898612 | 0.047542869 |
| rs34562820 | 3 | 46071081 | A | G | 0.5041333 | 3.27E-16 | IL22_A.fumigatusconidia_PBMC_7days | 0.38958022 | 0.047687787 |
| rs59166269 | 3 | 46070337 | G | C | 0.5113075 | 8.94E-17 | IL22_A.fumigatusconidia_PBMC_7days | 0.38929831 | 0.047833344 |
| rs13090194 | 3 | 46069573 | C | A | 0.5115833 | 8.64E-17 | IL22_A.fumigatusconidia_PBMC_7days | 0.38901547 | 0.04797954 |
| rs71327010 | 3 | 46068835 | T | G | 0.5039037 | 3.39E-16 | IL22_A.fumigatusconidia_PBMC_7days | 0.38844704 | 0.048273861 |
| rs35159820 | 3 | 46068473 | G | A | 0.5112475 | 9.02E-17 | IL22_A.fumigatusconidia_PBMC_7days | 0.38816144 | 0.04842199 |
| rs35161099 | 3 | 46067507 | G | T | 0.5040789 | 3.29E-16 | IL22_A.fumigatusconidia_PBMC_7days | 0.38787493 | 0.048570768 |
| rs35930050 | 3 | 46065763 | A | G | 0.5113015 | 8.93E-17 | IL22_A.fumigatusconidia_PBMC_7days | 0.38671979 | 0.049172402 |
| rs34584867 | 3 | 46065963 | A | G | 0.5112955 | 8.94E-17 | IL22_A.fumigatusconidia_PBMC_7days | 0.38671979 | 0.049172402 |
| rs35669129 | 3 | 46055250 | A | G | 0.5281192 | 6.78E-18 | IL22_A.fumigatusconidia_PBMC_7days | 0.39520776 | 0.049650748 |
| rs876668 | 3 | 46063753 | C | T | 0.5112415 | 9.01E-17 | IL22_A.fumigatusconidia_PBMC_7days | 0.38555021 | 0.049784575 |
| rs35751180 | 3 | 46056173 | T | C | 0.5342276 | 4.95E-18 | IL22_A.fumigatusconidia_PBMC_7days | 0.39443965 | 0.049914823 |
| rs13063033 | 3 | 46082481 | A | G | 0.4971019 | 7.78E-16 | IL22_B.burgdorferi_PBMC_7days | -0.30632643 | 0.044037466 |
| rs36122610 | 3 | 46022833 | A | G | 0.5300752 | 4.70E-18 | IL22_B.burgdorferi_PBMC_7days | -0.32616618 | 0.046707191 |
| rs34442130 | 3 | 46024529 | A | T | 0.5301222 | 4.67E-18 | IL22_B.burgdorferi_PBMC_7days | -0.32529503 | 0.047299832 |
| rs13075758 | 3 | 46025048 | A | G | 0.5301634 | 4.64E-18 | IL22_B.burgdorferi_PBMC_7days | -0.32520981 | 0.047345045 |
| rs13063033 | 3 | 46082481 | A | G | 0.4971019 | 7.78E-16 | IL22_C.albicansconidia_PBMC_7days | -0.58866533 | 0.000602469 |
| rs4493469 | 3 | 46177992 | C | T | 0.4298715 | 8.27E-11 | IL22_C.albicansconidia_PBMC_7days | -0.58256232 | 0.00150099 |
| rs4443214 | 3 | 46177864 | C | T | 0.4456917 | 5.27E-12 | IL22_C.albicansconidia_PBMC_7days | -0.56289561 | 0.001791915 |
| rs2230322 | 3 | 46063329 | C | T | 0.5110856 | 9.20E-17 | IL22_C.albicansconidia_PBMC_7days | -0.55908011 | 0.001814071 |
| rs4362758 | 3 | 46056911 | A | T | 0.5157296 | 5.39E-17 | IL22_C.albicansconidia_PBMC_7days | -0.57591464 | 0.001828601 |
| rs35159820 | 3 | 46068473 | G | A | 0.5112475 | 9.02E-17 | IL22_C.albicansconidia_PBMC_7days | -0.56108124 | 0.001892859 |
| rs35161099 | 3 | 46067507 | G | T | 0.5040789 | 3.29E-16 | IL22_C.albicansconidia_PBMC_7days | -0.56103651 | 0.001892859 |
| rs71327010 | 3 | 46068835 | T | G | 0.5039037 | 3.39E-16 | IL22_C.albicansconidia_PBMC_7days | -0.56112453 | 0.001892885 |
| rs13090194 | 3 | 46069573 | C | A | 0.5115833 | 8.64E-17 | IL22_C.albicansconidia_PBMC_7days | -0.56120678 | 0.001893014 |
| rs59166269 | 3 | 46070337 | G | C | 0.5113075 | 8.94E-17 | IL22_C.albicansconidia_PBMC_7days | -0.56124574 | 0.001893118 |
| rs35930050 | 3 | 46065763 | A | G | 0.5113015 | 8.93E-17 | IL22_C.albicansconidia_PBMC_7days | -0.56084322 | 0.001893119 |
| rs34584867 | 3 | 46065963 | A | G | 0.5112955 | 8.94E-17 | IL22_C.albicansconidia_PBMC_7days | -0.56084322 | 0.001893119 |
| rs34562820 | 3 | 46071081 | A | G | 0.5041333 | 3.27E-16 | IL22_C.albicansconidia_PBMC_7days | -0.56128326 | 0.001893248 |
| rs13082697 | 3 | 46071763 | C | T | 0.5096029 | 1.21E-16 | IL22_C.albicansconidia_PBMC_7days | -0.56131934 | 0.001893404 |
| rs13060287 | 3 | 46071868 | G | A | 0.5111396 | 9.18E-17 | IL22_C.albicansconidia_PBMC_7days | -0.56135397 | 0.001893586 |
| rs876668 | 3 | 46063753 | C | T | 0.5112415 | 9.01E-17 | IL22_C.albicansconidia_PBMC_7days | -0.56062697 | 0.001893793 |
| rs34168660 | 3 | 46056162 | A | G | 0.5435793 | 1.04E-18 | IL22_C.albicansconidia_PBMC_7days | -0.57391989 | 0.001901827 |
| rs34619093 | 3 | 46072519 | A | G | 0.5041695 | 3.25E-16 | IL22_C.albicansconidia_PBMC_7days | -0.56115027 | 0.001901837 |
| rs68087193 | 3 | 46072630 | T | C | 0.5113135 | 8.97E-17 | IL22_C.albicansconidia_PBMC_7days | -0.56115027 | 0.001901837 |
| rs2102055 | 3 | 46072704 | A | G | 0.5140265 | 4.75E-17 | IL22_C.albicansconidia_PBMC_7days | -0.56115027 | 0.001901837 |
| rs2102056 | 3 | 46072768 | A | G | 0.5114095 | 8.83E-17 | IL22_C.albicansconidia_PBMC_7days | -0.56115027 | 0.001901837 |
| rs2088690 | 3 | 46072839 | T | C | 0.5114095 | 8.83E-17 | IL22_C.albicansconidia_PBMC_7days | -0.56115027 | 0.001901837 |
| rs34774687 | 3 | 46073314 | C | T | 0.5114574 | 8.73E-17 | IL22_C.albicansconidia_PBMC_7days | -0.56118197 | 0.001902072 |
| rs35814488 | 3 | 46073923 | G | A | 0.5114394 | 8.75E-17 | IL22_C.albicansconidia_PBMC_7days | -0.56121223 | 0.001902334 |
| rs6790866 | 3 | 46074711 | A | G | 0.5114514 | 8.74E-17 | IL22_C.albicansconidia_PBMC_7days | -0.56124104 | 0.001902622 |
| rs6791016 | 3 | 46074858 | A | G | 0.5114394 | 8.76E-17 | IL22_C.albicansconidia_PBMC_7days | -0.5612684 | 0.001902936 |
| rs6780028 | 3 | 46075090 | C | A | 0.5114934 | 8.69E-17 | IL22_C.albicansconidia_PBMC_7days | -0.5612684 | 0.001902936 |
| rs35751180 | 3 | 46056173 | T | C | 0.5342276 | 4.95E-18 | IL22_C.albicansconidia_PBMC_7days | -0.57352032 | 0.001906391 |
| rs2088692 | 3 | 46077332 | G | A | 0.5114274 | 8.77E-17 | IL22_C.albicansconidia_PBMC_7days | -0.56111626 | 0.001912687 |
| rs34438204 | 3 | 46081306 | C | T | 0.504556 | 3.11E-16 | IL22_C.albicansconidia_PBMC_7days | -0.56123641 | 0.001916194 |
| rs72901034 | 3 | 46085321 | G | A | 0.4961586 | 1.34E-17 | IL22_C.albicansconidia_PBMC_7days | -0.56128262 | 0.001920236 |
| rs9877748 | 3 | 46111081 | G | A | 0.5070104 | 1.95E-16 | IL22_C.albicansconidia_PBMC_7days | -0.57003015 | 0.00192628 |
| rs34679077 | 3 | 46087992 | A | G | 0.5051656 | 2.90E-16 | IL22_C.albicansconidia_PBMC_7days | -0.56104222 | 0.001932537 |
| rs13081213 | 3 | 46055909 | T | C | 0.5281664 | 6.72E-18 | IL22_C.albicansconidia_PBMC_7days | -0.57304595 | 0.001935398 |
| rs13097556 | 3 | 46061997 | C | T | 0.5096389 | 1.23E-16 | IL22_C.albicansconidia_PBMC_7days | -0.5595787 | 0.001948125 |
| rs34863575 | 3 | 46094063 | G | A | 0.5076787 | 2.15E-16 | IL22_C.albicansconidia_PBMC_7days | -0.56073386 | 0.001953008 |
| rs35772789 | 3 | 46095104 | G | A | 0.5079254 | 2.08E-16 | IL22_C.albicansconidia_PBMC_7days | -0.56060104 | 0.001958772 |
| rs77399277 | 3 | 46096798 | C | T | 0.509008 | 1.80E-16 | IL22_C.albicansconidia_PBMC_7days | -0.5599339 | 0.001983987 |
| rs36040135 | 3 | 46061304 | G | A | 0.5113495 | 8.88E-17 | IL22_C.albicansconidia_PBMC_7days | -0.55875768 | 0.001987493 |
| rs34718164 | 3 | 46098526 | G | C | 0.5071188 | 2.42E-16 | IL22_C.albicansconidia_PBMC_7days | -0.55959419 | 0.001996244 |
| rs13096741 | 3 | 46099154 | T | C | 0.5072512 | 2.37E-16 | IL22_C.albicansconidia_PBMC_7days | -0.55948402 | 0.002000005 |
| rs35334665 | 3 | 46061218 | C | T | 0.5042903 | 3.19E-16 | IL22_C.albicansconidia_PBMC_7days | -0.55829459 | 0.002006736 |
| rs35420565 | 3 | 46100138 | G | C | 0.5074198 | 2.32E-16 | IL22_C.albicansconidia_PBMC_7days | -0.55924625 | 0.002007877 |
| rs34047915 | 3 | 46100491 | T | C | 0.5072994 | 2.36E-16 | IL22_C.albicansconidia_PBMC_7days | -0.55918317 | 0.002009918 |
| rs71327014 | 3 | 46101466 | T | C | 0.5076245 | 2.27E-16 | IL22_C.albicansconidia_PBMC_7days | -0.55911864 | 0.002011988 |
| rs71327015 | 3 | 46102173 | C | G | 0.5043145 | 4.35E-16 | IL22_C.albicansconidia_PBMC_7days | -0.55911864 | 0.002011988 |
| rs35669129 | 3 | 46055250 | A | G | 0.5281192 | 6.78E-18 | IL22_C.albicansconidia_PBMC_7days | -0.57087378 | 0.002018753 |
| rs34766614 | 3 | 46107601 | G | A | 0.500351 | 7.64E-16 | IL22_C.albicansconidia_PBMC_7days | -0.55887714 | 0.002020576 |
| rs34127208 | 3 | 46103680 | T | G | 0.5047371 | 4.15E-16 | IL22_C.albicansconidia_PBMC_7days | -0.55864762 | 0.002029788 |
| rs35454877 | 3 | 46059484 | C | T | 0.5121288 | 7.99E-17 | IL22_C.albicansconidia_PBMC_7days | -0.55627922 | 0.002102441 |
| rs71327007 | 3 | 46059249 | T | C | 0.5044473 | 3.14E-16 | IL22_C.albicansconidia_PBMC_7days | -0.5559965 | 0.002114927 |
| rs71327006 | 3 | 46058908 | A | G | 0.5044715 | 3.14E-16 | IL22_C.albicansconidia_PBMC_7days | -0.5552276 | 0.002148424 |
| rs1491951 | 3 | 46141844 | A | G | 0.5083165 | 2.07E-16 | IL22_C.albicansconidia_PBMC_7days | -0.54524321 | 0.00224495 |
| rs13075758 | 3 | 46025048 | A | G | 0.5301634 | 4.64E-18 | IL22_C.albicansconidia_PBMC_7days | -0.56067677 | 0.002474993 |
| rs34442130 | 3 | 46024529 | A | T | 0.5301222 | 4.67E-18 | IL22_C.albicansconidia_PBMC_7days | -0.56069562 | 0.002475623 |
| rs34460587 | 3 | 46142464 | T | C | 0.5045379 | 4.48E-16 | IL22_C.albicansconidia_PBMC_7days | -0.54538635 | 0.002478423 |
| rs13091868 | 3 | 46123043 | G | A | 0.509687 | 2.63E-16 | IL22_C.albicansconidia_PBMC_7days | -0.54481333 | 0.002488023 |
| rs13092030 | 3 | 46123055 | T | C | 0.5103595 | 2.38E-16 | IL22_C.albicansconidia_PBMC_7days | -0.54481333 | 0.002488023 |
| rs17215981 | 3 | 46124418 | G | A | 0.4971931 | 1.04E-15 | IL22_C.albicansconidia_PBMC_7days | -0.54481333 | 0.002488023 |
| rs13089554 | 3 | 46127557 | A | G | 0.4966395 | 1.13E-15 | IL22_C.albicansconidia_PBMC_7days | -0.54481333 | 0.002488023 |
| rs13095717 | 3 | 46128542 | A | G | 0.4967673 | 1.11E-15 | IL22_C.albicansconidia_PBMC_7days | -0.54481333 | 0.002488023 |
| rs13075528 | 3 | 46128709 | C | T | 0.5001448 | 7.62E-16 | IL22_C.albicansconidia_PBMC_7days | -0.54481333 | 0.002488023 |
| rs13095602 | 3 | 46128713 | G | A | 0.4997202 | 8.04E-16 | IL22_C.albicansconidia_PBMC_7days | -0.54481333 | 0.002488023 |
| rs71327023 | 3 | 46131225 | G | C | 0.49419 | 1.69E-15 | IL22_C.albicansconidia_PBMC_7days | -0.54481333 | 0.002488023 |
| rs13063527 | 3 | 46135690 | A | G | 0.4983724 | 9.69E-16 | IL22_C.albicansconidia_PBMC_7days | -0.54481333 | 0.002488023 |
| rs4373103 | 3 | 46138455 | A | G | 0.4974729 | 1.14E-15 | IL22_C.albicansconidia_PBMC_7days | -0.54481333 | 0.002488023 |
| rs71327024 | 3 | 46140073 | T | G | 0.5013388 | 6.45E-16 | IL22_C.albicansconidia_PBMC_7days | -0.54481333 | 0.002488023 |
| rs36122610 | 3 | 46022833 | A | G | 0.5300752 | 4.70E-18 | IL22_C.albicansconidia_PBMC_7days | -0.56018661 | 0.002498444 |
| rs71327017 | 3 | 46111213 | C | T | 0.4985668 | 8.96E-16 | IL22_C.albicansconidia_PBMC_7days | -0.54458986 | 0.002499018 |
| rs9845382 | 3 | 46111759 | A | G | 0.5066369 | 2.03E-16 | IL22_C.albicansconidia_PBMC_7days | -0.54458986 | 0.002499018 |
| rs13069742 | 3 | 46114216 | G | A | 0.5064079 | 2.08E-16 | IL22_C.albicansconidia_PBMC_7days | -0.54458986 | 0.002499018 |
| rs13072267 | 3 | 46116010 | A | T | 0.4979167 | 9.53E-16 | IL22_C.albicansconidia_PBMC_7days | -0.54458986 | 0.002499018 |
| rs36057789 | 3 | 46117096 | G | A | 0.4975033 | 1.01E-15 | IL22_C.albicansconidia_PBMC_7days | -0.54458986 | 0.002499018 |
| rs76647202 | 3 | 46117960 | G | A | 0.4978073 | 9.57E-16 | IL22_C.albicansconidia_PBMC_7days | -0.54458986 | 0.002499018 |
| rs71327021 | 3 | 46120517 | G | A | 0.4978316 | 9.43E-16 | IL22_C.albicansconidia_PBMC_7days | -0.54458986 | 0.002499018 |
| rs9824651 | 3 | 46108285 | T | C | 0.5120269 | 9.50E-17 | IL22_C.albicansconidia_PBMC_7days | -0.54436497 | 0.002510091 |
| rs9825081 | 3 | 46108400 | A | G | 0.5118771 | 9.67E-17 | IL22_C.albicansconidia_PBMC_7days | -0.54436497 | 0.002510091 |
| rs17330872 | 3 | 46035097 | G | A | 0.527665 | 7.34E-18 | IL22_C.albicansconidia_PBMC_7days | -0.55399015 | 0.002848216 |
| rs73833520 | 3 | 46057867 | G | A | 0.5115713 | 8.68E-17 | IL22_C.albicansconidia_PBMC_7days | -0.54062843 | 0.003038221 |
| rs71615438 | 3 | 46039868 | G | A | 0.5258581 | 9.97E-18 | IL22_C.albicansconidia_PBMC_7days | -0.54952646 | 0.003058006 |
| rs34754340 | 3 | 46041837 | T | C | 0.5256571 | 1.04E-17 | IL22_C.albicansconidia_PBMC_7days | -0.54747236 | 0.00315361 |
| rs34836513 | 3 | 46046459 | A | G | 0.5263958 | 9.51E-18 | IL22_C.albicansconidia_PBMC_7days | -0.54234187 | 0.003395566 |
| rs2088693 | 3 | 46077390 | C | T | 0.4958297 | 1.42E-17 | IL22_C.albicansconidia_PBMC_7days | -0.47600344 | 0.006025543 |
| rs58552652 | 3 | 46078292 | T | C | 0.4959028 | 1.40E-17 | IL22_C.albicansconidia_PBMC_7days | -0.47580855 | 0.00604634 |
| rs13087776 | 3 | 46079871 | G | A | 0.4931951 | 2.14E-17 | IL22_C.albicansconidia_PBMC_7days | -0.47546069 | 0.006082605 |
| rs13068410 | 3 | 46080176 | A | T | 0.4959272 | 1.38E-17 | IL22_C.albicansconidia_PBMC_7days | -0.47546069 | 0.006082605 |
| rs13094262 | 3 | 46080912 | T | C | 0.4960064 | 1.37E-17 | IL22_C.albicansconidia_PBMC_7days | -0.47546069 | 0.006082605 |
| rs34805664 | 3 | 46081108 | T | A | 0.5056844 | 2.69E-18 | IL22_C.albicansconidia_PBMC_7days | -0.4752636 | 0.006103698 |
| rs35028116 | 3 | 46081333 | C | T | 0.4961586 | 1.35E-17 | IL22_C.albicansconidia_PBMC_7days | -0.4752636 | 0.006103698 |
| rs34610609 | 3 | 46081707 | T | C | 0.4963534 | 1.31E-17 | IL22_C.albicansconidia_PBMC_7days | -0.4752636 | 0.006103698 |
| rs34254987 | 3 | 46083634 | A | G | 0.4959698 | 1.38E-17 | IL22_C.albicansconidia_PBMC_7days | -0.4750654 | 0.006124917 |
| rs4683163 | 3 | 46084372 | C | A | 0.495982 | 1.37E-17 | IL22_C.albicansconidia_PBMC_7days | -0.4750654 | 0.006124917 |
| rs4683164 | 3 | 46084461 | G | A | 0.4945133 | 1.71E-17 | IL22_C.albicansconidia_PBMC_7days | -0.47486611 | 0.006146262 |
| rs34155121 | 3 | 46084742 | A | G | 0.4955069 | 1.50E-17 | IL22_C.albicansconidia_PBMC_7days | -0.47486611 | 0.006146262 |
| rs4234453 | 3 | 46084949 | A | C | 0.4954216 | 1.50E-17 | IL22_C.albicansconidia_PBMC_7days | -0.47486611 | 0.006146262 |
| rs12108042 | 3 | 46086083 | G | A | 0.4961038 | 1.36E-17 | IL22_C.albicansconidia_PBMC_7days | -0.47486611 | 0.006146262 |
| rs34867672 | 3 | 46088282 | C | T | 0.4970593 | 1.20E-17 | IL22_C.albicansconidia_PBMC_7days | -0.47446423 | 0.006189333 |
| rs34924300 | 3 | 46088336 | T | G | 0.4970776 | 1.20E-17 | IL22_C.albicansconidia_PBMC_7days | -0.47446423 | 0.006189333 |
| rs7642229 | 3 | 46090557 | G | A | 0.4959576 | 1.47E-17 | IL22_C.albicansconidia_PBMC_7days | -0.47390561 | 0.006215782 |
| rs7652478 | 3 | 46090477 | A | G | 0.4976614 | 1.11E-17 | IL22_C.albicansconidia_PBMC_7days | -0.47370032 | 0.006270725 |
| rs7642320 | 3 | 46090622 | G | A | 0.4978316 | 1.08E-17 | IL22_C.albicansconidia_PBMC_7days | -0.47370032 | 0.006270725 |
| rs13088143 | 3 | 46080024 | T | A | 0.4959637 | 1.38E-17 | IL22_C.albicansconidia_PBMC_7days | -0.47402328 | 0.006487686 |
| rs4683162 | 3 | 46083941 | T | G | 0.4971505 | 1.12E-17 | IL22_C.albicansconidia_PBMC_7days | -0.46939533 | 0.006718722 |
| rs9845542 | 3 | 46111869 | A | G | 0.3328729 | 1.75E-09 | IL22_C.albicansconidia_PBMC_7days | -0.31262163 | 0.04039365 |
| rs35951367 | 3 | 46118439 | C | T | 0.3240092 | 5.94E-09 | IL22_C.albicansconidia_PBMC_7days | -0.31172104 | 0.040930116 |
| rs4443214 | 3 | 46177864 | C | T | 0.4456917 | 5.27E-12 | IL6_A.fumigatusconidiaSerum_PBMC_24h | -0.39642457 | 0.016631753 |
| rs2109069 | 19 | 4719443 | A | G | 0.3056227 | 3.98E-12 | IL6_A.fumigatusconidiaSerum_PBMC_24h | -0.25170374 | 0.017679114 |
| rs9838450 | 3 | 46123333 | C | A | 0.3461396 | 7.25E-10 | IL6_A.fumigatusconidiaSerum_PBMC_24h | -0.34894446 | 0.018860126 |
| rs4493469 | 3 | 46177992 | C | T | 0.4298715 | 8.27E-11 | IL6_A.fumigatusconidiaSerum_PBMC_24h | -0.39316539 | 0.019698594 |
| rs34836513 | 3 | 46046459 | A | G | 0.5263958 | 9.51E-18 | IL6_A.fumigatusconidiaSerum_PBMC_24h | -0.36604489 | 0.031311893 |
| rs34754340 | 3 | 46041837 | T | C | 0.5256571 | 1.04E-17 | IL6_A.fumigatusconidiaSerum_PBMC_24h | -0.36566199 | 0.031783607 |
| rs71615438 | 3 | 46039868 | G | A | 0.5258581 | 9.97E-18 | IL6_A.fumigatusconidiaSerum_PBMC_24h | -0.36522565 | 0.032087674 |
| rs17330872 | 3 | 46035097 | G | A | 0.527665 | 7.34E-18 | IL6_A.fumigatusconidiaSerum_PBMC_24h | -0.36410047 | 0.032769909 |
| rs9845542 | 3 | 46111869 | A | G | 0.3328729 | 1.75E-09 | IL6_A.fumigatusconidiaSerum_PBMC_24h | -0.29394808 | 0.035368317 |
| rs35951367 | 3 | 46118439 | C | T | 0.3240092 | 5.94E-09 | IL6_A.fumigatusconidiaSerum_PBMC_24h | -0.29335041 | 0.035702504 |
| rs9877748 | 3 | 46111081 | G | A | 0.5070104 | 1.95E-16 | IL6_A.fumigatusconidiaSerum_PBMC_24h | -0.35475511 | 0.035711146 |
| rs36122610 | 3 | 46022833 | A | G | 0.5300752 | 4.70E-18 | IL6_A.fumigatusconidiaSerum_PBMC_24h | -0.35601301 | 0.036490392 |
| rs34442130 | 3 | 46024529 | A | T | 0.5301222 | 4.67E-18 | IL6_A.fumigatusconidiaSerum_PBMC_24h | -0.3560114 | 0.03649144 |
| rs13075758 | 3 | 46025048 | A | G | 0.5301634 | 4.64E-18 | IL6_A.fumigatusconidiaSerum_PBMC_24h | -0.35593563 | 0.036520469 |
| rs35669129 | 3 | 46055250 | A | G | 0.5281192 | 6.78E-18 | IL6_A.fumigatusconidiaSerum_PBMC_24h | -0.35500292 | 0.036689575 |
| rs13063033 | 3 | 46082481 | A | G | 0.4971019 | 7.78E-16 | IL6_A.fumigatusconidiaSerum_PBMC_24h | -0.32978361 | 0.03676413 |
| rs13081213 | 3 | 46055909 | T | C | 0.5281664 | 6.72E-18 | IL6_A.fumigatusconidiaSerum_PBMC_24h | -0.35418827 | 0.037087634 |
| rs35751180 | 3 | 46056173 | T | C | 0.5342276 | 4.95E-18 | IL6_A.fumigatusconidiaSerum_PBMC_24h | -0.35351761 | 0.037335399 |
| rs34168660 | 3 | 46056162 | A | G | 0.5435793 | 1.04E-18 | IL6_A.fumigatusconidiaSerum_PBMC_24h | -0.35360583 | 0.037377714 |
| rs4362758 | 3 | 46056911 | A | T | 0.5157296 | 5.39E-17 | IL6_A.fumigatusconidiaSerum_PBMC_24h | -0.35264403 | 0.037860033 |
| rs73833520 | 3 | 46057867 | G | A | 0.5115713 | 8.68E-17 | IL6_A.fumigatusconidiaSerum_PBMC_24h | -0.34103833 | 0.04184029 |
| rs13091868 | 3 | 46123043 | G | A | 0.509687 | 2.63E-16 | IL6_A.fumigatusconidiaSerum_PBMC_24h | -0.33602075 | 0.04233629 |
| rs13092030 | 3 | 46123055 | T | C | 0.5103595 | 2.38E-16 | IL6_A.fumigatusconidiaSerum_PBMC_24h | -0.33602075 | 0.04233629 |
| rs17215981 | 3 | 46124418 | G | A | 0.4971931 | 1.04E-15 | IL6_A.fumigatusconidiaSerum_PBMC_24h | -0.33602075 | 0.04233629 |
| rs13089554 | 3 | 46127557 | A | G | 0.4966395 | 1.13E-15 | IL6_A.fumigatusconidiaSerum_PBMC_24h | -0.33602075 | 0.04233629 |
| rs13095717 | 3 | 46128542 | A | G | 0.4967673 | 1.11E-15 | IL6_A.fumigatusconidiaSerum_PBMC_24h | -0.33602075 | 0.04233629 |
| rs13075528 | 3 | 46128709 | C | T | 0.5001448 | 7.62E-16 | IL6_A.fumigatusconidiaSerum_PBMC_24h | -0.33602075 | 0.04233629 |

| | | | | | | | | | |
|---|---|---|---|---|---|---|---|---|---|
| rs13095602 | 3 | 46128713 | G | A | 0.4997202 | 8.04E-16 | IL6_A.fumigatusconidiaSerum_PBMC_24h | -0.33602075 | 0.04233629 |
| rs71327023 | 3 | 46131225 | G | C | 0.49419 | 1.69E-15 | IL6_A.fumigatusconidiaSerum_PBMC_24h | -0.33602075 | 0.04233629 |
| rs13063527 | 3 | 46135690 | A | G | 0.4983724 | 9.69E-16 | IL6_A.fumigatusconidiaSerum_PBMC_24h | -0.33602075 | 0.04233629 |
| rs4373103 | 3 | 46138455 | A | G | 0.4974729 | 1.14E-15 | IL6_A.fumigatusconidiaSerum_PBMC_24h | -0.33602075 | 0.04233629 |
| rs71327024 | 3 | 46140073 | T | G | 0.5013388 | 6.45E-16 | IL6_A.fumigatusconidiaSerum_PBMC_24h | -0.33602075 | 0.04233629 |
| rs71327017 | 3 | 46111213 | C | T | 0.4985668 | 8.96E-16 | IL6_A.fumigatusconidiaSerum_PBMC_24h | -0.33592105 | 0.042403215 |
| rs9845382 | 3 | 46111759 | A | G | 0.5066369 | 2.03E-16 | IL6_A.fumigatusconidiaSerum_PBMC_24h | -0.33592105 | 0.042403215 |
| rs13069742 | 3 | 46114216 | G | A | 0.5064079 | 2.08E-16 | IL6_A.fumigatusconidiaSerum_PBMC_24h | -0.33592105 | 0.042403215 |
| rs13072267 | 3 | 46116010 | A | T | 0.4979167 | 9.53E-16 | IL6_A.fumigatusconidiaSerum_PBMC_24h | -0.33592105 | 0.042403215 |
| rs36057789 | 3 | 46117096 | G | A | 0.4975033 | 1.01E-15 | IL6_A.fumigatusconidiaSerum_PBMC_24h | -0.33592105 | 0.042403215 |
| rs76647202 | 3 | 46117960 | G | A | 0.4978073 | 9.57E-16 | IL6_A.fumigatusconidiaSerum_PBMC_24h | -0.33592105 | 0.042403215 |
| rs71327021 | 3 | 46120517 | G | A | 0.4978316 | 9.43E-16 | IL6_A.fumigatusconidiaSerum_PBMC_24h | -0.33592105 | 0.042403215 |
| rs9824651 | 3 | 46108285 | T | C | 0.5120269 | 9.50E-17 | IL6_A.fumigatusconidiaSerum_PBMC_24h | -0.33582049 | 0.042470498 |
| rs9825081 | 3 | 46108400 | A | G | 0.5118771 | 9.67E-17 | IL6_A.fumigatusconidiaSerum_PBMC_24h | -0.33582049 | 0.042470498 |
| rs34460587 | 3 | 46142464 | T | C | 0.5045379 | 4.48E-16 | IL6_A.fumigatusconidiaSerum_PBMC_24h | -0.33592594 | 0.042533821 |
| rs4683162 | 3 | 46083941 | T | G | 0.4971505 | 1.12E-17 | IL6_A.fumigatusconidiaSerum_PBMC_24h | -0.31795938 | 0.045489722 |
| rs71327006 | 3 | 46058908 | A | G | 0.5044715 | 3.14E-16 | IL6_A.fumigatusconidiaSerum_PBMC_24h | -0.33117483 | 0.046425427 |
| rs71327007 | 3 | 46059249 | T | C | 0.5044473 | 3.14E-16 | IL6_A.fumigatusconidiaSerum_PBMC_24h | -0.33063446 | 0.046757395 |
| rs35454877 | 3 | 46059484 | C | T | 0.5121288 | 7.99E-17 | IL6_A.fumigatusconidiaSerum_PBMC_24h | -0.33031421 | 0.046958494 |
| rs1491951 | 3 | 46141844 | A | G | 0.5083165 | 2.07E-16 | IL6_A.fumigatusconidiaSerum_PBMC_24h | -0.32508073 | 0.047459586 |
| rs2088693 | 3 | 46077390 | C | T | 0.4958297 | 1.42E-17 | IL6_A.fumigatusconidiaSerum_PBMC_24h | -0.31313774 | 0.049094802 |
| rs58552652 | 3 | 46078292 | T | C | 0.4959028 | 1.40E-17 | IL6_A.fumigatusconidiaSerum_PBMC_24h | -0.3130405 | 0.049165149 |
| rs13087776 | 3 | 46079871 | G | A | 0.4931951 | 2.14E-17 | IL6_A.fumigatusconidiaSerum_PBMC_24h | -0.31281952 | 0.049320993 |
| rs13068410 | 3 | 46080176 | A | T | 0.4959272 | 1.38E-17 | IL6_A.fumigatusconidiaSerum_PBMC_24h | -0.31281952 | 0.049320993 |
| rs13094262 | 3 | 46080912 | T | C | 0.4960064 | 1.37E-17 | IL6_A.fumigatusconidiaSerum_PBMC_24h | -0.31281952 | 0.049320993 |
| rs35334665 | 3 | 46061218 | C | T | 0.5042903 | 3.19E-16 | IL6_A.fumigatusconidiaSerum_PBMC_24h | -0.32658951 | 0.049321313 |
| rs34805664 | 3 | 46081108 | T | A | 0.5056844 | 2.69E-18 | IL6_A.fumigatusconidiaSerum_PBMC_24h | -0.31272083 | 0.04939214 |
| rs35028116 | 3 | 46081333 | C | T | 0.4961586 | 1.35E-17 | IL6_A.fumigatusconidiaSerum_PBMC_24h | -0.31272083 | 0.04939214 |
| rs34610609 | 3 | 46081707 | T | C | 0.4963534 | 1.31E-17 | IL6_A.fumigatusconidiaSerum_PBMC_24h | -0.31272083 | 0.04939214 |
| rs34254987 | 3 | 46083634 | A | G | 0.4959698 | 1.38E-17 | IL6_A.fumigatusconidiaSerum_PBMC_24h | -0.31262142 | 0.049463639 |
| rs4683163 | 3 | 46084372 | C | A | 0.495982 | 1.37E-17 | IL6_A.fumigatusconidiaSerum_PBMC_24h | -0.31262142 | 0.049463639 |
| rs36040135 | 3 | 46061304 | G | A | 0.5113495 | 8.88E-17 | IL6_A.fumigatusconidiaSerum_PBMC_24h | -0.32635789 | 0.049463788 |
| rs4683164 | 3 | 46084461 | G | A | 0.4945133 | 1.71E-17 | IL6_A.fumigatusconidiaSerum_PBMC_24h | -0.31252129 | 0.049535489 |
| rs34155121 | 3 | 46084742 | A | G | 0.4955069 | 1.50E-17 | IL6_A.fumigatusconidiaSerum_PBMC_24h | -0.31252129 | 0.049535489 |
| rs4234453 | 3 | 46084949 | A | C | 0.4954216 | 1.50E-17 | IL6_A.fumigatusconidiaSerum_PBMC_24h | -0.31252129 | 0.049535489 |
| rs12108042 | 3 | 46086083 | G | A | 0.4961038 | 1.36E-17 | IL6_A.fumigatusconidiaSerum_PBMC_24h | -0.31252129 | 0.049535489 |
| rs34867672 | 3 | 46088282 | C | T | 0.4970593 | 1.20E-17 | IL6_A.fumigatusconidiaSerum_PBMC_24h | -0.31231885 | 0.04968025 |
| rs34924300 | 3 | 46088336 | T | G | 0.4970776 | 1.20E-17 | IL6_A.fumigatusconidiaSerum_PBMC_24h | -0.31231885 | 0.04968025 |
| rs13088143 | 3 | 46080024 | T | A | 0.4959637 | 1.38E-17 | IL6_A.fumigatusconidiaSerum_PBMC_24h | -0.31363603 | 0.049793898 |
| rs71327014 | 3 | 46101466 | T | C | 0.5076245 | 2.27E-16 | IL6_A.fumigatusconidiaSerum_PBMC_24h | -0.32627508 | 0.049936897 |
| rs71327015 | 3 | 46102173 | C | G | 0.5043145 | 4.35E-16 | IL6_A.fumigatusconidiaSerum_PBMC_24h | -0.32627508 | 0.049936897 |
| rs34047915 | 3 | 46100491 | T | C | 0.5072994 | 2.36E-16 | IL6_A.fumigatusconidiaSerum_PBMC_24h | -0.32625409 | 0.049955725 |
| rs35420565 | 3 | 46100138 | G | C | 0.5074198 | 2.32E-16 | IL6_A.fumigatusconidiaSerum_PBMC_24h | -0.32623226 | 0.049974859 |
| rs7652478 | 3 | 46090477 | A | G | 0.4976614 | 1.11E-17 | IL6_A.fumigatusconidiaSerum_PBMC_24h | -0.31188608 | 0.049986399 |
| rs7642320 | 3 | 46090622 | G | A | 0.4978316 | 1.08E-17 | IL6_A.fumigatusconidiaSerum_PBMC_24h | -0.31188608 | 0.049986399 |
| rs12705891 | 7 | 113317708 | C | T | 0.2753944 | 2.35E-10 | IL6_C.albicansconidia_PBMC_24h | 0.10500127 | 0.045933403 |
| rs9838450 | 3 | 46123333 | C | A | 0.3461396 | 7.25E-10 | IL6_C.burnetiininemileSerum_PBMC_24h | -0.18773669 | 0.024721135 |
| rs2109069 | 19 | 4719443 | A | G | 0.3056227 | 3.98E-12 | IL6_Cryptococcus_PBMC_24h | -0.43849043 | 0.023407941 |
| rs12705891 | 7 | 113317708 | C | T | 0.2753944 | 2.35E-10 | IL6_E.Coli_PBMC_24h | -0.17202437 | 0.018756444 |
| rs2109069 | 19 | 4719443 | A | G | 0.3056227 | 3.98E-12 | IL6_Influenza_PBMC_24h | -0.3948762 | 0.044475318 |
| rs3851347 | 3 | 46053778 | G | A | 0.2605395 | 4.30E-09 | IL6_LPS_macroPG_24h | -0.167785 | 0.046157366 |
| rs36122610 | 3 | 46022833 | A | G | 0.5300752 | 4.70E-18 | IL6_LPS100ng_PBMC_24h | -0.24634967 | 0.016412853 |
| rs34442130 | 3 | 46024529 | A | T | 0.5301222 | 4.67E-18 | IL6_LPS100ng_PBMC_24h | -0.24629561 | 0.016436759 |
| rs13075758 | 3 | 46025048 | A | G | 0.5301634 | 4.64E-18 | IL6_LPS100ng_PBMC_24h | -0.24627007 | 0.016441633 |
| rs17330872 | 3 | 46035097 | G | A | 0.527665 | 7.34E-18 | IL6_LPS100ng_PBMC_24h | -0.23887363 | 0.020254553 |
| rs71615438 | 3 | 46039868 | G | A | 0.5258581 | 9.97E-18 | IL6_LPS100ng_PBMC_24h | -0.23634601 | 0.021516651 |
| rs34754340 | 3 | 46041837 | T | C | 0.5256571 | 1.04E-17 | IL6_LPS100ng_PBMC_24h | -0.23520545 | 0.022082841 |
| rs34836513 | 3 | 46046459 | A | G | 0.5263958 | 9.51E-18 | IL6_LPS100ng_PBMC_24h | -0.23240961 | 0.023484604 |
| rs35669129 | 3 | 46055250 | A | G | 0.5281192 | 6.78E-18 | IL6_LPS100ng_PBMC_24h | -0.23171638 | 0.023809114 |
| rs13081213 | 3 | 46055909 | T | C | 0.5281664 | 6.72E-18 | IL6_LPS100ng_PBMC_24h | -0.23166322 | 0.023813451 |
| rs34168660 | 3 | 46056162 | A | G | 0.5435793 | 1.04E-18 | IL6_LPS100ng_PBMC_24h | -0.2315442 | 0.02386828 |
| rs4362758 | 3 | 46056911 | A | T | 0.5157296 | 5.39E-17 | IL6_LPS100ng_PBMC_24h | -0.23143883 | 0.023902514 |
| rs35751180 | 3 | 46056173 | T | C | 0.5342276 | 4.95E-18 | IL6_LPS100ng_PBMC_24h | -0.23108228 | 0.024084677 |
| rs9877748 | 3 | 46111081 | G | A | 0.5070104 | 1.95E-16 | IL6_LPS100ng_PBMC_24h | -0.22519849 | 0.027149952 |
| rs13063033 | 3 | 46082481 | A | G | 0.4971019 | 7.78E-16 | IL6_LPS100ng_PBMC_24h | -0.20919887 | 0.028138756 |
| rs9838450 | 3 | 46123333 | C | A | 0.3461396 | 7.25E-10 | IL6_LPS100ng_PBMC_24h | -0.18528942 | 0.038998646 |
| rs73833520 | 3 | 46057867 | G | A | 0.5115713 | 8.68E-17 | IL6_LPS100ng_PBMC_24h | -0.20343781 | 0.044315986 |
| rs77399277 | 3 | 46096798 | C | T | 0.509008 | 1.80E-16 | IL6_LPS100ng_PBMC_24h | -0.20133905 | 0.045046459 |
| rs34127208 | 3 | 46103680 | T | G | 0.5047371 | 4.15E-16 | IL6_LPS100ng_PBMC_24h | -0.20129305 | 0.045067522 |
| rs34718164 | 3 | 46098526 | G | C | 0.5071188 | 2.42E-16 | IL6_LPS100ng_PBMC_24h | -0.20130725 | 0.045075691 |
| rs13096741 | 3 | 46099154 | T | C | 0.5072512 | 2.37E-16 | IL6_LPS100ng_PBMC_24h | -0.20129344 | 0.045086574 |
| rs34863575 | 3 | 46094063 | G | A | 0.5076787 | 2.15E-16 | IL6_LPS100ng_PBMC_24h | -0.20128756 | 0.045088082 |
| rs35772789 | 3 | 46095104 | G | A | 0.5079254 | 2.08E-16 | IL6_LPS100ng_PBMC_24h | -0.2012914 | 0.045092995 |
| rs34766614 | 3 | 46107601 | G | A | 0.500351 | 7.64E-16 | IL6_LPS100ng_PBMC_24h | -0.20125588 | 0.045102575 |
| rs35420565 | 3 | 46100138 | G | C | 0.5074198 | 2.32E-16 | IL6_LPS100ng_PBMC_24h | -0.20125956 | 0.045111715 |
| rs34047915 | 3 | 46100491 | T | C | 0.5072994 | 2.36E-16 | IL6_LPS100ng_PBMC_24h | -0.20124977 | 0.045118705 |
| rs34679077 | 3 | 46087992 | A | G | 0.5051656 | 2.90E-16 | IL6_LPS100ng_PBMC_24h | -0.20116663 | 0.045121564 |
| rs71327014 | 3 | 46101466 | T | C | 0.5076245 | 2.27E-16 | IL6_LPS100ng_PBMC_24h | -0.20123947 | 0.045125975 |
| rs71327015 | 3 | 46102173 | C | G | 0.5043145 | 4.35E-16 | IL6_LPS100ng_PBMC_24h | -0.20123947 | 0.045125975 |
| rs72901034 | 3 | 46085321 | G | A | 0.4961586 | 1.34E-17 | IL6_LPS100ng_PBMC_24h | -0.20108567 | 0.045170912 |
| rs34438204 | 3 | 46081306 | C | T | 0.504556 | 3.11E-16 | IL6_LPS100ng_PBMC_24h | -0.20099217 | 0.045210696 |
| rs2088692 | 3 | 46077332 | G | A | 0.5114274 | 8.77E-17 | IL6_LPS100ng_PBMC_24h | -0.20085949 | 0.045269877 |
| rs6791016 | 3 | 46074858 | A | G | 0.5114394 | 8.76E-17 | IL6_LPS100ng_PBMC_24h | -0.20075662 | 0.045335239 |
| rs6780028 | 3 | 46075090 | C | A | 0.5114934 | 8.69E-17 | IL6_LPS100ng_PBMC_24h | -0.20075662 | 0.045335239 |
| rs6790866 | 3 | 46074711 | A | G | 0.5114514 | 8.74E-17 | IL6_LPS100ng_PBMC_24h | -0.20073406 | 0.045345663 |
| rs35814488 | 3 | 46073923 | G | A | 0.5114394 | 8.75E-17 | IL6_LPS100ng_PBMC_24h | -0.20071097 | 0.045356368 |
| rs34774687 | 3 | 46073314 | C | T | 0.5114574 | 8.73E-17 | IL6_LPS100ng_PBMC_24h | -0.20068738 | 0.045367354 |
| rs34619093 | 3 | 46072519 | A | G | 0.5041695 | 3.25E-16 | IL6_LPS100ng_PBMC_24h | -0.20066326 | 0.04537862 |
| rs68087193 | 3 | 46072630 | T | C | 0.5113135 | 8.97E-17 | IL6_LPS100ng_PBMC_24h | -0.20066326 | 0.04537862 |
| rs2102055 | 3 | 46072704 | A | G | 0.5140265 | 4.75E-17 | IL6_LPS100ng_PBMC_24h | -0.20066326 | 0.04537862 |
| rs2102056 | 3 | 46072768 | A | G | 0.5114095 | 8.83E-17 | IL6_LPS100ng_PBMC_24h | -0.20066326 | 0.04537862 |
| rs2088690 | 3 | 46072839 | T | C | 0.5114095 | 8.83E-17 | IL6_LPS100ng_PBMC_24h | -0.20066326 | 0.04537862 |
| rs13060287 | 3 | 46071868 | G | A | 0.5111396 | 9.18E-17 | IL6_LPS100ng_PBMC_24h | -0.20062085 | 0.045414428 |
| rs13082697 | 3 | 46071763 | C | T | 0.5096029 | 1.21E-16 | IL6_LPS100ng_PBMC_24h | -0.20059572 | 0.04542626 |
| rs34562820 | 3 | 46071081 | A | G | 0.5041333 | 3.27E-16 | IL6_LPS100ng_PBMC_24h | -0.20057007 | 0.045438372 |
| rs59166269 | 3 | 46070337 | G | C | 0.5113075 | 8.94E-17 | IL6_LPS100ng_PBMC_24h | -0.2005439 | 0.045450766 |
| rs13090194 | 3 | 46069573 | C | A | 0.5115833 | 8.64E-17 | IL6_LPS100ng_PBMC_24h | -0.20051723 | 0.04546344 |
| rs71327010 | 3 | 46068835 | T | G | 0.5039037 | 3.39E-16 | IL6_LPS100ng_PBMC_24h | -0.20046235 | 0.045489633 |
| rs35159820 | 3 | 46068473 | G | A | 0.5112475 | 9.02E-17 | IL6_LPS100ng_PBMC_24h | -0.20043414 | 0.04550315 |
| rs35161099 | 3 | 46067507 | G | T | 0.5040789 | 3.29E-16 | IL6_LPS100ng_PBMC_24h | -0.20040542 | 0.045516949 |
| rs35930050 | 3 | 46065763 | A | G | 0.5113015 | 8.93E-17 | IL6_LPS100ng_PBMC_24h | -0.20028546 | 0.045574956 |
| rs34584867 | 3 | 46065963 | A | G | 0.5112955 | 8.94E-17 | IL6_LPS100ng_PBMC_24h | -0.20028546 | 0.045574956 |
| rs35334665 | 3 | 46061218 | C | T | 0.5042903 | 3.19E-16 | IL6_LPS100ng_PBMC_24h | -0.20043668 | 0.045618593 |
| rs13097556 | 3 | 46061997 | C | T | 0.5096389 | 1.23E-16 | IL6_LPS100ng_PBMC_24h | -0.20033023 | 0.045622742 |
| rs876668 | 3 | 46063753 | C | T | 0.5112415 | 9.01E-17 | IL6_LPS100ng_PBMC_24h | -0.20015737 | 0.045637464 |
| rs36040135 | 3 | 46061304 | G | A | 0.5113495 | 8.88E-17 | IL6_LPS100ng_PBMC_24h | -0.20039904 | 0.045640771 |
| rs35454877 | 3 | 46059484 | C | T | 0.5121288 | 7.99E-17 | IL6_LPS100ng_PBMC_24h | -0.20054186 | 0.045685019 |
| rs71327006 | 3 | 46058908 | A | G | 0.5044715 | 3.14E-16 | IL6_LPS100ng_PBMC_24h | -0.20057166 | 0.045693047 |
| rs71327007 | 3 | 46059249 | T | C | 0.5044473 | 3.14E-16 | IL6_LPS100ng_PBMC_24h | -0.20054178 | 0.045698042 |
| rs2230322 | 3 | 46063329 | C | T | 0.5110856 | 9.20E-17 | IL6_LPS100ng_PBMC_24h | -0.19868669 | 0.045764183 |
| rs2109069 | 19 | 4719443 | A | G | 0.3056227 | 3.98E-12 | IL6_MSUC16_PBMC_24h | -0.18892984 | 0.044978066 |
| rs3851347 | 3 | 46053778 | G | A | 0.2605395 | 4.30E-09 | IL6_MTB_macroPG_24h | -0.25019388 | 0.027371315 |
| rs6764042 | 3 | 46059139 | G | T | 0.2550607 | 9.68E-09 | IL6_MTB_macroPG_24h | -0.22330895 | 0.048727079 |
| rs7623460 | 3 | 46068764 | C | A | 0.2603006 | 4.93E-09 | IL6_MTB_macroPG_24h | -0.22324046 | 0.049048019 |
| rs1392290 | 3 | 46069210 | A | G | 0.2486631 | 2.01E-08 | IL6_MTB_macroPG_24h | -0.22319739 | 0.049097386 |
| rs6805094 | 3 | 46071463 | G | C | 0.25647 | 7.95E-09 | IL6_MTB_macroPG_24h | -0.22301873 | 0.049307442 |
| rs13060901 | 3 | 46072006 | A | G | 0.2564932 | 7.93E-09 | IL6_MTB_macroPG_24h | -0.22297409 | 0.049357832 |
| rs4411920 | 3 | 46077525 | G | A | 0.2565551 | 7.89E-09 | IL6_MTB_macroPG_24h | -0.2226921 | 0.049696906 |
| rs6764042 | 3 | 46059139 | G | T | 0.2550607 | 9.68E-09 | IL6_Pam3Cys_PBMC_24h | 0.14238688 | 0.018346285 |
| rs13060901 | 3 | 46072006 | A | G | 0.2564932 | 7.93E-09 | IL6_Pam3Cys_PBMC_24h | 0.14148941 | 0.019238228 |
| rs4411920 | 3 | 46077525 | G | A | 0.2565551 | 7.89E-09 | IL6_Pam3Cys_PBMC_24h | 0.14149374 | 0.01926029 |
| rs6805094 | 3 | 46071463 | G | C | 0.25647 | 7.95E-09 | IL6_Pam3Cys_PBMC_24h | 0.14145894 | 0.019261858 |
| rs1392290 | 3 | 46069210 | A | G | 0.2486631 | 2.01E-08 | IL6_Pam3Cys_PBMC_24h | 0.14142335 | 0.019278624 |
| rs7623460 | 3 | 46068764 | C | A | 0.2603006 | 4.93E-09 | IL6_Pam3Cys_PBMC_24h | 0.14139191 | 0.019302717 |
| rs9849015 | 3 | 46056120 | C | T | 0.2579159 | 6.65E-09 | IL6_Pam3Cys_PBMC_24h | 0.14046263 | 0.020865344 |
| rs7623190 | 3 | 46068325 | C | T | 0.2556107 | 8.91E-09 | IL6_Pam3Cys_PBMC_24h | 0.13914734 | 0.021953652 |
| rs3851347 | 3 | 46053778 | G | A | 0.2605395 | 4.30E-09 | IL6_Pam3Cys_PBMC_24h | 0.12428326 | 0.040010997 |
| rs9845542 | 3 | 46111869 | A | G | 0.3328729 | 1.75E-09 | IL6_S.aureus_WB_48h | -0.22130739 | 0.027940074 |
| rs35951367 | 3 | 46118439 | C | T | 0.3240092 | 5.94E-09 | IL6_S.aureus_WB_48h | -0.22047148 | 0.02850311 |
| rs34906043 | 3 | 46094623 | T | C | 0.2717585 | 1.66E-09 | IL6_S.aureus_WB_48h | 0.16749927 | 0.040358854 |
| rs35657218 | 3 | 46094462 | G | A | 0.2755462 | 9.52E-10 | IL6_S.aureus_WB_48h | 0.16743041 | 0.040449627 |
| rs13098271 | 3 | 46093858 | A | G | 0.277071 | 7.60E-10 | IL6_S.aureus_WB_48h | 0.16731978 | 0.040585503 |
| rs7642701 | 3 | 46091058 | G | A | 0.2750678 | 9.82E-10 | IL6_S.aureus_WB_48h | 0.16675434 | 0.041303553 |
| rs2109069 | 19 | 4719443 | A | G | 0.3056227 | 3.98E-12 | IL6_S.typhimurium_macroPG_24h | 0.17357319 | 0.007838692 |
| rs35751180 | 3 | 46056173 | T | C | 0.5342276 | 4.95E-18 | TNFA_C.conidiaHK_WB_48h | -0.30473075 | 0.021070308 |
| rs4362758 | 3 | 46056911 | A | T | 0.5157296 | 5.39E-17 | TNFA_C.conidiaHK_WB_48h | -0.30444842 | 0.021223144 |
| rs34168660 | 3 | 46056162 | A | G | 0.5435793 | 1.04E-18 | TNFA_C.conidiaHK_WB_48h | -0.30400467 | 0.02144309 |
| rs13081213 | 3 | 46055909 | T | C | 0.5281664 | 6.72E-18 | TNFA_C.conidiaHK_WB_48h | -0.30398562 | 0.021467133 |
| rs35669129 | 3 | 46055250 | A | G | 0.5281192 | 6.78E-18 | TNFA_C.conidiaHK_WB_48h | -0.30401233 | 0.021481851 |
| rs34836513 | 3 | 46046459 | A | G | 0.5263958 | 9.51E-18 | TNFA_C.conidiaHK_WB_48h | -0.29841737 | 0.024123833 |
| rs13063033 | 3 | 46082481 | A | G | 0.4971019 | 7.78E-16 | TNFA_C.conidiaHK_WB_48h | -0.27375577 | 0.026041052 |
| rs34754340 | 3 | 46041837 | T | C | 0.5256571 | 1.04E-17 | TNFA_C.conidiaHK_WB_48h | -0.29486867 | 0.026139055 |
| rs71615438 | 3 | 46039868 | G | A | 0.5258581 | 9.97E-18 | TNFA_C.conidiaHK_WB_48h | -0.29337004 | 0.027006979 |
| rs9877748 | 3 | 46111081 | G | A | 0.5070104 | 1.95E-16 | TNFA_C.conidiaHK_WB_48h | -0.2878582 | 0.028627284 |
| rs73833520 | 3 | 46057867 | G | A | 0.5115713 | 8.68E-17 | TNFA_C.conidiaHK_WB_48h | -0.28555283 | 0.028666097 |
| rs17330872 | 3 | 46035097 | G | A | 0.527665 | 7.34E-18 | TNFA_C.conidiaHK_WB_48h | -0.28906016 | 0.029499172 |
| rs876668 | 3 | 46063753 | C | T | 0.5112415 | 9.01E-17 | TNFA_C.conidiaHK_WB_48h | -0.28091886 | 0.02969526 |
| rs35930050 | 3 | 46065763 | A | G | 0.5113015 | 8.93E-17 | TNFA_C.conidiaHK_WB_48h | -0.28067926 | 0.029902021 |
| rs34584867 | 3 | 46065963 | A | G | 0.5112955 | 8.94E-17 | TNFA_C.conidiaHK_WB_48h | -0.28067926 | 0.029902021 |
| rs13097556 | 3 | 46061997 | C | T | 0.5096389 | 1.23E-16 | TNFA_C.conidiaHK_WB_48h | -0.28069379 | 0.029951351 |
| rs35161099 | 3 | 46067507 | G | T | 0.5040789 | 3.29E-16 | TNFA_C.conidiaHK_WB_48h | -0.28042753 | 0.030113621 |
| rs35159820 | 3 | 46068473 | G | A | 0.5112475 | 9.02E-17 | TNFA_C.conidiaHK_WB_48h | -0.28036271 | 0.030167285 |
| rs71327010 | 3 | 46068835 | T | G | 0.5039037 | 3.39E-16 | TNFA_C.conidiaHK_WB_48h | -0.28029712 | 0.030221257 |
| rs36040135 | 3 | 46061304 | G | A | 0.5113495 | 8.88E-17 | TNFA_C.conidiaHK_WB_48h | -0.28034461 | 0.030226953 |
| rs35334665 | 3 | 46061218 | C | T | 0.5042903 | 3.19E-16 | TNFA_C.conidiaHK_WB_48h | -0.28032956 | 0.030250545 |
| rs13090194 | 3 | 46069573 | C | A | 0.5115833 | 8.64E-17 | TNFA_C.conidiaHK_WB_48h | -0.28016369 | 0.030330127 |
| rs59166269 | 3 | 46070337 | G | C | 0.5113075 | 8.94E-17 | TNFA_C.conidiaHK_WB_48h | -0.28009584 | 0.030385028 |
| rs34562820 | 3 | 46071081 | A | G | 0.5041333 | 3.27E-16 | TNFA_C.conidiaHK_WB_48h | -0.28002723 | 0.030440241 |
| rs13082697 | 3 | 46071763 | C | T | 0.5096029 | 1.21E-16 | TNFA_C.conidiaHK_WB_48h | -0.27995786 | 0.030495767 |
| rs13060287 | 3 | 46071868 | G | A | 0.5111396 | 9.18E-17 | TNFA_C.conidiaHK_WB_48h | -0.27988774 | 0.030551607 |
| rs34619093 | 3 | 46072519 | A | G | 0.5041695 | 3.25E-16 | TNFA_C.conidiaHK_WB_48h | -0.27981407 | 0.030606449 |
| rs68087193 | 3 | 46072630 | T | C | 0.5113135 | 8.97E-17 | TNFA_C.conidiaHK_WB_48h | -0.27981407 | 0.030606449 |
| rs2102055 | 3 | 46072704 | A | G | 0.5140265 | 4.75E-17 | TNFA_C.conidiaHK_WB_48h | -0.27981407 | 0.030606449 |
| rs2102056 | 3 | 46072768 | A | G | 0.5114095 | 8.83E-17 | TNFA_C.conidiaHK_WB_48h | -0.27981407 | 0.030606449 |
| rs2088690 | 3 | 46072839 | T | C | 0.5114095 | 8.83E-17 | TNFA_C.conidiaHK_WB_48h | -0.27981407 | 0.030606449 |
| rs34774687 | 3 | 46073314 | C | T | 0.5114574 | 8.73E-17 | TNFA_C.conidiaHK_WB_48h | -0.27974244 | 0.030662915 |
| rs35814488 | 3 | 46073923 | G | A | 0.5114394 | 8.75E-17 | TNFA_C.conidiaHK_WB_48h | -0.27967006 | 0.030719699 |
| rs6790866 | 3 | 46074711 | A | G | 0.5114514 | 8.74E-17 | TNFA_C.conidiaHK_WB_48h | -0.27959692 | 0.030776801 |
| rs6791016 | 3 | 46074858 | A | G | 0.5114394 | 8.76E-17 | TNFA_C.conidiaHK_WB_48h | -0.27952303 | 0.030834222 |
| rs6780028 | 3 | 46075090 | C | A | 0.5114934 | 8.69E-17 | TNFA_C.conidiaHK_WB_48h | -0.27952303 | 0.030834222 |
| rs35454877 | 3 | 46059484 | C | T | 0.5121288 | 7.99E-17 | TNFA_C.conidiaHK_WB_48h | -0.2794921 | 0.030883826 |
| rs71327007 | 3 | 46059249 | T | C | 0.5044473 | 3.14E-16 | TNFA_C.conidiaHK_WB_48h | -0.27945146 | 0.030917765 |
| rs71327006 | 3 | 46058908 | A | G | 0.5044715 | 3.14E-16 | TNFA_C.conidiaHK_WB_48h | -0.27938456 | 0.030979215 |
| rs2088692 | 3 | 46077332 | G | A | 0.5114274 | 8.77E-17 | TNFA_C.conidiaHK_WB_48h | -0.2792622 | 0.03103155 |
| rs34438204 | 3 | 46081306 | C | T | 0.504556 | 3.11E-16 | TNFA_C.conidiaHK_WB_48h | -0.27870789 | 0.031449675 |
| rs72901034 | 3 | 46085321 | G | A | 0.4961586 | 1.34E-17 | TNFA_C.conidiaHK_WB_48h | -0.27820343 | 0.031820988 |
| rs34679077 | 3 | 46087992 | A | G | 0.5051656 | 2.90E-16 | TNFA_C.conidiaHK_WB_48h | -0.2777591 | 0.032138506 |
| rs34863575 | 3 | 46094063 | G | A | 0.5076787 | 2.15E-16 | TNFA_C.conidiaHK_WB_48h | -0.27601744 | 0.033358138 |
| rs35772789 | 3 | 46095104 | G | A | 0.5079254 | 2.08E-16 | TNFA_C.conidiaHK_WB_48h | -0.27559786 | 0.03364501 |
| rs77399277 | 3 | 46096798 | C | T | 0.509008 | 1.80E-16 | TNFA_C.conidiaHK_WB_48h | -0.27501711 | 0.034036318 |
| rs34718164 | 3 | 46098526 | G | C | 0.5071188 | 2.42E-16 | TNFA_C.conidiaHK_WB_48h | -0.27422912 | 0.034566223 |
| rs13096741 | 3 | 46099154 | T | C | 0.5072512 | 2.37E-16 | TNFA_C.conidiaHK_WB_48h | -0.27399745 | 0.034721079 |
| rs35420565 | 3 | 46100138 | G | C | 0.5074198 | 2.32E-16 | TNFA_C.conidiaHK_WB_48h | -0.27352546 | 0.035035462 |
| rs34047915 | 3 | 46100491 | T | C | 0.5072994 | 2.36E-16 | TNFA_C.conidiaHK_WB_48h | -0.27340566 | 0.035115039 |
| rs34766614 | 3 | 46107601 | G | A | 0.500351 | 7.64E-16 | TNFA_C.conidiaHK_WB_48h | -0.27328078 | 0.035194056 |
| rs71327014 | 3 | 46101466 | T | C | 0.5076245 | 2.27E-16 | TNFA_C.conidiaHK_WB_48h | -0.27328515 | 0.03519501 |
| rs71327015 | 3 | 46102173 | C | G | 0.5043145 | 4.35E-16 | TNFA_C.conidiaHK_WB_48h | -0.27328515 | 0.03519501 |
| rs34127208 | 3 | 46103680 | T | G | 0.5047371 | 4.15E-16 | TNFA_C.conidiaHK_WB_48h | -0.27324392 | 0.03522059 |
| rs34460587 | 3 | 46142464 | T | C | 0.5045379 | 4.48E-16 | TNFA_C.conidiaHK_WB_48h | -0.26471059 | 0.040299893 |
| rs13091868 | 3 | 46123043 | G | A | 0.509687 | 2.63E-16 | TNFA_C.conidiaHK_WB_48h | -0.26296235 | 0.041512414 |
| rs13092030 | 3 | 46123055 | T | C | 0.5103595 | 2.38E-16 | TNFA_C.conidiaHK_WB_48h | -0.26296235 | 0.041512414 |
| rs17215981 | 3 | 46124418 | G | A | 0.4971931 | 1.04E-15 | TNFA_C.conidiaHK_WB_48h | -0.26296235 | 0.041512414 |
| rs13089554 | 3 | 46127557 | A | G | 0.4966395 | 1.13E-15 | TNFA_C.conidiaHK_WB_48h | -0.26296235 | 0.041512414 |
| rs13095717 | 3 | 46128542 | A | G | 0.4967673 | 1.11E-15 | TNFA_C.conidiaHK_WB_48h | -0.26296235 | 0.041512414 |
| rs13075528 | 3 | 46128709 | C | T | 0.5001448 | 7.62E-16 | TNFA_C.conidiaHK_WB_48h | -0.26296235 | 0.041512414 |
| rs13095602 | 3 | 46128713 | G | A | 0.4997202 | 8.04E-16 | TNFA_C.conidiaHK_WB_48h | -0.26296235 | 0.041512414 |
| rs71327023 | 3 | 46131225 | G | C | 0.49419 | 1.69E-15 | TNFA_C.conidiaHK_WB_48h | -0.26296235 | 0.041512414 |
| rs13063527 | 3 | 46135690 | A | G | 0.4983724 | 9.69E-16 | TNFA_C.conidiaHK_WB_48h | -0.26296235 | 0.041512414 |
| rs4373103 | 3 | 46138455 | A | G | 0.4974729 | 1.14E-15 | TNFA_C.conidiaHK_WB_48h | -0.26296235 | 0.041512414 |
| rs71327024 | 3 | 46140073 | T | G | 0.5013388 | 6.45E-16 | TNFA_C.conidiaHK_WB_48h | -0.26296235 | 0.041512414 |
| rs71327017 | 3 | 46111213 | C | T | 0.4985668 | 8.96E-16 | TNFA_C.conidiaHK_WB_48h | -0.26292806 | 0.041541923 |
| rs9845382 | 3 | 46111759 | A | G | 0.5066369 | 2.03E-16 | TNFA_C.conidiaHK_WB_48h | -0.26292806 | 0.041541923 |
| rs13069742 | 3 | 46114216 | G | A | 0.5064079 | 2.08E-16 | TNFA_C.conidiaHK_WB_48h | -0.26292806 | 0.041541923 |
| rs13072267 | 3 | 46116010 | A | T | 0.4979167 | 9.53E-16 | TNFA_C.conidiaHK_WB_48h | -0.26292806 | 0.041541923 |
| rs36057789 | 3 | 46117096 | G | A | 0.4975033 | 1.01E-15 | TNFA_C.conidiaHK_WB_48h | -0.26292806 | 0.041541923 |
| rs76647202 | 3 | 46117960 | G | A | 0.4978073 | 9.57E-16 | TNFA_C.conidiaHK_WB_48h | -0.26292806 | 0.041541923 |
| rs71327021 | 3 | 46120517 | G | A | 0.4978316 | 9.43E-16 | TNFA_C.conidiaHK_WB_48h | -0.26292806 | 0.041541923 |
| rs9824651 | 3 | 46108285 | T | C | 0.5120269 | 9.50E-17 | TNFA_C.conidiaHK_WB_48h | -0.26289308 | 0.041571715 |
| rs9825081 | 3 | 46108400 | A | G | 0.5118771 | 9.67E-17 | TNFA_C.conidiaHK_WB_48h | -0.26289308 | 0.041571715 |
| rs2230322 | 3 | 46063329 | C | T | 0.5110856 | 9.20E-17 | TNFA_C.conidiaHK_WB_48h | -0.26135966 | 0.041616076 |
| rs1491951 | 3 | 46141844 | A | G | 0.5083165 | 2.07E-16 | TNFA_C.conidiaHK_WB_48h | -0.25156042 | 0.048700477 |
| rs13075758 | 3 | 46025048 | A | G | 0.5301634 | 4.64E-18 | TNFA_C.conidiaHK_WB_48h | -0.26130281 | 0.048918743 |
| rs34442130 | 3 | 46024529 | A | T | 0.5301222 | 4.67E-18 | TNFA_C.conidiaHK_WB_48h | -0.26121747 | 0.049009203 |
| rs2109069 | 19 | 4719443 | A | G | 0.3056227 | 3.98E-12 | TNFA_Cryptococcus_PBMC_24h | -0.31039826 | 0.041205529 |
| rs7642701 | 3 | 46091058 | G | A | 0.2750678 | 9.82E-10 | TNFA_LPS_WB_48h | 0.13064524 | 0.044941874 |
| rs13098271 | 3 | 46093858 | A | G | 0.277071 | 7.60E-10 | TNFA_LPS_WB_48h | 0.13035264 | 0.045393181 |
| rs35657218 | 3 | 46094462 | G | A | 0.2755462 | 9.52E-10 | TNFA_LPS_WB_48h | 0.13028419 | 0.045504273 |
| rs34906043 | 3 | 46094623 | T | C | 0.2717585 | 1.66E-09 | TNFA_LPS_WB_48h | 0.13025128 | 0.045550436 |
| rs12705891 | 7 | 113317708 | C | T | 0.2753944 | 2.35E-10 | TNFA_Pam3Cys_PBMC_24h | 0.19437631 | 0.028029208 |

* all normal significant cQTLs have been listed

**Table S12: Overlapping variants in GWAS summary statistics and COVID-19 HGI meta analysis of severe respiratory COVID-19 versus population**

| RSid | Chromosome | Position | Effect Allele | Other Allele | Beta | SE | P | P-HGI |
|---|---|---|---|---|---|---|---|---|
| rs8176719 | 9 | 136132909 | T | TC | -0.2772 | 0.052 | 9.93E-08 | 8.88E-07 |
| rs633862 | 9 | 136155444 | T | C | 0.2534 | 0.0515 | 8.79E-07 | 2.80E-06 |

**Table S13: Comparsion polygeneic risk score between male and female in 500FG (N=478) and 300BCG (N=313) using independent sumamry statistics***.**

| Fraction | Risk group | Male -500FG | Female -500[illegible] | OR (95% CI) | Pvalue-500F[illegible] | Male -300BC | Female -300[illegible] | OR (95% CI) | Pvalue-300F[illegible] | Meta-Pvalue** |
|---|---|---|---|---|---|---|---|---|---|---|
| 10% | high risk | 15 | 32 | 0.556 (0.351 [illegible] | 0.998 | 13 | 18 | 0.77 (0.462 - [illegible] | 0.898 | 0.934 |
| 10% | low risk | 28 | 19 | | | 17 | 14 | | | |
| 15% | high risk | 29 | 42 | 0.795 (0.566 [illegible] | 0.935 | 21 | 25 | 0.877 (0.581 [illegible] | 0.798 | 0.822 |
| 15% | low risk | 37 | 34 | | | 24 | 22 | | | |
| 20% | high risk | 38 | 56 | 0.867 (0.645 [illegible] | 0.866 | 28 | 34 | 0.937 (0.657 [illegible] | 0.705 | 0.712 |
| 20% | low risk | 45 | 50 | | | 30 | 32 | | | |
| 25% | high risk | 50 | 68 | 0.924 (0.713 [illegible] | 0.766 | 35 | 43 | 0.95 (0.692 - [illegible] | 0.685 | 0.627 |
| 25% | low risk | 55 | 64 | | | 37 | 41 | | | |
| 30% | high risk | 58 | 84 | 0.897 (0.706 [illegible] | 0.847 | 42 | 51 | 0.979 (0.733 [illegible] | 0.616 | 0.642 |
| 30% | low risk | 66 | 77 | | | 43 | 50 | | | |

* the p value is obtained using the fisher extact test

** the p value is obtained using the meta analyzed z score approach

*** GWAS summary statistics were download from https://genomicc.org/data/ (Eurpean population) E. Pairo-Castineira et.al Nature 2020 , https://doi.org/10.1038/s41586-020-03065-y,

**Table S14: Numbers and basic characteristics of participants included in the study.**

| | 500FG (N = 451) | 300BCG (N = 313) |
|---|---|---|
| **Age (years)** | | |
| mean (SD) | 28.1 (13.5) | 25.9 (10.6) |
| median [Min, Max] | 23 [18, 75] | 23 [18, 71] |
| **Sex** | | |
| Male sex: N (%) | 211 (43.1%) | 141 (43.3%) |
| **BMI** | | |
| mean (SD) | 22.8 (2.91) | 22.5 (2.56) |
| median [Min, Max] | 22.2 [15.1, 34.6] | 22.1 [17.8, 34.2] |
| **Smoking behaviour** | | |
| Smoker *: N (%) | 85 (20.3%) | 63 (20.2%) |
| Missing | 32 | 0 |

SD = standard deviation

* Smokers were classified as people who regularly smoked or did so in the past.

| **Table S15: Numbers and basic characteristics of COVID-19 patients from Nijmegen included CCL25 Olink study** | | | |
|---|---|---|---|
| | Total (n=159) | non-ICU (n=102) | ICU (n=57) |
| **Age (years)** | | | |
| mean (SD) | 62.4 (13.0) | 63.3 (13.5) | 61.0 (12.1) |
| median [Min, Max] | 65 (18, 92) | 66 (28, 92) | 64 (18, 78) |
| **Sex** | | | |
| Male sex: N (%) | 105 (66.0) | 66 (64.7) | 39 (68.4) |
| **BMI** | | | |
| mean (SD) | 27,0 (4.1) | 27.0 (4.3) | 27.1 (3.6) |
| median [Min, Max] | 26.7 (17.8, 41.0) | 26.4 (17.8, 41.0) | 26.8 (19.7, 38.1) |
| **Mortality** | | | |
| non-survivor: N (%) | 21 (13.2) | 8 (7.8) | 13 (22.8) |

| Table S16: Numbers and basic characteristics of COVID-19 patients from Nijmegen included VWF and CXCR16 ELISA study | | | |
|---|---|---|---|
| | Total (n=46) | non-ICU (n=28) | ICU (n=18) |
| **Age (years)** | | | |
| mean (SD) | 62.9 (13.0) | 59.2 (14.4) | 68.6 (7.7) |
| median [Min, Max] | 67 [28, 81] | 60 [28, 81] | 70.5 [47, 81] |
| **Sex** | | | |
| Male sex: N (%) | 29 (63.0) | 16 (57.1) | 13 (72.2%) |
| **BMI** | | | |
| mean (SD) | 27.0 (4.5) | 25.8 (5.6) | 28.1 (3.4) |
| median [Min, Max] | 26.8 [21.0, 36.1] | 24.9 [21.0, 36.1] | 27.6 [23.1, 33.2] |
